# Clinical and genomic diversity of *Treponema pallidum subsp. pallidum:* A global, multi-center study of early syphilis to inform vaccine research

**DOI:** 10.1101/2023.07.19.23291250

**Authors:** Arlene C. Seña, Mitch M. Matoga, Ligang Yang, Eduardo Lopez-Medina, Farhang Aghakanian, Jane S. Chen, Everton B. Bettin, Melissa J. Caimano, Wentao Chen, Jonny A. Garcia-Luna, Christopher M. Hennelly, Yinbo Jiang, Jonathan J. Juliano, Petra Pospíšilová, Lady Ramirez, David Šmajs, Joseph D. Tucker, Fabio Vargas Cely, Heping Zheng, Irving F. Hoffman, Bin Yang, M. Anthony Moody, Kelly L. Hawley, Juan C. Salazar, Justin D. Radolf, Jonathan B. Parr

**Affiliations:** University of North Carolina at Chapel Hill, Chapel Hill, North Carolina, USA; UNC Project Malawi, Lilongwe, Malawi; Dermatology Hospital, Southern Medical University, Guangzhou, China; Centro Internacional de Entrenamiento e Investigaciones Medicas - CIDEIM, Cali, Colombia; Department of Pediatrics, Universidad del Valle; University of Connecticut School of Medicine, Farmington, Connecticut, USA; Universidad ICESI, Cali, Colombia; Division of Dermatology, School of Medicine, Universidad del Valle, Cali, Colombia; Masaryk University, Brno, Czech Republic; Duke University, Durham, North Carolina, USA; Connecticut Children’s, Hartford, Connecticut, USA

**Author notes:** Contributed equally as second authors (Mitch M. Matoga, Ligang Yang, Eduardo Lopez-Medina, Farhang Aghakanian, Jane S. Chen). Contributed equally as senior authors (Justin D. Radolf, Jonathan B. Parr). Correspondence to: Arlene C. Seña, MD, MPH, Department of Medicine, Institute for Global Health and Infectious Diseases, University of North Carolina at Chapel Hill, CB#7030, 130 Mason Farm Road, Chapel Hill, North Carolina 27599.

## Abstract

**Background:** The continuing increase in syphilis rates worldwide necessitates development of a vaccine with global efficacy. We conducted a multi-center, observational study to explore *Treponema pallidum* subsp. *pallidum* (*TPA*) molecular epidemiology essential for vaccine research by analyzing clinical data and specimens from early syphilis patients using whole-genome sequencing (WGS) and publicly available WGS data.

**Methods:** We enrolled patients with primary (PS), secondary (SS) or early latent (ELS) syphilis from clinics in China, Colombia, Malawi and the United States between November 2019 - May 2022. Inclusion criteria included age ≥18 years, and syphilis confirmation by direct detection methods and/or serological testing. *TPA* detection and WGS were conducted on lesion swabs, skin biopsies/scrapings, whole blood, and/or rabbit-passaged isolates. We compared our WGS data to publicly available genomes, and analysed *TPA* populations to identify mutations associated with lineage and geography.

**Findings:** We screened 2,820 patients and enrolled 233 participants - 77 (33%) with PS, 154 (66%) with SS, and two (1%) with ELS. Median age of participants was 28; 66% were *cis*-gender male, of which 43% reported identifying as “gay”, “bisexual”, or “other sexuality”. Among all participants, 56 (24%) had HIV co-infection. WGS data from 113 participants demonstrated a predominance of SS14-lineage strains with geographic clustering. Phylogenomic analysis confirmed that Nichols-lineage strains are more genetically diverse than SS14-lineage strains and cluster into more distinct subclades. Differences in single nucleotide variants (SNVs) were evident by *TPA* lineage and geography. Mapping of highly differentiated SNVs to three-dimensional protein models demonstrated population-specific substitutions, some in outer membrane proteins (OMPs) of interest.

**Interpretation:** Our study involving participants from four countries substantiates the global diversity of *TPA* strains. Additional analyses to explore *TPA* OMP variability within strains will be vital for vaccine development and improved understanding of syphilis pathogenesis on a population level.

**Funding:** National Institutes of Health, Bill and Melinda Gates Foundation

## Introduction

*Treponema pallidum* subsp. *pallidum* (*TPA*), the causative agent of syphilis, is highly invasive - resulting in protean clinical manifestations, adverse pregnancy outcomes, and increased HIV transmission.^1^ In 2020, the World Health Organization estimated 22.3 million prevalent syphilis cases and 7.1 million incident cases worldwide.^2^ Development of a syphilis vaccine will be key to stemming this global epidemic.^3^

Long-standing problems with *in vitro TPA* cultivation have complicated syphilis research in the past.^4^ Fortunately, advances in *TPA* whole-genome sequencing (WGS) performed directly from biological specimens have rapidly increased available genomes and our understanding of the pathogen’s genetic diversity.^5–8^ Multiple studies have demonstrated two deep-branching *TPA* phylogenetic lineages differentiated by their relationship to the Nichols or SS14 reference strains.^5–8^ Mapping of single nucleotide variants (SNVs) to structural models for *TPA* outer membrane proteins (OMPs) can identify regions of sequence variability relevant to syphilis vaccine design and pathogenesis.^9^

Most *TPA* genomic sequences have originated from strains circulating in high-income countries, ^5–8^ or from retrospective specimen analyses with limited accompanying clinical data. We hypothesized that *TPA* genetic diversity may exist between populations with differing demographics, clinical manifestations, and HIV status, as well as geography from low-middle income countries. Thus, we developed a global consortium to collect individual-level data and biological specimens for WGS analyses to explore strain diversity. By leveraging diverse patient populations, we evaluated characteristics associated with *TPA* strains, compared them to global strains published to-date, and analyzed SNVs among infecting strains to inform vaccine development.

## Methods

### Study participant population

We conducted a multi-center, observational, cross-sectional study by recruiting and enrolling a convenience sample of patients with early syphilis (primary, secondary, early latent syphilis) who presented to a provincial dermatology hospital in Guangzhou, China; a public healthcare sector network in Cali, Colombia; a sexually transmitted infection (STI) clinic in Lilongwe, Malawi, and an infectious disease clinic in Chapel Hill, North Carolina, United States (US), between November 28, 2019 and May 27, 2022. The study was approved by the institutional review board (IRB) at the University of North Carolina at Chapel Hill (UNC IRB protocol number 19-0311) and IRBs representing other sites (Supplementary Methods).

Eligibility screening criteria included age ≥ 18 years and one of the following: suspected primary syphilis (PS) with ulcerative lesions (*e.g*., chancres); secondary syphilis (SS) with mucocutaneous lesions (*e*.*g*., rashes, condyloma lata); or early latent syphilis (ELS) with seroconversion of serologies or exposure to early syphilis within the last 12 months (Supplementary Methods). Additional enrollment criteria included laboratory confirmation of PS with darkfield microscopy (DFM), polymerase chain reaction (PCR) or syphilis serology; SS with a non-treponemal antibody titer >1:8 and/or positive treponemal antibody test; and ELS with positive serologies. Individuals were excluded if they had antibiotics active against syphilis within 30 days of enrollment.

### Study procedures

Sexual histories, physical examinations, and HIV/STI testing were conducted as per routine care. Participants underwent screening using DFM from anogenital lesions, non-treponemal and/or rapid treponemal antibody testing. Urine pregnancy testing was performed for women of childbearing age.

Following written consent, participants underwent clinical data collection, photography of lesions, and specimen collection including anogenital lesion swabs, punch skin biopsies or scrapings of rashes. Multiple swabs were collected if participants had more than one lesion. Lesion swabs, skin biopsies/scrapings were collected in DNA/RNA Shield (Zymo Research; Irvine, California, US). Whole blood was collected for serum, plasma, and peripheral blood mononuclear cells depending on the clinical site. Lesion exudates and whole blood were collected for rabbit infectivity testing (RIT)^10^ in Guangzhou (Supplementary Methods).

Following specimen collection, all participants received treatment according to national guidelines. Behavioral counseling and partner services were provided per routine care.

### Data collection

Demographic data, sexual histories, clinical characteristics, laboratory results, and treatment were documented in case report forms (CRFs). English CRFs (Supplementary Methods) were translated into local languages. CRF data and photographs were uploaded into a secure Research Electronic Data Capture database.

### Sample processing and quantitative PCR

We performed quantitative PCR (qPCR) targeting the *TPA* polymerase I gene (*polA*/*tp0105*) on DNA extracted from specimens including lesion swabs, skin biopsies/scrapings, and whole blood.^11^ Any sample with >0 copy numbers was considered *TPA* positive. In addition, qPCR was conducted on rabbit-passaged isolates from PS lesion exudates and SS whole blood samples from Guangzhou (Supplementary Methods).

### TPA whole-genome sequencing

*TPA* WGS was performed on specimens with detectable DNA from enrolled participants and on other specimens including i) DFM-negative lesion swabs from participants screened in Lilongwe, Malawi, and ii) PS and SS lesions from participants enrolled in Cali, Colombia in separate IRB-approved studies (IRB reference numbers 1281, 1289, 1315). In general, available specimens with ≥40 *polA* copies/µL underwent *TPA* enrichment and WGS (Supplementary Methods). *TPA* enrichment was performed using parallel, pooled whole-genome amplification^12^ and custom 120-nucleotide RNA oligonucleotide baits (Agilent Technologies, Santa Clara, CA, US; Sure Select XT Low Input in Chapel Hill or Sure Select XTHS2 in Guangzhou). Pooled, *TPA*-enriched libraries were sequenced using MiSeq or NovaSeq platforms (Illumina, San Diego, CA, US) with paired-end, 150bp reads.

### Genome assembly, variant calling, and analysis

Sequencing data, along with publicly available data from geographically diverse locations,^8^ were analyzed using an adaptation of our bioinformatic pipeline,^13^ described at (https://github.com/IDEELResearch/Tpallidum_genomics). After read trimming, alignment, filtering and variant calling, consensus genomes were aligned using *MAFFT* (v7.490). Repetitive and putative recombination regions were masked using *Gubbins* (v3.2), before construction of maximum likelihood (ML) trees using *RAxML* (v8.2.12) with 1,000 rapid bootstraps. Trees were visualized and annotated in R (v4.1.2) using the *ggtree* package (v3.2.1), with clades and subclades assigned to facilitate comparison with published analyses.^8^ We identified macrolide-resistant strains using *ARIBA* (v2.14.6) on trimmed and host-removed *TPA* sequences, and a customized database to identify 23S rRNA A2058G and A2059G variants.^14, 15^ To evaluate population structure, we analyzed our samples alongside publicly available *TPA* genomes from Sequence Read Archive (SRA, Supplementary Table 1), using principal component analysis (PCA) and *fastBAPS* (Supplementary Methods) to identify *TPA* populations.^13^ SNVs were mapped to protein structural models of genes with high frequency, population-informative mutations, and visualized using *Chimera* (v1.16).^16^

### Data analyses

Descriptive statistics were used to describe participant characteristics. See Supplementary Methods for analysis of *TPA* qPCR and sequencing data. All analyses were performed using R (v.4.0.2).

## Results

### Characteristics of study participants

Over the study period, we screened 2,820 individuals with suspected syphilis (Figure 1); 2,407 did not meet inclusion criteria, while 181 declined participation. We enrolled 232 unique participants, including one enrolled in 2020 with SS and in 2021 with PS. Of the 233 total participant enrollments, 77 (33%) had PS, 154 (66%) had SS, and 2 (1%) had ELS (Table 1). Only two participants were enrolled from the US site, which halted enrollment due to the COVID-19 pandemic.

**Figure 1.**
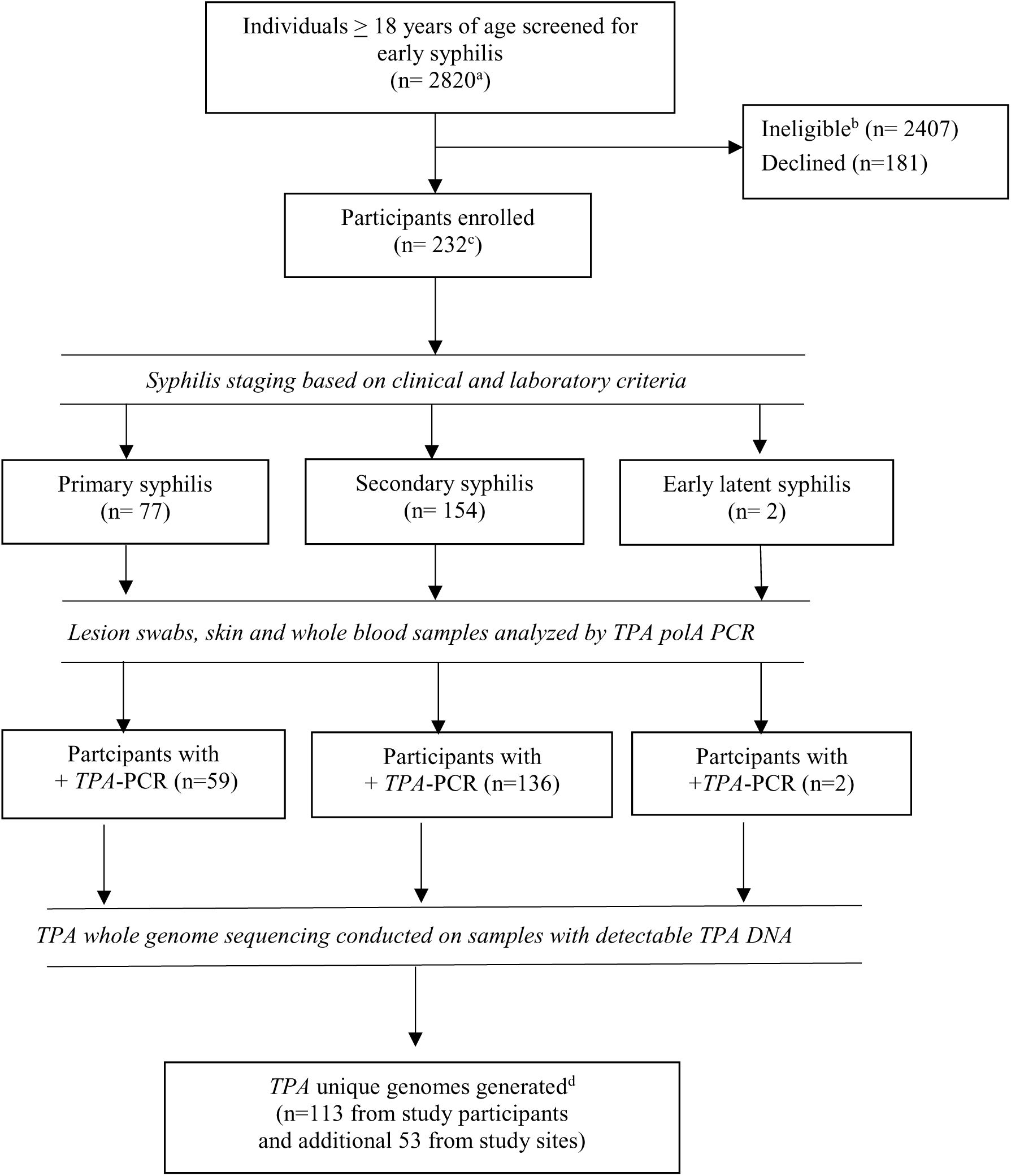
Screening, enrollment and testing algorithm for participants with early syphilis in this study. *TPA* = *Treponema pallidum* sub. *pallidum*; PCR = polymerase chain reaction; DNA = deoxyribonucleic acid ^a^Screening based on suspected primary, secondary or early latent syphilis among patients from China, Colombia, Malawi and US clinical sites. ^b^ Includes individuals who refused screening procedures in Malawi. ^c^ One participant was enrolled twice, initially with secondary syphilis and then with primary syphilis. ^d^ Total of unique genomes generated was 166, which includes sequences from enrolled study participants, 43 participants screened but not enrolled from Malawi with darkfield-negative lesions and 10 individuals from Colombia enrolled in a longitudinal study with available specimens

**Table 1.** Participant characteristics by site of enrollment in the multi-center study.

|  | China<br>n (%) | Colombia<br>n (%) | Malawi<br>n (%) | US<br>n (%) | Total<br>n (%) |
| --- | --- | --- | --- | --- | --- |
| **Total** | **69** | **74** | **88** | **2** | **233** |
| **Demographic Characteristics** |  |  |  |  |  |
| Age (median, IQR) | 27 (22, 32) | 28 (22, 39) | 28 (22, 33) | (51, 55)<sup>‡</sup> | 28 (22, 35) |
| Gender/sex |  |  |  |  |  |
| Cisgender female | 11 (16) | 20 (27) | 46 (52) | 0 (0) | 77 (33) |
| Cisgender male | 58 (84) | 53 (72) | 42 (48) | 1 (50) | 154 (66) |
| Transgender female | 0 (0) | 1 (1) | 0 (0) | 1 (50) | 2 (1) |
| Sexual orientation |  |  |  |  |  |
| Heterosexual | 44 (64) | 31 (42) | 86 (98) | 0 (0) | 161 (69) |
| Gay, bisexual, or other | 24 (35) | 43 (58) | 0 (0) | 1 (50) | 68 (29) |
| Unknown or unwilling to state | 1 (1) | 0 (0) | 2 (2) | 1 (50) | 4 (2) |
| Race/Ethnicity |  |  |  |  |  |
| African American | 0 (0) | 3 (4) | 0 (0) | 1 (50) | 4 (2) |
| African American; Hispanic/Latino | 0 (0) | 14 (19) | 0 (0) | 0 (0) | 14 (6) |
| Asian | 69 (100) | 0 (0) | 0 (0) | 0 (0) | 69 (30) |
| Black African | 0 (0) | 0 (0) | 88 (100) | 1 (50) | 89 (38) |
| Hispanic/Latino | 0 (0) | 48 (65) | 0 (0) | 0 (0) | 48 (21) |
| White; Hispanic Latino | 0 (0) | 9 (12) | 0 (0) | 0 (0) | 9 (4) |
| **Behavioral Characteristics** |  |  |  |  |  |
| Sexual partners in past year<sup>†</sup> | 3 (2, 5) | 2 (1, 4) | 1 (1, 2) | (3, 4)<sup>‡</sup> | 2 (1, 3) |
| New sexual partners in past year<sup>†</sup> | 1 (1, 3) | 1 (0, 3) | 1 (0, 1) | (3, 4)<sup>‡</sup> | 1 (0, 2) |
| Exchanged goods/money/favors for sex with a partner from past year<sup>‡</sup> | 13 (19) | 11 (15) | 31 (35) | 1 (50) | 56 (24) |
| Prior intravenous drug use | 0 (0) | 1 (1) | 0 (0) | 0 (0) | 1 (<1) |
| **Medical History** |  |  |  |  |  |
| HIV co-infection |  |  |  |  |  |
| Not Living with HIV | 68 (99) | 37 (50) | 72 (82) | 0 (0) | 177 (76) |
| Living with HIV | 1 (1) | 37 (50) | 16 (18) | 2 (100) | 56 (24) |
| Pregnant | 0 (0) | 5 (7) | 2 (2) | 0 (0) | 7 (3) |
| History of syphilis or GUD | 6 (9) | 24 (32) | 7 (8) | 1 (50) | 38 (16) |
| History of other STIs | 15 (22) | 17 (23) | 13 (15) | 2 (100) | 47 (20) |
| **Clinical Characteristics** |  |  |  |  |  |
| Syphilis stage |  |  |  |  |  |
| Primary syphilis | 18 (26) | 15 (20) | 44 (50) | 0 (0) | 77 (33) |
| Secondary syphilis | 51 (74) | 58 (78) | 44 (50) | 1 (50) | 154 (66) |
| Early latent syphilis | 0 (0) | 1 (1) | 0 (0) | 1 (50) | 2 (1) |
| Darkfield microscopy<sup>§</sup> |  |  |  |  |  |
| Positive | 12 (17) | 5 (7) | 59 (67) | 0 (0) | 76 (33) |
| Negative | 6 (9) | 9 (12) | 18 (20) | 0 (0) | 33 (14) |
| Equivocal | 0 (0) | 4 (5) | 0 (0) | 0 (0) | 4 (2) |
| Not applicable | 51 (74) | 56 (76) | 11 (12) | 2 (100) | 120 (52) |
| Nontreponemal antibody test |  |  |  |  |  |
| Negative | 9 (13) | 1 (1) | 10 (11) | 0 (0) | 20 (9) |
| Reactive | 60 (87) | 73 (99) | 78 (89) | 2 (100) | 213 (91) |
| Nontreponemal titer (median, IQR) | 32 (16, 64) | 32 (16, 64) | 32 (32, 128) | (64, 128)<sup>‡</sup> | 32 (16, 64) |
| Treponemal antibody test |  |  |  |  |  |
| Negative | 2 (3) | 0 (0) | 0 (0) | 0 (0) | 2 (1) |
| Positive | 67 (97) | 74 (100) | 78 (89) | 2 (100) | 221 (95) |
| Not performed<sup>¶</sup> | 0 (0) | 0 (0) | 10 (11) | 0 (0) | 10 (4) |
| Treatment for syphilis |  |  |  |  |  |
| Benzathine penicillin 2.4mu IM x 1 wk | 10 (14) | 23 (31) | 27 (31) | 2 (100) | 62 (27) |
| Benzathine penicillin 2.4mu IM x 2 wks | 45 (65) | 0 (0) | 0 (0) | 0 (0) | 45 (19) |
| Benzathine penicillin 2.4mu IM x 3 wks | 1 (1) | 50 (68) | 60 (68) | 0 (0) | 111 (48) |
| Doxycycline 100mg orally twice daily x 14 days | 12 (17) | 0 (0) | 1 (1) | 0 (0) | 13 (6) |
| Doxycycline 100mg orally twice daily x 28 days | 1 (1) | 1 (1) | 0 (0) | 0 (0) | 2 (1) |

IQR=interquartile range; GUD=genital ulcer disease; STI= sexually transmitted infections; IM=intramuscular
<sup>‡</sup>n=2, both values listed<sup>§</sup>Darkfield only performed on participants with genital ulcers, condyloma lata, and mucous patches<sup>†</sup>Sex exchanged for goods/money/favors was assessed by partner for up to 5 sexual partners from the past year. If no partners named, classified as no.<sup>¶</sup>At Malawi site, treponemal antibody testing was only performed for participants with a positive nontreponemal test.
NOTE: Includes two enrollments of a participant who enrolled twice in China.

Among participant enrollments, the median age was 28 years (interquartile range [IQR]: 22, 35). 66% (n=154) were *cis*-gender male, 33% (n=77) were *cis*-gender female, and 1% (n=2) were *trans*-gender female (Table 1). Forty-four percent (n=67) of *cis*-gender male participants reported identifying as “gay”, “bisexual”, or “other sexuality.” Among *cis*-gender women, 9% (n=7) were pregnant at enrollment. Study participants were 30% (n=69) Asian, 38% (n=89) Black African, and 30% (n=71) Hispanic/Latino. The median number of reported sexual partners in the past year was two (interquartile range [IQR]: 1,3), but 24% (n=56) reported exchanging goods, money or favors for sex in the past year (Table 1). Twenty-four percent (n=56) of participants were living with HIV or diagnosed with HIV at enrollment, 16% (n=38) had a history of prior syphilis or genital ulcer disease, and 20% (n=47) had a history of other STIs.

### Clinical and laboratory findings

DFM was performed from lesion swabs in 113 participants, 76 (67%) of whom were positive. Among all participants, 213 (91%) had reactive non-treponemal antibody tests and 221 (95%) had positive treponemal tests. The median non-treponemal titer was 1:32 for participants with PS (IQR: 1:8, 1:64) and SS (IQR: 1:32, 1:64). We observed a variety of clinical manifestations (Figure 2), including single and multiple anogenital ulcers in PS. SS manifestations ranged from typical maculopapular rashes to ulcerating, crusting lesions consistent with lues maligna, and extensive anogenital condyloma lata. Overall, 197 (85%) participants had specimens with detectable *TPA* DNA (Figure 1). *TPA* burdens varied by syphilis stage and clinical manifestations (Figure 2), as well as between specimen types and clinical sites (Table 2). Skin biopsies/scrapings and whole blood specimens had lower geometric mean copy numbers than lesion swabs (Supplementary Results).

**Figure 2.**
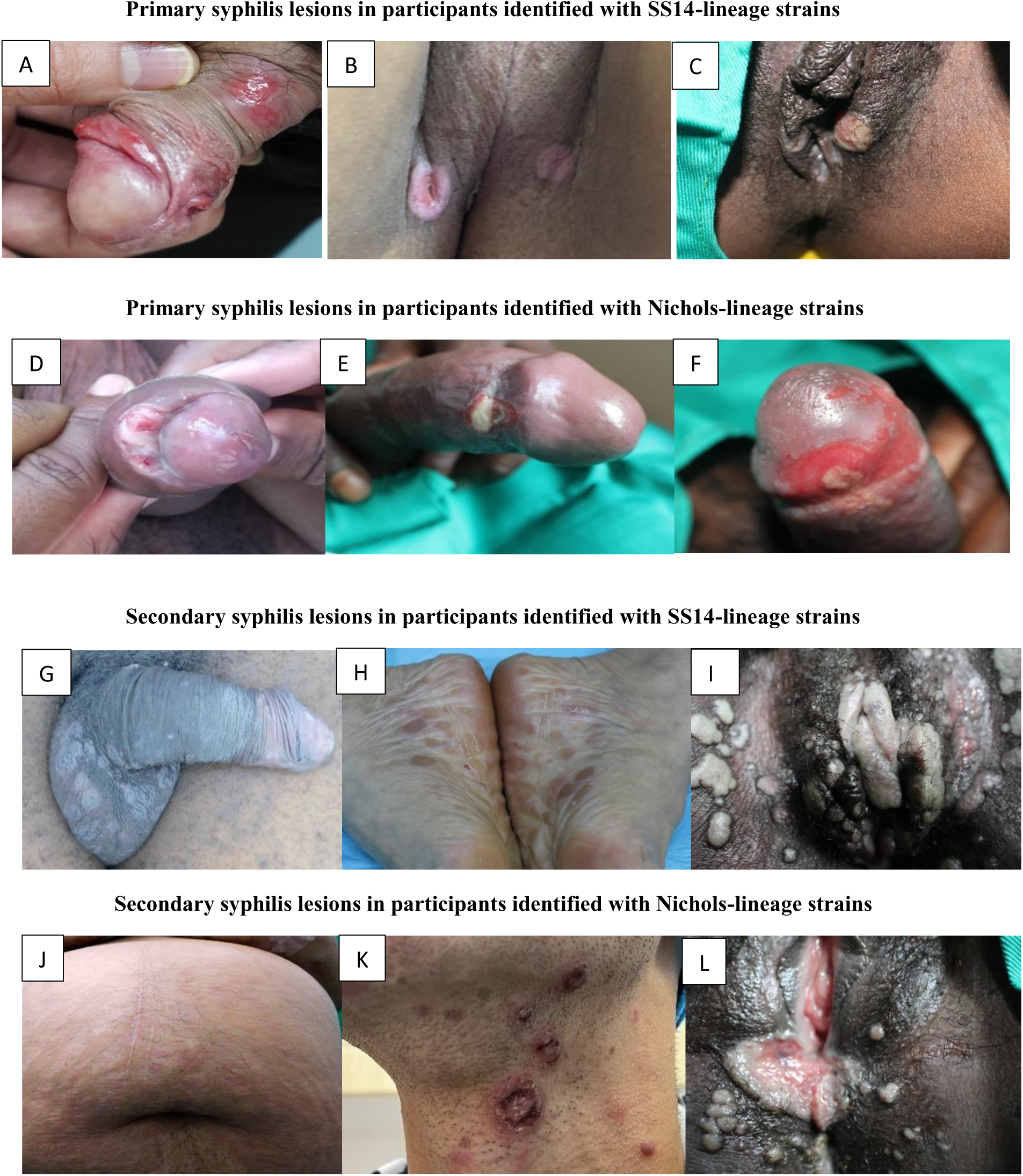
**Clinical manifestations among study participants with primary and secondary syphilis**, by *TPA* clade, geographical location, and quantitative PCR result (copies/µl). A - multiple, shallow penile ulcers (China; lesion swab = 22 copies/µl); B - multiple, deep perineal ulcers (Colombia; lesion swab = 2117 copies/µl); C - multiple vulvar chancres (Malawi; 8764 copies/µl); D - penile ulcer underneath the foreskin with a purulent discharge (Colombia; lesion swab = 3034 copies/µl); E and F - penile chancres with purulent base (Malawi; lesion swabs = 6897 and 2281 copies/µl, respectively); G - diffuse scaly, macular rash involving testicles (Colombia, skin biopsy = 53 copies/µl); H - hyperpigmented macules involving soles (China, skin biopsy = 305 copies/µl); I - extensive condylomata lata involving labia and perineum (Malawi; lesion swab = 2281 copies/µl); J - erythematous macular rash in pregnancy (Colombia; skin biopsy = 2460 copies/µl); K - diffuse rash with crusting lesions consistent with lues maligna (China; skin biopsy = 250 copies/µl); L - moist, condylomata lata involving vulvar area and perineum (Malawi; lesion swab = 6897 copies/µl). *TPA* = *Treponema pallidum* sub. *pallidum;* PCR= polymerase chain reaction

**Table 2:** Qualitative and quantitative *T. pallidum polA* PCR results based on stage of syphilis, specimen type and site of enrollment.

| Characteristic | China<sup>a</sup> |  | Colombia<sup>b</sup> |  | Malawi<sup>c</sup> |  | USA<sup>c</sup> |  |
| --- | --- | --- | --- | --- | --- | --- | --- | --- |
|  | Qualitative *polA* PCR Results |  |  |  |  |  |  |  |
|  | n | (%) | n | (%) | n | (%) | n | (%) |
|  | By Participant and Stage (n=233) |  |  |  |  |  |  |  |
| **Participants with primary syphilis** |  |  |  |  |  |  |  |  |
| Negative (0 copies/μl) | 2 | (11) | 4 | (27) | 12 | (27) | .. | .. |
| Positive (>0 copies/μl) | 16 | (89) | 11 | (73) | 32 | (73) | .. | .. |
| **Participants with secondary syphilis** |  |  |  |  |  |  |  |  |
| Negative (0 copies/μl) | 0 | (0) | 10 | (17) | 8 | (18) | 0 | (0) |
| Positive (>0 copies/μl) | 51 | (100) | 48 | (83) | 36 | (82) | 1 | (100) |
| **Participants with early latent syphilis** |  |  |  |  |  |  |  |  |
| Negative (0 copies/μl) | .. | .. | 0 | (0) | .. | .. | 0 | (0) |
| Positive (>0 copies/μl) | .. | .. | 1 | (100) | .. | .. | 1 | (100) |
|  | By Specimen Type (n=395) |  |  |  |  |  |  |  |
| **Lesion swab** |  |  |  |  |  |  |  |  |
| Negative (0 copies/μl) | 2 | (12) | 8 | (22) | 12 | (16) | .. | .. |
| Positive (>0 copies/μl) | 15 | (88) | 28 | (78) | 65 | (84) | .. | .. |
| **Skin biopsy** |  |  |  |  |  |  |  |  |
| Negative (0 copies/μl) | 1 | (2) | 15 | (27) | .. | .. | 0 | (0) |
| Positive (>0 copies/μl) | 51 | (98) | 40 | (73) | .. | .. | 1 | (100) |
| **Skin scraping** |  |  |  |  |  |  |  |  |
| Negative (0 copies/μl) | .. | .. | .. | .. | 12 | (63) | .. | .. |
| Positive (>0 copies/μl) | .. | .. | .. | .. | 7 | (37) | .. | .. |
| **Whole blood** |  |  |  |  |  |  |  |  |
| Negative (0 copies/μl) | 39 | (57) | 29 | (43) | .. | .. | 0 | (0) |
| Positive (>0 copies/μl) | 30 | (43) | 39 | (57) | .. | .. | 1 | (100) |
|  | Quantitative *polA* Results (copies/μl) (n=277)<sup>d</sup> |  |  |  |  |  |  |  |
|  | n | GMean (Range) | n | GMean (Range) | n | GMean (Range) | n | GMean (Range) |
| **Primary syphilis** |  |  |  |  |  |  |  |  |
| Lesion swabs |  |  |  |  |  |  |  |  |
| Darkfield positive | 12 | 39 (1 - 862) | 5 | 194 (11 - 8835) | 31 | 1016 (12 - 10438) | .. | .. |
| Darkfield negative | 3 | 29 (2 - 2250) | 4 | 20 (3 - 600) | 1 | 1996 (n/a) | .. | .. |
| Darkfield equivocal | .. | .. | 1 | 46200 (n/a) | .. | .. | .. | .. |
| Darkfield not performed | .. | .. | 1 | 9942 (n/a) | .. | .. | .. | .. |
| Skin biopsy | 1 | 1780 (n/a) | -- | -- | .. | .. | .. | .. |
| Blood | 3 | 6 (<1 - 38) | 5 | <1 (<1, 2) | .. | .. | .. | .. |
| **Secondary syphilis** |  |  |  |  |  |  |  |  |
| Lesion swabs |  |  |  |  |  |  |  |  |
| Darkfield positive | .. | .. | .. | .. | 16 | 1858 (115 - 82147) | .. | .. |
| Darkfield negative | .. | .. | 1 | <1 (n/a) | 17 | 2830 (235 - 20143) | .. | .. |
| Darkfield equivocal | .. | .. | 1 | 107 (n/a) | .. | .. | .. | .. |
| Darkfield not performed | .. | .. | 15 | 33 (<1 - 3294) | .. | .. | .. | .. |
| Skin biopsies | 50 | 40 (1 - 10200) | 40 | 41 (<1 - 5249) | .. | .. | 1 | 341 (n/a) |
| Skin scrapings | .. | .. | .. | .. | 7 | 160 (7-7996) | .. | .. |
| Blood |  |  |  |  |  |  |  |  |
| NT antibody titer ≤ 1:32 | 14 | 14 (<1 - 3850) | 17 | 2 (<1 - 8) | .. | .. | .. | .. |
| NT antibody titer >1:32 | 13 | 2 (<1, 232) | 16 | 2 (<1 - 17) | .. | .. | .. | .. |
| **Early latent syphilis** |  |  |  |  |  |  |  |  |
| Blood | .. | .. | 1 | <1 (n/a) | .. | .. | 1 | 2 (n/a) |

PCR= polymerase chain reaction; GMean = geometric mean; n/a = not applicable; NT=nontreponemal antibody titer
<sup>§</sup>Darkfield only performed on participants with genital ulcers, ulcerated condyloma lata, and mucous patches (depending on site)
<sup>a</sup>Gmeans for *TPA* PCR results were calculated from one specimen per participant performed at SMU, with the exception of one participant with two different specimens from 2 separate enrollments.
<sup>b</sup>Gmeans for *TPA* PCR results were calculated from multiple specimens per participant performed at CIDEIM.
<sup>c</sup>Gmeans for *TPA* PCR results were calculated from multiple specimens per participant performed at UNC.
<sup>d</sup>Quantitative PCR geometric mean values were calculated from PCR positive specimens.

### Genomic diversity of infecting TPA strains

WGS was conducted on 174 samples, achieving acceptable coverage (Supplementary Methods) for phylogenomic analysis in 166 (95%) (Figure 1). These included *TPA* genomes from 113 (97%) of 117 participants with sufficient *TPA* copy numbers, 43 individuals from Malawi with DFM-negative lesions, and ten individuals from Colombia enrolled in a separate study.^17^ Samples collected at both visits from the reinfected participant had *TPA* copy numbers insufficient for WGS. In total, our phylogenomic analysis (Figure 3) included 166 new genomes from this study, 62 recently published genomes from diverse geographical locations,^8^ and five reference genomes (233 total genomes).

**Figure 3.**
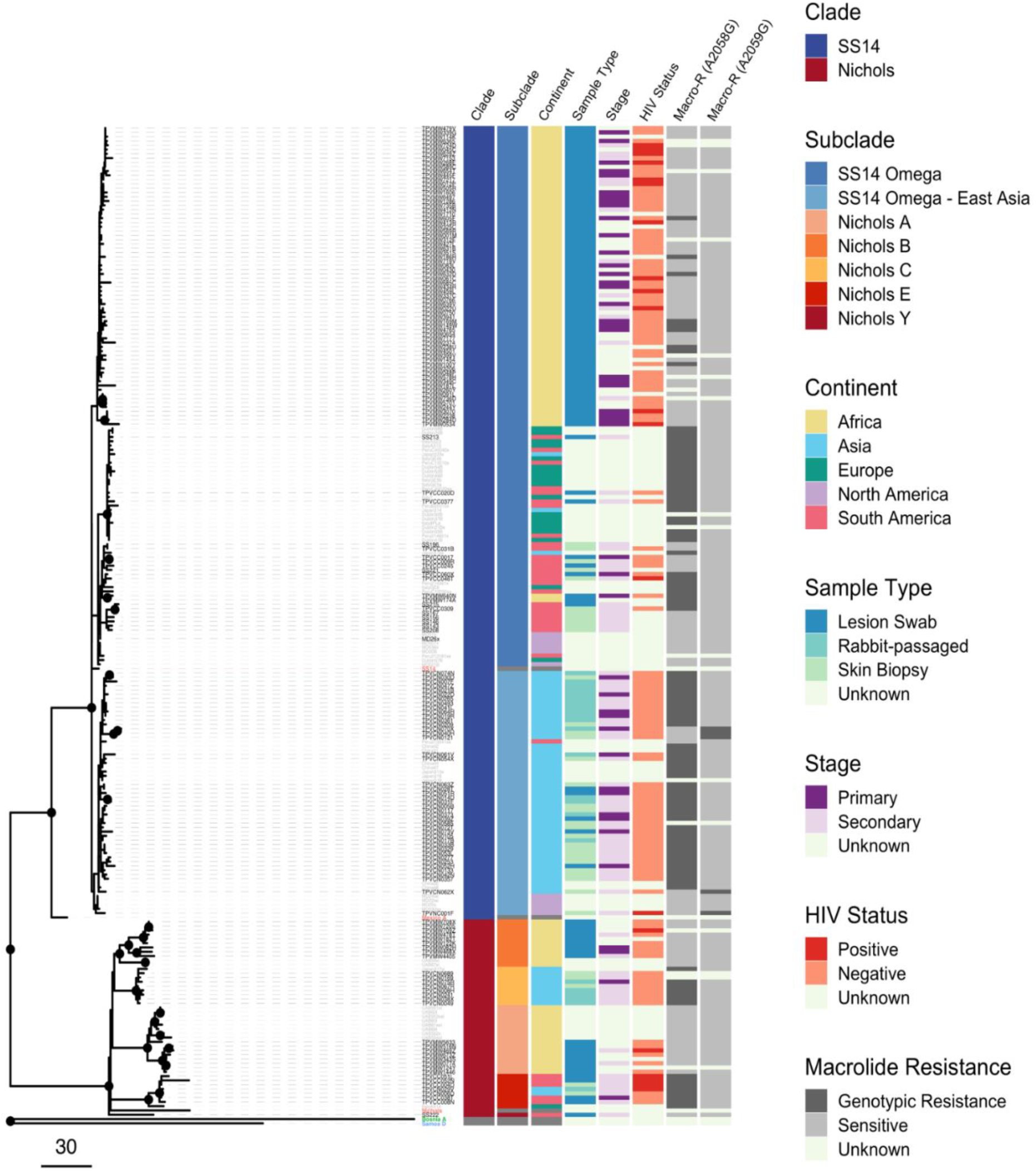
Recombination-masked *TPA* whole-genome phylogeny. derived from 166 individuals in this study,^a^ 62 recently published genomes, and 5 reference genomes (*TPA*, red; *Treponema pallidum* subsp. *pertenue*, blue; *Treponema pallidum* subsp. *endemicum*, green). Nodes with >80% bootstrap support are highlighted with a black circle. *TPA* = *Treponema pallidum* sub. *Pallidum* ^a^ Includes 43 DFM-negative specimens from Lilongwe, Malawi, and 10 specimens from Cali, Colombia, collected in a prior study.

Although 133 (80%) strains belonged to the SS14 lineage, we observed genomically diverse strains across clinical sites (Figure 3). SS14-lineage and Nichols-lineage strains were similarly distributed among groups with differing age, sex/gender, race/ethnicity, sexual orientation and behaviors, syphilis stage and HIV status (Supplementary Figure 1); clinical manifestations did not clearly differ by lineage (Figure 2). SS14-lineage strains, however, clustered by country with all Malawi SS14-lineage strains grouped within the previously described SS14-Omega subclade^8^ (Figure 3). While only 33 (20%) of our genomes belonged to the Nichols clade, Nichols-lineage strains were identified across continents and diverse participants (Figure 4). Compared to SS14-lineage strains, Nichols-lineage strains clustered into more distinct subclades and exhibited longer branch lengths, consistent with greater genetic divergence. Half of participants in the Nichols E subclade were living with HIV. This subclade consisted of strains collected from Colombia and China, as well as one published genome from Europe from an individual with unknown HIV status.

**Figure 4:**
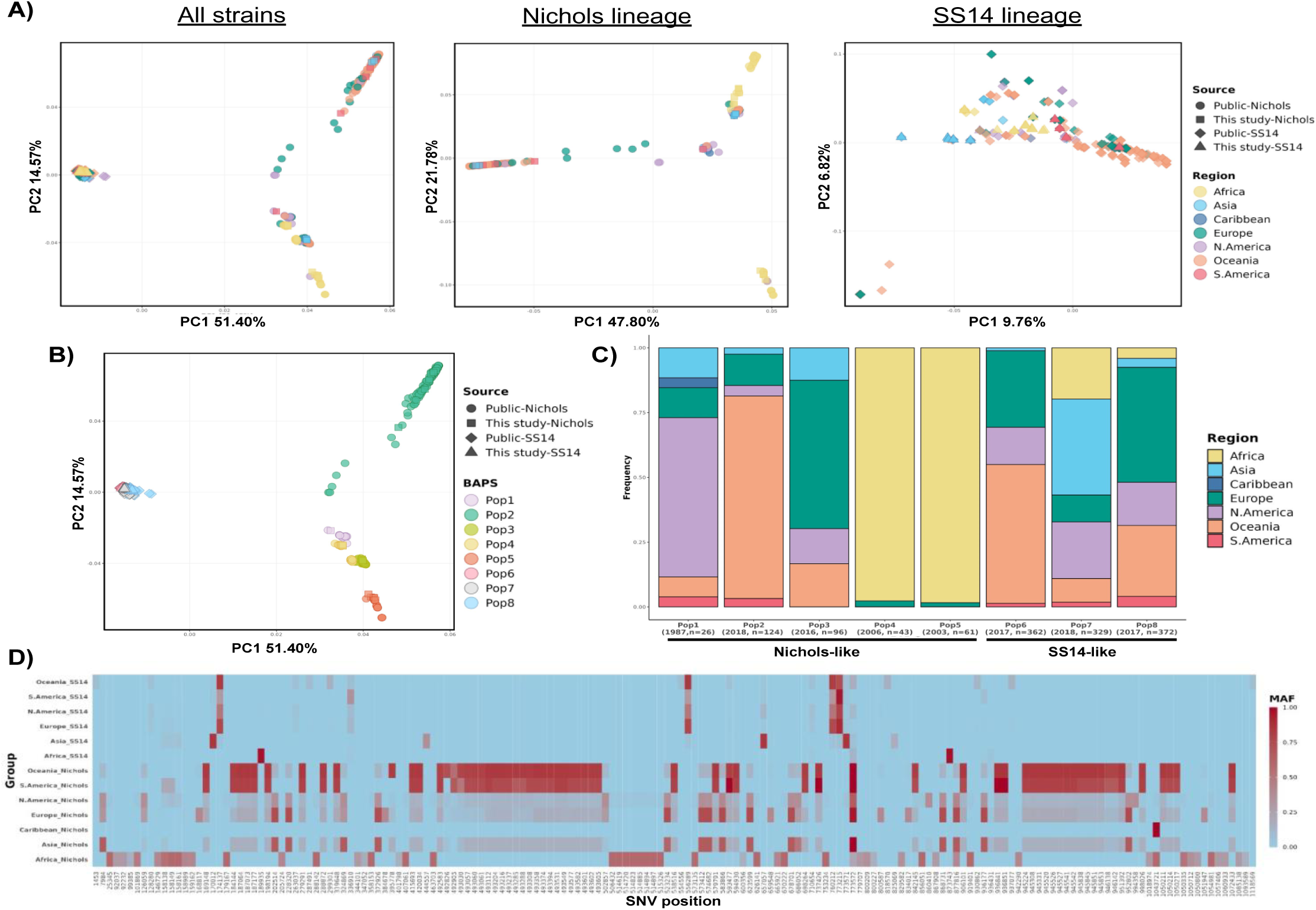
Global TPA population structure. A) Principal components analyses (PCA) of global *TPA* strains, overall and by lineage. Analysis of 1,413 *TPA* genomes derived from diverse studies in global sites confirms that membership in Nichols (n=350) vs. SS14 (n=1,063) clade accounts for the majority of genetic variation (excluding *tpr* family, *tp0470,* and *arp* genes). When PCA was restricted to members of the same clade, more distinct population structure is evident among Nichols-like strains. B) Annotation by *TPA* population determined using *baps* Bayesian modeling confirmed C) five Nichols- and three SS14-like populations with distinct geographical composition and temporal sampling. Median year of sample collection and number of samples are included. D) Allele frequencies of lineage-informative, highly differentiated (Fisher exact *p* < 4.451×10^-6^) SNVs reveal lineage- and geography-specific mutation patterns. SNV coordinates reference the Nichols genome; annotated SNVs are described in Supplementary Figure 4. *TPA* = *Treponema pallidum* sub. *pallidum; SNV* = single nucleotide variant

Macrolide susceptibility varied by geography, with A2058G or A2059G mutations in 75 (47%) of 159 samples with calls for both loci (Figure 3). The SS14 Omega East-Asia subclade had the highest proportion of macrolide resistance mutations. The proportion of specimens with resistance mutations ranged from 100% in China (n=49) and the US (n=1), to 50% (n=13) in Colombia and 15% (n=12) in Malawi.

### TPA genetic population structure

Principal components analysis of 1,413 *TPA* genomes confirmed that our study strains reflect the global diversity of strains across published studies (Supplementary Figure 2, Supplementary Table 1).^5–8^ Over half (51.4%, principal component one) of genetic variation in our recombination-masked analysis was explained by membership in the SS14 versus Nichols clades, with more distinct population structure observed in Nichols-lineage strains (Figure 4A). Using Bayesian modeling, we identified eight distinct *TPA* populations worldwide. These included five Nichols-lineage (Pop1-Pop5, n=350) and three SS14-lineage populations (Pop6-Pop8, n=1063) (Figure 4B). Within the Nichols-lineage, one population was dominated by well-characterized laboratory strains (Pop1), one was dominated by Australian samples (Pop2), and two (Pop4-Pop5) by African samples. Pop4-Pop5 samples originated from Madagascar (n_Pop 4_ =31 and n_Pop 5_ =52), Malawi (n_Pop 4_ =7 and n_Pop 5_ =7) and Zimbabwe (n_Pop 4_ =4 and n_Pop 5_ =1), with median collection dates before 2010 (Figure 4C). *TPA* populations determined by Bayesian modeling were generally concordant with previously described Nichols subclades, whereas concordance with SS14 subclades was less clear (Supplementary Figure 3). We identified *TPA* strains from all eight populations in our study, with the greatest number in the SS14-lineage Pop7 and Pop8 (Supplementary Table 2).

We found 103 SNVs that were fixed or absent in all SS14- or Nichols-lineage strains (Supplementary Table 3), including 57 non-synonymous (missense), 30 synonymous (sense), and 16 upstream (possible promoter disrupting) SNVs. Among 344 biallelic SNVs with minor allelic frequencies (MAF) >1%, there were 169 significantly differentiated SNVs between the two lineages (Fisher’s exact p< 4.451×10^-6^). These include 118 non-synonymous, 39 synonymous, and 12 upstream SNVs (Supplementary Table 4) collectively referred to as ‘highly differentiated’.

We observed an accumulation of non-synonymous fixed SNVs in *lptD* (*tp0515*), which encodes a homolog for the Gram-negative OMP that mediates the final step in the transport of lipopolysaccharide to the outer leaflet of the outer membrane,^9^ and in *tp0179* which encodes a hypothetical protein. The highest number of highly differentiated SNVs (including two fixed and 21 highly differentiated non-synonymous SNVs) were observed in *tp0462*, encoding a putative lipoprotein of unknown function. Our results also showed an accumulation of non-synonymous, highly differentiated SNVs in *tp0865* and *tp0858*, encoding two members of the FadL family of fatty acid importers, and the *tolC* homologue *tp0966*, which encodes the outer membrane factor (OMF) component of an efflux pump.^8^ Overall, 21% of non-synonymous fixed and 26% of non-synonymous highly differentiated SNVs occurred in known or putative genes for OMPs.^9^

Geographical differences were apparent when comparing MAF patterns by lineage and sampling location (Figure 4D, Supplementary Figures). Among Nichols-lineage strains, highly differentiated SNVs were common across the *tp0164* (*troB*, encoding an ATPase), *tp0462* (putative lipoprotein), and *tp0865* (*fadL*) genes with high frequencies in Oceanian and South American strains. In SS14-lineage strains, common non-synonymous SNVs included mutations in *tp0151* (*rnfD*)*, tp0515 (lptD*), and *tp0705* (*mrcA*). These mutations further segregated by *TPA* population assignment (Supplementary Figure 4). Asian SS14-lineage strains often had non-synonymous SNVs in *tp0705* (*mrcA,* encoding penicillin binding protein 1a), synonymous mutations in *tp0603* and *tp0416,* and mutations upstream of *tp0143.* Among SS14-lineage strains, the variant *mrcA* c.1873A>G was common in Europe, America and Oceania, and c.1517C>T was predominant in Asia. The variant c.1516G>A was common in Nichols-lineage strains.

The limited number of African *TPA* genomes sequenced to-date were distinct from the global population, with unique SNV signatures. We observed two high-frequency SNVs in African SS14-lineage strains, a non-synonymous mutation in *troR* (*tp0167*, allele frequency 97.5%), encoding a regulator of a transition metal uptake permease system, and synonymous mutation in the *tp0803* gene. Several SNVs were more common among African (Pop4 - Pop5) than global Nichols-lineage strains, including multiple non-synonymous SNVs in the *tp0483* and *tp0136* genes encoding putative fibronectin-binding proteins. All non-synonymous SNVs in *tp0483* worldwide were exclusive to Pop5. TP0136 p.Ser66Gly occurred at high frequency in Pop5, while p.Asp349Glu and p.Ser407Leu were exclusive to Pop4.

### Localization of amino acid substitutions in predicted protein models

To explore the potential impact of non-synonymous SNVs on protein structures, we localized amino acid changes to three-dimensional models of proteins encoded by genes affected by the most distinct mutations (Figure 5).^9^ In *lptD* (*tp0515*), nine fixed missense SNVs were present in all SS14-lineage strains and affected amino acids concentrated in its predicted C-terminal extension (Supplementary Table 5). Most non-synonymous, highly differentiated SNVs in the FadL proteins TP0865 and TP0858 were in predicted extracellular loops, with TP0858 mutations segregated by distinct *TPA* populations. Nearly all isolates from Nichols-lineage Pop2 had numerous mutations in TP0865, two distinct mutations in a TP0858 extracellular loop, and in the β-barrel and extracellular loop of OMF TP0966. These findings confirm *TPA* lineage- and population-specific genetic variation across global populations that can alter the structures of multiple proteins.

**Figure 5:**
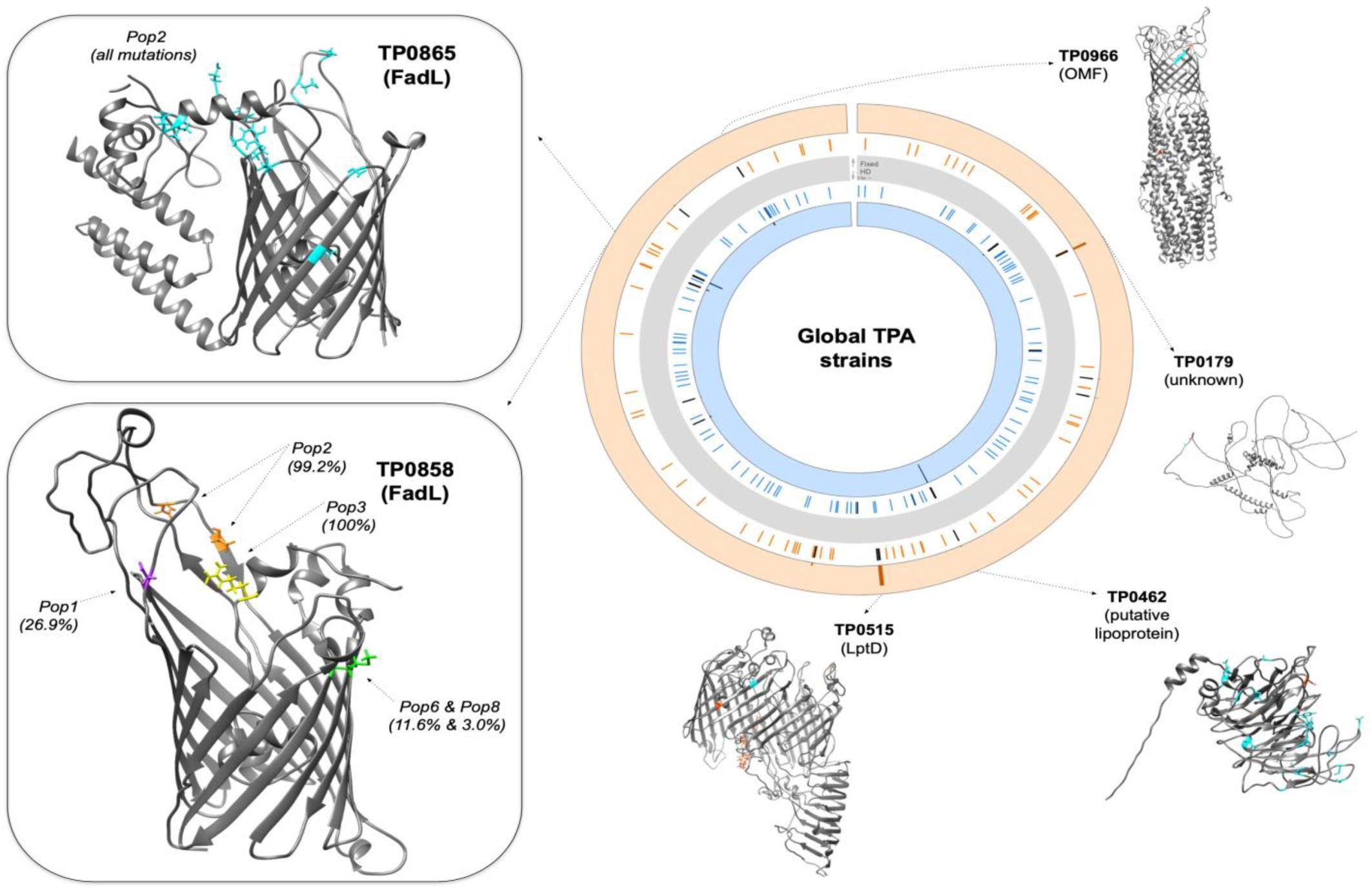
Accumulation of lineage- and population-associated missense mutations in select TPA proteins. Evaluation of predicted protein structures for genes with multiple lineage-informative missense SNVs confirms population-specific mutations across global TPA isolates. Genes with lineage-informative fixed (orange) and highly differentiated (HD; blue) SNVs identified during comparison of 1,413 TPA genomes are highlighted in the circular plot (excluding *tpr* family, *tp0470,* and *arp* genes). Starting from the gray bar: i) colored lines highlight genes affected by fixed (outer; orange) or highly differentiated (inner; blue) SNVs, with black lines highlighting genes with ≥2 distinct missense mutations, and ii) histograms depict the frequency of fixed (orange background) and highly differentiated (blue background) missense mutations by gene. Nichols-lineage three-dimensional structural protein models were previously predicted by Hawley *et al*. (TP0515, TP0858, TP0865, TP0966) or by *AlphaFold2* (TP0462, TP0179). Boxes highlight differences by TPA population for the FadLs TP0865 and TP0858; within-population allele frequencies >1% are annotated. Frequencies of lineage-informative fixed and highly informative SNVs are provided in Supplementary Tables 3-5. Genomic coordinates correspond to the Nichols reference strain. *TPA* = *Treponema pallidum* sub. *pallidum; SNV* = single nucleotide variant

## Discussion

Our consortium provided a unique infrastructure for assessing the clinical and genomic diversity of circulating *TPA* strains in the context of efforts to develop a syphilis vaccine with global efficacy. We enrolled racially/ethnically distinct populations with early syphilis, a fourth of whom were living with HIV, who provided samples that enabled analysis of genetically diverse SS14- and Nichols-lineage strains. Genomic analyses of participant samples, identification of SNVs across *TPA* populations, and mapping to protein models revealed variants of periplasmic proteins and OMPs that may influence protective immunity and spirochetal virulence.

We identified *TPA* in samples from PS and SS participants and found a range of qPCR results (Figure 2). Compared to the literature,^18–20^ we observed a higher proportion of lesion swabs and skin biopsies with detectable *TPA* (>80%), and high PCR positivity rates from skin scrapings and whole blood. Lesion swabs from genital ulcers and condyloma lata had the highest *TPA* qPCR values (ranging between 1,000-3,000 copies/µl among Malawi participants), confirming their suitability as specimens for syphilis detection.^21^

Our *TPA* WGS data support findings from the literature describing syphilis genetic epidemiology,^5–8^ while revealing several new findings. SS14-lineage strains predominated among participants recruited from clinical populations with different geographic, demographic, behavioral and clinical characteristics. Nichols-lineage strains had greater genetic divergence and clustered into more distinct subclades as previously observed.^5, 6, 8^ Interestingly, over half of participants infected by Nichols E subclade strains were living with HIV infection. These strains were distinct from HIV-associated BP1.3 *TPA* sublineage (SS14 lineage) described from Australia,^7^ suggesting spread within different networks. Consistent with prior reports of widespread macrolide-resistance,^22, 23^ most of our study sites had predominantly macrolide-resistant strains with the exception of Malawi. Although macrolides are no longer recommended for syphilis treatment, resistant strains remain highly prevalent suggesting that resistance may be driven by antibiotic use for other infections.^23^

In our large-scale genomic data analysis, we identified multiple lineage-informative mutations that differentiate SS14- and Nichols-lineage strains as well as *TPA* populations worldwide. Variation at the protein level was evident, with several *TPA* proteins affected by distinct mutations, including variation in extracellular loops of OMPs that may be driven by host immune pressure. Using new *TPA* genomes from our study and a large number of publicly available genomes, we observed eight distinct *TPA* populations consistent with the report by Leiberman, *et al*.^8^ Most of these are widely distributed, with two SS14-lineage populations (Pop7 and Pop8) consisting of sequences from Africa, Asia, Europe, North America, Oceania, and South America. Two Nichols-lineage populations (Pop4 and Pop5) were comprised almost entirely of sequences from Madagascar, Malawi and Zimbabwe.

We observed genetic differences by lineage and geography, cataloguing hundreds of fixed and significantly differentiated SNVs that distinguish SS14-versus Nichols-lineage strains. Among SS14-lineage strains from Africa, the *troR* c.314A>G (p.Glu105Gly) mutation was highly prevalent; *troR* is a member of the *tro* operon, encoding an ATP-binding cassette importer of transition metals in *TPA* and *T. denticola*.^24–26^ Non-synonymous mutations in genes encoding periplasmic proteins (*tp0151*) and OMPs (*tp0515, tp0858, tp0865* and *tp0966*) were highly differentiated between Nichols- and SS14-lineage strains. The *rnfD* (*tp0151*) gene product belongs to the cytoplasmic membrane Rnf complex involved in oxidation of flavodoxin and generation of a transmembrane ion gradient.^27^ The *tp0515* gene encodes an OMP orthologue of the LPS transport system (Lpt) of Gram-negative bacteria (LptABCDEFG).^9^ In *TPA*, LptE is replaced by a C-terminal extension to LptD in which we observed numerous fixed mutations. We also observed multiple non-synonymous SNVs in extracellular loops of the FadLs TP0858 and TP0865 that segregated by lineage and geography. The FadL OMPs are presumably involved in importing long-chain fatty acids and are important targets for syphilis vaccine development.^9^ Our findings emphasize the importance of studying genetic variation of these and other candidate *TPA* proteins in diverse populations.

We also observed differences by geography in three non-synonymous variants in the *tp0705* (*mrcA*) gene coding for penicillin-binding protein 1a. Variations in the *TPA mrcA* gene have been previously identified.^7, 22, 28^ Their biological function and clinical relevance are unclear given that penicillin remains highly effective for syphilis and confirmed penicillin resistance has never been reported.

While our study provides an important source of *TPA* clinical and genomic data from underrepresented areas, several limitations should be noted. First, we chose to enroll participants from four sites resulting in small sample sizes at each location. Thus, we could not evaluate *TPA* strains in the context of sexual networks. Furthermore, there were differences in recruitment strategies, resulting in dissimilar participant populations in each country. Our data are therefore clustered, limiting our ability to compare and draw larger conclusions based on participant characteristics. However, inclusion of diverse study participants improved the generalizability of our findings and was supported by genomic data confirming sampling of globally dominant *TPA* populations. Second, we excluded several recombinant loci demonstrated to be under positive and purifying selection from our analysis;^29^ novel methods for resolving these loci are greatly needed.^30^ Finally, our *TPA* genomic analysis focused on lineage-informative variants and does not include OMP mutations subject to recombination, which are relevant to vaccine design (*e.g., TPA* repeat proteins). Additional investigations should be conducted to explore the full array of *TPA* protein variants and their impacts on vaccine efficacy and *TPA* biology.

Our multi-center study delineated clinical and genomic features of contemporary SS14- and Nichols-lineage *TPA* infection among different populations. Although our data contributed to the number of available *TPA* genomes from Africa and South America, more sampling from these regions is needed to define local strain diversity. We observed an array of manifestations that did not appear to differ by lineage among our participants with syphilis, although genomic analysis confirmed lineage-informative mutations that varied by *TPA* population. Several of these mutations localized to OMP extracellular loops and segregated by geography, confirming the importance of representative sampling across global sites. Further comprehensive analyses of *TPA* OMP variability within circulating strains worldwide will be essential for syphilis vaccine development.

## Data Availability

The study protocol and English case report forms are included in the Supplementary Methods. The deidentified participant REDCap database will be made publicly available at the time of publication. Requests to share the informed consent forms and database can be made by emailing the corresponding author at. Raw sequencing data from this study with residual human reads removed are available through the Sequence Read Archive (SRA, BioProject PRJNA815321). Data supporting the findings of this study are available within the manuscript and Supplementary material.

## Data sharing

The study protocol and English case report forms are included in the Supplementary Methods. The deidentified participant REDCap database will be made publicly available at the time of peer-reviewed journal publication. Requests to share the informed consent forms and database can be made by emailing the corresponding author at. Raw sequencing data from this study with residual human reads removed are available through the Sequence Read Archive (SRA, BioProject PRJNA815321). Data supporting the findings of this study are available within the manuscript and Supplementary material.

## Acknowledgments

This project was funded by the US National Institutes of Health National Institute for Allergy and Infectious Disease (U19AI144177 to JDR and MAM). This work also was supported, in part, by the Bill & Melinda Gates Foundation (INV-036560 to ACS), strategic research dollars from Connecticut Children’s, the National Institute of Virology and Bacteriology (Programme EXCELES, ID Project No. LX22NPO5103, Funded by the European Union - Next Generation EU) to DS. Under the grant conditions of the Foundation, a Creative Commons Attribution 4.0 Generic License has already been assigned to the Author Accepted Manuscript version that might arise from this submission. JAGL was supported by an International Master’s Fellowship of the Wellcome Trust (reference number: 214641/Z/18/Z) and a Global Infectious Diseases Research Training grant of the Fogarty International Center (award number D43TW006589). Finally, the collection of samples of the other longitudinal study in Colombia that were included in the genomic analysis of *TPA* was supported by funds provided by Connecticut Children’s.

The authors would like to thank all of the study participants and research staff in Guangzhou, Cali, Lilongwe, and Chapel Hill. They would like to thank Myron Cohen for his support and guidance as a prior member of the Scientific Review Committee for the grant. They would also like to thank Julia Sung for contributing to participant enrollment; Sebastian Silva and Carol Ospina for their support in sample collection and management; Santiago Camacho, Maria Fernanda Amortegui and Nelson Romero for their work enrolling participants in the study; Andreea Waltmann and Fredrick Nindo for early contributions to genomic sequencing methods development and data analysis pipelines, respectively; Julie Nelson for supporting Malawi sample processing; Nancy Saravia for her guidance; and Julie Vigil for her clinical site regulatory oversight and monitoring. They would also like to thank the clinical and administrative staff of private and public health institutions in the Cali network, who referred potential participants to be included in this study.

Initial analysis of *TPA* genomes from 16 individuals enrolled in this study was included in a multi-institution NIH Cooperative Research Centers for Sexually Transmitted Infection manuscript as previously described.^30^ Protein structure visualization was performed using UCSF *Chimera*, developed by the Resource for Biocomputing, Visualization, and Informatics at the University of California, San Francisco, with support from NIH P41-GM103311.

## References

1. Ghanem KG, Ram S, Rice PA. The modern epidemic of syphilis. N Engl J Med 2020; 382(9): 845–54.

2. Organization WH. Web Annex 1. Key data at a glance. In: Global progress report on HIV, viral hepatitis and sexually transmitted infections, 2021. Accountability for the global health sector strategies 2016–2021: actions for impact.; 2021.

3. Gottlieb SL, Deal CD, Giersing B, et al. The global roadmap for advancing development of vaccines against sexually transmitted infections: Update and next steps. Vaccine 2016; 34(26): 2939–47.

4. Edmondson DG, Norris SJ. *In vitro* cultivation of the syphilis spirochete *Treponema pallidum*. Curr Protoc 2021; 1(2): e44.

5. Arora N, Schuenemann VJ, Jager G, et al. Origin of modern syphilis and emergence of a pandemic Treponema pallidum cluster. Nat Microbiol 2016; 2: 16245.

6. Beale MA, Marks M, Cole MJ, et al. Global phylogeny of Treponema pallidum lineages reveals recent expansion and spread of contemporary syphilis. Nat Microbiol 2021; 6(12): 1549–60.

7. Taouk ML, Taiaroa G, Pasricha S, et al. Characterisation of *Treponema pallidum* lineages within the contemporary syphilis outbreak in Australia: a genomic epidemiological analysis. The Lancet Microbe 2022.

8. Lieberman NAP, Lin MJ, Xie H, et al. Treponema pallidum genome sequencing from six continents reveals variability in vaccine candidate genes and dominance of Nichols clade strains in Madagascar. PLoS Negl Trop Dis 2021; 15(12): e0010063.

9. Hawley KL, Montezuma-Rusca JM, Delgado KN, et al. Structural modeling of the *Treponema pallidum* OMPeome: a roadmap for deconvolution of syphilis pathogenesis and development of a syphilis vaccine. J Bacteriol 2021; 203(15): e0008221.

10. Lukehart SA, Marra CM. Isolation and laboratory maintenance of *Treponema pallidum*. Curr Protoc Microbiol 2007; Chapter 12: Unit 12A 1.

11. Marfin AA, Liu H, Sutton MY, Steiner B, Pillay A, Markowitz LE. Amplification of the DNA polymerase I gene of *Treponema pallidum* from whole blood of persons with syphilis. Diagn Microbiol Infect Dis 2001; 40(4): 163–6.

12. Chen W, Smajs D, Hu Y, et al. Analysis of *Treponema pallidum* strains from China using improved methods for whole-genome sequencing from primary syphilis chancres. J Infect Dis 2021; 223(5): 848–53.

13. Delgado KN, Montezuma-Rusca JM, Orbe IC, et al. Extracellular loops of the *Treponema pallidum* FadL orthologs TP0856 and TP0858 elicit IgG antibodies and IgG(+)-specific B-cells in the rabbit model of experimental syphilis. mBio 2022; 13: e0163922.

14. Stamm LV, Bergen HL. A point mutation associated with bacterial macrolide resistance is present in both 23S rRNA genes of an erythromycin-resistant *Treponema pallidum* clinical isolate. Antimicrob Agents Chemother 2000; 44(3): 806–7.

15. Hunt M, Mather AE, Sanchez-Buso L, et al. ARIBA: rapid antimicrobial resistance genotyping directly from sequencing reads. Microb Genom 2017; 3(10): e000131.

16. Pettersen EF, Goddard TD, Huang CC, et al. UCSF Chimera--a visualization system for exploratory research and analysis. J Comput Chem 2004; 25(13): 1605–12.

17. Cruz AR, Ramirez LG, Zuluaga AV, et al. Immune evasion and recognition of the syphilis spirochete in blood and skin of secondary syphilis patients: two immunologically distinct compartments. PLoS Negl Trop Dis 2012; 6(7): e1717.

18. Grange PA, Gressier L, Dion PL, et al. Evaluation of a PCR test for detection of *Treponema pallidum i*n swabs and blood. J Clin Microbiol 2012; 50(3): 546–52.

19. Towns JM, Leslie DE, Denham I, et al. *Treponema pallidum* detection in lesion and non-lesion sites in men who have sex with men with early syphilis: a prospective, cross-sectional study. Lancet Infect Dis 2021; 21(9): 1324–31.

20. Theel ES, Katz SS, Pillay A. Molecular and direct detection tests for *Treponema pallidum* subspecies *pallidum*: a review of the literature, 1964–2017. Clin Infect Dis 2020; 71(Supplement_1): S4–S12.

21. Lukehart SA, Godornes C, Molini BJ, et al. Macrolide resistance in *Treponema pallidum* in the United States and Ireland. N Engl J Med 2004; 351(2): 154–8.

22. Beale MA, Marks M, Sahi SK, et al. Genomic epidemiology of syphilis reveals independent emergence of macrolide resistance across multiple circulating lineages. Nat Commun 2019; 10(1): 3255.

23. Machalek DA, Tao Y, Shilling H, et al. Prevalence of mutations associated with resistance to macrolides and fluoroquinolones in *Mycoplasma genitalium*: a systematic review and meta-analysis. Lancet Infect Dis 2020; 20(11): 1302–14.

24. Saraithong P, Goetting-Minesky MP, Durbin PM, Olson SW, Gherardini FC, Fenno JC. Roles of TroA and TroR in metalloregulated growth and gene expression in *Treponema denticola*. J Bacteriol 2020; 202(7).

25. Posey JE, Hardham JM, Norris SJ, Gherardini FC. Characterization of a manganese-dependent regulatory protein, TroR, from *Treponema pallidum*. Proc Natl Acad Sci U S A 1999; 96(19): 10887–92.

26. Desrosiers DC, Sun YC, Zaidi AA, Eggers CH, Cox DL, Radolf JD. The general transition metal (Tro) and Zn2+ (Znu) transporters in *Treponema pallidum*: analysis of metal specificities and expression profiles. Mol Microbiol 2007; 65(1): 137–52.

27. Radolf JD, Deka RK, Anand A, Smajs D, Norgard MV, Yang XF. *Treponema pallidum*, the syphilis spirochete: making a living as a stealth pathogen. Nat Rev Microbiol 2016.

28. Grillova L, Bawa T, Mikalova L, et al. Molecular characterization of *Treponema pallidum* subsp. *pallidum* in Switzerland and France with a new multilocus sequence typing scheme. PLoS One 2018; 13(7): e0200773.

29. Pla-Diaz M, Sanchez-Buso L, Giacani L, et al. Evolutionary processes in the emergence and recent spread of the syphilis agent, *Treponema pallidum*. Mol Biol Evol 2022; 39(1).

30. Lieberman NAP, Armstrong TD, Chung B, et al. High-throughput nanopore sequencing of *Treponema pallidum* tandem repeat genes arp and tp0470 reveals clade-specific patterns and recapitulates global whole genome phylogeny. Front Microbiol 2022; 13: 1007056.

