## Supplementary Material for "Clinical and genomic diversity of *Treponema pallidum subsp. pallidum:* A global, multi-center study of early syphilis to inform vaccine research"

**Seña AC, et al.**

**Table of Contents** **Page**

### Supplementary Methods

*Site-specific ethical review*

Each clinical site requested protocol review and approvals from their local institutional review board (IRB) prior to study recruitment and enrollment. In Guangzhou, China, participants were recruited from outpatient clinics of the Dermatology Hospital of the Southern Medical University (SMU; Medical Ethics Committee IRB protocol number GDDHLS-20181202 [R3]). In Cali, Colombia, participants were identified through a public health network of approximately 100 community health centers and referred if eligible to the Centro Internacional de Entrenamiento e Investigaciones Medicas (CIDEIM) in Colombia (CIDEIM Institutional Human Research Ethics Committee [IHREC] IRB 163, protocol number 1289). In Lilongwe, Malawi, participants were recruited from the Bwaila District Hospital sexually transmitted infection (STI) clinic and skin clinic in Malawi (National Health Sciences Research Committee Ministry of Health and 161 Population (IRB Approval Number 2252). In North Carolina, US, eligible participants were recruited as part of routine care at an infectious disease clinic in North Carolina (University of North Carolina [UNC] IRB Protocol Number 19-0311). Study participant enrollment was continued during the COVID-19 pandemic at each site based on local and country-specific policies.

*Study protocol*

| | |
|---|---|
| Title: | Global Sequence and Surface Antigenic Diversity of *Treponema pallidum* Outer Membrane Proteins |
| Study Cohort: | Approximately 1850 adults aged ≥18 years enrolled with untreated early (primary, secondary, or early latent) syphilis |
| Number of Sites: | Four |
| Subject Duration: | Two clinic visits, including baseline and follow-up 1-6 months after treatment (site specific duration). |

Objectives:

Primary:

- To establish a global clinical research consortium for syphilis vaccine development.
- To characterize *T. pallidum* (*TPA*) genomic sequences and catalog the global repertoire of *TPA* outer membrane proteins (OMPs) based on strains circulating in affected populations in consortium sites.
- To identify *TPA* OMPs and OMP extracellular loop epitopes that are recognized by sera and antigen-specific B-cells from syphilis patients in the global consortia sites.

Secondary:

- To develop a new classification system of TPA strains based on the variable outer membrane protein alleles.
- To estimate the impact of a vaccine candidate based on circulating strains in the global consortia sites.

Schematic of Overall Study Design:

Figure 1. Enrollment and Study Procedures

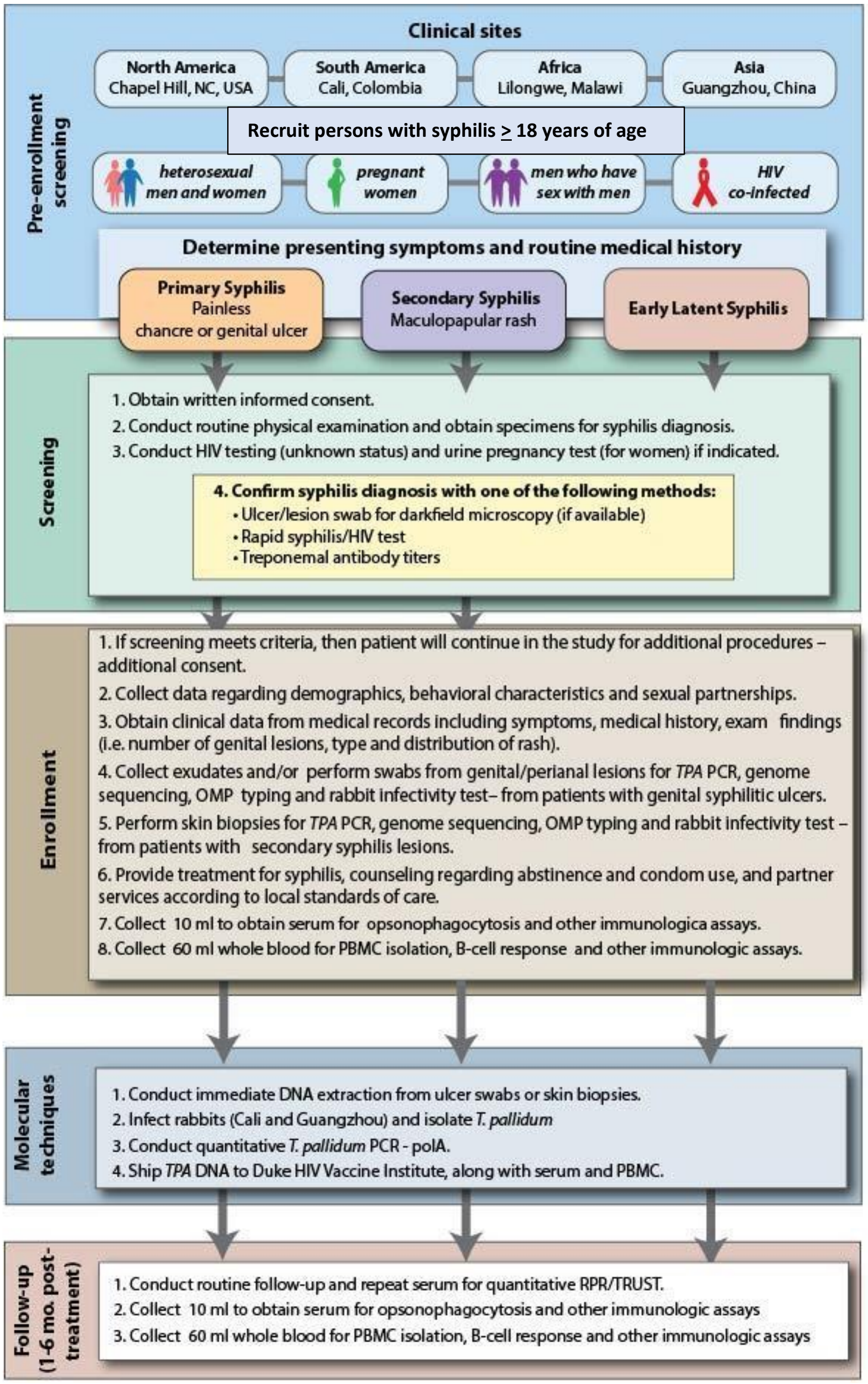

**1.** STUDY DESIGN

The study will be a longitudinal observational study of patients with primary, secondary, or early latent syphilis.

From our well-established syphilis network (Figure 2), we will recruit, screen, and enroll patients presenting with primary or SS (Figure 1). We will also enroll a smaller number of patients from the UNC and Cali sites with early latent syphilis. We will also enroll pregnant women with early syphilis at our Cali site. Our clinical sites, located in four different continents, will provide access to a diversity of patient populations with early syphilis, including heterosexual men and women, men who have sex with men, pregnant women, and HIV co-infected persons.

Figure 2: Global Syphilis Clinical Network

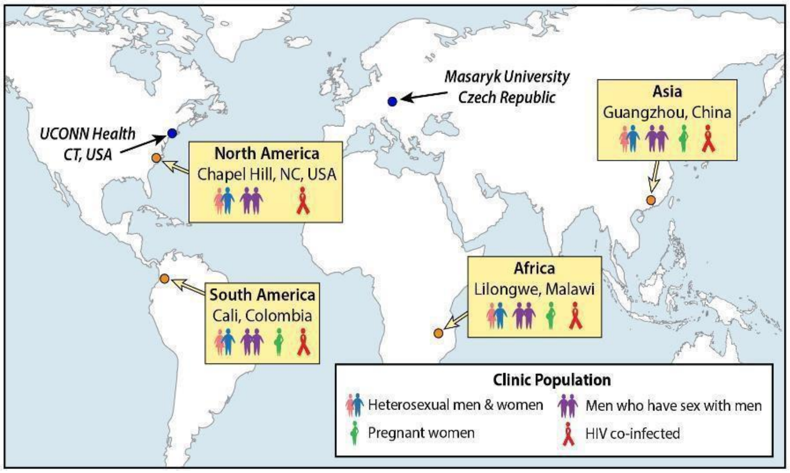

In order to participate, participants must be 18 years of age or older; be willing to provide informed consent; have signs or symptoms of primary or SS, or documented seroconversion within the previous 12 months, or sexual exposure to an individual known to have early syphilis diagnosed within the last 12 months. It is important to note that each of the four sites has slightly different capabilities for screening and testing patients for syphilis (See Study Procedures).

Following written consent, patients will undergo evaluation to confirm a diagnosis of early syphilis using direct detection and/or rapid treponemal Ab testing and stat non-treponemal antibody testing, (for patients with a known history of prior syphilis). Participants with unknown or prior HIV- negative status will undergo HIV testing. Additional STI testing will be conducted as per standard, country-specific care, and urine pregnancy testing will be performed in women with childbearing potential.

Study participants with initial confirmation of early syphilis diagnosis will be consented for additional study procedures including a detailed standardized questionnaire and specimen collection, including exudate collection and ulcer swabs from genital/perianal lesions, skin biopsies from patients with secondary syphilis, and serum and whole blood collection. All patients will receive syphilis therapy as per country specific standards of care. Patients enrolled in Cali and UNC will be asked to return for a follow up visit within 6 months after initial diagnosis. At the follow up visit, PBMC and serum samples will be obtained for Ab binding experiments and immunologic studies. If a patient is re-infected and clinically symptomatic with either primary or secondary syphilis, new samples will be obtained for TPA DNA analysis and OMP sequencing.

### 2. STUDY POPULATION

#### 2.1 Selection of the Study Population

Site 1: UNC Infectious Disease Clinic, Chapel Hill, North Carolina: This clinical site will recruit and enroll known HIV-infected patients who present to the walk-in clinic or their routine appointments with genital lesions or a rash suggestive of early syphilis, and patients with early latent syphilis.

Site 2: CIDEIM, Cali, Colombia: This clinical site will recruit and enroll early syphilis patients identified through the clinical network and referred for enrollment to the Clinical Research Unit at CIDEIM.

Site 3: UNC Project, Lilongwe, Malawi: This clinical site will recruit and enroll patients who present to the Bwaila Hospital STI Clinic with genital ulcer disease (GUD).

Site 4: Southern Medical University Dermatology Hospital, Guangzhou, China: This clinical site will recruit and enroll patients who present to the STI clinic with a presumptive diagnosis of primary or secondary syphilis.

### 2.2 Inclusion/Exclusion Criteria

Inclusion criteria for screening (based on site capability for routine syphilis testing):

1. Age 18 years of age or older;
2. Any of the following presentations:
   a) Suspected primary syphilis characterized by one or more painless ulcerative lesions (e.g. chancre); with or without a reactive non-treponemal or rapid treponemal antibody test.
   b) Suspected secondary syphilis characteristic mucocutaneous lesions (e.g., macular, maculopapular, papular, or pustular cutaneous lesions, mucous patches and/or condyloma lata ), with or without generalized lymphadenopathy AND with reactive non- treponemal AND treponemal antibody tests.
   c) Suspected early latent syphilis with no reported symptoms but documented seroconversion of syphilis serologies within the previous 12 months or documented sexual exposure to an individual known to have early syphilis diagnosed within the last 12 months; with reactive non-treponemal (i.e. RPR) titers >1:8 dilutions and treponemal antibody tests;
3. Willingness to have an HIV test if HIV status is unknown;
4. Willingness to have a rapid or urine pregnancy test for female subjects of child-bearing potential;
5. Willingness to provide informed consent for screening.

Inclusion criteria for enrollment (syphilis patients only):

1. Any of the following presentations:
   a) Suspected primary syphilis characterized by one or more painless ulcerative lesions (e.g. chancre); with or without a positive darkfield microscopy and/or reactive non-treponemal or rapid treponemal antibody test.
   b) Study subject has untreated secondary syphilis characterized by clinical findings as described above AND initial serologic confirmation defined by either a positive non- treponemal antibody test (i.e. RPR) titers >1:8 dilutions and/or a positive rapid treponemal antibody test.
   c) Suspected early latent syphilis with no reported symptoms but documented seroconversion of syphilis serologies within the previous 12 months or documented sexual exposure to an individual known to have early syphilis diagnosed within the last 12 months; with reactive non-treponemal (i.e. RPR) titers >1:8 dilutions and treponemal antibody tests;
2. Willingness to provide informed consent for additional study procedures;
3. Willingness to undergo ulcer swabs or skin biopsies for *T. pallidum* DNA extraction and genomics (for participants with suspected primary or secondary syphilis);
4. Willingness to provide serum and whole blood for peripheral blood mononuclear cells;

5. Study subject is willing to return to the clinic for study follow-up between 1-6 months following enrollment (UNC and Cali sites only).

Pregnant women, HIV-infected persons or patients who have been previously enrolled in the study will not be excluded. We will actively recruit participants who are suspected of reinfection with primary, secondary, or early latent syphilis. Exclusion criteria:

1. Patient is unwilling or unable to give informed consent;
2. Patient does not understand the purpose of the study or the nature of their participation;
3. Patient has received antibiotics active against TPA in the last 30 days.

### 3. STUDY PROCEDURES/EVALUATIONS

#### 3.1 Study Procedures

Screening: Potential participants with either signs/symptoms of early syphilis, history of a recent exposure to a syphilis infected sexual partner, and/or reactive syphilis screening test will present to the clinics for routine evaluation. This may involve a routine history, targeted physical examination, and initial evaluation for syphilis. Subjects with unknown HIV status or prior HIV- negative status will undergo HIV testing. Additional STI testing will be conducted and pregnancy testing will be performed in women of child-bearing potential.

If a patient is deemed to be potentially eligible, study staff will review the eligibility criteria and will obtain written informed consent before screening and additional study procedures. An ICF example is provided in Appendix A. Study specific procedures are those tests that are not routinely performed in the clinic as part of standard of care.

Enrollment: Upon initial confirmation of primary, secondary or early latent syphilis with at least one of the tests above at each clinical site, study participants will undergo additional informed consent for the following study procedures. All study related forms and specimens will be labeled with study identification numbers and no private identifiable information will be collected in this study.

1) Case report forms (CRFs, Appendix B) will be reviewed to collect additional demographic characteristics (i.e., age, sex/gender, race/ethnicity, marital status, education); behavioral characteristics (i.e., sexual orientation, age at first sexual debut,); sexual partnerships (i.e. number and gender of long-term and short-term sexual partners, number of sexual acts per week with long-term and short-term sexual partners, number of sex acts with use of condoms, number of concurrent sexual partners, duration of long-term and short-term partnerships, history of sexual contact with sex workers);

2) If the subject is pregnant, we will also collect obstetric characteristics (i.e., prenatal care, existing number of children, number of pregnancies and parity, and previous pregnancy loss).

3) Clinical data will be collected from the medical records and documented in CRFs, including syphilis symptoms, medical history (i.e. HIV status, history of syphilis or other STDs, history of viral hepatitis, history of drug use), physical examination findings (i.e. size, number, and location of genital ulcers lesions; type and distribution of rash, mucous patches, lymphadenopathy, condylomata lata), number of days from onset of symptoms to initiation of therapy, and treatment for syphilis.

4) Specimen collection for research purposes.

5) Treatment for syphilis, counseling regarding abstinence and condom use, and partner services will be provided according to standards of care at each clinical site.

Site specific procedures are outlined below. Each site will develop and follow procedures based on site specific algorithms that consider standard of care for syphilis management in their country.

Site 1 (UNC ID Clinic, Chapel Hill) –

1) Patient ≥ 18 years of age presents for routine HIV care with a characteristic genital lesion or rash suggestive of early syphilis, or a newly reactive RPR (>1:8 titer), or exposure to a sexual partner diagnosed with syphilis within last 12 months.
2) Refer patient to research staff for recruitment and study screening.
3) Research staff will obtain screening consent.
4) After screening consent, review eligibility criteria and complete screening CRF.
5) Perform stat RPR test and/or rapid treponemal test in patient without a history of prior syphilis (*Darkfield microscopy not available).
6) If the stat RPR (conducted for determination of eligibility) and/or rapid treponemal tests are positive, obtain additional consent for enrollment. (If negative, patient is not eligible. Provide screening incentive.)
7) Proceed with enrollment procedures: additional CRFs, study specimen collection (e.g. lesions swab, skin biopsy, whole blood, serum), urine pregnancy testing (if not already performed for routine care among women of childbearing potential), specimen processing.
8) Provide antibiotic treatment per CDC protocol for appropriate stage of syphilis.
9) Provide study incentive, and schedule study follow-up visit at 1-6 months.
10) Other routine care (e.g. counseling, partner services, condoms)

Site 2 (CIDEIM, Cali) –

1) Patient ≥ 18 years of age presents for routine care with a characteristic genital lesion or rash suggestive of early syphilis, or a newly reactive RPR (>1:8 titer) or positive rapid treponemal syphilis test, or recent exposure to a sexual partner diagnosed with syphilis within last 12 months.
2) Health care institution staff identify and refer patient to Clinical Research Unit at CIDEIM for the study.
3) CIDEIM staff will obtain screening consent for HIV, consent for enrollment and complete CRF.
4) HIV screening and rapid syphilis test will be performed.
5) Perform stat RPR and rapid treponemal test in patient without a history of prior syphilis, pregnancy test in women.
6) If genital ulcer is suggestive of primary syphilis, or any of the syphilis tests are positive, patient is eligible for additional procedures. (If negative, patient is not eligible. Provide screening incentive).
7) Proceed with enrollment procedures: additional CRFs, study specimen collection (e.g. dark- field microscopy, exudate collection, lesions swab, skin biopsy, whole blood, serum), specimen processing.
8) Provide study incentive, and schedule study follow-up visit at 1-6 months.
9) Provide antibiotic treatment per country specific protocol for appropriate stage of syphilis.
10) Other routine care (e.g. counseling, partner services, condoms, STI testing)

Site 3 (UNC Malawi, Lilongwe) –

1) Patient ≥ 18 years of age presents for routine care with a genital ulcerative lesion suggestive of primary syphilis.
2) Refer patient to research staff for recruitment and study screening.
3) Research staff will obtain screening consent.
4) After screening consent, review eligibility criteria and complete screening CRF.
5) Obtain ulcer swab and perform darkfield microscopy.
6) If darkfield microscopy is positive, obtain additional consent for enrollment. (If negative, patient is not eligible. Provide screening incentive).
7) Proceed with enrollment procedures: additional CRFs, RPR, treponemal antibody test in patient without a history of prior syphilis, HIV test (unless known HIV-positive), and pregnancy test in women, study specimen collection (e.g. lesions swab, serum), specimen processing.
8) Provide study incentive, and antibiotic treatment per country specific protocol for appropriate stage of syphilis.
9) Other routine care (e.g. counseling, partner services, condoms).

Site 4 (SMU Dermatology Hospital, Guangzhou) –

1) Patient ≥ 18 years of age presents for routine care to the STD clinic with a characteristic genital lesion or rash suggestive of early syphilis.
2) Perform darkfield microscopy, stat TRUST/RPR, rapid treponemal test in patient without a history of prior syphilis, HIV test (unless known HIV-positive), and pregnancy test in women.
3) If any of the syphilis tests are positive, refer patient to research staff for recruitment and study enrollment.
4) Research staff will obtain consent for enrollment.
5) After consent, review eligibility criteria and complete screening CRFs.
6) Proceed with enrollment procedures: additional CRFs, study specimen collection (e.g. lesions swab, whole blood, serum) and specimen processing.
7) Provide antibiotic treatment per country specific protocol for appropriate stage of syphilis.
8) Provide study incentive,
9) Other routine care (e.g. HIV care, partner services, counseling, condoms)

3.2 Clinical Evaluations (Post-screening)

Complete medical history will be obtained by a personal interview and questionnaire of each subject upon enrollment. A targeted physical examination (genital, rectal, oral, skin, and lymph node examinations) will be performed at each visit. All physical examinations will be performed by a qualified study clinician. Clinical photography will be performed from skin or mucosal lesions if patient consents.

Specimens to be collected specifically for research purposes from study subjects who meet criteria for early syphilis will include exudate collection and ulcer swabs from genital/perianal lesions and/or skin biopsies for *TPA* PCR and amplicon-based OMP sequencing. Study participants will undergo phlebotomy to collect 10 mL of whole blood to obtain serum for opsonophagocytosis assays and up to 60 mL of whole blood for peripheral blood mononuclear cells and immununology assays.

3.3 Laboratory Evaluations

3.3.1 Clinical Laboratory Evaluations

Diagnostic laboratory tests for syphilis, other STIs, HIV, CD4 count, hepatitis, and inflammatory markers (e.g. ESR, CRP) as indicated will be performed locally according to local protocols. Phlebotomy for serological testing and specimen storage to determine study outcomes will be performed locally. A pregnancy test will be performed on all subjects of childbearing potential.

*Specimen collection for research purposes*

Specimens collected from study participants who meet criteria for early syphilis and signed informed consent for the full study will include exudate swabs from genital/perianal lesions and/or biopsies from SS skin lesions for qPCR, genome sequencing, and OMP typing. Study participants will undergo phlebotomy to collect 10 mL of whole blood for serum for opsonophagocytosis assays and up to 60 mL of whole blood for PBMCs extraction and immunology assays.

*TPA DNA sample procurement and processing*

In addition to whole blood and serum samples, genital ulcer material (CIDEIM, Malawi, China) and skin biopsies (CIDEIM and UNC only) will be used to extract *TPA* DNA from patients with primary and SS. *TPA* DNA samples will be initially stored locally at each clinic, and shipped in batches to the University of North Carolina at Chapel Hill Genomics Core or the Duke Human Vaccine Institute (under the direction of Dr. Moody, U19 co-PI). Samples can then be transferred to Dr. Moody's immunology laboratory at Duke, the GGC at UNC, and the Salazar/Hawley laboratory in Connecticut for additional molecular and immunologic analyses (i.e. opsonophagocytosis). DNAs will be extracted from fresh genital ulcer material on site within 2 hours of collection.

##### 3.3.2 Laboratory Evaluations/Assays

Spirochetal burdens can be quantitated by qPCR using for *TPA polA*[17] by study site personnel, and in the case of the ID clinic in Chapel Hill and UNC Project Malawi, by the Genetics and Genomics Core. As previously stated above and in Table 3, each site will have slightly different screening procedures and in the case of Malawi and China, will enroll mostly primary syphilis cases, while in Cali and Chapel Hill, we will enroll a combination of primary, secondary, and early latent syphilis patients. A detailed description and site-specific enrollment is described in the Human Subjects section of this application.

##### 3.3.3 Special Assays or Procedures

*Rabbit infectivity testing (RIT)*

SMU (China) has the ability to isolate *TPA* from syphilis patients' blood and genital ulcer material, a technique known as rabbit infectivity testing. Cali has a new rabbit facility on site at CIDEIM. Procedures for rabbit injection and *TPA* isolation have been previously described. Approximately 20 live *TPA* isolates will also be isolated from a cohort of primary or secondary syphilis patients by RIT for whole genome sequencing (WGS).

*Genomic sequencing*

The Genetics and Genomics Core (GGC) will be housed primarily at UNC, with support from a second laboratory facility at Masaryk University (MU) in the Czech Republic. Each GGC laboratory will be responsible for sequencing of samples from different field collection sites. Specifically, UNC will process and sequence de-identified DNA from samples collected in North America, South America, Africa, and Asia. MU will process and sequence archived, de-identified DNA from samples previously collected in Europe. The GGC will employ next-generation sequencing techniques for whole-genome and/or targeted sequencing of known OMP genes. Sequencing data will be analyzed using established bioinformatics pipelines that permit assessment of minority variants in polyclonal infections. The GGC will also perform WGS on a subset of clinical isolates, including those successfully passaged in rabbits as described above. Sequences will be deposited in public repositories as described below (see *Publication Policy*).

### 4. STUDY SCHEDULE

We will recruit adult patients identified with early syphilis from four clinical sites located at the UNC- Chapel Hill Infectious Disease Clinic in Chapel Hill, North Carolina; CIDEIM in Cali, Colombia; Bwaila Hospital STI Clinic in Lilongwe, Malawi; and the Dermatology Hospital of SMU in Guangzhou, China. Heterosexual men and women, men who have sex with men, pregnant women, and HIV co-infected persons presenting for routine evaluation with symptoms suggestive of primary or secondary syphilis or suspected early latent syphilis will be recruited by study staff located at each clinical site. Recruitment will also be conducted through outreach activities and IRB-approved materials to be shared in person or via social media to promote syphilis screening among high-risk populations in the communities.

#### 4.1 Screening and Enrollment

The study procedures for each clinical site are outlined and discussed in Section 6, Study Procedures

- The potential study participant will be provided with a description of the study (purpose and study procedures) and asked to read the ICF or have it read to him/her. The ICF must be signed before any screening or study procedures are performed.
- Demographic information will be collected from the study participant.
- Eligibility criteria will be reviewed, and the following procedures will be conducted.

- Complete medical history will be obtained by interviewing subjects to assure eligibility.
- Sexual history for the past 60 days will be collected.
- A targeted physical examination will be performed by a qualified study clinician.
- If primary or secondary lesions are present, clinical staff may swab the lesions.
- A urine or serum pregnancy test will be performed on all subjects of childbearing potential.
- Blood will be collected for RPR test, storage, and HIV assays (including a CD4 if not available for HIV infected patients). Clinicians will provide pre- and post-test counseling, obtain written consent if required for HIV testing by local authorities, treatment, and referrals per local standard of care.

- Protocol requirements will be reviewed with the participant.

- Contact information will be collected, the preferred method of contact will be noted for any follow-up visit (s) at each site.

4.2 Follow-up and Final Visits, if applicable

To improve retention (UNC and Cali only), enrolled participants will be provided an appointment card for the follow-up visit to be scheduled after enrollment and to monitor response to syphilis therapy (defined as 3-6 months after treatment for early syphilis, or monthly for pregnant women with syphilis). Appointment reminders via telephone calls will also be provided to increase the likelihood of follow-up. Three separate attempts (e.g. via telephone calls) to contact participants for follow-up will be conducted by study staff. Participants will be considered lost-to-followup if there is no response or return to clinic after three attempts.

4.3 Criteria for Discontinuation or Withdrawal of a Subject (or a Cohort), if applicable

Study participants may voluntarily withdraw their consent for further study participation at any time and for any reason without penalty or prejudice to future medical care.

5. ASSESSMENT OF OUTCOME MEASURES

5.1 Specification of the Appropriate Outcome Measures

Significant advances have been made in the characterization of outer membrane proteins of TPA. However, developing a syphilis vaccine requires a better understanding of the OMPeome in different geographic areas and different transmission groups. The variety of TPA strains and intra-strain variability that has been observed in clinical isolates indicates that immune protection will require and effective immune response across multiple TPA strains.

5. 2 Primary Outcome Measures

Generate a catalog of the global repertoire of TPA outer membrane proteins, establishing the frequency of TPA clades and strains within each geographic area, as well as within specific transmission groups. Genome sequencing and OMP typing in clinical isolate samples will allow us to characterize the distribution and frequency of the alleles of current OMPs and may allow to identify new OMPs.

Structural analysis of OMPs identified in clinical isolates will be used to identify proposed extracellular loops which will then be used to evaluate serum reactivity and to identify epitope-specific B-cells. Epitope-specific B-cells will be utilized to generate monoclonal antibodies (MAbs) to be used in opsonization experiments. Epitope-specific MAbs found to have higher opsonization capacity will be considered to be directed against epitopes that will potentially have higher immunogenicity, and therefore could be used for immunogenicity experiments in animal models.

#### 5.3 Secondary Outcome Measures

Based on the frequency of the OMP alleles, recombination of alleles from different strains within clinical isolates, and the number of mutations within variable regions of the OMPs, a new classification system will be developed, which then can be used for local surveillance studies to make sure vaccine cocktails in specific locales have the correct representation of OMP variants.

Demographic and behavioral surveys of the study participants will be used to generate models of transmission dynamics and sexual partnership, which will be helpful to analyze the impact of vaccine candidates in different geographic areas and transmission groups.

*Case report forms*

Paper or electronic case report forms were used to document eligibility criteria for screening and enrollment, and review participants' demographics and sexual histories. In depth information regarding male or female sexual partnerships in the past year were also collected in this study to assist with future syphilis vaccine modeling. Clinical data were collected from participants and their medical records to include the following: history of sexually transmitted infections (STIs) and HIV; medical history and drug use; pregnancy status (from women); symptoms and physical examination findings; point-of-care and clinical laboratory testing; and research specimen collection. Data on other STI testing and repeat syphilis testing following treatment were collected if conducted as part of routine clinical care at each site. Any missing data or data anomalies were communicated to the sites for clarification and resolution. All queries were addressed locally and REDCap records were corrected when applicable.

Screening Eligibility Form

Inclusion Criteria: 0=No 1=Yes

*Participants must respond "Yes" to all site specific questions below to be eligible for screening.*

| |
|---|
| 1. Willingness to provide informed consent for screening *(all sites)*; |
| 2. Age 18 years of age or older *(all sites);* |
| 3. Any of the following:<br><br>a. Suspected primary syphilis characterized by one or more moist, ulcerative lesions (i.e. chancre) OR<br>b. Suspected secondary syphilis characterized by mucocutaneous lesions (e.g., rash – maculopapular, papular, or pustular lesions) with generalized lymphadenopathy, mucous patches and/or condyloma lata |
| 4. Willingness to undergo ulcer swab for darkfield microscopy |
| 5. Willingness to undergo finger prick for hemoglobin test *(for Lilongwe only)*; |
| 6. Patient has not received antibiotics active against syphilis in the last 30 days *(all sites: penicillin, azithromycin, doxycycline or syndromic GUD treatment)* |
| 7. Patient understands the purpose of the study or the nature of their participation *(all sites)*; |
| Study Eligibility Determination: *(all sites)* |
| 8. Is the patient eligible to participate?<br>*(responded "yes" to all inclusion criteria above)*: |
| 9. Patient has provided informed consent for screening procedures |

Enrollment Eligibility Form

Inclusion Criteria:

*Participants must respond "Yes" to 1-6, 7 (females only), and 8* 0=No 1=Yes

*in order to be eligible for enrollment*

| |
|---|
| 1. Willingness to provide informed consent for study procedures *(all sites);* |
| 2. Any of the following:<br>a. Patient has untreated primary syphilis based on clinical findings AND confirmation by darkfield microscopy, OR<br>b. Patient has untreated secondary syphilis based on clinical findings AND initial confirmation by stat non-treponemal antibody test and/or rapid syphilis test (if no prior history of syphilis) OR<br>c. Patient has untreated early latent syphilis based on documented seroconversion of syphilis serologies within past 12 months, or sexual exposure to an individual diagnosed with early syphilis within last 12 months |
| 3. Patient has a hemoglobin level ≥ 10.5 *(for Lilongwe only);* |
| 4. Willingness to undergo skin biopsies for *T. pallidum* genomics if patient has secondary syphilis |
| 5. Willingness to provide serum and whole blood for peripheral blood mononuclear cells |
| 6. Willingness to have an HIV test, if HIV status is unknown, uninfected, or not already performed; |
| 7. Willingness (for female subjects in childbearing age only) to have a urine pregnancy test if not already performed; |
| 8. Patient understands the purpose of the study or the nature of their participation; |
| Study Eligibility Determination: |
| 9. Is the patient eligible to participate?<br>*(responded "yes" to 1-8, (7 females only)* |
| 10. Patient has provided informed consent for enrollment procedures |

Demographics Form

| | |
|---|---|
| 1. Date of Birth | (DD-MMM-YY) |
| 2. Race/ethnicity | 1 = African American<br>2 = White<br>3 = Hispanic/Latino<br>4 = Asian<br>5 = Black African<br>6 = Hawaiian/Pacific Islander<br>7 = Native American<br>8 = Other, *specify*: |
| 3. Gender | 1 = *Cis*-gender male<br>2 = *Cis*-gender female<br>3 = *Trans*-gender male<br>4 = *Trans*-gender female<br>5 = Other, specify: |
| 4. Sexual orientation | 1 = Heterosexual<br>2 = Gay<br>3 = Bisexual<br>4 = Other, *specify:*<br>5 = Unknown *(for Malawi site)*<br>9 = Decline to answer |

| | | |
|---|---|---|
| 5. Marital status | 1 = Single<br>2 =Married/Cohabiting<br>3 = Divorced/separated/widowed | |
| 6. Educational status (*Highest completed*) | 1 = No formal school<br>2 = Elementary/Primary<br>3 = High school/Secondary<br>4 = Associate degree/Trade school | 5 = College/University<br>6 = Post-graduate<br>7 = Other, *specify*: |
| 7. Residence | 1 = Chapel Hill/Durham<br>2 = Cali<br>3 = Lilongwe<br>4 = Guangzhou | 5 = Other, North Carolina<br>6 = Other, Colombia<br>7 = Other, Malawi<br>8 = Other, China<br>*Specify town for #5-8:* |

Sexual History Form

1. Age of first sexual encounter *(includes oral, anal, penile/vaginal*)

2. Number of sexual partners in past 3 months

3. Number of *new* sexual partners in past 3 months

4. Number of sexual partners in the past year

5. Number of *new* sexual partners in the past year

*Loop through the next set of partner questions for the number of partners reported in the past year [Q4], up to a maximum of 5 partners.*

*Interviewer should use the following prompt, "You reported [Q4] number of sex partners in the past year. The following questions will ask about each of those partners, starting with your most recent."*

*For each set of questions in the loop, also use the following prompt: "The following questions are about partner [#]," or ask the subject to give the partner a nickname, an alias or initials, and then substitute that nickname/alias/initials in for every instance of "this partner" in a loop.*

1. Is this partner…
   1 = A spouse
   2 = A steady, long-term partner other than a spouse
   3 = A "one-night" stand or hookup
   4 = A casual partner with whom you have sex from time to time
   9 = Decline to answer

2. Have you *received* money/goods/favors to have sex with this partner?
   1 = Yes
   2 = No
   9 = Decline to answer

3. Have you *given* money/goods/favors to have sex with this partner?
   1 = Yes
   2 = No
   9 = Decline to answer

4. What is this partner's gender?
   1 = *Cis*-gender male
   2 = *Cis*-gender female
   3 = *Trans*-gender male
   4 = *Trans*-gender female
   9 = Decline to answer

5. How old is this partner?
   *(Estimated age)* 99=Decline to answer

6. What is this partner's race/ethnicity?
   1 = African American
   2 = White
   3 = Hispanic/Latino
   4 = Asian
   5 = Black African
   6 = Hawaiian/Pacific Islander

7 = Native American
8 = Other, *specify*:
9 = Decline to answer

7. What is this partner's HIV status?
1 = This partner told you he/she was HIV-negative
2 = You know this partner is HIV-positive.
3 = You are not completely sure of this partner's HIV status.
9 = Decline to answer

8. Has this partner had any symptoms of a sexually transmitted infection in the last year?
1 = Yes
2 = No (*skip Q9*)
3 = Don't know (*skip Q9*)
9 = Decline to answer (*skip Q9*)

9. What symptom did this partner have in the last year? *(mark all that apply)*
1 = Ulcer(s) or sore(s) on the genitals
2 = Ulcer(s) or sore(s) in the mouth
3 = Ulcer(s), sore(s) or bumps on the anus
4 = Rash on the body
5 = Discharge from genital area
6 = Pain or burning during urination *(peeing or emptying the bladder)*
7 = Other, *specify*:
9 = Decline to answer

10. Did this partner have syphilis in the last year?
1 = Yes
2 = No (*skip Q11*)
3 = Don't know (*skip Q11*)
9 = Decline to answer (*skip Q11*)

11. Did this partner receive treatment for syphilis?
1 = Yes, *specify type (e.g. injections, pills)*
2 = No
3 = Don't know
9 = Decline to answer

12. Has this partner had other sexual partners in the last year?
1 = Yes
2 = No
3 = Don't know
9 = Decline to answer

13. How long ago did you first have sex with this partner?
99 = Decline to answer

14. How long ago was your most recent sex with this partner?
99 = Decline to answer

15. Do you anticipate having sex with this partner again in the future?
1 = Yes
2 = No
3 = Don't know
9 = Decline to answer

FOR MALE subjects **(***For cis-gender males, and possibly trans-gender females depending on transition process)*

<u>If sexual partner is female:</u>

16. In an average month during which you were sexually active with this partner in the past year, how many times per month did you have insertive *vaginal* intercourse with this partner (*you inserted your penis in your partner's vagina)?*
99 = Decline to answer
*If 0, skip Q16a.*

16a. Of those times that you had insertive *vaginal* intercourse, how many times did you put your penis in your partner's vagina *without* a condom?
99 = Decline to Answer
*Error message if Q16>Q16a: The number of times without a condom cannot be greater than the total number of times reported for insertive vaginal intercourse.*

If sexual partner is female or male:

17. In an average month during which you were sexually active with this partner in the past year, how many times per month did you have insertive *anal* intercourse with this partner (*you inserted your penis in your partner's butt or anus)*?
99 = Decline to answer
*If 0, skip Q17a.*

17a. Of those times that you had insertive *anal* intercourse, how many times did you put your penis in your partner's rectum *(butt or anus) without* a condom?
99 = Decline to answer
*Error message if Q17>Q17a: The number of times without a condom cannot be greater than the total number of times reported for insertive anal intercourse.*

18. In an average month during which you were sexually active with this partner in the past year, how many times per month did you perform oral sex on this partner (*you put your mouth on your partner's genitals)*?
99 = Decline to answer

19. In an average month during which you were sexually active with this partner in the past year, how many times per month did this partner perform oral sex on you (*you inserted your penis in your partner's mouth*?
99 = Decline to answer

If sexual partner is male:

20. In an average month during which you were sexually active with this partner in the past year, how many times per month did you have *receptive anal* intercourse with this partner (*your partner inserted his penis in your butt or anus*)?
99 = Decline to answer
*If 0, skip Q20a.*

20a. Of those times that you had *receptive anal* intercourse, how many times did this partner put his penis in your rectum *without* a condom?
99 = Decline to Answer
*Error message if Q20>Q20a: The number of times without a condom cannot be greater than the total number of times reported for receptive anal intercourse.*

For FEMALE subjects **(***For cis-gender females, and possibly trans-gender males depending on transition process)*

If sexual partner is male:

21. In an average month during which you were sexually active with this partner in the past year, how many times per month did you have receptive *vaginal* intercourse with this partner (*your partner inserted his penis in your vagina)*?
99 = Decline to Answer
*If 0, skip Q21a.*

21a. Of those times that you had receptive *vaginal* intercourse, how many times did your partner put his penis in your vagina *without* a condom?
99 = Decline to Answer
*Error message if Q21>Q21a: The number of times without a condom cannot be greater than the total number of times reported for receptive vaginal intercourse.*

22. In an average month during which you were sexually active with this partner in the past year, how many times per month did you have receptive *anal* intercourse with this partner (*your partner inserted his penis in your butt or anus*)?
99 = Decline to Answer
*If 0, skip Q22a.*

22a. Of those times that you had receptive *anal* intercourse, how many times did this partner put his penis in your rectum *(butt or anus) without* a condom?
99 = Decline to Answer
*Error message if Q22>Q22a: The number of times without a condom cannot be greater than the total number of times reported for receptive anal intercourse.*

If sexual partner is male or female:

23. In an average month during which you were sexually active with this partner in the past year, how many times per month did you perform oral sex on this partner (*you put your mouth on your partner's genitals)*?
99 = Decline to Answer

24. In an average month during which you were sexually active with this partner in the past year, how many times per month did this partner perform oral sex on you (*he/she put his/her mouth on your vaginal area)*?
99 = Decline to Answer

STI and HIV History Form

1. Previous self-testing *(Self-test defined as you take your own sample and interpret the result yourself)*
   0 = No
   1 =Syphilis test, *specify result*:
   2 = HIV test, *specify result*:
   9 = Unknown

For ALL subjects: *Responses to the following questions can be from obtained from the patient. However, review of medical records should be conducted for additional documentation.*

2. Current syphilis diagnosis
   1 = Primary syphilis
   2 = Secondary syphilis
   3 = Early latent syphilis
   4 = Other, *specify:*

3. Prior history of syphilis or GUD
   0 = No (*skip to Q8*)
   1 = Yes
   9 = Unknown

4. Date of *last* syphilis diagnosis (*DD-MMM-YY*)
   99 = Unknown

5. Prior syphilis diagnoses (*mark all that apply*)
   1 = Primary syphilis
   2 = Secondary syphilis
   3 = Early latent syphilis
   4 = Late latent or unknown duration
   5 = Neurosyphilis
   6 = Other, tertiary
   7 = Other, *specify*:
   9 = Unknown

6. Date of *last* syphilis treatment, including syndromic treatment for GUD (*DD-MMM-YY*)
   99 = Unknown

7. *Last* treatment for syphilis (or as part of GUD treatment)
   1 = Benzathine penicillin 2.4mu intramuscular injections (IM) x 1
   2 = Benzathine penicillin 2.4mu IM weekly x 2 weeks
   3 = Benzathine penicillin 2.4mu IM weekly x 3 weeks
   4 = Doxycycline 100mg orally twice daily (BID) x 14 days
   5 = Doxycycline 100mg orally twice daily (BID) x 28 days
   6 = Other, *specify*
   9 = Unknown

8. Serologic response to *last* syphilis treatment (*Based on RPR/VDRL/TRUST titer*)
   1 = At least 4-fold decline
   2 = Less than 4-fold decline
   3 = Serofast
   4 = Seroreversion
   5 = Other, *specify:*
   9 = Unknown

9. Clinical response to *last* syphilis treatment
   1 = Yes
   2 = No
   3 = Other, *specify*:
   9 = Unknown

10. History of other STIs (*mark all that apply*)
   0 = No prior STIs
   1 = Chlamydia
   2 = Gonorrhea
   3 = Trichomonas
   4 =Genital herpes
   5 = Genital warts
   6 = Genital ulcer disease
   7 = Vaginitis
   8 = Urethritis
   9 =Other, specify:

11. HIV-infected
0 = No; *If "No", end of form*
1 = Yes
9 = Unknown

12. Date of HIV diagnosis (*MMM-YYYY*)
99 = Unknown

13. Last CD4 count (*cells/uL*)
9999 = Unknown

13a. Date of CD4 test: (*DD-MMM-YY*)
99 = Unknown

14. Last HIV viral load (*copies/ml)*
000 = Undetectable
999 = Unknown

14a. Date of HIV viral load test (*DD-MMM-YY)*
99 = Unknown

15. Currently on antiretroviral therapy 0 = No 1 = Yes 9 = Unknown

Medical History and Drug Use Form

1. History of chronic hepatitis B 0 = No 1 = Yes 9 = Unknown

2. History of chronic hepatitis C 0 = No 1 = Yes 9 = Unknown

3. History of other immunodeficiency 0 = No 1=Yes 9 = Unknown
(*Other than HIV, if "Yes", specify):*

4. Prior intravenous drug use 0 = No 1 = Yes
(*If "No," skip to #7)*

5. Last injection drug use
1 = Within past year
2 = Greater than a year ago
9 = Unknown

6. Type of injection drugs used: (*mark all that apply*)
1 = Cocaine 5 = Methamphetamine
2 = Crack 6 = Stimulants
3 = Heroin 7 = Other, *specify:*
4 = Opiates 9 = Unknown

7. Other current drug use: (*mark all that apply*)
0 = No drug use
1 = Alcohol
2 = Marijuana
3 = Cocaine/crack
4 = Narcotics/Opioids
5 = Methamphetamine
6 = Stimulants
7 = Nitrates/Poppers
8 = Other, *specify*:

Current Pregnancy Form: For ALL Females

1. Currently pregnant, as determined by pregnancy test
0 = No (*skip to #4*)
1 = Yes

2. Gestational age
1 = <13 weeks (first trimester)
2 =14-27 weeks (second trimester)
3 = 28-40 weeks (third trimester)

9 = Unknown or not applicable

3. Prenatal care — 0 = No
1=Yes
9 =Unknown or not applicable

4. Number of prior pregnancies (*not including this pregnancy, if currently pregnant*);
*If '00' then end of form.*

5. Number of live births

6. History of stillbirths — 0 = No
1 = Yes
9 = Unknown

7. History of premature births — 0 = No
1 = Yes
9 = Unknown

Symptoms and Physical Exam Form

1. Presence of ulcer (chancre) at sites of sexual exposure?
*If "No" or "Unknown" skip to #2 and mark 1a and 1b as not applicable*

0 = No 1 = Yes 9 = Unknown

1a. Number of days since ulcer first appeared
999 = Unknown or not applicable

2. Presence of rash — 0 = No 1 = Yes *(Obtain photographs of rash)*
*If "No" skip to #2c and mark 2a and 2b as not applicable*

2a. Type of rash — 1 = Macular 4 = Papulo-squamous
2 = Papular 5 = Other, *specify:*
3 = Maculo-papular 9 = Not applicable

2b. Number of days since rash first appeared
999 = Unknown or not applicable

2c. Presence or absence of rash: 0=Absent 1=Present 9=Not applicable

| Location | Presence |
|---|---|
| Face and head | |
| Trunk | |
| Genital region | |
| Palms or soles | |
| Extremities | |
| Other, *specify:* | |

3. Other signs of secondary syphilis: 0=Absent 1=Present 9=Not applicable

| Sign | Presence |
|---|---|
| Alopecia | |
| Mucous patch | |
| Condyloma lata | |
| Lymphadenopathy | |
| Other, *specify:* | |

| |
|---|
| 4. No current signs or symptoms |

Point-of-Care Test Form

| | | |
|---|---|---|
| **1.** Hemoglobin Result *(for Lilongwe only)* | | |
| 2. Pregnancy test (*for females only*) | 0 = Negative<br>1 = Positive | 2 = Equivocal<br>9 = Not applicable |
| 3. Rapid HIV test | 0 = Negative<br>1 = Positive | 2 = Equivocal<br>9 = Not performed |
| 3a. Name of rapid HIV test: | | |
| 4. Rapid syphilis test | 0 = Negative<br>1 = Positive | 2 = Equivocal<br>9 = Not performed |
| 4a. Name of rapid syphilis test: | | |

Clinical Laboratory Specimen Form

| | | |
|---|---|---|
| 1. Darkfield microscopy | 0 = Negative<br>1=Positive | 2 = Equivocal<br>9 = Not applicable |
| 2. Nontreponemal antibody test | 0 = Negative<br>1 = Reactive<br>9 = Not performed | |
| 2a. Type of test | 1 = RPR<br>2 = VDRL<br>3 = T RUST | |
| 3. Treponemal antibody test | 0 = Negative<br>1 = Positive | 2 = Equivocal |
| 3a. Name of treponemal test: | | |
| 4. HIV-1 ELISA or antibody test | 0 = Negative<br>1 = Reactive | 2 = Equivocal<br>9 =Not applicable |
| 4a. Type of HIV test: | | |
| 5. HIV Western blot or PCR | 0 = Negative<br>1= Positive | 2 = Equivocal<br>9 = Not applicable |
| 6. Gonorrhea test<br>*(Oral, genital, rectal)* | 0 = Detected<br>1 = Not detected | 9 = Not performed |
| 6a. Type of gonorrhea test: | | |
| 7. Chlamydia test<br>*(Oral, genital, rectal)* | 0 = Detected<br>1 = Not detected | 9 = Not performed |
| 7a. Type of chlamydia test: | | |
| 8. Herpes simplex virus (HSV) test | 0 = Negative<br>1 = Reactive | 9 = Not performed |

8a. Type of herpes test:

9. Other STD or viral hepatitis test — 0 = Negative; 1 = Reactive; 9 = Not performed

9a. Name of other test (s):

Research Specimen Collection Form

1. Genital ulcer swabs for PCR (*for Cali, Lilongwe, Guangzhou*);
   1 = Collected
   2 = Not collected
   9 = Not applicable

1a. Number of ulcers swabbed for PCR testing
   9 = Not applicable

**2.** Skin biopsies for PCR (*for Chapel Hill, Cali, Guangzhou*)
   1 = Collected
   2 = Not collected
   9 = Not applicable

2a. Number of biopsies collected for PCR testing
   9 = Not applicable

2b. Location of biopsies:
   1 = Chest
   2 = Back
   3 = Extremities
   4 = Other, *specify*

**3.** Mucous ulcer and/or exudate for rabbit infectivity testing
   1 = Collected
   2 = Not collected
   9 = Not applicable

3b. Number of exudates collected for rabbit infectivity testing

4. Whole blood to obtain serum for opsonophagocytosis
   1 = Collected
   2 = Not collected

4a. Specimen volume (ml) — 999= Not applicable

5. Whole blood for PBMCs
   1 = Collected
   2 = Not collected

5a. Specimen volume (ml) — 999 = Not applicable

Study Follow-up Form

1. Months after study enrollment

2. Syphilis treatment provided (*at enrollment visit*)
   1 = Benzathine penicillin 2.4 mu IM x 1
   2 = Benzathine penicillin 2.4 mu IM x 3
   3 = Doxycycline 100mg po BID x 14 days
   4 = Doxycycline 100mg po BID x 28 days
   5 = Other, *specify:*

3. Nontreponemal antibody test — 0 = Negative; 1 = Reactive

| | |
|---|---|
| 3a. Type of test | 1 = RPR<br>2 = VDRL<br>3 = TRUST |
| 3b. Titer (*at follow-up visit*) | |
| 4. Serologic response to syphilis treatment (*Based on RPR/VDRL/TRUST titer*) | 1 = At least 4-fold decline<br>2 = Less than 4-fold decline<br>3 = Serofast<br>4 = Seroreversion<br>5 = Other, *specify:* |
| 5. Clinical response to syphilis treatment | 1 = Yes<br>2 = No<br>3 = Other, *specify*: |
| 6. Whole blood to obtain PBMCs | 1 = Collected<br>2 = Not collected |
| 6a. Specimen volume (*ml)* | 999 = Not applicable |

*DNA extraction and T. pallidum isolation from rabbit infectivity testing*

*Treponema pallidum* subsp. *pallidum* (*TPA*) DNA was extracted from lesion swabs, skin biopsies, and 2 ml of whole blood, using the QIAmp DNA Blood Mini Kit (Qiagen Inc; Venlo, Netherlands) according to a standardized protocol across all sites. DNA was stored at −80°C until further testing.

At the SMU site in China, rabbit experimentation was approved by the Ethics Committee of SMU Experimental Animal Ethics Committee (L2018277) and South China Agricultural University Experimental Animal Ethics Committee (2020C004). Exudates from lesion swabs from participants with PS and 1ml of whole blood from participants with SS were used to immediately inoculate New Zealand White rabbits in their each testicle. The animals were monitored for a maximum of 3 months to look for the presence of seroconversion. *TPA* isolation was performed from rabbit testicles after *TPA* specific antibody titer raised to 1:1280, and isolates subjected to serial passage before DNA extraction using the DNeasy Blood & Tissue Kit (QIAGEN, Hilden, Germany; 69506) which usually takes no more than three rabbits. Animals that did not seroconvert underwent a blind pass and only two rabbits were observed consecutively in this procedure. If seroconversion occurred in the second rabbit, the passage was continued, and the procedure was terminated while no seroconversion occurred.

*TPA quantitative PCR*

*TPA* PCR was conducted on samples collected from participants enrolled in the primary study; samples previously collected as part of four different studies in Colombia were also sequenced (CIDEIM IRB reference numbers 1281, 1289, 1315). Spirochetal burdens were analyzed using quantitative real-time polymerase chain reaction (qPCR) targeting the *TPA* DNA polymerase I (*polA; tp0105*) gene from lesion swabs, skin biopsies or scrapings, and whole blood as previously described,[1] using a standardized protocol across sites. *TPA* qPCR was performed at CIDEIM and SMU before DNA shipment to the UNC Genomics and Genetics Core; UNC Project-Malawi and UNC DNA samples underwent qPCR at the UNC Genetics and Genomics Core. All samples had repeat qPCR performed immediately before *TPA* WGS. From Colombia and Malawi, the first lesion swab for darkfield microscopy (DFM) underwent *TPA* PCR analyses; in China, the second lesion swab was used for PCR since the first specimen was prioritized for rabbit infectivity testing. Each PCR run included negative (no template) controls. *PolA* copy numbers were extrapolated from a standard curve generated using 10-fold serial dilutions ($10^6$

to $10^1$ copies) of purified plasmid DNA containing the *polA* amplicon cloned into pCR2·1-TOPO vector (Invitrogen; Waltham, Massachusetts, US) and shared across sites. The positivity threshold was established at >0 copies/µL.

For participants with multiple *TPA* qPCR results per sample type, overall *TPA* qPCR sample results were classified as follows: if at least one individual sample result was >0 copies/µl, the overall sample type result was classified as positive with an overall quantitative result as the geometric mean of all >0 copies/µl results. Otherwise, the overall sample result was classified as negative (0 copies/µl). Only results from the local laboratories were included in this determination, with the exception of Malawi, which had all *TPA* PCR labs performed at UNC. Genomic data analyses are described above.

*TPA enrichment, library preparation, and sequencing*

Determination of *TPA polA* copy number and total DNA concentration was conducted on samples by qPCR and Qubit fluorometer with dsDNA HS2 reagents (Thermo Fischer Scientific, Waltham, MA, USA), respectively. Specimens with ≥40 *polA* copies/µL were selected for WGS, with the exception of two low concentration samples from China. For participants with multiple specimens, we selected one sample for inclusion in the study by prioritizing DNA samples that had been extracted directly from chancre swabs or skin biopsies (without rabbit passage) when available and had the highest *polA* copy number.

*TPA* enrichment and WGS were conducted as previously described,[1] with several alterations. In brief, samples with total DNA concentrations <2ng/µL (i.e. <10ng DNA in 50µL) were first subjected to parallel, pooled whole-genome amplification (ppWGA) with at least three replicates using random hexamer primers and *phi29* polymerase (Genomiphi v2, Cytiva, Marlborough, MA, USA). Amplified products were pooled and cleaned up using 1·8x AMPure XP beads (Beckman Coulter, Brea, CA, USA) as previously described.[2] DNA or cleaned-up ppWGA product was then acoustically sheered and enriched for TPA DNA using Sure Select XT Low Input (Agilent Technologies, Santa Clara, CA) or Sure Select XTHS2 custom 120-nucleotide RNA oligonucleotide baits at UNC or SMU, respectively, according to manufacturer instructions. Efforts were made to normalize capture efficiency and sequencing depth of all samples. Samples were pooled for hybrid capture by relative TPA input (TPA:total DNA ratio). Rabbit-passaged samples were pooled separately from directly extracted samples. Capture pools consisted of up to 40 libraries per capture reaction. These pools were combined and sequenced using the MiSeq platform (Illumina, San Diego, CA, USA) at UNC or NovaSeq platform at SMU with paired-end, 150bp reads. Raw sequencing data after removal residual human reads are available through the Sequence Read Archive (SRA, BioProject PRJNA815321).

*Sequencing data processing and phylogenomic analysis*

Sequencing data, along with publicly available data from geographically diverse locations published by Lieberman *et al.*,[3] was processed, aligned, and analyzed using an adaptation of our previously described bioinformatic pipeline,[2] as described at https://github.com/IDEELResearch/Tpallidum_genomics. In brief, adapter sequences were trimmed using *trimmomatic* (v0·36), and trimmed reads analyzed for contaminants using *strainseeker* (v1·5·1); aligned to the SS14 (accession CP004011.1) and Nichols (accession CP004010.2) reference genomes using *bwa* (v0·7·17); filtered using *samtools* (v1·16), *picard* (v2.26.11), and custom shell scripts; and variant called using *GATK*'s (v3·8-0) *haplotypecaller* utility. Downstream analyses were conducted using a consensus genome FASTA file constructed based on single nucleotide variant (SNVs); only loci with ≥3 unique mapping reads were called. Consensus genomes for SS14- and Nichols-like strains were derived from the SS14- and Nichols-reference-based alignments, respectively. Genomes were aligned using *MAFFT* (v7.490), including well-characterized reference genomes for TPA (Nichols, SS14, Mexico A), *T. pallidum* subsp. *pertenue* (TPE, Samoa D), and *T. pallidum* subp. *endemicum* (TEN, Bosnia A). Repetitive and putative recombination regions were identified and masked using *Gubbins* (v3·2) before construction of a maximum likelihood (ML) tree using *RAxML* (v8·2·12), performed with 1,000 rapid bootstraps. Trees were visualized and annotated in R (v4·1·2) using the *ggtree* package (v3·2·1). Clades and subclades were assigned by visual inspection and comparison to those defined by Lieberman *et al*.[3] We identified macrolide-resistant strains using *ARIBA*[4] (v2·14·6) applied to trimmed and host-removed TPA sequences and a customized reference database based on the Nichols and SS14 coordinate system to identify 23S rRNA A2058G and A2059G variants.

*Analysis of global TPA genomic population structure*

A separate analysis pipeline was used to align, call, and analyze these and all publicly available *TPA* genomes as described at https://github.com/IDEELResearch/Tpallidum_genomics. Following adapter trimming using *trimmomatic* (v0.39), we assessed sequences for traces of host genome using *bbmap* (v38·82) by mapping reads against human (hg19) and rabbit (oryCun2) reference genomes and removing reads with ≥2 hits against the reference genomes. Broken reads were removed using repaired.sh embedded in *bbmap*. We mapped quality-filtered reads to a modified version of Nichols (accession CP004010.2, with 23S *rRNA*, *tpr*, *arp* and *tp0470* genes masked using *BEDTools* (v2·30)) using *bwa* (v0.7.17). Sequence alignments were subjected to post-alignment filtering, including removing duplicate reads, indel realignment, removing reads with excessive mismatches, soft and hard clips, chimeric alignment, repetitively aligned reads and low mapping quality reads. Following post-alignment filtering, genomic sequences with at ≥3 unique reads mapping across >80% of the genome were retained for analysis. Variant calling was performed using *GATK* (v3·8·1).

We evaluated TPA population structure using several approaches. First, we performed principal component analysis (PCA) using the *smartpca* package in EIGENSOFT (v6·1·4), including only biallelic single nucleotide variant (SNV) sites with <20% missingness and >1% minor allele frequency (MAF) across the global TPA gene pool. PCAs were visualized and annotated using *ggplot2* (v3·3·6) in R. In order to assign SS14 versus Nichols clade membership, a maximum likelihood phylogeny tree of clinical isolates alongside Nichols (CP004010·2) and SS14 (CP004011·1) reference genomes was constructed using IQ-tree (v1·6·12)[5] and clades were assigned according to clustering with the corresponding reference genomes. Next, we employed *fastbaps* (v1·0·8) to identify clades and subclades using a Bayesian approach.[6] Fisher's exact test was used to identify significantly differentiated SNVs between SS14 and Nichols clades using a Bonferonni-corrected p value of $< 4.451 \times 10^{-6}$ to account for multiple comparisons. For all SNVs with >1% MAF, we compared allele frequencies by clade and continent using a heatmap generated using *ggplot2* in R. We used *snpEff* (v5·1) for functional annotation estimation of significant SNVs. SNVs then were mapped to 3-dimensional protein models previously predicted by Hawley *et al*.[7] (TP0515, TP0858, TP0865, TP0966) or by *AlphaFold2* (TP0462, TP0179) for genes with high frequency, population-informative mutations, and visualized using *Chimera* (v1·16).

*Study outcomes and sample size*

Primary outcomes of this study included the number and frequency of unique *TPA* genomic sequences distributed geographically and associated individual-level characteristics. Secondary outcomes include the predominant clades, subclades and alleles identified in our study population and relative to the *TPA* global population structure. Target sample sizes at each site were informed by rarefaction curves, which are classically used in ecological studies to depict observed species richness at different sample sizes[8] but also used to assess allelic diversity in genomic analyses. Based on estimates of the genetic diversity of a subset of *TPA* outer membrane protein genes available at the time of study launch,[9] we estimated that sequences of 30-50 *TPA* strains from each clinical site (target total sample size of 120-200 for four sites) were required in order to adequately sample each site's *TPA* OMP genetic diversity with 95% confidence.

**Supplementary Results**

*TPA qPCR*

*TPA polA* qPCR was positive from 77% (n=59) of study participants with PS, 88% (n=136) with SS, and 100% (n=2) with ELS (Figure 1); however, the two participants with ELS had very low qPCR copy numbers from whole blood compared to other sample types from participants with PS or SS. Overall, there were 277 samples with positive PCR results, including 108 (83%) lesion swabs, 92 (85%) skin biopsies, seven (37%) skin scrapings, and 70 (51%) whole blood specimens (Table 2). Genital ulcer swabs from Malawi had qPCR values with geometric means of 1016 copies/µl (range 12-10,438), compared to 39 (range 1-862) and 194 (range 11-8,835) copies/µl from the China and Colombia sites, respectively (Table 2). SS lesion swabs from Malawi had the highest mean values of 1858 (range 115-82,147) and 2830 (range 235-20,143) copies/µl for both DFM-positive and DFM-negative samples, respectively. *TPA* qPCR copy numbers in blood from SS participants based on nontreponemal titers ≤ 1:32 versus > 1:32 were similar.

**Supplementary Table 1:** Genomes and metadata for samples included in this study, comprising both new and publicly available genomes.

| Run | BioProject | BioSample | Sample_Name | Geo_country | Continent | Date_collected | Publication | Clade | BAPS |
|---|---|---|---|---|---|---|---|---|---|
| DRR213710 | PRJDB9408 | SAMD00208592 | 14B001UK | Japan | Asia | 2014 | Nishiki_2021 | Nichols | Pop1 |
| DRR213711 | PRJDB9408 | SAMD00208593 | 15A011MM | Japan | Asia | 2015 | Nishiki_2021 | SS14 | Pop7 |
| DRR213712 | PRJDB9408 | SAMD00208594 | 15A019HM | Japan | Asia | 2015 | Nishiki_2021 | SS14 | Pop7 |
| DRR213713 | PRJDB9408 | SAMD00208595 | 16A013MM | Japan | Asia | 2016 | Nishiki_2021 | SS14 | Pop8 |
| DRR213714 | PRJDB9408 | SAMD00208596 | 17A003MM | Japan | Asia | 2017 | Nishiki_2021 | Nichols | Pop1 |
| DRR213715 | PRJDB9408 | SAMD00208597 | 17A010MM | Japan | Asia | 2017 | Nishiki_2021 | SS14 | Pop8 |
| DRR213716 | PRJDB9408 | SAMD00208598 | 17A014MM | Japan | Asia | 2017 | Nishiki_2021 | SS14 | Pop8 |
| DRR213717 | PRJDB9408 | SAMD00208599 | 17A017MM | Japan | Asia | 2017 | Nishiki_2021 | SS14 | Pop7 |
| DRR213718 | PRJDB9408 | SAMD00208600 | 17A021MM | Japan | Asia | 2017 | Nishiki_2021 | Nichols | Pop1 |
| DRR213719 | PRJDB9408 | SAMD00208601 | 17C005HM | Japan | Asia | 2017 | Nishiki_2021 | SS14 | Pop8 |
| DRR213720 | PRJDB9408 | SAMD00208602 | 17C009HM | Japan | Asia | 2017 | Nishiki_2021 | SS14 | Pop8 |
| DRR213721 | PRJDB9408 | SAMD00208603 | 17D002HM | Japan | Asia | 2017 | Nishiki_2021 | SS14 | Pop7 |
| DRR213722 | PRJDB9408 | SAMD00208604 | 17D006HM | Japan | Asia | 2017 | Nishiki_2021 | SS14 | Pop7 |
| DRR213723 | PRJDB9408 | SAMD00208605 | 17D009HM | Japan | Asia | 2017 | Nishiki_2021 | SS14 | Pop7 |
| DRR213724 | PRJDB9408 | SAMD00208606 | 17E002HF | Japan | Asia | 2017 | Nishiki_2021 | SS14 | Pop7 |
| DRR213725 | PRJDB9408 | SAMD00208607 | 18C004HM | Japan | Asia | 2018 | Nishiki_2021 | SS14 | Pop7 |
| DRR213726 | PRJDB9408 | SAMD00208608 | 18D001HM | Japan | Asia | 2018 | Nishiki_2021 | SS14 | Pop7 |
| DRR213727 | PRJDB9408 | SAMD00208609 | 18D002HM | Japan | Asia | 2018 | Nishiki_2021 | SS14 | Pop7 |
| DRR213728 | PRJDB9408 | SAMD00208610 | 18E001HF | Japan | Asia | 2018 | Nishiki_2021 | SS14 | Pop7 |
| DRR213729 | PRJDB9408 | SAMD00208611 | 18E002HF | Japan | Asia | 2018 | Nishiki_2021 | SS14 | Pop7 |
| DRR213730 | PRJDB9408 | SAMD00208612 | 18E016HF | Japan | Asia | 2018 | Nishiki_2021 | SS14 | Pop8 |
| ERR2756160 | PRJEB20795 | SAMEA4857071 | Nichols Houston J | USA | N.America | 1912 | Beale_2019 | Nichols | Pop1 |
| ERR2756162 | PRJEB20795 | SAMEA4857073 | Nichols Houston E | USA | N.America | 1912 | Beale_2019 | Nichols | Pop1 |
| ERR2756163 | PRJEB20795 | SAMEA4857074 | Nichols Houston O | USA | N.America | 1912 | Beale_2019 | Nichols | Pop1 |
| ERR2756168 | PRJEB20795 | SAMEA104225534 | UW298B | USA | N.America | 2004 | Beale_2019 | SS14 | Pop7 |
| ERR2756169 | PRJEB20795 | SAMEA104225539 | UW370B | USA | N.America | 2005 | Beale_2019 | SS14 | Pop7 |
| ERR2756170 | PRJEB20795 | SAMEA104225540 | UW824B | USA | N.America | 2011 | Beale_2019 | SS14 | Pop6 |
| ERR2756171 | PRJEB20795 | SAMEA104225549 | UW116B | USA | N.America | 2002 | Beale_2019 | SS14 | Pop7 |
| ERR2756172 | PRJEB20795 | SAMEA104225554 | UW262B | USA | N.America | 2004 | Beale_2019 | SS14 | Pop7 |
| ERR2756173 | PRJEB20795 | SAMEA104225555 | UW337B | USA | N.America | 2005 | Beale_2019 | SS14 | Pop7 |
| ERR2756174 | PRJEB20795 | SAMEA104225558 | UW391B | USA | N.America | 2005 | Beale_2019 | SS14 | Pop7 |
| ERR2756175 | PRJEB20795 | SAMEA104225561 | UW099B | USA | N.America | 2001 | Beale_2019 | SS14 | Pop7 |
| ERR2756176 | PRJEB20795 | SAMEA104225562 | UW291B | USA | N.America | 2005 | Beale_2019 | SS14 | Pop7 |
| ERR2756177 | PRJEB20795 | SAMEA104225567 | UW376B | USA | N.America | 2005 | Beale_2019 | SS14 | Pop7 |
| ERR2756178 | PRJEB20795 | SAMEA104225568 | UW248B | USA | N.America | 2004 | Beale_2019 | SS14 | Pop7 |
| ERR2756179 | PRJEB20795 | SAMEA104225570 | UW823B | USA | N.America | 2011 | Beale_2019 | SS14 | Pop6 |
| ERR2756181 | PRJEB20795 | SAMEA104225574 | UW397B | USA | N.America | 2006 | Beale_2019 | SS14 | Pop6 |
| ERR2756182 | PRJEB20795 | SAMEA104225575 | UW257B | USA | N.America | 2004 | Beale_2019 | SS14 | Pop7 |
| ERR2756184 | PRJEB20795 | SAMEA104225578 | UW473B | USA | N.America | 2006 | Beale_2019 | SS14 | Pop7 |
| ERR2756185 | PRJEB20795 | SAMEA104225579 | UW140B | USA | N.America | 2002 | Beale_2019 | SS14 | Pop7 |

| | | | | | | | | | |
|---|---|---|---|---|---|---|---|---|---|
| ERR2756186 | PRJEB20795 | SAMEA104225580 | UW186B | USA | N.America | 2003 | Beale_2019 | SS14 | Pop7 |
| ERR2756187 | PRJEB20795 | SAMEA104225581 | UW189B | USA | N.America | 2004 | Beale_2019 | Nichols | Pop2 |
| ERR2756188 | PRJEB20795 | SAMEA104225583 | UW133B | USA | N.America | 2002 | Beale_2019 | SS14 | Pop7 |
| ERR2756189 | PRJEB20795 | SAMEA104225584 | UW344B | USA | N.America | 2005 | Beale_2019 | SS14 | Pop7 |
| ERR2756190 | PRJEB20795 | SAMEA104225585 | UW254B | USA | N.America | 2004 | Beale_2019 | SS14 | Pop7 |
| ERR2756191 | PRJEB20795 | SAMEA104225586 | UW411B | USA | N.America | 2006 | Beale_2019 | SS14 | Pop6 |
| ERR2756192 | PRJEB20795 | SAMEA104225587 | UW187B | USA | N.America | 2003 | Beale_2019 | SS14 | Pop7 |
| ERR2756193 | PRJEB20795 | SAMEA104225588 | UW228B | USA | N.America | 2004 | Beale_2019 | SS14 | Pop7 |
| ERR2756194 | PRJEB20795 | SAMEA104225589 | UW102B | USA | N.America | 2002 | Beale_2019 | SS14 | Pop7 |
| ERR2756195 | PRJEB20795 | SAMEA104225590 | UW074B | USA | N.America | 2004 | Beale_2019 | SS14 | Pop6 |
| ERR2756196 | PRJEB20795 | SAMEA104225591 | UW368B | USA | N.America | 2005 | Beale_2019 | SS14 | Pop7 |
| ERR2756197 | PRJEB20795 | SAMEA104225592 | UW279B | USA | N.America | 2004 | Beale_2019 | Nichols | Pop2 |
| ERR2756198 | PRJEB20795 | SAMEA104225594 | UW125B | USA | N.America | 2002 | Beale_2019 | SS14 | Pop7 |
| ERR2756199 | PRJEB20795 | SAMEA104225595 | UW330B | USA | N.America | 2005 | Beale_2019 | SS14 | Pop7 |
| ERR2756200 | PRJEB20795 | SAMEA104225597 | UW147B | USA | N.America | 2003 | Beale_2019 | SS14 | Pop7 |
| ERR2756201 | PRJEB20795 | SAMEA104225598 | UW138B | USA | N.America | 2002 | Beale_2019 | SS14 | Pop7 |
| ERR2756202 | PRJEB20795 | SAMEA104225600 | UW181B | USA | N.America | 2003 | Beale_2019 | SS14 | Pop7 |
| ERR2756203 | PRJEB20795 | SAMEA104225601 | UW148B | USA | N.America | 2003 | Beale_2019 | SS14 | Pop7 |
| ERR2756204 | PRJEB20795 | SAMEA104225602 | UW852B | USA | N.America | 2011 | Beale_2019 | SS14 | Pop6 |
| ERR2756205 | PRJEB20795 | SAMEA104225603 | UW213B | USA | N.America | 2003 | Beale_2019 | SS14 | Pop7 |
| ERR2756206 | PRJEB20795 | SAMEA104225605 | UW379B | USA | N.America | 2005 | Beale_2019 | SS14 | Pop6 |
| ERR2756207 | PRJEB20795 | SAMEA104225606 | UW126B | USA | N.America | 2002 | Beale_2019 | SS14 | Pop7 |
| ERR2756208 | PRJEB20795 | SAMEA104225607 | UW383B | USA | N.America | 2005 | Beale_2019 | SS14 | Pop6 |
| ERR2756210 | PRJEB20795 | SAMEA104225609 | UW231B | USA | N.America | 2004 | Beale_2019 | SS14 | Pop7 |
| ERR2756211 | PRJEB20795 | SAMEA104225610 | UW281B | USA | N.America | 2004 | Beale_2019 | SS14 | Pop7 |
| ERR2756212 | PRJEB20795 | SAMEA104225611 | UW211B | USA | N.America | 2003 | Beale_2019 | SS14 | Pop7 |
| ERR2756213 | PRJEB20795 | SAMEA104225613 | UW215B | USA | N.America | 2004 | Beale_2019 | SS14 | Pop7 |
| ERR2756214 | PRJEB20795 | SAMEA104225615 | UW093B | USA | N.America | 2001 | Beale_2019 | SS14 | Pop7 |
| ERR2756216 | PRJEB20795 | SAMEA104225618 | UW492B | USA | N.America | 2006 | Beale_2019 | SS14 | Pop7 |
| ERR2756217 | PRJEB20795 | SAMEA104225487 | UW244B | USA | N.America | 2004 | Beale_2019 | SS14 | Pop7 |
| ERR2756218 | PRJEB20795 | SAMEA104225488 | UW259B | USA | N.America | 2004 | Beale_2019 | SS14 | Pop6 |
| ERR2756219 | PRJEB20795 | SAMEA104225489 | UW134B | USA | N.America | 2002 | Beale_2019 | SS14 | Pop7 |
| ERR2756220 | PRJEB20795 | SAMEA104225490 | UW526B | USA | N.America | 2007 | Beale_2019 | SS14 | Pop6 |
| ERR2756221 | PRJEB20795 | SAMEA104225493 | UW155B | USA | N.America | 2003 | Beale_2019 | SS14 | Pop7 |
| ERR2756222 | PRJEB20795 | SAMEA104225494 | UW280B | USA | N.America | 2004 | Beale_2019 | SS14 | Pop7 |
| ERR2756223 | PRJEB20795 | SAMEA104225496 | UW195B | USA | N.America | 2003 | Beale_2019 | SS14 | Pop7 |
| ERR2756224 | PRJEB20795 | SAMEA104225497 | UW304B | USA | N.America | 2005 | Beale_2019 | SS14 | Pop6 |
| ERR2756225 | PRJEB20795 | SAMEA104225498 | UW327B | USA | N.America | 2005 | Beale_2019 | SS14 | Pop6 |
| ERR2756226 | PRJEB20795 | SAMEA104225500 | UW264B | USA | N.America | 2004 | Beale_2019 | SS14 | Pop6 |
| ERR2756227 | PRJEB20795 | SAMEA104225501 | UW149B | USA | N.America | 2003 | Beale_2019 | SS14 | Pop7 |
| ERR2756228 | PRJEB20795 | SAMEA104225502 | UW303B | USA | N.America | 2005 | Beale_2019 | SS14 | Pop6 |
| ERR2756229 | PRJEB20795 | SAMEA104225503 | UW104B | USA | N.America | 2002 | Beale_2019 | SS14 | Pop7 |
| ERR3684452 | PRJEB28546 | SAMEA5145700 | PHE140085A | UK | Europe | 2014 | Beale_2021 | Nichols | Pop3 |
| ERR3684453 | PRJEB28546 | SAMEA5145701 | PHE150121A | UK | Europe | 2015 | Beale_2021 | Nichols | Pop3 |

| | | | | | | | | | |
|---|---|---|---|---|---|---|---|---|---|
| ERR3684454 | PRJEB28546 | SAMEA5145702 | PHE150145A | UK | Europe | 2015 | Beale_2021 | Nichols | Pop3 |
| ERR3684455 | PRJEB28546 | SAMEA5145703 | PHE150150A | UK | Europe | 2015 | Beale_2021 | Nichols | Pop3 |
| ERR3684456 | PRJEB28546 | SAMEA5145704 | PHE160190A | UK | Europe | 2016 | Beale_2021 | SS14 | Pop6 |
| ERR3684457 | PRJEB28546 | SAMEA5145705 | PHE160196A | UK | Europe | 2016 | Beale_2021 | Nichols | Pop3 |
| ERR3684458 | PRJEB28546 | SAMEA5145706 | PHE160224A | UK | Europe | 2016 | Beale_2021 | Nichols | Pop3 |
| ERR3684459 | PRJEB28546 | SAMEA5145707 | PHE160243A | UK | Europe | 2016 | Beale_2021 | Nichols | Pop3 |
| ERR3684460 | PRJEB28546 | SAMEA5145708 | PHE160249A | UK | Europe | 2016 | Beale_2021 | SS14 | Pop7 |
| ERR3684461 | PRJEB28546 | SAMEA5145709 | PHE160255A | UK | Europe | 2016 | Beale_2021 | Nichols | Pop3 |
| ERR3684462 | PRJEB28546 | SAMEA5145710 | PHE160263A | UK | Europe | 2016 | Beale_2021 | Nichols | Pop3 |
| ERR3684463 | PRJEB28546 | SAMEA5145711 | PHE160270A | UK | Europe | 2016 | Beale_2021 | Nichols | Pop3 |
| ERR3684464 | PRJEB28546 | SAMEA5145712 | PHE160274A | UK | Europe | 2016 | Beale_2021 | Nichols | Pop3 |
| ERR3684467 | PRJEB28546 | SAMEA5145715 | PHE160287A | UK | Europe | 2016 | Beale_2021 | Nichols | Pop3 |
| ERR3684468 | PRJEB28546 | SAMEA5145716 | PHE160294A | UK | Europe | 2016 | Beale_2021 | Nichols | Pop3 |
| ERR3684469 | PRJEB28546 | SAMEA5145717 | PHE160316A | UK | Europe | 2016 | Beale_2021 | Nichols | Pop3 |
| ERR3684470 | PRJEB28546 | SAMEA5145718 | PHE160317A | UK | Europe | 2016 | Beale_2021 | Nichols | Pop3 |
| ERR3684471 | PRJEB28546 | SAMEA5145719 | PHE170322A | UK | Europe | 2017 | Beale_2021 | Nichols | Pop2 |
| ERR3684472 | PRJEB28546 | SAMEA5145720 | PHE170333A | UK | Europe | 2017 | Beale_2021 | Nichols | Pop3 |
| ERR3684474 | PRJEB28546 | SAMEA5145722 | PHE170349A | UK | Europe | 2017 | Beale_2021 | Nichols | Pop3 |
| ERR3684475 | PRJEB28546 | SAMEA5145723 | PHE170372A | UK | Europe | 2017 | Beale_2021 | Nichols | Pop3 |
| ERR3684476 | PRJEB28546 | SAMEA5145724 | PHE170374A | UK | Europe | 2017 | Beale_2021 | Nichols | Pop3 |
| ERR3684477 | PRJEB28546 | SAMEA5145733 | PHE170381A | UK | Europe | 2017 | Beale_2021 | Nichols | Pop3 |
| ERR3684478 | PRJEB28546 | SAMEA5145734 | PHE170386A | UK | Europe | 2017 | Beale_2021 | Nichols | Pop3 |
| ERR3684479 | PRJEB28546 | SAMEA5145735 | PHE170405A | UK | Europe | 2017 | Beale_2021 | Nichols | Pop3 |
| ERR3684480 | PRJEB28546 | SAMEA5145736 | PHE170407A | UK | Europe | 2017 | Beale_2021 | SS14 | Pop6 |
| ERR3684481 | PRJEB28546 | SAMEA5145737 | PHE170413A | UK | Europe | 2017 | Beale_2021 | Nichols | Pop3 |
| ERR3684482 | PRJEB28546 | SAMEA5145738 | PHE120011A | UK | Europe | 2012 | Beale_2021 | SS14 | Pop6 |
| ERR3684484 | PRJEB28546 | SAMEA5145740 | PHE120029A | UK | Europe | 2012 | Beale_2021 | Nichols | Pop1 |
| ERR3684485 | PRJEB28546 | SAMEA5145741 | PHE130043A | UK | Europe | 2013 | Beale_2021 | SS14 | Pop8 |
| ERR3684486 | PRJEB28546 | SAMEA5145742 | PHE140089A | UK | Europe | 2014 | Beale_2021 | Nichols | Pop3 |
| ERR3684487 | PRJEB28546 | SAMEA5145743 | PHE150114A | UK | Europe | 2015 | Beale_2021 | Nichols | Pop2 |
| ERR3684488 | PRJEB28546 | SAMEA5145744 | PHE150115A | UK | Europe | 2015 | Beale_2021 | Nichols | Pop2 |
| ERR3684489 | PRJEB28546 | SAMEA5145745 | PHE150126A | UK | Europe | 2015 | Beale_2021 | SS14 | Pop6 |
| ERR3684490 | PRJEB28546 | SAMEA5145746 | PHE150133A | UK | Europe | 2015 | Beale_2021 | Nichols | Pop3 |
| ERR3684491 | PRJEB28546 | SAMEA5145747 | PHE150149A | UK | Europe | 2015 | Beale_2021 | Nichols | Pop3 |
| ERR3684492 | PRJEB28546 | SAMEA5145748 | PHE150162A | UK | Europe | 2015 | Beale_2021 | Nichols | Pop3 |
| ERR3684493 | PRJEB28546 | SAMEA5145749 | PHE150166A | UK | Europe | 2015 | Beale_2021 | Nichols | Pop3 |
| ERR3684494 | PRJEB28546 | SAMEA5145750 | PHE150170A | UK | Europe | 2015 | Beale_2021 | Nichols | Pop3 |
| ERR3684495 | PRJEB28546 | SAMEA5145751 | PHE160203A | UK | Europe | 2016 | Beale_2021 | SS14 | Pop6 |
| ERR3684496 | PRJEB28546 | SAMEA5145752 | PHE160206A | UK | Europe | 2016 | Beale_2021 | Nichols | Pop3 |
| ERR3684497 | PRJEB28546 | SAMEA5145753 | PHE160214A | UK | Europe | 2016 | Beale_2021 | SS14 | Pop6 |
| ERR3684498 | PRJEB28546 | SAMEA5145754 | PHE160240A | UK | Europe | 2016 | Beale_2021 | SS14 | Pop8 |
| ERR3684499 | PRJEB28546 | SAMEA5145755 | PHE160253A | UK | Europe | 2016 | Beale_2021 | SS14 | Pop6 |
| ERR3684500 | PRJEB28546 | SAMEA5145756 | PHE160259A | UK | Europe | 2016 | Beale_2021 | SS14 | Pop7 |
| ERR3684501 | PRJEB28546 | SAMEA5145757 | PHE160262A | UK | Europe | 2016 | Beale_2021 | SS14 | Pop6 |

| | | | | | | | | | |
|---|---|---|---|---|---|---|---|---|---|
| ERR3684502 | PRJEB28546 | SAMEA5145758 | PHE160264A | UK | Europe | 2016 | Beale_2021 | SS14 | Pop6 |
| ERR3684504 | PRJEB28546 | SAMEA5145760 | PHE160280A | UK | Europe | 2016 | Beale_2021 | SS14 | Pop8 |
| ERR3684507 | PRJEB28546 | SAMEA5145725 | PHE160298A | UK | Europe | 2016 | Beale_2021 | SS14 | Pop6 |
| ERR3684509 | PRJEB28546 | SAMEA5145727 | PHE160312A | UK | Europe | 2016 | Beale_2021 | SS14 | Pop6 |
| ERR3684510 | PRJEB28546 | SAMEA5145728 | PHE160315A | UK | Europe | 2016 | Beale_2021 | SS14 | Pop7 |
| ERR3684511 | PRJEB28546 | SAMEA5145729 | PHE170328A | UK | Europe | 2017 | Beale_2021 | SS14 | Pop6 |
| ERR3684512 | PRJEB28546 | SAMEA5145730 | PHE170346A | UK | Europe | 2017 | Beale_2021 | SS14 | Pop6 |
| ERR3684515 | PRJEB28546 | SAMEA5145763 | PHE170387A | UK | Europe | 2017 | Beale_2021 | SS14 | Pop6 |
| ERR3684516 | PRJEB28546 | SAMEA5145764 | PHE170397A | UK | Europe | 2017 | Beale_2021 | Nichols | Pop3 |
| RR3684517 | PRJEB28546 | SAMEA5145765 | PHE170408A | UK | Europe | 2017 | Beale_2021 | SS14 | Pop7 |
| ERR3684519 | PRJEB28546 | SAMEA5145767 | PHE120009B | UK | Europe | 2012 | Beale_2021 | SS14 | Pop6 |
| ERR3684520 | PRJEB28546 | SAMEA5145768 | PHE120021A | UK | Europe | 2012 | Beale_2021 | SS14 | Pop8 |
| ERR3684521 | PRJEB28546 | SAMEA5145769 | PHE120024A | UK | Europe | 2012 | Beale_2021 | SS14 | Pop8 |
| ERR3684522 | PRJEB28546 | SAMEA5145770 | PHE130039A | UK | Europe | 2013 | Beale_2021 | SS14 | Pop7 |
| ERR3684523 | PRJEB28546 | SAMEA5145771 | PHE130045A | UK | Europe | 2013 | Beale_2021 | SS14 | Pop6 |
| ERR3684524 | PRJEB28546 | SAMEA5145772 | PHE130064A | UK | Europe | 2013 | Beale_2021 | SS14 | Pop6 |
| ERR3684525 | PRJEB28546 | SAMEA5145773 | PHE140074A | UK | Europe | 2014 | Beale_2021 | SS14 | Pop6 |
| ERR3684526 | PRJEB28546 | SAMEA5145774 | PHE140095A | UK | Europe | 2014 | Beale_2021 | SS14 | Pop6 |
| ERR3684527 | PRJEB28546 | SAMEA5145775 | PHE140107A | UK | Europe | 2014 | Beale_2021 | Nichols | Pop3 |
| ERR3684528 | PRJEB28546 | SAMEA5145776 | PHE150110A | UK | Europe | 2015 | Beale_2021 | SS14 | Pop6 |
| ERR3684529 | PRJEB28546 | SAMEA5145777 | PHE150139A | UK | Europe | 2015 | Beale_2021 | Nichols | Pop3 |
| ERR3684530 | PRJEB28546 | SAMEA5145778 | PHE150148A | UK | Europe | 2015 | Beale_2021 | SS14 | Pop6 |
| ERR3684531 | PRJEB28546 | SAMEA5145779 | PHE150177A | UK | Europe | 2015 | Beale_2021 | SS14 | Pop8 |
| ERR3684532 | PRJEB28546 | SAMEA5145780 | PHE150181A | UK | Europe | 2015 | Beale_2021 | Nichols | Pop3 |
| ERR3684533 | PRJEB28546 | SAMEA5145781 | PHE160198A | UK | Europe | 2016 | Beale_2021 | SS14 | Pop6 |
| ERR3684534 | PRJEB28546 | SAMEA5145782 | PHE160211A | UK | Europe | 2016 | Beale_2021 | SS14 | Pop6 |
| ERR3684535 | PRJEB28546 | SAMEA5145783 | PHE160217A | UK | Europe | 2016 | Beale_2021 | SS14 | Pop6 |
| ERR3684536 | PRJEB28546 | SAMEA5145784 | PHE160233A | UK | Europe | 2016 | Beale_2021 | Nichols | Pop3 |
| ERR3684537 | PRJEB28546 | SAMEA5145785 | PHE160239A | UK | Europe | 2016 | Beale_2021 | SS14 | Pop6 |
| ERR3684538 | PRJEB28546 | SAMEA5145786 | PHE160256A | UK | Europe | 2016 | Beale_2021 | SS14 | Pop8 |
| ERR3684539 | PRJEB28546 | SAMEA5145787 | PHE160268A | UK | Europe | 2016 | Beale_2021 | SS14 | Pop8 |
| ERR3684540 | PRJEB28546 | SAMEA5145788 | PHE160277A | UK | Europe | 2016 | Beale_2021 | SS14 | Pop8 |
| ERR3684542 | PRJEB28546 | SAMEA5145790 | PHE160299A | UK | Europe | 2016 | Beale_2021 | SS14 | Pop6 |
| ERR3684543 | PRJEB28546 | SAMEA5145791 | PHE160300A | UK | Europe | 2016 | Beale_2021 | SS14 | Pop6 |
| ERR3684544 | PRJEB28546 | SAMEA5145792 | PHE160301A | UK | Europe | 2016 | Beale_2021 | SS14 | Pop6 |
| ERR3684547 | PRJEB28546 | SAMEA5145795 | PHE160309A | UK | Europe | 2016 | Beale_2021 | SS14 | Pop8 |
| ERR3684548 | PRJEB28546 | SAMEA5145796 | PHE170329A | UK | Europe | 2017 | Beale_2021 | SS14 | Pop6 |
| ERR3684549 | PRJEB28546 | SAMEA5145797 | PHE170336A | UK | Europe | 2017 | Beale_2021 | Nichols | Pop2 |
| ERR3684550 | PRJEB28546 | SAMEA5145798 | PHE170343A | UK | Europe | 2017 | Beale_2021 | SS14 | Pop6 |
| ERR3684551 | PRJEB28546 | SAMEA5145799 | PHE170351A | UK | Europe | 2017 | Beale_2021 | SS14 | Pop6 |
| ERR3684552 | PRJEB28546 | SAMEA5145800 | PHE170352A | UK | Europe | 2017 | Beale_2021 | SS14 | Pop7 |
| ERR3684553 | PRJEB28546 | SAMEA5145801 | PHE170356A | UK | Europe | 2017 | Beale_2021 | SS14 | Pop6 |
| ERR3684555 | PRJEB28546 | SAMEA5145803 | PHE170367A | UK | Europe | 2017 | Beale_2021 | SS14 | Pop8 |
| ERR3684556 | PRJEB28546 | SAMEA5145804 | PHE170380A | UK | Europe | 2017 | Beale_2021 | SS14 | Pop7 |

| | | | | | | | | | |
|---|---|---|---|---|---|---|---|---|---|
| ERR3684557 | PRJEB28546 | SAMEA5145805 | PHE170384A | UK | Europe | 2017 | Beale_2021 | SS14 | Pop6 |
| ERR3684558 | PRJEB28546 | SAMEA5145806 | PHE170388A | UK | Europe | 2017 | Beale_2021 | SS14 | Pop6 |
| ERR3684560 | PRJEB28546 | SAMEA5145808 | PHE170402A | UK | Europe | 2017 | Beale_2021 | SS14 | Pop7 |
| ERR3684564 | PRJEB28546 | SAMEA5145812 | PHE120007A | UK | Europe | 2012 | Beale_2021 | SS14 | Pop6 |
| ERR3684565 | PRJEB28546 | SAMEA5145813 | PHE120013A | UK | Europe | 2012 | Beale_2021 | SS14 | Pop6 |
| ERR3684566 | PRJEB28546 | SAMEA5145814 | PHE120019A | UK | Europe | 2012 | Beale_2021 | SS14 | Pop6 |
| ERR3684567 | PRJEB28546 | SAMEA5145815 | PHE120022A | UK | Europe | 2012 | Beale_2021 | SS14 | Pop8 |
| ERR3684568 | PRJEB28546 | SAMEA5145816 | PHE130036A | UK | Europe | 2013 | Beale_2021 | SS14 | Pop8 |
| ERR3684569 | PRJEB28546 | SAMEA5145817 | PHE130047A | UK | Europe | 2013 | Beale_2021 | SS14 | Pop8 |
| ERR3684571 | PRJEB28546 | SAMEA5145819 | PHE130054A | UK | Europe | 2013 | Beale_2021 | SS14 | Pop8 |
| ERR3684572 | PRJEB28546 | SAMEA5145820 | PHE130056A | UK | Europe | 2013 | Beale_2021 | SS14 | Pop6 |
| ERR3684573 | PRJEB28546 | SAMEA5145821 | PHE140073A | UK | Europe | 2014 | Beale_2021 | SS14 | Pop7 |
| ERR3684574 | PRJEB28546 | SAMEA5145822 | PHE150130A | UK | Europe | 2015 | Beale_2021 | SS14 | Pop6 |
| ERR3684575 | PRJEB28546 | SAMEA5145823 | PHE150131A | UK | Europe | 2015 | Beale_2021 | SS14 | Pop7 |
| ERR3684576 | PRJEB28546 | SAMEA5145824 | PHE150137A | UK | Europe | 2015 | Beale_2021 | SS14 | Pop7 |
| ERR3684577 | PRJEB28546 | SAMEA5145825 | PHE150138A | UK | Europe | 2015 | Beale_2021 | SS14 | Pop6 |
| ERR3684580 | PRJEB28546 | SAMEA5145828 | PHE150153A | UK | Europe | 2015 | Beale_2021 | SS14 | Pop6 |
| ERR3684581 | PRJEB28546 | SAMEA5145829 | PHE150154A | UK | Europe | 2015 | Beale_2021 | SS14 | Pop8 |
| ERR3684582 | PRJEB28546 | SAMEA5145830 | PHE150160A | UK | Europe | 2015 | Beale_2021 | SS14 | Pop6 |
| ERR3684583 | PRJEB28546 | SAMEA5145831 | PHE150163A | UK | Europe | 2015 | Beale_2021 | SS14 | Pop8 |
| ERR3684584 | PRJEB28546 | SAMEA5145832 | PHE160197A | UK | Europe | 2016 | Beale_2021 | SS14 | Pop8 |
| ERR3684585 | PRJEB28546 | SAMEA5145833 | PHE160215A | UK | Europe | 2016 | Beale_2021 | SS14 | Pop7 |
| ERR3684587 | PRJEB28546 | SAMEA5145835 | PHE160234A | UK | Europe | 2016 | Beale_2021 | SS14 | Pop8 |
| ERR3684588 | PRJEB28546 | SAMEA5145836 | PHE160247A | UK | Europe | 2016 | Beale_2021 | SS14 | Pop8 |
| ERR3684589 | PRJEB28546 | SAMEA5145837 | PHE160254A | UK | Europe | 2016 | Beale_2021 | SS14 | Pop8 |
| ERR3684590 | PRJEB28546 | SAMEA5145838 | PHE160257A | UK | Europe | 2016 | Beale_2021 | SS14 | Pop8 |
| ERR3684592 | PRJEB28546 | SAMEA5145840 | PHE160292A | UK | Europe | 2016 | Beale_2021 | SS14 | Pop8 |
| ERR3684593 | PRJEB28546 | SAMEA5145841 | PHE160308A | UK | Europe | 2016 | Beale_2021 | SS14 | Pop8 |
| ERR3684594 | PRJEB28546 | SAMEA5145842 | PHE160313A | UK | Europe | 2016 | Beale_2021 | SS14 | Pop8 |
| ERR3684596 | PRJEB28546 | SAMEA5145844 | PHE170355A | UK | Europe | 2017 | Beale_2021 | SS14 | Pop6 |
| ERR3684599 | PRJEB28546 | SAMEA5145847 | PHE170385A | UK | Europe | 2017 | Beale_2021 | SS14 | Pop8 |
| ERR3684600 | PRJEB28546 | SAMEA5145848 | PHE170398A | UK | Europe | 2017 | Beale_2021 | SS14 | Pop7 |
| ERR3684601 | PRJEB28546 | SAMEA5145849 | PHE170411A | UK | Europe | 2017 | Beale_2021 | SS14 | Pop8 |
| ERR3684602 | PRJEB28546 | SAMEA5145850 | PHE170412A | UK | Europe | 2017 | Beale_2021 | SS14 | Pop8 |
| ERR3684606 | PRJEB28546 | SAMEA5145855 | PHE160628A | UK | Europe | 2016 | Beale_2021 | SS14 | Pop8 |
| ERR3684609 | PRJEB28546 | SAMEA5145858 | PHE120001A | UK | Europe | 2012 | Beale_2021 | SS14 | Pop8 |
| ERR3684610 | PRJEB28546 | SAMEA5145859 | PHE120006A | UK | Europe | 2012 | Beale_2021 | SS14 | Pop8 |
| ERR3684613 | PRJEB28546 | SAMEA5145862 | PHE130053A | UK | Europe | 2013 | Beale_2021 | SS14 | Pop8 |
| ERR3684615 | PRJEB28546 | SAMEA5145864 | PHE140093A | UK | Europe | 2014 | Beale_2021 | SS14 | Pop8 |
| ERR3684616 | PRJEB28546 | SAMEA5145865 | PHE150113A | UK | Europe | 2015 | Beale_2021 | SS14 | Pop6 |
| ERR3684618 | PRJEB28546 | SAMEA5145867 | PHE150119A | UK | Europe | 2015 | Beale_2021 | SS14 | Pop8 |
| ERR3684619 | PRJEB28546 | SAMEA5145868 | PHE150122A | UK | Europe | 2015 | Beale_2021 | SS14 | Pop8 |
| ERR3684620 | PRJEB28546 | SAMEA5145869 | PHE150161A | UK | Europe | 2015 | Beale_2021 | SS14 | Pop8 |
| ERR3684622 | PRJEB28546 | SAMEA5145871 | PHE150175A | UK | Europe | 2015 | Beale_2021 | SS14 | Pop8 |

| | | | | | | | | | |
|---|---|---|---|---|---|---|---|---|---|
| ERR3684623 | PRJEB28546 | SAMEA5145872 | PHE160189A | UK | Europe | 2016 | Beale_2021 | SS14 | Pop6 |
| ERR3684624 | PRJEB28546 | SAMEA5145873 | PHE160199A | UK | Europe | 2016 | Beale_2021 | SS14 | Pop8 |
| ERR3684626 | PRJEB28546 | SAMEA5145875 | PHE160238A | UK | Europe | 2016 | Beale_2021 | SS14 | Pop6 |
| ERR3684628 | PRJEB28546 | SAMEA5145877 | PHE160245A | UK | Europe | 2016 | Beale_2021 | SS14 | Pop8 |
| ERR3684630 | PRJEB28546 | SAMEA5145879 | PHE160248A | UK | Europe | 2016 | Beale_2021 | SS14 | Pop8 |
| ERR3684631 | PRJEB28546 | SAMEA5145880 | PHE160260A | UK | Europe | 2016 | Beale_2021 | SS14 | Pop8 |
| ERR3684632 | PRJEB28546 | SAMEA5145881 | PHE160261A | UK | Europe | 2016 | Beale_2021 | SS14 | Pop8 |
| ERR3684634 | PRJEB28546 | SAMEA5145883 | PHE160267A | UK | Europe | 2016 | Beale_2021 | SS14 | Pop8 |
| ERR3684635 | PRJEB28546 | SAMEA5145884 | PHE160272A | UK | Europe | 2016 | Beale_2021 | SS14 | Pop8 |
| ERR3684638 | PRJEB28546 | SAMEA5145887 | PHE160303A | UK | Europe | 2016 | Beale_2021 | SS14 | Pop8 |
| ERR3684639 | PRJEB28546 | SAMEA5145888 | PHE160311A | UK | Europe | 2016 | Beale_2021 | SS14 | Pop8 |
| ERR3684641 | PRJEB28546 | SAMEA5145890 | PHE170345A | UK | Europe | 2017 | Beale_2021 | SS14 | Pop8 |
| ERR3684643 | PRJEB28546 | SAMEA5145892 | PHE170366A | UK | Europe | 2017 | Beale_2021 | SS14 | Pop8 |
| ERR3684644 | PRJEB28546 | SAMEA5145893 | PHE170378A | UK | Europe | 2017 | Beale_2021 | SS14 | Pop8 |
| ERR3684646 | PRJEB28546 | SAMEA5145895 | PHE170393A | UK | Europe | 2017 | Beale_2021 | SS14 | Pop8 |
| ERR3684647 | PRJEB28546 | SAMEA5145896 | PHE170403A | UK | Europe | 2017 | Beale_2021 | SS14 | Pop6 |
| ERR3684652 | PRJEB28546 | SAMEA5145903 | PHE120025A | UK | Europe | 2012 | Beale_2021 | SS14 | Pop8 |
| ERR3684653 | PRJEB28546 | SAMEA5145904 | PHE120027A | UK | Europe | 2012 | Beale_2021 | SS14 | Pop8 |
| ERR3684654 | PRJEB28546 | SAMEA5145905 | PHE120028A | UK | Europe | 2012 | Beale_2021 | SS14 | Pop8 |
| ERR3684655 | PRJEB28546 | SAMEA5145906 | PHE130038A | UK | Europe | 2013 | Beale_2021 | SS14 | Pop8 |
| ERR3684656 | PRJEB28546 | SAMEA5145907 | PHE130041A | UK | Europe | 2013 | Beale_2021 | SS14 | Pop8 |
| ERR3684657 | PRJEB28546 | SAMEA5145908 | PHE130046A | UK | Europe | 2013 | Beale_2021 | SS14 | Pop8 |
| ERR3684658 | PRJEB28546 | SAMEA5145909 | PHE130049A | UK | Europe | 2013 | Beale_2021 | SS14 | Pop8 |
| ERR3684659 | PRJEB28546 | SAMEA5145910 | PHE130050A | UK | Europe | 2013 | Beale_2021 | SS14 | Pop8 |
| ERR3684660 | PRJEB28546 | SAMEA5145911 | PHE140081A | UK | Europe | 2014 | Beale_2021 | SS14 | Pop6 |
| ERR3684661 | PRJEB28546 | SAMEA5145912 | PHE140083A | UK | Europe | 2014 | Beale_2021 | SS14 | Pop8 |
| ERR3684665 | PRJEB28546 | SAMEA5145916 | PHE150155A | UK | Europe | 2015 | Beale_2021 | SS14 | Pop8 |
| ERR3684666 | PRJEB28546 | SAMEA5145917 | PHE150159A | UK | Europe | 2015 | Beale_2021 | SS14 | Pop7 |
| ERR3684667 | PRJEB28546 | SAMEA5145918 | PHE150173A | UK | Europe | 2015 | Beale_2021 | SS14 | Pop8 |
| ERR3684669 | PRJEB28546 | SAMEA5145920 | PHE160184A | UK | Europe | 2016 | Beale_2021 | SS14 | Pop8 |
| ERR3684671 | PRJEB28546 | SAMEA5145922 | PHE160216A | UK | Europe | 2016 | Beale_2021 | SS14 | Pop8 |
| ERR3684672 | PRJEB28546 | SAMEA5145923 | PHE160222A | UK | Europe | 2016 | Beale_2021 | SS14 | Pop8 |
| ERR3684673 | PRJEB28546 | SAMEA5145924 | PHE160227A | UK | Europe | 2016 | Beale_2021 | SS14 | Pop8 |
| ERR3684676 | PRJEB28546 | SAMEA5145927 | PHE160291A | UK | Europe | 2016 | Beale_2021 | SS14 | Pop8 |
| ERR3684678 | PRJEB28546 | SAMEA5145929 | PHE160304A | UK | Europe | 2016 | Beale_2021 | SS14 | Pop8 |
| ERR3684680 | PRJEB28546 | SAMEA5145931 | PHE170320A | UK | Europe | 2017 | Beale_2021 | SS14 | Pop8 |
| ERR3684681 | PRJEB28546 | SAMEA5145932 | PHE170326A | UK | Europe | 2017 | Beale_2021 | SS14 | Pop8 |
| ERR3684682 | PRJEB28546 | SAMEA5145933 | PHE170344A | UK | Europe | 2017 | Beale_2021 | SS14 | Pop8 |
| ERR3684684 | PRJEB28546 | SAMEA5145935 | PHE170353A | UK | Europe | 2017 | Beale_2021 | SS14 | Pop7 |
| ERR3684685 | PRJEB28546 | SAMEA5145936 | PHE170358A | UK | Europe | 2017 | Beale_2021 | SS14 | Pop8 |
| ERR3684686 | PRJEB28546 | SAMEA5145937 | PHE170373A | UK | Europe | 2017 | Beale_2021 | SS14 | Pop8 |
| ERR3684687 | PRJEB28546 | SAMEA5145938 | PHE170392A | UK | Europe | 2017 | Beale_2021 | SS14 | Pop8 |
| ERR3684694 | PRJEB28546 | SAMEA5145950 | PHE120014A | UK | Europe | 2012 | Beale_2021 | SS14 | Pop8 |
| ERR3684695 | PRJEB28546 | SAMEA5145951 | PHE120016A | UK | Europe | 2012 | Beale_2021 | SS14 | Pop6 |

| | | | | | | | | | |
|---|---|---|---|---|---|---|---|---|---|
| ERR3684696 | PRJEB28546 | SAMEA5145952 | PHE120032A | UK | Europe | 2012 | Beale_2021 | SS14 | Pop8 |
| ERR3684697 | PRJEB28546 | SAMEA5145953 | PHE120034A | UK | Europe | 2012 | Beale_2021 | SS14 | Pop8 |
| ERR3684698 | PRJEB28546 | SAMEA5145954 | PHE130060A | UK | Europe | 2013 | Beale_2021 | SS14 | Pop8 |
| ERR3684700 | PRJEB28546 | SAMEA5145956 | PHE140084A | UK | Europe | 2014 | Beale_2021 | SS14 | Pop8 |
| ERR3684702 | PRJEB28546 | SAMEA5145958 | PHE140105A | UK | Europe | 2014 | Beale_2021 | SS14 | Pop8 |
| ERR3684703 | PRJEB28546 | SAMEA5145959 | PHE150123A | UK | Europe | 2015 | Beale_2021 | SS14 | Pop8 |
| ERR3684704 | PRJEB28546 | SAMEA5145960 | PHE150128A | UK | Europe | 2015 | Beale_2021 | SS14 | Pop8 |
| ERR3684706 | PRJEB28546 | SAMEA5145962 | PHE150164A | UK | Europe | 2015 | Beale_2021 | SS14 | Pop8 |
| ERR3684707 | PRJEB28546 | SAMEA5145963 | PHE150178A | UK | Europe | 2015 | Beale_2021 | SS14 | Pop8 |
| ERR3684708 | PRJEB28546 | SAMEA5145964 | PHE150179A | UK | Europe | 2015 | Beale_2021 | SS14 | Pop8 |
| ERR3684712 | PRJEB28546 | SAMEA5145968 | PHE160237A | UK | Europe | 2016 | Beale_2021 | SS14 | Pop8 |
| ERR3684713 | PRJEB28546 | SAMEA5145969 | PHE160242A | UK | Europe | 2016 | Beale_2021 | SS14 | Pop6 |
| ERR3684714 | PRJEB28546 | SAMEA5145970 | PHE160250A | UK | Europe | 2016 | Beale_2021 | SS14 | Pop8 |
| ERR3684722 | PRJEB28546 | SAMEA5145978 | PHE170335A | UK | Europe | 2017 | Beale_2021 | SS14 | Pop8 |
| ERR3684723 | PRJEB28546 | SAMEA5145979 | PHE170362A | UK | Europe | 2017 | Beale_2021 | SS14 | Pop8 |
| ERR3684724 | PRJEB28546 | SAMEA5145980 | PHE170369A | UK | Europe | 2017 | Beale_2021 | SS14 | Pop8 |
| ERR3684725 | PRJEB28546 | SAMEA5145981 | PHE170370A | UK | Europe | 2017 | Beale_2021 | SS14 | Pop6 |
| ERR3684727 | PRJEB28546 | SAMEA5145983 | PHE170391A | UK | Europe | 2017 | Beale_2021 | SS14 | Pop8 |
| ERR3684728 | PRJEB28546 | SAMEA5145984 | PHE170394A | UK | Europe | 2017 | Beale_2021 | SS14 | Pop8 |
| ERR3684739 | PRJEB28546 | SAMEA5146015 | PHE120003A | UK | Europe | 2012 | Beale_2021 | SS14 | Pop8 |
| ERR3684741 | PRJEB28546 | SAMEA5146017 | PHE120030A | UK | Europe | 2012 | Beale_2021 | SS14 | Pop8 |
| ERR3684742 | PRJEB28546 | SAMEA5146018 | PHE130063A | UK | Europe | 2013 | Beale_2021 | SS14 | Pop8 |
| ERR3684743 | PRJEB28546 | SAMEA5146019 | PHE140066A | UK | Europe | 2014 | Beale_2021 | SS14 | Pop8 |
| ERR3684744 | PRJEB28546 | SAMEA5146020 | PHE140077A | UK | Europe | 2014 | Beale_2021 | SS14 | Pop8 |
| ERR3684745 | PRJEB28546 | SAMEA5146021 | PHE140090A | UK | Europe | 2014 | Beale_2021 | SS14 | Pop8 |
| ERR3684746 | PRJEB28546 | SAMEA5146023 | PHE150125A | UK | Europe | 2015 | Beale_2021 | SS14 | Pop8 |
| ERR3684747 | PRJEB28546 | SAMEA5146024 | PHE150132A | UK | Europe | 2015 | Beale_2021 | SS14 | Pop8 |
| ERR3684748 | PRJEB28546 | SAMEA5146025 | PHE150151A | UK | Europe | 2015 | Beale_2021 | SS14 | Pop8 |
| ERR3684749 | PRJEB28546 | SAMEA5146026 | PHE150158A | UK | Europe | 2015 | Beale_2021 | SS14 | Pop8 |
| ERR3684750 | PRJEB28546 | SAMEA5146027 | PHE150168A | UK | Europe | 2015 | Beale_2021 | Nichols | Pop3 |
| ERR3684752 | PRJEB28546 | SAMEA5145995 | PHE150174A | UK | Europe | 2015 | Beale_2021 | SS14 | Pop8 |
| ERR3684753 | PRJEB28546 | SAMEA5145996 | PHE160192A | UK | Europe | 2016 | Beale_2021 | SS14 | Pop8 |
| ERR3699556 | PRJEB28546 | SAMEA5146045 | PHE120008A | UK | Europe | 2012 | Beale_2021 | SS14 | Pop8 |
| ERR3699557 | PRJEB28546 | SAMEA5146046 | PHE120012A | UK | Europe | 2012 | Beale_2021 | SS14 | Pop8 |
| ERR3699566 | PRJEB28546 | SAMEA5146055 | PHE140100A | UK | Europe | 2014 | Beale_2021 | SS14 | Pop8 |
| ERR3699567 | PRJEB28546 | SAMEA5146056 | PHE140101A | UK | Europe | 2014 | Beale_2021 | SS14 | Pop6 |
| ERR3699570 | PRJEB28546 | SAMEA5146059 | PHE150136A | UK | Europe | 2015 | Beale_2021 | SS14 | Pop8 |
| ERR3699576 | PRJEB28546 | SAMEA5146065 | PHE160202A | UK | Europe | 2016 | Beale_2021 | SS14 | Pop8 |
| ERR3811625 | PRJEB28546 | SAMEA5146094 | PHE130051A | UK | Europe | 2013 | Beale_2021 | SS14 | Pop8 |
| ERR3811628 | PRJEB28546 | SAMEA5146097 | PHE140072A | UK | Europe | 2014 | Beale_2021 | SS14 | Pop8 |
| ERR3811629 | PRJEB28546 | SAMEA5146098 | PHE140091A | UK | Europe | 2014 | Beale_2021 | SS14 | Pop8 |
| ERR3811630 | PRJEB28546 | SAMEA5146099 | PHE140094A | UK | Europe | 2014 | Beale_2021 | SS14 | Pop8 |
| ERR3811631 | PRJEB28546 | SAMEA5146100 | PHE150129A | UK | Europe | 2015 | Beale_2021 | SS14 | Pop8 |
| ERR3811643 | PRJEB28546 | SAMEA5146113 | PHE170395A | UK | Europe | 2017 | Beale_2021 | SS14 | Pop8 |

| | | | | | | | | | |
|---|---|---|---|---|---|---|---|---|---|
| ERR4045376 | PRJEB28546 | SAMEA5145789 | PHE160282A | UK | Europe | 2016 | Beale_2021 | Nichols | Pop2 |
| ERR4045377 | PRJEB28546 | SAMEA5145793 | PHE160302A | UK | Europe | 2016 | Beale_2021 | Nichols | Pop3 |
| ERR4045378 | PRJEB28546 | SAMEA5145809 | PHE170409A | UK | Europe | 2017 | Beale_2021 | SS14 | Pop8 |
| ERR4045379 | PRJEB28546 | SAMEA5145759 | PHE160276A | UK | Europe | 2016 | Beale_2021 | Nichols | Pop3 |
| ERR4045380 | PRJEB28546 | SAMEA5145762 | PHE160290A | UK | Europe | 2016 | Beale_2021 | Nichols | Pop3 |
| ERR4045381 | PRJEB28546 | SAMEA5145726 | PHE160306A | UK | Europe | 2016 | Beale_2021 | Nichols | Pop3 |
| ERR4045382 | PRJEB28546 | SAMEA5145766 | PHE140455A | UK | Europe | 2014 | Beale_2021 | Nichols | Pop3 |
| ERR4045383 | PRJEB28546 | SAMEA5145861 | PHE120033A | UK | Europe | 2012 | Beale_2021 | Nichols | Pop1 |
| ERR4045384 | PRJEB28546 | SAMEA5145863 | PHE140076A | UK | Europe | 2014 | Beale_2021 | Nichols | Pop3 |
| ERR4045385 | PRJEB28546 | SAMEA5145866 | PHE150118A | UK | Europe | 2015 | Beale_2021 | Nichols | Pop3 |
| ERR4045386 | PRJEB28546 | SAMEA5145878 | PHE160246A | UK | Europe | 2016 | Beale_2021 | SS14 | Pop8 |
| ERR4045387 | PRJEB28546 | SAMEA5145882 | PHE160265A | UK | Europe | 2016 | Beale_2021 | SS14 | Pop8 |
| ERR4045388 | PRJEB28546 | SAMEA5145794 | PHE160307A | UK | Europe | 2016 | Beale_2021 | Nichols | Pop3 |
| ERR4045389 | PRJEB28546 | SAMEA5145886 | PHE160295A | UK | Europe | 2016 | Beale_2021 | SS14 | Pop8 |
| ERR4045390 | PRJEB28546 | SAMEA5145894 | PHE170379A | UK | Europe | 2017 | Beale_2021 | SS14 | Pop8 |
| ERR4045391 | PRJEB28546 | SAMEA5145807 | PHE170401A | UK | Europe | 2017 | Beale_2021 | Nichols | Pop2 |
| ERR4045392 | PRJEB28546 | SAMEA5145900 | PHE160602A | UK | Europe | 2016 | Beale_2021 | SS14 | Pop6 |
| ERR4045393 | PRJEB28546 | SAMEA5145811 | PHE170664A | UK | Europe | 2017 | Beale_2021 | Nichols | Pop3 |
| ERR4045395 | PRJEB28546 | SAMEA5145826 | PHE150143A | UK | Europe | 2015 | Beale_2021 | Nichols | Pop3 |
| ERR4045396 | PRJEB28546 | SAMEA5145839 | PHE160271A | UK | Europe | 2016 | Beale_2021 | SS14 | Pop8 |
| ERR4045397 | PRJEB28546 | SAMEA5145843 | PHE170319A | UK | Europe | 2017 | Beale_2021 | SS14 | Pop8 |
| ERR4853527 | PRJEB33181 | SAMEA5753815 | TPA_BCC061 | Canada | N.America | 2012 | Beale_2021 | SS14 | Pop6 |
| ERR4853528 | PRJEB33181 | SAMEA5753816 | TPA_BCC075 | Canada | N.America | 2012 | Beale_2021 | SS14 | Pop7 |
| ERR4853529 | PRJEB33181 | SAMEA5753817 | TPA_BCC079 | Canada | N.America | 2012 | Beale_2021 | SS14 | Pop6 |
| ERR4853530 | PRJEB33181 | SAMEA5753818 | TPA_BCC081 | Canada | N.America | 2013 | Beale_2021 | Nichols | Pop3 |
| ERR4853531 | PRJEB33181 | SAMEA5753821 | TPA_BCC101 | Canada | N.America | 2014 | Beale_2021 | Nichols | Pop3 |
| ERR4853532 | PRJEB33181 | SAMEA5753830 | TPA_USL-SEA-81- | USA | N.America | 1981 | Beale_2021 | SS14 | Pop7 |
| ERR4853533 | PRJEB33181 | SAMEA5753840 | TPA_BCC008 | Canada | N.America | 2001 | Beale_2021 | SS14 | Pop7 |
| ERR4853534 | PRJEB33181 | SAMEA5753843 | TPA_BCC023 | Canada | N.America | 2003 | Beale_2021 | SS14 | Pop7 |
| ERR4853535 | PRJEB33181 | SAMEA5753844 | TPA_BCC049 | Canada | N.America | 2011 | Beale_2021 | SS14 | Pop6 |
| ERR4853536 | PRJEB33181 | SAMEA5753847 | TPA_BCC089 | Canada | N.America | 2013 | Beale_2021 | Nichols | Pop3 |
| ERR4853537 | PRJEB33181 | SAMEA5753849 | TPA_BCC109 | Canada | N.America | 2014 | Beale_2021 | SS14 | Pop6 |
| ERR4853538 | PRJEB33181 | SAMEA5753851 | TPA_BCC124 | Canada | N.America | 2015 | Beale_2021 | Nichols | Pop3 |
| ERR4853539 | PRJEB33181 | SAMEA5753852 | TPA_BCC126 | Canada | N.America | 2015 | Beale_2021 | Nichols | Pop3 |
| ERR4853540 | PRJEB33181 | SAMEA5753854 | TPA_BCC136 | Canada | N.America | 2015 | Beale_2021 | Nichols | Pop3 |
| ERR4853541 | PRJEB33181 | SAMEA5753855 | TPA_BCC139 | Canada | N.America | 2015 | Beale_2021 | SS14 | Pop7 |
| ERR4853542 | PRJEB33181 | SAMEA5753856 | TPA_BCC142 | Canada | N.America | 2016 | Beale_2021 | Nichols | Pop2 |
| ERR4853543 | PRJEB33181 | SAMEA5753857 | TPA_BCC154 | Canada | N.America | 2016 | Beale_2021 | SS14 | Pop6 |
| ERR4853544 | PRJEB33181 | SAMEA5753858 | TPA_BCC157 | Canada | N.America | 2016 | Beale_2021 | SS14 | Pop6 |
| ERR4853545 | PRJEB33181 | SAMEA5753859 | TPA_BCC163 | Canada | N.America | 2017 | Beale_2021 | SS14 | Pop6 |
| ERR4853546 | PRJEB33181 | SAMEA5753861 | TPA_BCC169 | Canada | N.America | 2017 | Beale_2021 | Nichols | Pop3 |
| ERR4853547 | PRJEB33181 | SAMEA5753862 | TPA_BCC187 | Canada | N.America | 2018 | Beale_2021 | SS14 | Pop6 |
| ERR4853548 | PRJEB33181 | SAMEA5753866 | TPA_ALC011 | Canada | N.America | 2018 | Beale_2021 | SS14 | Pop8 |
| ERR4853549 | PRJEB33181 | SAMEA5753868 | TPA_ALC023 | Canada | N.America | 2018 | Beale_2021 | SS14 | Pop8 |

| | | | | | | | | | |
|---|---|---|---|---|---|---|---|---|---|
| ERR4853550 | PRJEB33181 | SAMEA5753872 | TPA_BCC034 | Canada | N.America | 2005 | Beale_2021 | SS14 | Pop8 |
| ERR4853551 | PRJEB33181 | SAMEA5753878 | TPA_BCC064 | Canada | N.America | 2012 | Beale_2021 | SS14 | Pop8 |
| ERR4853552 | PRJEB33181 | SAMEA5753880 | TPA_BCC112 | Canada | N.America | 2014 | Beale_2021 | Nichols | Pop3 |
| ERR4853553 | PRJEB33181 | SAMEA5753882 | TPA_BCC130 | Canada | N.America | 2015 | Beale_2021 | SS14 | Pop8 |
| ERR4853554 | PRJEB33181 | SAMEA5753883 | TPA_BCC132 | Canada | N.America | 2015 | Beale_2021 | SS14 | Pop8 |
| ERR4853555 | PRJEB33181 | SAMEA5753884 | TPA_BCC141 | Canada | N.America | 2016 | Beale_2021 | SS14 | Pop6 |
| ERR4853556 | PRJEB33181 | SAMEA5753885 | TPA_BCC147 | Canada | N.America | 2016 | Beale_2021 | SS14 | Pop6 |
| ERR4853557 | PRJEB33181 | SAMEA5753888 | TPA_BCC158 | Canada | N.America | 2016 | Beale_2021 | Nichols | Pop3 |
| ERR4853558 | PRJEB33181 | SAMEA5753889 | TPA_BCC168 | Canada | N.America | 2017 | Beale_2021 | Nichols | Pop3 |
| ERR4853559 | PRJEB33181 | SAMEA5753890 | TPA_BCC174 | Canada | N.America | 2017 | Beale_2021 | SS14 | Pop6 |
| ERR4853560 | PRJEB33181 | SAMEA5753891 | TPA_BCC175 | Canada | N.America | 2017 | Beale_2021 | SS14 | Pop6 |
| ERR4853561 | PRJEB33181 | SAMEA5753892 | TPA_BCC181 | Canada | N.America | 2018 | Beale_2021 | SS14 | Pop6 |
| ERR4853562 | PRJEB33181 | SAMEA5753896 | TPA_BCC198 | Canada | N.America | 2018 | Beale_2021 | SS14 | Pop6 |
| ERR4853563 | PRJEB33181 | SAMEA5753899 | TPA_ALC005 | Canada | N.America | 2018 | Beale_2021 | SS14 | Pop8 |
| ERR4853564 | PRJEB33181 | SAMEA5753901 | TPA_ALC035 | Canada | N.America | 2018 | Beale_2021 | SS14 | Pop8 |
| ERR4853565 | PRJEB33181 | SAMEA5753812 | TPA_BCC014 | Canada | N.America | 2002 | Beale_2021 | SS14 | Pop7 |
| ERR4853566 | PRJEB33181 | SAMEA5753822 | TPA_BCC122 | Canada | N.America | 2015 | Beale_2021 | Nichols | Pop3 |
| ERR4853568 | PRJEB33181 | SAMEA5753826 | TPA_USL-BAL-6 | USA | N.America | 1950-1980 | Beale_2021 | Nichols | Pop1 |
| ERR4853569 | PRJEB33181 | SAMEA5753831 | TPA_USL-SEA-83- | USA | N.America | 1983 | Beale_2021 | Nichols | Pop1 |
| ERR4853571 | PRJEB33181 | SAMEA5753833 | TPA_USL-SEA-86- | USA | N.America | 1986 | Beale_2021 | Nichols | Pop2 |
| ERR4853572 | PRJEB33181 | SAMEA5753835 | TPA_USL-Haiti-B | Haiti | Caribbean | 1951 | Beale_2021 | Nichols | Pop1 |
| ERR4853573 | PRJEB33181 | SAMEA5753838 | TPA_USL-Phil-3 | USA | N.America | 1987 | Beale_2021 | Nichols | Pop1 |
| ERR4853575 | PRJEB33181 | SAMEA5857571 | TPA_BCC017 | Canada | N.America | 2002 | Beale_2021 | SS14 | Pop8 |
| ERR4853577 | PRJEB33181 | SAMEA5857617 | TPA_UKMAN047 | UK | Europe | 2018 | Beale_2021 | SS14 | Pop6 |
| ERR4853578 | PRJEB33181 | SAMEA5857618 | TPA_UKMAN049 | UK | Europe | 2018 | Beale_2021 | SS14 | Pop8 |
| ERR4853579 | PRJEB33181 | SAMEA5857619 | TPA_UKMAN054 | UK | Europe | 2018 | Beale_2021 | SS14 | Pop8 |
| ERR4853580 | PRJEB33181 | SAMEA5857620 | TPA_UKMAN003 | UK | Europe | 2018 | Beale_2021 | SS14 | Pop6 |
| ERR4853582 | PRJEB33181 | SAMEA5753845 | TPA_BCC055 | Canada | N.America | 2011 | Beale_2021 | SS14 | Pop8 |
| ERR4853583 | PRJEB33181 | SAMEA5753848 | TPA_BCC106 | Canada | N.America | 2014 | Beale_2021 | SS14 | Pop6 |
| ERR4853584 | PRJEB33181 | SAMEA5753853 | TPA_BCC127 | Canada | N.America | 2015 | Beale_2021 | SS14 | Pop6 |
| ERR4853585 | PRJEB33181 | SAMEA5753863 | TPA_BCC196 | Canada | N.America | 2018 | Beale_2021 | SS14 | Pop6 |
| ERR4853586 | PRJEB33181 | SAMEA5753864 | TPA_BCC199 | Canada | N.America | 2018 | Beale_2021 | SS14 | Pop6 |
| ERR4853587 | PRJEB33181 | SAMEA5753865 | TPA_EIR017 | Ireland | Europe | 2018 | Beale_2021 | SS14 | Pop6 |
| ERR4853588 | PRJEB33181 | SAMEA5753869 | TPA_ALC034 | Canada | N.America | 2018 | Beale_2021 | SS14 | Pop6 |
| ERR4853589 | PRJEB33181 | SAMEA5753870 | TPA_ALC055 | Canada | N.America | 2018 | Beale_2021 | SS14 | Pop8 |
| ERR4853590 | PRJEB33181 | SAMEA5753871 | TPA_BCC032 | Canada | N.America | 2004 | Beale_2021 | SS14 | Pop6 |
| ERR4853591 | PRJEB33181 | SAMEA5753873 | TPA_BCC038 | Canada | N.America | 2005 | Beale_2021 | SS14 | Pop8 |
| ERR4853592 | PRJEB33181 | SAMEA5753877 | TPA_BCC063 | Canada | N.America | 2012 | Beale_2021 | SS14 | Pop6 |
| ERR4853593 | PRJEB33181 | SAMEA5753879 | TPA_BCC102 | Canada | N.America | 2014 | Beale_2021 | SS14 | Pop6 |
| ERR4853594 | PRJEB33181 | SAMEA5753881 | TPA_BCC123 | Canada | N.America | 2015 | Beale_2021 | SS14 | Pop8 |
| ERR4853596 | PRJEB33181 | SAMEA5753893 | TPA_BCC185 | Canada | N.America | 2018 | Beale_2021 | SS14 | Pop6 |
| ERR4853597 | PRJEB33181 | SAMEA5753894 | TPA_BCC186 | Canada | N.America | 2018 | Beale_2021 | SS14 | Pop8 |
| ERR4853598 | PRJEB33181 | SAMEA5753895 | TPA_BCC197 | Canada | N.America | 2018 | Beale_2021 | SS14 | Pop6 |
| ERR4853599 | PRJEB33181 | SAMEA5753902 | TPA_ALC077 | Canada | N.America | 2018 | Beale_2021 | SS14 | Pop8 |

| | | | | | | | | | |
|---|---|---|---|---|---|---|---|---|---|
| ERR4853601 | PRJEB33181 | SAMEA5857542 | TPA_ZIM025 | Zimbabwe | Africa | 2015 | Beale_2021 | SS14 | Pop7 |
| ERR4853602 | PRJEB33181 | SAMEA5857502 | TPA_UKBRG012 | UK | Europe | 2018 | Beale_2021 | SS14 | Pop6 |
| ERR4853603 | PRJEB33181 | SAMEA5857503 | TPA_UKBRG007 | UK | Europe | 2018 | Beale_2021 | Nichols | Pop3 |
| ERR4853604 | PRJEB33181 | SAMEA5857504 | TPA_UKBRG010 | UK | Europe | 2018 | Beale_2021 | SS14 | Pop8 |
| ERR4853605 | PRJEB33181 | SAMEA5857506 | TPA_UKBRG017 | UK | Europe | 2018 | Beale_2021 | SS14 | Pop7 |
| ERR4853606 | PRJEB33181 | SAMEA5857507 | TPA_UKBRG009 | UK | Europe | 2018 | Beale_2021 | SS14 | Pop8 |
| ERR4853610 | PRJEB33181 | SAMEA5857515 | TPA_UKMAN019 | UK | Europe | 2018 | Beale_2021 | SS14 | Pop6 |
| ERR4853611 | PRJEB33181 | SAMEA5857516 | TPA_UKMAN027 | UK | Europe | 2018 | Beale_2021 | SS14 | Pop6 |
| ERR4853612 | PRJEB33181 | SAMEA5857519 | TPA_UKBIR033 | UK | Europe | 2018 | Beale_2021 | Nichols | Pop2 |
| ERR4853613 | PRJEB33181 | SAMEA5857521 | TPA_UKBIR032 | UK | Europe | 2018 | Beale_2021 | Nichols | Pop3 |
| ERR4853614 | PRJEB33181 | SAMEA5857523 | TPA_UKBIR052 | UK | Europe | 2018 | Beale_2021 | SS14 | Pop6 |
| ERR4853615 | PRJEB33181 | SAMEA5857524 | TPA_UKLEE004 | UK | Europe | 2018 | Beale_2021 | SS14 | Pop6 |
| ERR4853620 | PRJEB33181 | SAMEA5857535 | TPA_ZIM005 | Zimbabwe | Africa | 2015 | Beale_2021 | SS14 | Pop7 |
| ERR4853621 | PRJEB33181 | SAMEA5857537 | TPA_ZIM014 | Zimbabwe | Africa | 2015 | Beale_2021 | Nichols | Pop4 |
| ERR4853623 | PRJEB33181 | SAMEA5857540 | TPA_ZIM018 | Zimbabwe | Africa | 2015 | Beale_2021 | Nichols | Pop4 |
| ERR4853624 | PRJEB33181 | SAMEA5857541 | TPA_OMI017 | UK | Europe | 2017 | Beale_2021 | Nichols | Pop4 |
| ERR4853625 | PRJEB33181 | SAMEA5857543 | TPA_ZIM028 | Zimbabwe | Africa | 2015 | Beale_2021 | SS14 | Pop7 |
| ERR4853626 | PRJEB33181 | SAMEA5857548 | TPA_OMI061 | UK | Europe | 2018 | Beale_2021 | Nichols | Pop2 |
| ERR4853629 | PRJEB33181 | SAMEA5753876 | TPA_BCC058 | Canada | N.America | 2011 | Beale_2021 | SS14 | Pop8 |
| ERR4853631 | PRJEB33181 | SAMEA5857545 | TPA_OMI002 | UK | Europe | 2018 | Beale_2021 | SS14 | Pop7 |
| ERR4853633 | PRJEB33181 | SAMEA5857556 | TPA_ALC036 | Canada | N.America | 2018 | Beale_2021 | SS14 | Pop8 |
| ERR4853634 | PRJEB33181 | SAMEA5857559 | TPA_ALC048 | Canada | N.America | 2018 | Beale_2021 | SS14 | Pop8 |
| ERR4853635 | PRJEB33181 | SAMEA5857564 | TPA_ALC073 | Canada | N.America | 2018 | Beale_2021 | SS14 | Pop8 |
| ERR4853636 | PRJEB33181 | SAMEA5857565 | TPA_ALC076 | Canada | N.America | 2018 | Beale_2021 | SS14 | Pop8 |
| ERR4853640 | PRJEB33181 | SAMEA5857576 | TPA_BCC036 | Canada | N.America | 2005 | Beale_2021 | SS14 | Pop8 |
| ERR4853641 | PRJEB33181 | SAMEA5857579 | TPA_BCC052 | Canada | N.America | 2011 | Beale_2021 | SS14 | Pop8 |
| ERR4853643 | PRJEB33181 | SAMEA5857581 | TPA_BCC085 | Canada | N.America | 2013 | Beale_2021 | SS14 | Pop7 |
| ERR4853644 | PRJEB33181 | SAMEA5857582 | TPA_BCC107 | Canada | N.America | 2014 | Beale_2021 | SS14 | Pop6 |
| ERR4853645 | PRJEB33181 | SAMEA5857583 | TPA_BCC108 | Canada | N.America | 2014 | Beale_2021 | SS14 | Pop8 |
| ERR4853647 | PRJEB33181 | SAMEA5857613 | TPA_ESBCN004 | Spain | Europe | 2015 | Beale_2021 | SS14 | Pop8 |
| ERR4853648 | PRJEB33181 | SAMEA5857614 | TPA_ESBCN002 | Spain | Europe | 2015 | Beale_2021 | SS14 | Pop8 |
| ERR4853650 | PRJEB33181 | SAMEA5857616 | TPA_ESBCN001 | Spain | Europe | 2015 | Beale_2021 | SS14 | Pop8 |
| ERR4853651 | PRJEB33181 | SAMEA5753887 | TPA_BCC156 | Canada | N.America | 2016 | Beale_2021 | SS14 | Pop6 |
| ERR4853652 | PRJEB33181 | SAMEA5753897 | TPA_USL-SEA-87- | USA | N.America | 1987 | Beale_2021 | SS14 | Pop7 |
| ERR4853653 | PRJEB33181 | SAMEA5753898 | TPA_EIR013 | Ireland | Europe | 2018 | Beale_2021 | SS14 | Pop6 |
| ERR4853655 | PRJEB33181 | SAMEA5857554 | TPA_ALC009 | Canada | N.America | 2018 | Beale_2021 | SS14 | Pop8 |
| ERR4853657 | PRJEB33181 | SAMEA5857560 | TPA_ALC052 | Canada | N.America | 2018 | Beale_2021 | SS14 | Pop8 |
| ERR4853658 | PRJEB33181 | SAMEA5857561 | TPA_ALC053 | Canada | N.America | 2018 | Beale_2021 | SS14 | Pop8 |
| ERR4853659 | PRJEB33181 | SAMEA5857566 | TPA_ALC078 | Canada | N.America | 2018 | Beale_2021 | SS14 | Pop8 |
| ERR4853661 | PRJEB33181 | SAMEA5857585 | TPA_BCC125 | Canada | N.America | 2015 | Beale_2021 | SS14 | Pop8 |
| ERR4853662 | PRJEB33181 | SAMEA5857586 | TPA_BCC128 | Canada | N.America | 2015 | Beale_2021 | SS14 | Pop8 |
| ERR4853663 | PRJEB33181 | SAMEA5857587 | TPA_BCC129 | Canada | N.America | 2015 | Beale_2021 | SS14 | Pop6 |
| ERR4853664 | PRJEB33181 | SAMEA5857588 | TPA_BCC135 | Canada | N.America | 2015 | Beale_2021 | SS14 | Pop8 |
| ERR4853665 | PRJEB33181 | SAMEA5857589 | TPA_BCC137 | Canada | N.America | 2015 | Beale_2021 | SS14 | Pop8 |

| | | | | | | | | | |
|---|---|---|---|---|---|---|---|---|---|
| ERR4853666 | PRJEB33181 | SAMEA5857590 | TPA_BCC138 | Canada | N.America | 2015 | Beale_2021 | SS14 | Pop8 |
| ERR4853667 | PRJEB33181 | SAMEA5857591 | TPA_BCC140 | Canada | N.America | 2016 | Beale_2021 | SS14 | Pop8 |
| ERR4853668 | PRJEB33181 | SAMEA5857592 | TPA_BCC143 | Canada | N.America | 2016 | Beale_2021 | SS14 | Pop8 |
| ERR4853670 | PRJEB33181 | SAMEA5857594 | TPA_BCC145 | Canada | N.America | 2016 | Beale_2021 | SS14 | Pop8 |
| ERR4853671 | PRJEB33181 | SAMEA5857595 | TPA_BCC146 | Canada | N.America | 2016 | Beale_2021 | SS14 | Pop8 |
| ERR4853672 | PRJEB33181 | SAMEA5857596 | TPA_BCC153 | Canada | N.America | 2016 | Beale_2021 | SS14 | Pop6 |
| ERR4853673 | PRJEB33181 | SAMEA5857597 | TPA_BCC161 | Canada | N.America | 2017 | Beale_2021 | SS14 | Pop8 |
| ERR4853674 | PRJEB33181 | SAMEA5857598 | TPA_BCC166 | Canada | N.America | 2017 | Beale_2021 | SS14 | Pop8 |
| ERR4853675 | PRJEB33181 | SAMEA5857599 | TPA_BCC172 | Canada | N.America | 2017 | Beale_2021 | SS14 | Pop8 |
| ERR4853676 | PRJEB33181 | SAMEA5857600 | TPA_BCC176 | Canada | N.America | 2017 | Beale_2021 | SS14 | Pop6 |
| ERR4853677 | PRJEB33181 | SAMEA5857601 | TPA_BCC180 | Canada | N.America | 2018 | Beale_2021 | SS14 | Pop8 |
| ERR4853678 | PRJEB33181 | SAMEA5857602 | TPA_BCC182 | Canada | N.America | 2018 | Beale_2021 | SS14 | Pop8 |
| ERR4853679 | PRJEB33181 | SAMEA5857603 | TPA_BCC183 | Canada | N.America | 2018 | Beale_2021 | SS14 | Pop7 |
| ERR4853680 | PRJEB33181 | SAMEA5857605 | TPA_BCC191 | Canada | N.America | 2018 | Beale_2021 | SS14 | Pop8 |
| ERR4853681 | PRJEB33181 | SAMEA5857606 | TPA_BCC194 | Canada | N.America | 2018 | Beale_2021 | SS14 | Pop8 |
| ERR4853682 | PRJEB33181 | SAMEA5857607 | TPA_BCC195 | Canada | N.America | 2018 | Beale_2021 | SS14 | Pop8 |
| ERR4853683 | PRJEB33181 | SAMEA5857608 | TPA_UKBRG015 | UK | Europe | 2018 | Beale_2021 | SS14 | Pop8 |
| ERR4853684 | PRJEB33181 | SAMEA5857610 | TPA_EIR008 | Ireland | Europe | 2018 | Beale_2021 | SS14 | Pop6 |
| ERR4899197 | PRJEB33181 | SAMEA5753839 | TPA_BCC005 | Canada | N.America | 2000 | Beale_2021 | SS14 | Pop7 |
| ERR4899198 | PRJEB33181 | SAMEA5753841 | TPA_BCC009 | Canada | N.America | 2001 | Beale_2021 | SS14 | Pop7 |
| ERR4899199 | PRJEB33181 | SAMEA5753842 | TPA_BCC013 | Canada | N.America | 2002 | Beale_2021 | Nichols | Pop1 |
| ERR4899200 | PRJEB33181 | SAMEA5753850 | TPA_BCC111 | Canada | N.America | 2014 | Beale_2021 | SS14 | Pop6 |
| ERR4899201 | PRJEB33181 | SAMEA5753860 | TPA_BCC165 | Canada | N.America | 2017 | Beale_2021 | SS14 | Pop7 |
| ERR4899202 | PRJEB33181 | SAMEA5753867 | TPA_ALC015 | Canada | N.America | 2018 | Beale_2021 | SS14 | Pop6 |
| ERR4899203 | PRJEB33181 | SAMEA5857501 | TPA_UKBRG004 | UK | Europe | 2018 | Beale_2021 | SS14 | Pop8 |
| ERR4899204 | PRJEB33181 | SAMEA5857505 | TPA_UKBRG018 | UK | Europe | 2018 | Beale_2021 | SS14 | Pop7 |
| ERR4899206 | PRJEB33181 | SAMEA5857514 | TPA_ESBCN005 | Spain | Europe | 2015 | Beale_2021 | Nichols | Pop3 |
| ERR4899209 | PRJEB33181 | SAMEA5857520 | TPA_UKBIR028 | UK | Europe | 2018 | Beale_2021 | Nichols | Pop1 |
| ERR4899210 | PRJEB33181 | SAMEA5857522 | TPA_UKBIR044 | UK | Europe | 2018 | Beale_2021 | Nichols | Pop3 |
| ERR4899213 | PRJEB33181 | SAMEA5857536 | TPA_ZIM008 | Zimbabwe | Africa | 2015 | Beale_2021 | Nichols | Pop5 |
| ERR4899214 | PRJEB33181 | SAMEA5857538 | TPA_ZIM015 | Zimbabwe | Africa | 2015 | Beale_2021 | Nichols | Pop4 |
| ERR4899216 | PRJEB33181 | SAMEA5857546 | TPA_OMI015 | UK | Europe | 2017 | Beale_2021 | SS14 | Pop6 |
| ERR4899217 | PRJEB33181 | SAMEA5857547 | TPA_OMI021 | UK | Europe | 2017 | Beale_2021 | Nichols | Pop2 |
| ERR4899222 | PRJEB33181 | SAMEA5857577 | TPA_BCC044 | Canada | N.America | 2009 | Beale_2021 | SS14 | Pop8 |
| ERR4899223 | PRJEB33181 | SAMEA5857578 | TPA_BCC045 | Canada | N.America | 2010 | Beale_2021 | SS14 | Pop8 |
| ERR4899224 | PRJEB33181 | SAMEA5857623 | TPA_UKBIR026 | UK | Europe | 2018 | Beale_2021 | SS14 | Pop7 |
| ERR4899226 | PRJEB33181 | SAMEA5857625 | TPA_UKBIR043 | UK | Europe | 2018 | Beale_2021 | SS14 | Pop6 |
| ERR4899227 | PRJEB33181 | SAMEA5857627 | TPA_UKBIR049 | UK | Europe | 2018 | Beale_2021 | SS14 | Pop8 |
| ERR4899228 | PRJEB33181 | SAMEA5857628 | TPA_UKLEE006 | UK | Europe | 2018 | Beale_2021 | SS14 | Pop8 |
| ERR4899229 | PRJEB33181 | SAMEA5857629 | TPA_UKLEE012 | UK | Europe | 2018 | Beale_2021 | SS14 | Pop8 |
| ERR4899230 | PRJEB33181 | SAMEA5857630 | TPA_USL-Phil-1 | USA | N.America | 1987 | Beale_2021 | SS14 | Pop7 |
| ERR4899231 | PRJEB33181 | SAMEA5857631 | TPA_USL-Grady-1 | USA | N.America | 1994 | Beale_2021 | SS14 | Pop7 |
| ERR4899233 | PRJEB33181 | SAMEA5857636 | TPA_ZIM003 | Zimbabwe | Africa | 2015 | Beale_2021 | SS14 | Pop7 |
| ERR4899234 | PRJEB33181 | SAMEA5857637 | TPA_ZIM004 | Zimbabwe | Africa | 2015 | Beale_2021 | SS14 | Pop8 |

| | | | | | | | | | |
|---|---|---|---|---|---|---|---|---|---|
| ERR4899235 | PRJEB33181 | SAMEA5857638 | TPA_ZIM006 | Zimbabwe | Africa | 2015 | Beale_2021 | SS14 | Pop8 |
| ERR4899236 | PRJEB33181 | SAMEA5857639 | TPA_ZIM007 | Zimbabwe | Africa | 2015 | Beale_2021 | SS14 | Pop7 |
| ERR4899237 | PRJEB33181 | SAMEA5857640 | TPA_ZIM009 | Zimbabwe | Africa | 2015 | Beale_2021 | SS14 | Pop7 |
| ERR4899239 | PRJEB33181 | SAMEA5857642 | TPA_ZIM020 | Zimbabwe | Africa | 2015 | Beale_2021 | Nichols | Pop4 |
| ERR4899241 | PRJEB33181 | SAMEA5857644 | TPA_ZIM024 | Zimbabwe | Africa | 2015 | Beale_2021 | SS14 | Pop7 |
| ERR4899243 | PRJEB33181 | SAMEA5857646 | TPA_OMI075 | UK | Europe | 2017 | Beale_2021 | SS14 | Pop6 |
| ERR4899245 | PRJEB33181 | SAMEA5753828 | TPA_USL-BAL-8 | USA | N.America | 1960-1980 | Beale_2021 | Nichols | Pop1 |
| ERR4899246 | PRJEB33181 | SAMEA5753846 | TPA_BCC083 | Canada | N.America | 2013 | Beale_2021 | Nichols | Pop3 |
| ERR4993349 | PRJEB33181 | SAMEA5753809 | TPA_EIR015 | Ireland | Europe | 2018 | Beale_2021 | Nichols | Pop2 |
| ERR4993350 | PRJEB33181 | SAMEA5753810 | TPA_BCC004 | Canada | N.America | 2000 | Beale_2021 | SS14 | Pop7 |
| ERR4993351 | PRJEB33181 | SAMEA5753811 | TPA_BCC012 | Canada | N.America | 2002 | Beale_2021 | Nichols | Pop1 |
| ERR4993352 | PRJEB33181 | SAMEA5753813 | TPA_BCC030 | Canada | N.America | 2004 | Beale_2021 | SS14 | Pop6 |
| ERR4993353 | PRJEB33181 | SAMEA5753814 | TPA_BCC040 | Canada | N.America | 2006 | Beale_2021 | SS14 | Pop7 |
| ERR4993354 | PRJEB33181 | SAMEA5753819 | TPA_BCC088 | Canada | N.America | 2013 | Beale_2021 | Nichols | Pop3 |
| ERR4993355 | PRJEB33181 | SAMEA5753823 | TPA_BCC134 | Canada | N.America | 2015 | Beale_2021 | SS14 | Pop6 |
| ERR4993356 | PRJEB33181 | SAMEA5753824 | TPA_USL-BAL-2 | USA | N.America | 1971 | Beale_2021 | SS14 | Pop7 |
| ERR4993357 | PRJEB33181 | SAMEA5753827 | TPA_USL-BAL-7 | USA | N.America | 1976 | Beale_2021 | Nichols | Pop1 |
| ERR4993358 | PRJEB33181 | SAMEA5753829 | TPA_USL-SEA-81- | USA | N.America | 1981 | Beale_2021 | SS14 | Pop7 |
| ERR4993359 | PRJEB33181 | SAMEA5753834 | TPA_USL-SEA-84- | USA | N.America | 1984 | Beale_2021 | SS14 | Pop7 |
| ERR4993361 | PRJEB33181 | SAMEA5753837 | TPA_USL-Phil-2 | USA | N.America | 1987 | Beale_2021 | Nichols | Pop1 |
| ERR4993362 | PRJEB33181 | SAMEA5857457 | TPA_UKBRG008 | UK | Europe | 2018 | Beale_2021 | Nichols | Pop3 |
| ERR5206970 | PRJEB33181 | SAMEA6224016 | TPA_ALC115 | Canada | N.America | 2019 | Beale_2021 | SS14 | Pop8 |
| ERR5206972 | PRJEB33181 | SAMEA6224019 | TPA_ALC124 | Canada | N.America | 2019 | Beale_2021 | SS14 | Pop8 |
| ERR5206974 | PRJEB33181 | SAMEA6224021 | TPA_ALC126 | Canada | N.America | 2019 | Beale_2021 | SS14 | Pop6 |
| ERR5206976 | PRJEB33181 | SAMEA6224023 | TPA_ALC135 | Canada | N.America | 2019 | Beale_2021 | SS14 | Pop8 |
| ERR5206981 | PRJEB33181 | SAMEA6224029 | TPA_ALC151 | Canada | N.America | 2019 | Beale_2021 | SS14 | Pop8 |
| ERR5206984 | PRJEB33181 | SAMEA6224033 | TPA_ALC162 | Canada | N.America | 2019 | Beale_2021 | SS14 | Pop8 |
| ERR5206988 | PRJEB33181 | SAMEA6224040 | TPA_ALC180 | Canada | N.America | 2019 | Beale_2021 | SS14 | Pop8 |
| ERR5206989 | PRJEB33181 | SAMEA6224042 | TPA_ALC181 | Canada | N.America | 2019 | Beale_2021 | SS14 | Pop8 |
| ERR5206990 | PRJEB33181 | SAMEA6224044 | TPA_ALC105 | Canada | N.America | 2019 | Beale_2021 | SS14 | Pop8 |
| ERR5206991 | PRJEB33181 | SAMEA6224045 | TPA_ALC107 | Canada | N.America | 2019 | Beale_2021 | SS14 | Pop8 |
| ERR5206992 | PRJEB33181 | SAMEA6224051 | TPA_ALC123 | Canada | N.America | 2019 | Beale_2021 | SS14 | Pop8 |
| ERR5206993 | PRJEB33181 | SAMEA6224053 | TPA_ALC133 | Canada | N.America | 2019 | Beale_2021 | SS14 | Pop8 |
| ERR5206994 | PRJEB33181 | SAMEA6224055 | TPA_ALC136 | Canada | N.America | 2019 | Beale_2021 | SS14 | Pop8 |
| ERR5206995 | PRJEB33181 | SAMEA6224061 | TPA_ALC152 | Canada | N.America | 2019 | Beale_2021 | SS14 | Pop8 |
| ERR5207003 | PRJEB33181 | SAMEA6224092 | TPA_BCC103 | Canada | N.America | 2014 | Beale_2021 | SS14 | Pop6 |
| ERR5207005 | PRJEB33181 | SAMEA6224102 | TPA_UKBIR030 | UK | Europe | 2018 | Beale_2021 | SS14 | Pop8 |
| ERR5207006 | PRJEB33181 | SAMEA6224098 | TPA_UKBIR050 | UK | Europe | 2018 | Beale_2021 | SS14 | Pop8 |
| ERR5207007 | PRJEB33181 | SAMEA6224099 | TPA_ZIM019 | Zimbabwe | Africa | 2015 | Beale_2021 | SS14 | Pop7 |
| ERR5207008 | PRJEB33181 | SAMEA6224104 | TPA_OMI006 | UK | Europe | 2018 | Beale_2021 | SS14 | Pop7 |
| ERR5207009 | PRJEB33181 | SAMEA6224103 | TPA_OMI022 | UK | Europe | 2018 | Beale_2021 | SS14 | Pop6 |
| ERR5207010 | PRJEB33181 | SAMEA6224100 | TPA_OMI033 | UK | Europe | 2017 | Beale_2021 | SS14 | Pop8 |
| ERR5207011 | PRJEB33181 | SAMEA6224101 | TPA_OMI029 | UK | Europe | 2017 | Beale_2021 | SS14 | Pop6 |
| ERR5207013 | PRJEB33181 | SAMEA6626607 | TPA_HUN180001 | Hungary | Europe | 2018 | Beale_2021 | SS14 | Pop8 |

| | | | | | | | | | |
|---|---|---|---|---|---|---|---|---|---|
| ERR5207014 | PRJEB33181 | SAMEA6626610 | TPA_HUN190008 | Hungary | Europe | 2019 | Beale_2021 | SS14 | Pop8 |
| ERR5207015 | PRJEB33181 | SAMEA6626614 | TPA_HUN190017 | Hungary | Europe | 2019 | Beale_2021 | SS14 | Pop8 |
| ERR5207016 | PRJEB33181 | SAMEA6626616 | TPA_HUN190018 | Hungary | Europe | 2019 | Beale_2021 | SS14 | Pop8 |
| ERR5207017 | PRJEB33181 | SAMEA6626618 | TPA_HUN190020 | Hungary | Europe | 2019 | Beale_2021 | Nichols | Pop3 |
| ERR5207018 | PRJEB33181 | SAMEA6626620 | TPA_HUN200024 | Hungary | Europe | 2019 | Beale_2021 | SS14 | Pop8 |
| ERR5207019 | PRJEB33181 | SAMEA6626621 | TPA_RUS_Tuva-41 | Russia | Asia | 2014 | Beale_2021 | SS14 | Pop7 |
| ERR5207020 | PRJEB33181 | SAMEA6626624 | TPA_AUSBR-41 | Australia | Oceania | 2019 | Beale_2021 | Nichols | Pop2 |
| ERR5207022 | PRJEB33181 | SAMEA6626643 | TPA_HUN180006 | Hungary | Europe | 2018 | Beale_2021 | SS14 | Pop8 |
| ERR5207023 | PRJEB33181 | SAMEA6626646 | TPA_HUN190011 | Hungary | Europe | 2019 | Beale_2021 | SS14 | Pop8 |
| ERR5207025 | PRJEB33181 | SAMEA6626652 | TPA_HUN190022 | Hungary | Europe | 2019 | Beale_2021 | SS14 | Pop8 |
| ERR5207026 | PRJEB33181 | SAMEA6626654 | TPA_RUS_Tuva-39 | Russia | Asia | 2014 | Beale_2021 | SS14 | Pop7 |
| ERR5207027 | PRJEB33181 | SAMEA6626656 | TPA_RUS_Tuva-58 | Russia | Asia | 2014 | Beale_2021 | SS14 | Pop7 |
| ERR5207028 | PRJEB33181 | SAMEA6626658 | TPA_RUS_Tuva-67 | Russia | Asia | 2014 | Beale_2021 | SS14 | Pop8 |
| ERR5207029 | PRJEB33181 | SAMEA6626660 | TPA_RUS_Tuva-59 | Russia | Asia | 2014 | Beale_2021 | SS14 | Pop7 |
| ERR5207030 | PRJEB33181 | SAMEA6626662 | TPA_SWE-575 | Sweden | Europe | 2019 | Beale_2021 | SS14 | Pop8 |
| ERR5207031 | PRJEB33181 | SAMEA6626664 | TPA_SWE-996 | Sweden | Europe | 2019 | Beale_2021 | SS14 | Pop6 |
| ERR5207032 | PRJEB33181 | SAMEA6626666 | TPA_SWE-2189 | Sweden | Europe | 2019 | Beale_2021 | SS14 | Pop8 |
| ERR5207033 | PRJEB33181 | SAMEA6626669 | TPA_AUSBR-83 | Australia | Oceania | 2019 | Beale_2021 | SS14 | Pop8 |
| ERR5207034 | PRJEB33181 | SAMEA6626671 | TPA_RUS_Tuva-26 | Russia | Asia | 2013 | Beale_2021 | SS14 | Pop8 |
| ERR5210563 | PRJEB33181 | SAMEA6626609 | TPA_HUN180004 | Hungary | Europe | 2018 | Beale_2021 | Nichols | Pop3 |
| ERR5210564 | PRJEB33181 | SAMEA6626611 | TPA_HUN190009 | Hungary | Europe | 2019 | Beale_2021 | SS14 | Pop8 |
| ERR5210567 | PRJEB33181 | SAMEA6626617 | TPA_HUN190019 | Hungary | Europe | 2019 | Beale_2021 | Nichols | Pop3 |
| ERR5210568 | PRJEB33181 | SAMEA6626622 | TPA_RUS_Tuva-61 | Russia | Asia | 2014 | Beale_2021 | SS14 | Pop7 |
| ERR5210569 | PRJEB33181 | SAMEA6626623 | TPA_SWE-467 | Sweden | Europe | 2019 | Beale_2021 | SS14 | Pop6 |
| ERR5210570 | PRJEB33181 | SAMEA6626625 | TPA_AUSBR-45 | Australia | Oceania | 2019 | Beale_2021 | SS14 | Pop6 |
| ERR5210572 | PRJEB33181 | SAMEA6626644 | TPA_HUN180007 | Hungary | Europe | 2018 | Beale_2021 | SS14 | Pop6 |
| ERR5210574 | PRJEB33181 | SAMEA6626653 | TPA_HUN190023 | Hungary | Europe | 2019 | Beale_2021 | SS14 | Pop8 |
| ERR5210575 | PRJEB33181 | SAMEA6626655 | TPA_RUS_Tuva-49 | Russia | Asia | 2014 | Beale_2021 | SS14 | Pop7 |
| ERR5210576 | PRJEB33181 | SAMEA6626657 | TPA_RUS_Tuva-62 | Russia | Asia | 2014 | Beale_2021 | SS14 | Pop7 |
| ERR5210577 | PRJEB33181 | SAMEA6626659 | TPA_RUS_Tuva-54 | Russia | Asia | 2014 | Beale_2021 | SS14 | Pop7 |
| ERR5210578 | PRJEB33181 | SAMEA6626661 | TPA_SWE-507 | Sweden | Europe | 2019 | Beale_2021 | SS14 | Pop8 |
| ERR5210579 | PRJEB33181 | SAMEA6626663 | TPA_SWE-662 | Sweden | Europe | 2019 | Beale_2021 | SS14 | Pop6 |
| ERR5210580 | PRJEB33181 | SAMEA6626665 | TPA_SWE-1352 | Sweden | Europe | 2019 | Beale_2021 | SS14 | Pop8 |
| ERR5210581 | PRJEB33181 | SAMEA6626667 | TPA_AUSBR-39 | Australia | Oceania | 2019 | Beale_2021 | SS14 | Pop6 |
| ERR5210582 | PRJEB33181 | SAMEA6626670 | TPA_AUSBR-113 | Australia | Oceania | 2019 | Beale_2021 | SS14 | Pop8 |
| SRR10041768 | PRJNA562373 | SAMN12638760 | SMUTp_08 | China | Asia | 2018 | Chen_2021 | SS14 | Pop8 |
| SRR10041769 | PRJNA562373 | SAMN12638759 | SMUTp_07 | China | Asia | 2018 | Chen_2021 | Nichols | Pop3 |
| SRR10041772 | PRJNA562373 | SAMN12638756 | SMUTp_04 | China | Asia | 2018 | Chen_2021 | SS14 | Pop7 |
| SRR10041774 | PRJNA562373 | SAMN12638754 | SMUTp_02 | China | Asia | 2017 | Chen_2021 | SS14 | Pop7 |
| SRR10041775 | PRJNA562373 | SAMN12638753 | SMUTp_01 | China | Asia | 2018 | Chen_2021 | SS14 | Pop7 |
| SRR14277265 | PRJNA723099 | SAMN18804944 | Japan323x | Japan | Asia | 2019 | Lieberman_2022 | SS14 | Pop6 |
| SRR14277268 | PRJNA723099 | SAMN18804941 | Japan319 | Japan | Asia | 2019 | Lieberman_2022 | SS14 | Pop7 |
| SRR14277269 | PRJNA723099 | SAMN18804940 | Japan318 | Japan | Asia | 2019 | Lieberman_2022 | SS14 | Pop7 |
| SRR14277270 | PRJNA723099 | SAMN18804939 | Japan317xi | Japan | Asia | 2019 | Lieberman_2022 | Nichols | Pop3 |

| | | | | | | | | | |
|---|---|---|---|---|---|---|---|---|---|
| SRR14277271 | PRJNA723099 | SAMN18804894 | China44 | China | Asia | 2018 | Lieberman_2022 | SS14 | Pop7 |
| SRR14277272 | PRJNA723099 | SAMN18804938 | Japan316x | Japan | Asia | 2019 | Lieberman_2022 | SS14 | Pop8 |
| SRR14277273 | PRJNA723099 | SAMN18804937 | Japan314 | Japan | Asia | 2019 | Lieberman_2022 | SS14 | Pop6 |
| SRR14277274 | PRJNA723099 | SAMN18804936 | Japan313x | Japan | Asia | 2019 | Lieberman_2022 | SS14 | Pop7 |
| SRR14277275 | PRJNA723099 | SAMN18804935 | Japan312xe | Japan | Asia | 2019 | Lieberman_2022 | SS14 | Pop7 |
| SRR14277276 | PRJNA723099 | SAMN18804934 | Japan311 | Japan | Asia | 2019 | Lieberman_2022 | SS14 | Pop7 |
| SRR14277277 | PRJNA723099 | SAMN18804933 | Japan309x | Japan | Asia | 2019 | Lieberman_2022 | SS14 | Pop7 |
| SRR14277278 | PRJNA723099 | SAMN18804932 | Japan308 | Japan | Asia | 2019 | Lieberman_2022 | SS14 | Pop7 |
| SRR14277279 | PRJNA723099 | SAMN18804931 | Japan307 | Japan | Asia | 2019 | Lieberman_2022 | SS14 | Pop6 |
| SRR14277280 | PRJNA723099 | SAMN18804930 | Japan306 | Japan | Asia | 2019 | Lieberman_2022 | SS14 | Pop7 |
| SRR14277281 | PRJNA723099 | SAMN18804929 | Japan305 | Japan | Asia | 2019 | Lieberman_2022 | SS14 | Pop7 |
| SRR14277282 | PRJNA723099 | SAMN18804893 | China42 | China | Asia | 2018 | Lieberman_2022 | SS14 | Pop7 |
| SRR14277283 | PRJNA723099 | SAMN18804928 | Japan303 | Japan | Asia | 2019 | Lieberman_2022 | Nichols | Pop2 |
| SRR14277284 | PRJNA723099 | SAMN18804927 | Japan302x | Japan | Asia | 2019 | Lieberman_2022 | SS14 | Pop7 |
| SRR14277285 | PRJNA723099 | SAMN18804926 | Japan301 | Japan | Asia | 2019 | Lieberman_2022 | SS14 | Pop7 |
| SRR14277286 | PRJNA723099 | SAMN18804925 | Japan300 | Japan | Asia | 2019 | Lieberman_2022 | SS14 | Pop6 |
| SRR14277287 | PRJNA723099 | SAMN18804924 | Japan298x | Japan | Asia | 2019 | Lieberman_2022 | SS14 | Pop7 |
| SRR14277288 | PRJNA723099 | SAMN18804923 | Japan295x | Japan | Asia | 2019 | Lieberman_2022 | SS14 | Pop7 |
| SRR14277289 | PRJNA723099 | SAMN18804922 | Japan294x | Japan | Asia | 2019 | Lieberman_2022 | SS14 | Pop7 |
| SRR14277290 | PRJNA723099 | SAMN18804921 | Japan291x | Japan | Asia | 2019 | Lieberman_2022 | SS14 | Pop7 |
| SRR14277291 | PRJNA723099 | SAMN18804920 | Japan290 | Japan | Asia | 2019 | Lieberman_2022 | SS14 | Pop7 |
| SRR14277292 | PRJNA723099 | SAMN18804919 | Japan288 | Japan | Asia | 2019 | Lieberman_2022 | SS14 | Pop7 |
| SRR14277293 | PRJNA723099 | SAMN18804892 | China31 | China | Asia | 2018 | Lieberman_2022 | SS14 | Pop7 |
| SRR14277294 | PRJNA723099 | SAMN18804918 | Japan287e | Japan | Asia | 2019 | Lieberman_2022 | SS14 | Pop7 |
| SRR14277295 | PRJNA723099 | SAMN18804917 | ItalyGE6 | Italy | Europe | 2017 | Lieberman_2022 | SS14 | Pop7 |
| SRR14277296 | PRJNA723099 | SAMN18804916 | ItalyGE5b | Italy | Europe | 2017 | Lieberman_2022 | SS14 | Pop6 |
| SRR14277297 | PRJNA723099 | SAMN18804915 | ItalyGE5a | Italy | Europe | 2017 | Lieberman_2022 | SS14 | Pop6 |
| SRR14277298 | PRJNA723099 | SAMN18804914 | ItalyGE4b | Italy | Europe | 2017 | Lieberman_2022 | SS14 | Pop8 |
| SRR14277299 | PRJNA723099 | SAMN18804913 | ItalyAS23 | Italy | Europe | 2017 | Lieberman_2022 | SS14 | Pop6 |
| SRR14277300 | PRJNA723099 | SAMN18804912 | ItalyAS15 | Italy | Europe | 2017 | Lieberman_2022 | SS14 | Pop6 |
| SRR14277301 | PRJNA723099 | SAMN18804911 | ItalyAS14 | Italy | Europe | 2017 | Lieberman_2022 | Nichols | Pop2 |
| SRR14277302 | PRJNA723099 | SAMN18804910 | Italy8PLe | Italy | Europe | 2017 | Lieberman_2022 | SS14 | Pop6 |
| SRR14277303 | PRJNA723099 | SAMN18804909 | Italy7CGx | Italy | Europe | 2017 | Lieberman_2022 | SS14 | Pop6 |
| SRR14277304 | PRJNA723099 | SAMN18804891 | China30 | China | Asia | 2018 | Lieberman_2022 | SS14 | Pop7 |
| SRR14277305 | PRJNA723099 | SAMN18805084 | UAB89xi | Madagascar | Africa | 2001 | Lieberman_2022 | Nichols | Pop4 |
| SRR14277306 | PRJNA723099 | SAMN18805083 | UAB81xi | Madagascar | Africa | 2001 | Lieberman_2022 | Nichols | Pop4 |
| SRR14277307 | PRJNA723099 | SAMN18805082 | UAB76xei | Madagascar | Africa | 2001 | Lieberman_2022 | Nichols | Pop4 |
| SRR14277308 | PRJNA723099 | SAMN18805081 | UAB75 | Madagascar | Africa | 2001 | Lieberman_2022 | Nichols | Pop5 |
| SRR14277309 | PRJNA723099 | SAMN18805080 | UAB71i | Madagascar | Africa | 2001 | Lieberman_2022 | Nichols | Pop5 |
| SRR14277310 | PRJNA723099 | SAMN18805079 | UAB6xi | Madagascar | Africa | 2001 | Lieberman_2022 | Nichols | Pop5 |
| SRR14277311 | PRJNA723099 | SAMN18804908 | Italy17LGMi | Italy | Europe | 2017 | Lieberman_2022 | Nichols | Pop3 |
| SRR14277312 | PRJNA723099 | SAMN18805078 | UAB69i | Madagascar | Africa | 2001 | Lieberman_2022 | Nichols | Pop5 |
| SRR14277313 | PRJNA723099 | SAMN18805077 | UAB67xi | Madagascar | Africa | 2001 | Lieberman_2022 | Nichols | Pop4 |
| SRR14277314 | PRJNA723099 | SAMN18805076 | UAB62i | Madagascar | Africa | 2001 | Lieberman_2022 | Nichols | Pop5 |

| | | | | | | | | | |
|---|---|---|---|---|---|---|---|---|---|
| SRR14277315 | PRJNA723099 | SAMN18805075 | UAB61xei | Madagascar | Africa | 2001 | Lieberman_2022 | Nichols | Pop5 |
| SRR14277316 | PRJNA723099 | SAMN18805074 | UAB60i | Madagascar | Africa | 2001 | Lieberman_2022 | Nichols | Pop5 |
| SRR14277317 | PRJNA723099 | SAMN18805073 | UAB554xi | Madagascar | Africa | 2007 | Lieberman_2022 | Nichols | Pop5 |
| SRR14277318 | PRJNA723099 | SAMN18805072 | UAB553xei | Madagascar | Africa | 2007 | Lieberman_2022 | Nichols | Pop5 |
| SRR14277319 | PRJNA723099 | SAMN18805071 | UAB552x | Madagascar | Africa | 2007 | Lieberman_2022 | Nichols | Pop5 |
| SRR14277320 | PRJNA723099 | SAMN18805070 | UAB551xi | Madagascar | Africa | 2007 | Lieberman_2022 | Nichols | Pop5 |
| SRR14277321 | PRJNA723099 | SAMN18805069 | UAB54xi | Madagascar | Africa | 2001 | Lieberman_2022 | Nichols | Pop4 |
| SRR14277322 | PRJNA723099 | SAMN18804907 | Dublin58B | Ireland | Europe | 2002 | Lieberman_2022 | SS14 | Pop6 |
| SRR14277323 | PRJNA723099 | SAMN18805068 | UAB549xe | Madagascar | Africa | 2007 | Lieberman_2022 | Nichols | Pop4 |
| SRR14277324 | PRJNA723099 | SAMN18805067 | UAB547xei | Madagascar | Africa | 2007 | Lieberman_2022 | Nichols | Pop4 |
| SRR14277325 | PRJNA723099 | SAMN18805066 | UAB546i | Madagascar | Africa | 2007 | Lieberman_2022 | Nichols | Pop4 |
| SRR14277326 | PRJNA723099 | SAMN18805065 | UAB545ei | Madagascar | Africa | 2006 | Lieberman_2022 | Nichols | Pop5 |
| SRR14277327 | PRJNA723099 | SAMN18805064 | UAB538xi | Madagascar | Africa | 2007 | Lieberman_2022 | Nichols | Pop4 |
| SRR14277328 | PRJNA723099 | SAMN18805063 | UAB535i | Madagascar | Africa | 2006 | Lieberman_2022 | Nichols | Pop4 |
| SRR14277329 | PRJNA723099 | SAMN18805062 | UAB534xi | Madagascar | Africa | 2006 | Lieberman_2022 | Nichols | Pop5 |
| SRR14277330 | PRJNA723099 | SAMN18805061 | UAB533i | Madagascar | Africa | 2006 | Lieberman_2022 | Nichols | Pop5 |
| SRR14277331 | PRJNA723099 | SAMN18805060 | UAB532xi | Madagascar | Africa | 2006 | Lieberman_2022 | Nichols | Pop4 |
| SRR14277332 | PRJNA723099 | SAMN18805059 | UAB531i | Madagascar | Africa | 2006 | Lieberman_2022 | Nichols | Pop5 |
| SRR14277333 | PRJNA723099 | SAMN18804906 | Dublin57B | Ireland | Europe | 2002 | Lieberman_2022 | SS14 | Pop7 |
| SRR14277334 | PRJNA723099 | SAMN18805058 | UAB530i | Madagascar | Africa | 2006 | Lieberman_2022 | Nichols | Pop4 |
| SRR14277335 | PRJNA723099 | SAMN18805057 | UAB52xi | Madagascar | Africa | 2001 | Lieberman_2022 | Nichols | Pop5 |
| SRR14277336 | PRJNA723099 | SAMN18805056 | UAB529i | Madagascar | Africa | 2006 | Lieberman_2022 | Nichols | Pop4 |
| SRR14277337 | PRJNA723099 | SAMN18805055 | UAB528ei | Madagascar | Africa | 2006 | Lieberman_2022 | Nichols | Pop4 |
| SRR14277338 | PRJNA723099 | SAMN18805054 | UAB526xi | Madagascar | Africa | 2006 | Lieberman_2022 | Nichols | Pop5 |
| SRR14277339 | PRJNA723099 | SAMN18805053 | UAB525xe | Madagascar | Africa | 2006 | Lieberman_2022 | Nichols | Pop5 |
| SRR14277340 | PRJNA723099 | SAMN18805052 | UAB523x | Madagascar | Africa | 2006 | Lieberman_2022 | Nichols | Pop5 |
| SRR14277342 | PRJNA723099 | SAMN18805050 | UAB521xi | Madagascar | Africa | 2006 | Lieberman_2022 | Nichols | Pop5 |
| SRR14277343 | PRJNA723099 | SAMN18805049 | UAB516xi | Madagascar | Africa | 2006 | Lieberman_2022 | Nichols | Pop5 |
| SRR14277344 | PRJNA723099 | SAMN18804905 | Dublin55B | Ireland | Europe | 2002 | Lieberman_2022 | SS14 | Pop6 |
| SRR14277345 | PRJNA723099 | SAMN18805048 | UAB515xi | Madagascar | Africa | 2006 | Lieberman_2022 | Nichols | Pop5 |
| SRR14277346 | PRJNA723099 | SAMN18805047 | UAB512x | Madagascar | Africa | 2006 | Lieberman_2022 | Nichols | Pop4 |
| SRR14277347 | PRJNA723099 | SAMN18805046 | UAB511xi | Madagascar | Africa | 2006 | Lieberman_2022 | Nichols | Pop5 |
| SRR14277348 | PRJNA723099 | SAMN18805045 | UAB510i | Madagascar | Africa | 2006 | Lieberman_2022 | Nichols | Pop5 |
| SRR14277349 | PRJNA723099 | SAMN18805044 | UAB509ei | Madagascar | Africa | 2006 | Lieberman_2022 | Nichols | Pop4 |
| SRR14277350 | PRJNA723099 | SAMN18805043 | UAB508 | Madagascar | Africa | 2006 | Lieberman_2022 | Nichols | Pop5 |
| SRR14277351 | PRJNA723099 | SAMN18805042 | UAB507ei | Madagascar | Africa | 2006 | Lieberman_2022 | Nichols | Pop5 |
| SRR14277352 | PRJNA723099 | SAMN18805041 | UAB505ei | Madagascar | Africa | 2006 | Lieberman_2022 | Nichols | Pop5 |
| SRR14277353 | PRJNA723099 | SAMN18805040 | UAB504xi | Madagascar | Africa | 2006 | Lieberman_2022 | Nichols | Pop5 |
| SRR14277354 | PRJNA723099 | SAMN18805039 | UAB503i | Madagascar | Africa | 2006 | Lieberman_2022 | Nichols | Pop5 |
| SRR14277355 | PRJNA723099 | SAMN18804904 | Dublin54B | Ireland | Europe | 2002 | Lieberman_2022 | SS14 | Pop6 |
| SRR14277356 | PRJNA723099 | SAMN18805038 | UAB501 | Madagascar | Africa | 2003 | Lieberman_2022 | Nichols | Pop5 |
| SRR14277357 | PRJNA723099 | SAMN18805037 | UAB49xi | Madagascar | Africa | 2000 | Lieberman_2022 | Nichols | Pop4 |
| SRR14277358 | PRJNA723099 | SAMN18805036 | UAB46xei | Madagascar | Africa | 2000 | Lieberman_2022 | Nichols | Pop4 |
| SRR14277359 | PRJNA723099 | SAMN18805035 | UAB44xi | Madagascar | Africa | 2000 | Lieberman_2022 | Nichols | Pop4 |

| | | | | | | | | | |
|---|---|---|---|---|---|---|---|---|---|
| SRR14277360 | PRJNA723099 | SAMN18805034 | UAB40xi | Madagascar | Africa | 2000 | Lieberman_2022 | Nichols | Pop4 |
| SRR14277361 | PRJNA723099 | SAMN18805033 | UAB38xi | Madagascar | Africa | 2000 | Lieberman_2022 | Nichols | Pop5 |
| SRR14277362 | PRJNA723099 | SAMN18805032 | UAB28xi | Madagascar | Africa | 2000 | Lieberman_2022 | Nichols | Pop5 |
| SRR14277363 | PRJNA723099 | SAMN18805031 | UAB278i | Madagascar | Africa | 2003 | Lieberman_2022 | Nichols | Pop5 |
| SRR14277364 | PRJNA723099 | SAMN18805030 | UAB276i | Madagascar | Africa | 2003 | Lieberman_2022 | Nichols | Pop5 |
| SRR14277365 | PRJNA723099 | SAMN18805029 | UAB275i | Madagascar | Africa | 2003 | Lieberman_2022 | Nichols | Pop5 |
| SRR14277366 | PRJNA723099 | SAMN18804903 | Dublin52B | Ireland | Europe | 2002 | Lieberman_2022 | SS14 | Pop6 |
| SRR14277367 | PRJNA723099 | SAMN18805028 | UAB268xi | Madagascar | Africa | 2003 | Lieberman_2022 | Nichols | Pop5 |
| SRR14277368 | PRJNA723099 | SAMN18805027 | UAB267i | Madagascar | Africa | 2003 | Lieberman_2022 | Nichols | Pop5 |
| SRR14277369 | PRJNA723099 | SAMN18805026 | UAB266i | Madagascar | Africa | 2003 | Lieberman_2022 | Nichols | Pop5 |
| SRR14277370 | PRJNA723099 | SAMN18805025 | UAB263xi | Madagascar | Africa | 2003 | Lieberman_2022 | Nichols | Pop5 |
| SRR14277371 | PRJNA723099 | SAMN18805024 | UAB237 | Madagascar | Africa | 2003 | Lieberman_2022 | Nichols | Pop5 |
| SRR14277372 | PRJNA723099 | SAMN18805023 | UAB236i | Madagascar | Africa | 2003 | Lieberman_2022 | Nichols | Pop4 |
| SRR14277373 | PRJNA723099 | SAMN18805022 | UAB231i | Madagascar | Africa | 2003 | Lieberman_2022 | Nichols | Pop4 |
| SRR14277374 | PRJNA723099 | SAMN18805021 | UAB229xe | Madagascar | Africa | 2003 | Lieberman_2022 | Nichols | Pop5 |
| SRR14277375 | PRJNA723099 | SAMN18805020 | UAB218i | Madagascar | Africa | 2003 | Lieberman_2022 | Nichols | Pop5 |
| SRR14277376 | PRJNA723099 | SAMN18805019 | UAB216i | Madagascar | Africa | 2003 | Lieberman_2022 | Nichols | Pop5 |
| SRR14277377 | PRJNA723099 | SAMN18804902 | Dublin50B | Ireland | Europe | 2002 | Lieberman_2022 | SS14 | Pop6 |
| SRR14277378 | PRJNA723099 | SAMN18805018 | UAB211i | Madagascar | Africa | 2002 | Lieberman_2022 | Nichols | Pop5 |
| SRR14277379 | PRJNA723099 | SAMN18805017 | UAB204xe | Madagascar | Africa | 2002 | Lieberman_2022 | Nichols | Pop5 |
| SRR14277380 | PRJNA723099 | SAMN18805016 | UAB203i | Madagascar | Africa | 2002 | Lieberman_2022 | Nichols | Pop4 |
| SRR14277381 | PRJNA723099 | SAMN18805015 | UAB202xi | Madagascar | Africa | 2002 | Lieberman_2022 | Nichols | Pop4 |
| SRR14277382 | PRJNA723099 | SAMN18805014 | UAB197i | Madagascar | Africa | 2002 | Lieberman_2022 | Nichols | Pop4 |
| SRR14277383 | PRJNA723099 | SAMN18805013 | UAB195xi | Madagascar | Africa | 2002 | Lieberman_2022 | Nichols | Pop4 |
| SRR14277384 | PRJNA723099 | SAMN18805012 | UAB194xi | Madagascar | Africa | 2002 | Lieberman_2022 | Nichols | Pop4 |
| SRR14277385 | PRJNA723099 | SAMN18805011 | UAB193i | Madagascar | Africa | 2002 | Lieberman_2022 | Nichols | Pop4 |
| SRR14277386 | PRJNA723099 | SAMN18805010 | UAB191xei | Madagascar | Africa | 2002 | Lieberman_2022 | Nichols | Pop5 |
| SRR14277387 | PRJNA723099 | SAMN18805009 | UAB17x | Madagascar | Africa | 2000 | Lieberman_2022 | Nichols | Pop5 |
| SRR14277388 | PRJNA723099 | SAMN18804901 | Dublin46B | Ireland | Europe | 2002 | Lieberman_2022 | SS14 | Pop6 |
| SRR14277389 | PRJNA723099 | SAMN18805008 | UAB178ei | Madagascar | Africa | 2002 | Lieberman_2022 | Nichols | Pop5 |
| SRR14277390 | PRJNA723099 | SAMN18805007 | UAB16xi | Madagascar | Africa | 2000 | Lieberman_2022 | Nichols | Pop5 |
| SRR14277391 | PRJNA723099 | SAMN18805006 | UAB167xei | Madagascar | Africa | 2002 | Lieberman_2022 | Nichols | Pop4 |
| SRR14277393 | PRJNA723099 | SAMN18805004 | UAB165xi | Madagascar | Africa | 2002 | Lieberman_2022 | Nichols | Pop5 |
| SRR14277394 | PRJNA723099 | SAMN18805003 | UAB164xei | Madagascar | Africa | 2002 | Lieberman_2022 | Nichols | Pop5 |
| SRR14277395 | PRJNA723099 | SAMN18805002 | UAB162xi | Madagascar | Africa | 2002 | Lieberman_2022 | Nichols | Pop4 |
| SRR14277396 | PRJNA723099 | SAMN18805001 | UAB160xe | Madagascar | Africa | 2001 | Lieberman_2022 | Nichols | Pop5 |
| SRR14277397 | PRJNA723099 | SAMN18805000 | UAB12xi | Madagascar | Africa | 2000 | Lieberman_2022 | Nichols | Pop4 |
| SRR14277399 | PRJNA723099 | SAMN18804900 | Dublin41B | Ireland | Europe | 2002 | Lieberman_2022 | SS14 | Pop6 |
| SRR14277400 | PRJNA723099 | SAMN18804998 | PeruC40040x | Peru | S.America | 2019 | Lieberman_2022 | SS14 | Pop6 |
| SRR14277401 | PRJNA723099 | SAMN18804997 | PeruC40020xe | Peru | S.America | 2019 | Lieberman_2022 | SS14 | Pop8 |
| SRR14277402 | PRJNA723099 | SAMN18804996 | PeruC10050xe | Peru | S.America | 2019 | Lieberman_2022 | SS14 | Pop8 |
| SRR14277403 | PRJNA723099 | SAMN18804995 | PeruC10010x | Peru | S.America | 2019 | Lieberman_2022 | SS14 | Pop6 |
| SRR14277404 | PRJNA723099 | SAMN18804994 | PeruB20010x | Peru | S.America | 2019 | Lieberman_2022 | SS14 | Pop6 |
| SRR14277405 | PRJNA723099 | SAMN18804993 | Peru214601x | Peru | S.America | 2018 | Lieberman_2022 | SS14 | Pop6 |

| | | | | | | | | | |
|---|---|---|---|---|---|---|---|---|---|
| SRR14277406 | PRJNA723099 | SAMN18804992 | Peru214361x | Peru | S.America | 2018 | Lieberman_2022 | SS14 | Pop7 |
| SRR14277407 | PRJNA723099 | SAMN18804991 | Peru213161xe | Peru | S.America | 2018 | Lieberman_2022 | SS14 | Pop8 |
| SRR14277408 | PRJNA723099 | SAMN18804990 | Peru213041xe | Peru | S.America | 2018 | Lieberman_2022 | SS14 | Pop8 |
| SRR14277409 | PRJNA723099 | SAMN18804989 | MD57x | USA | N.America | 2000 | Lieberman_2022 | SS14 | Pop7 |
| SRR14277410 | PRJNA723099 | SAMN18804899 | Dublin37B | Ireland | Europe | 2002 | Lieberman_2022 | SS14 | Pop6 |
| SRR14277411 | PRJNA723099 | SAMN18804890 | China23 | China | Asia | 2018 | Lieberman_2022 | SS14 | Pop7 |
| SRR14277412 | PRJNA723099 | SAMN18804889 | China11 | China | Asia | 2018 | Lieberman_2022 | SS14 | Pop7 |
| SRR14277413 | PRJNA723099 | SAMN18804988 | MD536x | USA | N.America | 2001 | Lieberman_2022 | SS14 | Pop7 |
| SRR14277414 | PRJNA723099 | SAMN18804987 | MD530xe | USA | N.America | 2000 | Lieberman_2022 | SS14 | Pop7 |
| SRR14277415 | PRJNA723099 | SAMN18804986 | MD521B | USA | N.America | 2001 | Lieberman_2022 | SS14 | Pop7 |
| SRR14277416 | PRJNA723099 | SAMN18804985 | MD51x | USA | N.America | 2000 | Lieberman_2022 | SS14 | Pop7 |
| SRR14277417 | PRJNA723099 | SAMN18804984 | MD507x | USA | N.America | 1998 | Lieberman_2022 | SS14 | Pop7 |
| SRR14277418 | PRJNA723099 | SAMN18804983 | MD26x | USA | N.America | 1999 | Lieberman_2022 | SS14 | Pop7 |
| SRR14277419 | PRJNA723099 | SAMN18804982 | MD25x | USA | N.America | 1999 | Lieberman_2022 | SS14 | Pop7 |
| SRR14277420 | PRJNA723099 | SAMN18804981 | MD24xe | USA | N.America | 1999 | Lieberman_2022 | SS14 | Pop7 |
| SRR14277421 | PRJNA723099 | SAMN18804980 | MD22xe | USA | N.America | 1999 | Lieberman_2022 | SS14 | Pop8 |
| SRR14277422 | PRJNA723099 | SAMN18804979 | MD20B | USA | N.America | 1998 | Lieberman_2022 | SS14 | Pop7 |
| SRR14277423 | PRJNA723099 | SAMN18804898 | Dublin24B | Ireland | Europe | 2002 | Lieberman_2022 | SS14 | Pop6 |
| SRR14277425 | PRJNA723099 | SAMN18804977 | MD16x | USA | N.America | 1998 | Lieberman_2022 | SS14 | Pop6 |
| SRR14277426 | PRJNA723099 | SAMN18804976 | MD09B | USA | N.America | 1998 | Lieberman_2022 | SS14 | Pop7 |
| SRR14277428 | PRJNA723099 | SAMN18804974 | Japan368i | Japan | Asia | 2020 | Lieberman_2022 | Nichols | Pop3 |
| SRR14277429 | PRJNA723099 | SAMN18804973 | Japan365x | Japan | Asia | 2020 | Lieberman_2022 | SS14 | Pop7 |
| SRR14277430 | PRJNA723099 | SAMN18804972 | Japan364 | Japan | Asia | 2020 | Lieberman_2022 | SS14 | Pop7 |
| SRR14277431 | PRJNA723099 | SAMN18804971 | Japan362x | Japan | Asia | 2020 | Lieberman_2022 | SS14 | Pop7 |
| SRR14277432 | PRJNA723099 | SAMN18804970 | Japan361 | Japan | Asia | 2020 | Lieberman_2022 | SS14 | Pop7 |
| SRR14277433 | PRJNA723099 | SAMN18804969 | Japan360x | Japan | Asia | 2020 | Lieberman_2022 | SS14 | Pop7 |
| SRR14277434 | PRJNA723099 | SAMN18804897 | Dublin21Be | Ireland | Europe | 2002 | Lieberman_2022 | SS14 | Pop8 |
| SRR14277435 | PRJNA723099 | SAMN18804968 | Japan358 | Japan | Asia | 2020 | Lieberman_2022 | SS14 | Pop7 |
| SRR14277436 | PRJNA723099 | SAMN18804967 | Japan357 | Japan | Asia | 2019 | Lieberman_2022 | SS14 | Pop7 |
| SRR14277437 | PRJNA723099 | SAMN18804966 | Japan356 | Japan | Asia | 2019 | Lieberman_2022 | SS14 | Pop7 |
| SRR14277438 | PRJNA723099 | SAMN18804965 | Japan355 | Japan | Asia | 2019 | Lieberman_2022 | SS14 | Pop7 |
| SRR14277439 | PRJNA723099 | SAMN18804964 | Japan354x | Japan | Asia | 2019 | Lieberman_2022 | SS14 | Pop7 |
| SRR14277440 | PRJNA723099 | SAMN18804963 | Japan352xe | Japan | Asia | 2019 | Lieberman_2022 | SS14 | Pop7 |
| SRR14277441 | PRJNA723099 | SAMN18804962 | Japan351 | Japan | Asia | 2019 | Lieberman_2022 | SS14 | Pop7 |
| SRR14277442 | PRJNA723099 | SAMN18804961 | Japan349 | Japan | Asia | 2019 | Lieberman_2022 | SS14 | Pop7 |
| SRR14277443 | PRJNA723099 | SAMN18804960 | Japan348 | Japan | Asia | 2019 | Lieberman_2022 | SS14 | Pop7 |
| SRR14277445 | PRJNA723099 | SAMN18804896 | China48 | China | Asia | 2018 | Lieberman_2022 | SS14 | Pop7 |
| SRR14277446 | PRJNA723099 | SAMN18804958 | Japan345 | Japan | Asia | 2019 | Lieberman_2022 | SS14 | Pop7 |
| SRR14277447 | PRJNA723099 | SAMN18804957 | Japan344e | Japan | Asia | 2019 | Lieberman_2022 | SS14 | Pop7 |
| SRR14277448 | PRJNA723099 | SAMN18804956 | Japan343 | Japan | Asia | 2019 | Lieberman_2022 | SS14 | Pop7 |
| SRR14277449 | PRJNA723099 | SAMN18804955 | Japan342e | Japan | Asia | 2019 | Lieberman_2022 | SS14 | Pop7 |
| SRR14277450 | PRJNA723099 | SAMN18804954 | Japan339 | Japan | Asia | 2019 | Lieberman_2022 | SS14 | Pop7 |
| SRR14277451 | PRJNA723099 | SAMN18804953 | Japan338xei | Japan | Asia | 2019 | Lieberman_2022 | Nichols | Pop3 |
| SRR14277452 | PRJNA723099 | SAMN18804952 | Japan337 | Japan | Asia | 2019 | Lieberman_2022 | SS14 | Pop7 |

| | | | | | | | | | |
|---|---|---|---|---|---|---|---|---|---|
| SRR14277453 | PRJNA723099 | SAMN18804951 | Japan332 | Japan | Asia | 2019 | Lieberman_2022 | SS14 | Pop7 |
| SRR14277454 | PRJNA723099 | SAMN18804950 | Japan331 | Japan | Asia | 2019 | Lieberman_2022 | SS14 | Pop8 |
| SRR14277455 | PRJNA723099 | SAMN18804949 | Japan329 | Japan | Asia | 2019 | Lieberman_2022 | SS14 | Pop7 |
| SRR14277456 | PRJNA723099 | SAMN18804895 | China47 | China | Asia | 2018 | Lieberman_2022 | SS14 | Pop7 |
| SRR14277457 | PRJNA723099 | SAMN18804948 | Japan328x | Japan | Asia | 2019 | Lieberman_2022 | SS14 | Pop7 |
| SRR14277459 | PRJNA723099 | SAMN18804946 | Japan325xe | Japan | Asia | 2019 | Lieberman_2022 | SS14 | Pop7 |
| SRR14277460 | PRJNA723099 | SAMN18804945 | Japan324x | Japan | Asia | 2019 | Lieberman_2022 | SS14 | Pop8 |
| SRR15440024 | PRJNA754263 | SAMN20751642 | TPAUS_244 | Australia | Oceania | 2020 | Taouk_2022 | SS14 | Pop6 |
| SRR15440025 | PRJNA754263 | SAMN20751641 | TPAUS_243 | Australia | Oceania | 2015 | Taouk_2022 | SS14 | Pop7 |
| SRR15440026 | PRJNA754263 | SAMN20751640 | TPAUS_240 | Australia | Oceania | 2016 | Taouk_2022 | SS14 | Pop8 |
| SRR15440027 | PRJNA754263 | SAMN20751639 | TPAUS_237 | Australia | Oceania | 2016 | Taouk_2022 | SS14 | Pop6 |
| SRR15440028 | PRJNA754263 | SAMN20751638 | TPAUS_235 | Australia | Oceania | 2020 | Taouk_2022 | SS14 | Pop8 |
| SRR15440029 | PRJNA754263 | SAMN20751637 | TPAUS_234 | Australia | Oceania | 2020 | Taouk_2022 | SS14 | Pop6 |
| SRR15440030 | PRJNA754263 | SAMN20751493 | TPAUS_042 | Australia | Oceania | 2019 | Taouk_2022 | SS14 | Pop8 |
| SRR15440032 | PRJNA754263 | SAMN20751635 | TPAUS_231 | Australia | Oceania | 2019 | Taouk_2022 | SS14 | Pop8 |
| SRR15440033 | PRJNA754263 | SAMN20751634 | TPAUS_230 | Australia | Oceania | 2020 | Taouk_2022 | SS14 | Pop6 |
| SRR15440034 | PRJNA754263 | SAMN20751633 | TPAUS_229 | Australia | Oceania | 2020 | Taouk_2022 | SS14 | Pop6 |
| SRR15440035 | PRJNA754263 | SAMN20751632 | TPAUS_228 | Australia | Oceania | 2020 | Taouk_2022 | Nichols | Pop2 |
| SRR15440036 | PRJNA754263 | SAMN20751631 | TPAUS_226 | Australia | Oceania | 2020 | Taouk_2022 | SS14 | Pop6 |
| SRR15440037 | PRJNA754263 | SAMN20751630 | TPAUS_225 | Australia | Oceania | 2020 | Taouk_2022 | SS14 | Pop7 |
| SRR15440038 | PRJNA754263 | SAMN20751629 | TPAUS_224 | Australia | Oceania | 2020 | Taouk_2022 | SS14 | Pop6 |
| SRR15440039 | PRJNA754263 | SAMN20751628 | TPAUS_223 | Australia | Oceania | 2020 | Taouk_2022 | SS14 | Pop6 |
| SRR15440040 | PRJNA754263 | SAMN20751627 | TPAUS_222 | Australia | Oceania | 2020 | Taouk_2022 | SS14 | Pop8 |
| SRR15440041 | PRJNA754263 | SAMN20751492 | TPAUS_040 | Australia | Oceania | 2021 | Taouk_2022 | SS14 | Pop6 |
| SRR15440042 | PRJNA754263 | SAMN20751626 | TPAUS_221 | Australia | Oceania | 2021 | Taouk_2022 | SS14 | Pop6 |
| SRR15440043 | PRJNA754263 | SAMN20751625 | TPAUS_218 | Australia | Oceania | 2021 | Taouk_2022 | SS14 | Pop6 |
| SRR15440045 | PRJNA754263 | SAMN20751623 | TPAUS_211 | Australia | Oceania | 2021 | Taouk_2022 | SS14 | Pop8 |
| SRR15440046 | PRJNA754263 | SAMN20751622 | TPAUS_210 | Australia | Oceania | 2021 | Taouk_2022 | SS14 | Pop8 |
| SRR15440047 | PRJNA754263 | SAMN20751621 | TPAUS_208 | Australia | Oceania | 2021 | Taouk_2022 | SS14 | Pop6 |
| SRR15440048 | PRJNA754263 | SAMN20751620 | TPAUS_206 | Australia | Oceania | 2019 | Taouk_2022 | SS14 | Pop6 |
| SRR15440049 | PRJNA754263 | SAMN20751619 | TPAUS_205 | Australia | Oceania | 2019 | Taouk_2022 | Nichols | Pop3 |
| SRR15440050 | PRJNA754263 | SAMN20751618 | TPAUS_204 | Australia | Oceania | 2020 | Taouk_2022 | SS14 | Pop8 |
| SRR15440051 | PRJNA754263 | SAMN20751617 | TPAUS_203 | Australia | Oceania | 2020 | Taouk_2022 | SS14 | Pop6 |
| SRR15440052 | PRJNA754263 | SAMN20751491 | TPAUS_038 | Australia | Oceania | 2020 | Taouk_2022 | Nichols | Pop2 |
| SRR15440053 | PRJNA754263 | SAMN20751616 | TPAUS_201 | Australia | Oceania | 2020 | Taouk_2022 | SS14 | Pop8 |
| SRR15440054 | PRJNA754263 | SAMN20751615 | TPAUS_200 | Australia | Oceania | 2020 | Taouk_2022 | SS14 | Pop6 |
| SRR15440055 | PRJNA754263 | SAMN20751614 | TPAUS_199 | Australia | Oceania | 2020 | Taouk_2022 | SS14 | Pop6 |
| SRR15440056 | PRJNA754263 | SAMN20751613 | TPAUS_198 | Australia | Oceania | 2020 | Taouk_2022 | SS14 | Pop6 |
| SRR15440057 | PRJNA754263 | SAMN20751612 | TPAUS_197 | Australia | Oceania | 2020 | Taouk_2022 | SS14 | Pop6 |
| SRR15440058 | PRJNA754263 | SAMN20751611 | TPAUS_196 | Australia | Oceania | 2020 | Taouk_2022 | SS14 | Pop8 |
| SRR15440059 | PRJNA754263 | SAMN20751610 | TPAUS_195 | Australia | Oceania | 2020 | Taouk_2022 | SS14 | Pop6 |
| SRR15440060 | PRJNA754263 | SAMN20751609 | TPAUS_194 | Australia | Oceania | 2020 | Taouk_2022 | SS14 | Pop8 |
| SRR15440061 | PRJNA754263 | SAMN20751608 | TPAUS_190 | Australia | Oceania | 2020 | Taouk_2022 | SS14 | Pop6 |
| SRR15440062 | PRJNA754263 | SAMN20751607 | TPAUS_189 | Australia | Oceania | 2020 | Taouk_2022 | SS14 | Pop6 |

| | | | | | | | | | |
|---|---|---|---|---|---|---|---|---|---|
| SRR15440063 | PRJNA754263 | SAMN20751490 | TPAUS_037 | Australia | Oceania | 2020 | Taouk_2022 | Nichols | Pop2 |
| SRR15440064 | PRJNA754263 | SAMN20751606 | TPAUS_188 | Australia | Oceania | 2020 | Taouk_2022 | SS14 | Pop6 |
| SRR15440065 | PRJNA754263 | SAMN20751605 | TPAUS_187 | Australia | Oceania | 2020 | Taouk_2022 | SS14 | Pop8 |
| SRR15440066 | PRJNA754263 | SAMN20751604 | TPAUS_185 | Australia | Oceania | 2020 | Taouk_2022 | Nichols | Pop2 |
| SRR15440067 | PRJNA754263 | SAMN20751603 | TPAUS_184 | Australia | Oceania | 2020 | Taouk_2022 | Nichols | Pop2 |
| SRR15440068 | PRJNA754263 | SAMN20751602 | TPAUS_183 | Australia | Oceania | 2020 | Taouk_2022 | SS14 | Pop6 |
| SRR15440069 | PRJNA754263 | SAMN20751601 | TPAUS_182 | Australia | Oceania | 2020 | Taouk_2022 | SS14 | Pop6 |
| SRR15440070 | PRJNA754263 | SAMN20751600 | TPAUS_181 | Australia | Oceania | 2020 | Taouk_2022 | SS14 | Pop8 |
| SRR15440071 | PRJNA754263 | SAMN20751599 | TPAUS_180 | Australia | Oceania | 2020 | Taouk_2022 | SS14 | Pop6 |
| SRR15440072 | PRJNA754263 | SAMN20751598 | TPAUS_179 | Australia | Oceania | 2020 | Taouk_2022 | SS14 | Pop6 |
| SRR15440073 | PRJNA754263 | SAMN20751597 | TPAUS_178 | Australia | Oceania | 2020 | Taouk_2022 | Nichols | Pop2 |
| SRR15440074 | PRJNA754263 | SAMN20751489 | TPAUS_036 | Australia | Oceania | 2021 | Taouk_2022 | SS14 | Pop6 |
| SRR15440075 | PRJNA754263 | SAMN20751596 | TPAUS_177 | Australia | Oceania | 2021 | Taouk_2022 | SS14 | Pop6 |
| SRR15440076 | PRJNA754263 | SAMN20751595 | TPAUS_176 | Australia | Oceania | 2021 | Taouk_2022 | SS14 | Pop6 |
| SRR15440077 | PRJNA754263 | SAMN20751594 | TPAUS_175 | Australia | Oceania | 2021 | Taouk_2022 | SS14 | Pop8 |
| SRR15440078 | PRJNA754263 | SAMN20751593 | TPAUS_174 | Australia | Oceania | 2021 | Taouk_2022 | SS14 | Pop6 |
| SRR15440079 | PRJNA754263 | SAMN20751592 | TPAUS_173 | Australia | Oceania | 2021 | Taouk_2022 | SS14 | Pop6 |
| SRR15440080 | PRJNA754263 | SAMN20751591 | TPAUS_172 | Australia | Oceania | 2022 | Taouk_2022 | Nichols | Pop2 |
| SRR15440081 | PRJNA754263 | SAMN20751590 | TPAUS_171 | Australia | Oceania | 2021 | Taouk_2022 | Nichols | Pop2 |
| SRR15440082 | PRJNA754263 | SAMN20751589 | TPAUS_169 | Australia | Oceania | Unknown | Taouk_2022 | SS14 | Pop7 |
| SRR15440083 | PRJNA754263 | SAMN20751588 | TPAUS_168 | Australia | Oceania | 2021 | Taouk_2022 | SS14 | Pop6 |
| SRR15440084 | PRJNA754263 | SAMN20751587 | TPAUS_167 | Australia | Oceania | 2021 | Taouk_2022 | SS14 | Pop6 |
| SRR15440085 | PRJNA754263 | SAMN20751488 | TPAUS_035 | Australia | Oceania | 2021 | Taouk_2022 | Nichols | Pop2 |
| SRR15440086 | PRJNA754263 | SAMN20751586 | TPAUS_166 | Australia | Oceania | 2021 | Taouk_2022 | SS14 | Pop6 |
| SRR15440087 | PRJNA754263 | SAMN20751585 | TPAUS_165 | Australia | Oceania | 2021 | Taouk_2022 | SS14 | Pop6 |
| SRR15440088 | PRJNA754263 | SAMN20751584 | TPAUS_164 | Australia | Oceania | 2021 | Taouk_2022 | Nichols | Pop3 |
| SRR15440089 | PRJNA754263 | SAMN20751583 | TPAUS_163 | Australia | Oceania | 2021 | Taouk_2022 | SS14 | Pop6 |
| SRR15440090 | PRJNA754263 | SAMN20751582 | TPAUS_162 | Australia | Oceania | 2021 | Taouk_2022 | SS14 | Pop7 |
| SRR15440091 | PRJNA754263 | SAMN20751581 | TPAUS_161 | Australia | Oceania | 2021 | Taouk_2022 | SS14 | Pop6 |
| SRR15440092 | PRJNA754263 | SAMN20751580 | TPAUS_160 | Australia | Oceania | 2021 | Taouk_2022 | Nichols | Pop2 |
| SRR15440093 | PRJNA754263 | SAMN20751579 | TPAUS_159 | Australia | Oceania | 2021 | Taouk_2022 | SS14 | Pop6 |
| SRR15440094 | PRJNA754263 | SAMN20751578 | TPAUS_157 | Australia | Oceania | 2021 | Taouk_2022 | SS14 | Pop7 |
| SRR15440095 | PRJNA754263 | SAMN20751577 | TPAUS_156 | Australia | Oceania | 2022 | Taouk_2022 | SS14 | Pop6 |
| SRR15440096 | PRJNA754263 | SAMN20751487 | TPAUS_034 | Australia | Oceania | 2022 | Taouk_2022 | SS14 | Pop8 |
| SRR15440097 | PRJNA754263 | SAMN20751478 | TPAUS_007 | Australia | Oceania | 2022 | Taouk_2022 | Nichols | Pop2 |
| SRR15440098 | PRJNA754263 | SAMN20751477 | TPAUS_006 | Australia | Oceania | 2022 | Taouk_2022 | SS14 | Pop8 |
| SRR15440099 | PRJNA754263 | SAMN20751576 | TPAUS_155 | Australia | Oceania | 2022 | Taouk_2022 | SS14 | Pop6 |
| SRR15440100 | PRJNA754263 | SAMN20751575 | TPAUS_154 | Australia | Oceania | 2020 | Taouk_2022 | Nichols | Pop2 |
| SRR15440101 | PRJNA754263 | SAMN20751574 | TPAUS_153 | Australia | Oceania | 2020 | Taouk_2022 | SS14 | Pop6 |
| SRR15440102 | PRJNA754263 | SAMN20751573 | TPAUS_152 | Australia | Oceania | 2020 | Taouk_2022 | Nichols | Pop2 |
| SRR15440103 | PRJNA754263 | SAMN20751572 | TPAUS_151 | Australia | Oceania | 2020 | Taouk_2022 | SS14 | Pop8 |
| SRR15440104 | PRJNA754263 | SAMN20751571 | TPAUS_150 | Australia | Oceania | 2020 | Taouk_2022 | Nichols | Pop2 |
| SRR15440105 | PRJNA754263 | SAMN20751570 | TPAUS_149 | Australia | Oceania | 2020 | Taouk_2022 | SS14 | Pop8 |
| SRR15440106 | PRJNA754263 | SAMN20751569 | TPAUS_148 | Australia | Oceania | 2020 | Taouk_2022 | SS14 | Pop7 |

| | | | | | | | | | |
|---|---|---|---|---|---|---|---|---|---|
| SRR15440107 | PRJNA754263 | SAMN20751568 | TPAUS_147 | Australia | Oceania | 2020 | Taouk_2022 | SS14 | Pop6 |
| SRR15440108 | PRJNA754263 | SAMN20751567 | TPAUS_146 | Australia | Oceania | 2020 | Taouk_2022 | SS14 | Pop7 |
| SRR15440109 | PRJNA754263 | SAMN20751486 | TPAUS_029 | Australia | Oceania | 2020 | Taouk_2022 | Nichols | Pop2 |
| SRR15440110 | PRJNA754263 | SAMN20751566 | TPAUS_145 | Australia | Oceania | 2020 | Taouk_2022 | Nichols | Pop2 |
| SRR15440111 | PRJNA754263 | SAMN20751565 | TPAUS_144 | Australia | Oceania | 2020 | Taouk_2022 | Nichols | Pop2 |
| SRR15440112 | PRJNA754263 | SAMN20751564 | TPAUS_143 | Australia | Oceania | 2020 | Taouk_2022 | SS14 | Pop6 |
| SRR15440113 | PRJNA754263 | SAMN20751563 | TPAUS_142 | Australia | Oceania | 2020 | Taouk_2022 | Nichols | Pop3 |
| SRR15440114 | PRJNA754263 | SAMN20751562 | TPAUS_141 | Australia | Oceania | 2020 | Taouk_2022 | SS14 | Pop6 |
| SRR15440115 | PRJNA754263 | SAMN20751561 | TPAUS_140 | Australia | Oceania | 2020 | Taouk_2022 | Nichols | Pop2 |
| SRR15440116 | PRJNA754263 | SAMN20751560 | TPAUS_139 | Australia | Oceania | 2020 | Taouk_2022 | Nichols | Pop2 |
| SRR15440117 | PRJNA754263 | SAMN20751559 | TPAUS_138 | Australia | Oceania | 2020 | Taouk_2022 | SS14 | Pop6 |
| SRR15440118 | PRJNA754263 | SAMN20751558 | TPAUS_137 | Australia | Oceania | 2020 | Taouk_2022 | SS14 | Pop6 |
| SRR15440119 | PRJNA754263 | SAMN20751557 | TPAUS_136 | Australia | Oceania | 2020 | Taouk_2022 | SS14 | Pop6 |
| SRR15440120 | PRJNA754263 | SAMN20751485 | TPAUS_027 | Australia | Oceania | 2020 | Taouk_2022 | SS14 | Pop6 |
| SRR15440121 | PRJNA754263 | SAMN20751556 | TPAUS_135 | Australia | Oceania | 2020 | Taouk_2022 | Nichols | Pop2 |
| SRR15440122 | PRJNA754263 | SAMN20751555 | TPAUS_134 | Australia | Oceania | 2020 | Taouk_2022 | SS14 | Pop6 |
| SRR15440123 | PRJNA754263 | SAMN20751554 | TPAUS_133 | Australia | Oceania | 2020 | Taouk_2022 | SS14 | Pop6 |
| SRR15440124 | PRJNA754263 | SAMN20751553 | TPAUS_132 | Australia | Oceania | 2020 | Taouk_2022 | SS14 | Pop6 |
| SRR15440125 | PRJNA754263 | SAMN20751552 | TPAUS_131 | Australia | Oceania | 2020 | Taouk_2022 | SS14 | Pop6 |
| SRR15440126 | PRJNA754263 | SAMN20751551 | TPAUS_128 | Australia | Oceania | 2020 | Taouk_2022 | SS14 | Pop6 |
| SRR15440127 | PRJNA754263 | SAMN20751550 | TPAUS_127 | Australia | Oceania | 2020 | Taouk_2022 | Nichols | Pop2 |
| SRR15440128 | PRJNA754263 | SAMN20751549 | TPAUS_126 | Australia | Oceania | 2020 | Taouk_2022 | SS14 | Pop6 |
| SRR15440129 | PRJNA754263 | SAMN20751548 | TPAUS_125 | Australia | Oceania | 2020 | Taouk_2022 | Nichols | Pop2 |
| SRR15440130 | PRJNA754263 | SAMN20751547 | TPAUS_124 | Australia | Oceania | 2020 | Taouk_2022 | SS14 | Pop6 |
| SRR15440131 | PRJNA754263 | SAMN20751484 | TPAUS_025 | Australia | Oceania | 2020 | Taouk_2022 | SS14 | Pop6 |
| SRR15440132 | PRJNA754263 | SAMN20751546 | TPAUS_123 | Australia | Oceania | 2020 | Taouk_2022 | Nichols | Pop2 |
| SRR15440133 | PRJNA754263 | SAMN20751545 | TPAUS_122 | Australia | Oceania | 2020 | Taouk_2022 | SS14 | Pop6 |
| SRR15440134 | PRJNA754263 | SAMN20751544 | TPAUS_121 | Australia | Oceania | 2020 | Taouk_2022 | SS14 | Pop6 |
| SRR15440135 | PRJNA754263 | SAMN20751543 | TPAUS_120 | Australia | Oceania | 2020 | Taouk_2022 | SS14 | Pop7 |
| SRR15440136 | PRJNA754263 | SAMN20751542 | TPAUS_119 | Australia | Oceania | 2020 | Taouk_2022 | SS14 | Pop6 |
| SRR15440137 | PRJNA754263 | SAMN20751541 | TPAUS_118 | Australia | Oceania | 2020 | Taouk_2022 | SS14 | Pop7 |
| SRR15440138 | PRJNA754263 | SAMN20751540 | TPAUS_117 | Australia | Oceania | 2020 | Taouk_2022 | Nichols | Pop2 |
| SRR15440139 | PRJNA754263 | SAMN20751539 | TPAUS_116 | Australia | Oceania | 2020 | Taouk_2022 | SS14 | Pop6 |
| SRR15440140 | PRJNA754263 | SAMN20751538 | TPAUS_113 | Australia | Oceania | 2020 | Taouk_2022 | SS14 | Pop6 |
| SRR15440141 | PRJNA754263 | SAMN20751537 | TPAUS_111 | Australia | Oceania | 2020 | Taouk_2022 | SS14 | Pop6 |
| SRR15440142 | PRJNA754263 | SAMN20751483 | TPAUS_022 | Australia | Oceania | 2020 | Taouk_2022 | Nichols | Pop2 |
| SRR15440143 | PRJNA754263 | SAMN20751536 | TPAUS_109 | Australia | Oceania | 2020 | Taouk_2022 | Nichols | Pop2 |
| SRR15440144 | PRJNA754263 | SAMN20751535 | TPAUS_108 | Australia | Oceania | 2021 | Taouk_2022 | SS14 | Pop6 |
| SRR15440145 | PRJNA754263 | SAMN20751534 | TPAUS_107 | Australia | Oceania | 2021 | Taouk_2022 | SS14 | Pop6 |
| SRR15440146 | PRJNA754263 | SAMN20751533 | TPAUS_106 | Australia | Oceania | 2021 | Taouk_2022 | SS14 | Pop6 |
| SRR15440147 | PRJNA754263 | SAMN20751532 | TPAUS_102 | Australia | Oceania | 2021 | Taouk_2022 | SS14 | Pop6 |
| SRR15440148 | PRJNA754263 | SAMN20751531 | TPAUS_101 | Australia | Oceania | 2021 | Taouk_2022 | SS14 | Pop6 |
| SRR15440149 | PRJNA754263 | SAMN20751530 | TPAUS_100 | Australia | Oceania | 2021 | Taouk_2022 | Nichols | Pop2 |
| SRR15440150 | PRJNA754263 | SAMN20751529 | TPAUS_099 | Australia | Oceania | 2021 | Taouk_2022 | SS14 | Pop6 |

| | | | | | | | | | |
|---|---|---|---|---|---|---|---|---|---|
| SRR15440151 | PRJNA754263 | SAMN20751528 | TPAUS_098 | Australia | Oceania | 2021 | Taouk_2022 | SS14 | Pop6 |
| SRR15440152 | PRJNA754263 | SAMN20751527 | TPAUS_097 | Australia | Oceania | 2021 | Taouk_2022 | SS14 | Pop7 |
| SRR15440153 | PRJNA754263 | SAMN20751482 | TPAUS_018 | Australia | Oceania | 2021 | Taouk_2022 | SS14 | Pop7 |
| SRR15440154 | PRJNA754263 | SAMN20751526 | TPAUS_096 | Australia | Oceania | 2021 | Taouk_2022 | SS14 | Pop6 |
| SRR15440155 | PRJNA754263 | SAMN20751525 | TPAUS_094 | Australia | Oceania | 2021 | Taouk_2022 | SS14 | Pop6 |
| SRR15440156 | PRJNA754263 | SAMN20751524 | TPAUS_093 | Australia | Oceania | 2021 | Taouk_2022 | SS14 | Pop7 |
| SRR15440157 | PRJNA754263 | SAMN20751523 | TPAUS_092 | Australia | Oceania | 2021 | Taouk_2022 | Nichols | Pop2 |
| SRR15440158 | PRJNA754263 | SAMN20751932 | TPAUS_813 | Australia | Oceania | 2021 | Taouk_2022 | SS14 | Pop6 |
| SRR15440159 | PRJNA754263 | SAMN20751931 | TPAUS_811 | Australia | Oceania | 2021 | Taouk_2022 | SS14 | Pop8 |
| SRR15440160 | PRJNA754263 | SAMN20751930 | TPAUS_810 | Australia | Oceania | 2021 | Taouk_2022 | SS14 | Pop6 |
| SRR15440161 | PRJNA754263 | SAMN20751929 | TPAUS_809 | Australia | Oceania | 2021 | Taouk_2022 | SS14 | Pop8 |
| SRR15440163 | PRJNA754263 | SAMN20751927 | TPAUS_806 | Australia | Oceania | 2021 | Taouk_2022 | SS14 | Pop6 |
| SRR15440164 | PRJNA754263 | SAMN20751522 | TPAUS_091 | Australia | Oceania | 2021 | Taouk_2022 | SS14 | Pop8 |
| SRR15440165 | PRJNA754263 | SAMN20751926 | TPAUS_805 | Australia | Oceania | 2021 | Taouk_2022 | SS14 | Pop6 |
| SRR15440166 | PRJNA754263 | SAMN20751925 | TPAUS_804 | Australia | Oceania | 2021 | Taouk_2022 | SS14 | Pop6 |
| SRR15440167 | PRJNA754263 | SAMN20751924 | TPAUS_801 | Australia | Oceania | 2021 | Taouk_2022 | SS14 | Pop8 |
| SRR15440168 | PRJNA754263 | SAMN20751923 | TPAUS_799 | Australia | Oceania | 2021 | Taouk_2022 | SS14 | Pop6 |
| SRR15440169 | PRJNA754263 | SAMN20751922 | TPAUS_798 | Australia | Oceania | 2021 | Taouk_2022 | SS14 | Pop8 |
| SRR15440170 | PRJNA754263 | SAMN20751921 | TPAUS_797 | Australia | Oceania | 2021 | Taouk_2022 | SS14 | Pop6 |
| SRR15440171 | PRJNA754263 | SAMN20751920 | TPAUS_796 | Australia | Oceania | 2021 | Taouk_2022 | SS14 | Pop8 |
| SRR15440172 | PRJNA754263 | SAMN20751919 | TPAUS_795 | Australia | Oceania | 2021 | Taouk_2022 | SS14 | Pop8 |
| SRR15440173 | PRJNA754263 | SAMN20751918 | TPAUS_794 | Australia | Oceania | 2021 | Taouk_2022 | SS14 | Pop8 |
| SRR15440174 | PRJNA754263 | SAMN20751917 | TPAUS_793 | Australia | Oceania | 2021 | Taouk_2022 | SS14 | Pop6 |
| SRR15440175 | PRJNA754263 | SAMN20751521 | TPAUS_090 | Australia | Oceania | 2021 | Taouk_2022 | SS14 | Pop6 |
| SRR15440176 | PRJNA754263 | SAMN20751916 | TPAUS_791 | Australia | Oceania | 2020 | Taouk_2022 | SS14 | Pop6 |
| SRR15440177 | PRJNA754263 | SAMN20751915 | TPAUS_786 | Australia | Oceania | 2020 | Taouk_2022 | SS14 | Pop6 |
| SRR15440178 | PRJNA754263 | SAMN20751914 | TPAUS_785 | Australia | Oceania | 2020 | Taouk_2022 | SS14 | Pop8 |
| SRR15440179 | PRJNA754263 | SAMN20751913 | TPAUS_784 | Australia | Oceania | 2020 | Taouk_2022 | SS14 | Pop8 |
| SRR15440180 | PRJNA754263 | SAMN20751912 | TPAUS_783 | Australia | Oceania | 2020 | Taouk_2022 | SS14 | Pop8 |
| SRR15440181 | PRJNA754263 | SAMN20751911 | TPAUS_781 | Australia | Oceania | 2020 | Taouk_2022 | SS14 | Pop8 |
| SRR15440182 | PRJNA754263 | SAMN20751910 | TPAUS_780 | Australia | Oceania | 2020 | Taouk_2022 | SS14 | Pop6 |
| SRR15440183 | PRJNA754263 | SAMN20751909 | TPAUS_779 | Australia | Oceania | 2020 | Taouk_2022 | SS14 | Pop6 |
| SRR15440184 | PRJNA754263 | SAMN20751908 | TPAUS_775 | Australia | Oceania | 2020 | Taouk_2022 | SS14 | Pop7 |
| SRR15440185 | PRJNA754263 | SAMN20751907 | TPAUS_772 | Australia | Oceania | 40779 | Taouk_2022 | SS14 | Pop6 |
| SRR15440186 | PRJNA754263 | SAMN20751520 | TPAUS_089 | Australia | Oceania | 42391 | Taouk_2022 | Nichols | Pop3 |
| SRR15440187 | PRJNA754263 | SAMN20751906 | TPAUS_768 | Australia | Oceania | 40676 | Taouk_2022 | SS14 | Pop6 |
| SRR15440188 | PRJNA754263 | SAMN20751905 | TPAUS_766 | Australia | Oceania | 40525 | Taouk_2022 | SS14 | Pop6 |
| SRR15440189 | PRJNA754263 | SAMN20751904 | TPAUS_765 | Australia | Oceania | 40369 | Taouk_2022 | SS14 | Pop6 |
| SRR15440190 | PRJNA754263 | SAMN20751903 | TPAUS_764 | Australia | Oceania | 40197 | Taouk_2022 | SS14 | Pop7 |
| SRR15440191 | PRJNA754263 | SAMN20751902 | TPAUS_762 | Australia | Oceania | 40532 | Taouk_2022 | SS14 | Pop8 |
| SRR15440192 | PRJNA754263 | SAMN20751901 | TPAUS_761 | Australia | Oceania | 40529 | Taouk_2022 | SS14 | Pop6 |
| SRR15440193 | PRJNA754263 | SAMN20751900 | TPAUS_760 | Australia | Oceania | 40542 | Taouk_2022 | SS14 | Pop6 |
| SRR15440194 | PRJNA754263 | SAMN20751899 | TPAUS_759 | Australia | Oceania | 40198 | Taouk_2022 | SS14 | Pop8 |
| SRR15440195 | PRJNA754263 | SAMN20751898 | TPAUS_758 | Australia | Oceania | 40505 | Taouk_2022 | SS14 | Pop7 |

| | | | | | | | | | |
|---|---|---|---|---|---|---|---|---|---|
| SRR15440196 | PRJNA754263 | SAMN20751897 | TPAUS_757 | Australia | Oceania | 40506 | Taouk_2022 | SS14 | Pop7 |
| SRR15440197 | PRJNA754263 | SAMN20751519 | TPAUS_088 | Australia | Oceania | 42679 | Taouk_2022 | SS14 | Pop8 |
| SRR15440199 | PRJNA754263 | SAMN20751895 | TPAUS_755 | Australia | Oceania | 40338 | Taouk_2022 | SS14 | Pop6 |
| SRR15440200 | PRJNA754263 | SAMN20751894 | TPAUS_754 | Australia | Oceania | 40379 | Taouk_2022 | SS14 | Pop6 |
| SRR15440201 | PRJNA754263 | SAMN20751893 | TPAUS_753 | Australia | Oceania | 40315 | Taouk_2022 | SS14 | Pop8 |
| SRR15440202 | PRJNA754263 | SAMN20751892 | TPAUS_750 | Australia | Oceania | 40233 | Taouk_2022 | Nichols | Pop3 |
| SRR15440203 | PRJNA754263 | SAMN20751891 | TPAUS_746 | Australia | Oceania | 39884 | Taouk_2022 | SS14 | Pop8 |
| SRR15440204 | PRJNA754263 | SAMN20751890 | TPAUS_742 | Australia | Oceania | 39948 | Taouk_2022 | SS14 | Pop6 |
| SRR15440205 | PRJNA754263 | SAMN20751889 | TPAUS_738 | Australia | Oceania | 39874 | Taouk_2022 | SS14 | Pop8 |
| SRR15440206 | PRJNA754263 | SAMN20751888 | TPAUS_736 | Australia | Oceania | 39449 | Taouk_2022 | SS14 | Pop7 |
| SRR15440207 | PRJNA754263 | SAMN20751887 | TPAUS_733 | Australia | Oceania | 39610 | Taouk_2022 | SS14 | Pop8 |
| SRR15440208 | PRJNA754263 | SAMN20751518 | TPAUS_087 | Australia | Oceania | 43026 | Taouk_2022 | SS14 | Pop8 |
| SRR15440209 | PRJNA754263 | SAMN20751886 | TPAUS_732 | Australia | Oceania | 39623 | Taouk_2022 | SS14 | Pop8 |
| SRR15440210 | PRJNA754263 | SAMN20751885 | TPAUS_730 | Australia | Oceania | 39633 | Taouk_2022 | SS14 | Pop8 |
| SRR15440211 | PRJNA754263 | SAMN20751884 | TPAUS_729 | Australia | Oceania | 39535 | Taouk_2022 | SS14 | Pop6 |
| SRR15440212 | PRJNA754263 | SAMN20751883 | TPAUS_726 | Australia | Oceania | 39436 | Taouk_2022 | SS14 | Pop8 |
| SRR15440213 | PRJNA754263 | SAMN20751882 | TPAUS_724 | Australia | Oceania | 39338 | Taouk_2022 | SS14 | Pop6 |
| SRR15440214 | PRJNA754263 | SAMN20751881 | TPAUS_722 | Australia | Oceania | 39393 | Taouk_2022 | SS14 | Pop6 |
| SRR15440215 | PRJNA754263 | SAMN20751880 | TPAUS_718 | Australia | Oceania | 39157 | Taouk_2022 | SS14 | Pop6 |
| SRR15440216 | PRJNA754263 | SAMN20751879 | TPAUS_716 | Australia | Oceania | 39417 | Taouk_2022 | SS14 | Pop7 |
| SRR15440217 | PRJNA754263 | SAMN20751878 | TPAUS_713 | Australia | Oceania | 38867 | Taouk_2022 | SS14 | Pop8 |
| SRR15440218 | PRJNA754263 | SAMN20751877 | TPAUS_711 | Australia | Oceania | 39037 | Taouk_2022 | SS14 | Pop6 |
| SRR15440219 | PRJNA754263 | SAMN20751517 | TPAUS_086 | Australia | Oceania | 42514 | Taouk_2022 | SS14 | Pop7 |
| SRR15440220 | PRJNA754263 | SAMN20751481 | TPAUS_013 | Australia | Oceania | 43487 | Taouk_2022 | SS14 | Pop8 |
| SRR15440221 | PRJNA754263 | SAMN20751876 | TPAUS_710 | Australia | Oceania | 38974 | Taouk_2022 | SS14 | Pop6 |
| SRR15440222 | PRJNA754263 | SAMN20751875 | TPAUS_703 | Australia | Oceania | 38644 | Taouk_2022 | Nichols | Pop2 |
| SRR15440223 | PRJNA754263 | SAMN20751874 | TPAUS_699 | Australia | Oceania | 43534 | Taouk_2022 | SS14 | Pop6 |
| SRR15440224 | PRJNA754263 | SAMN20751873 | TPAUS_698 | Australia | Oceania | 43738 | Taouk_2022 | SS14 | Pop7 |
| SRR15440225 | PRJNA754263 | SAMN20751872 | TPAUS_697 | Australia | Oceania | 43542 | Taouk_2022 | SS14 | Pop6 |
| SRR15440226 | PRJNA754263 | SAMN20751871 | TPAUS_696 | Australia | Oceania | 43497 | Taouk_2022 | SS14 | Pop6 |
| SRR15440227 | PRJNA754263 | SAMN20751870 | TPAUS_695 | Australia | Oceania | 43525 | Taouk_2022 | SS14 | Pop8 |
| SRR15440228 | PRJNA754263 | SAMN20751869 | TPAUS_694 | Australia | Oceania | 43434 | Taouk_2022 | Nichols | Pop2 |
| SRR15440229 | PRJNA754263 | SAMN20751868 | TPAUS_693 | Australia | Oceania | 43424 | Taouk_2022 | Nichols | Pop2 |
| SRR15440230 | PRJNA754263 | SAMN20751867 | TPAUS_692 | Australia | Oceania | 43262 | Taouk_2022 | SS14 | Pop6 |
| SRR15440231 | PRJNA754263 | SAMN20751516 | TPAUS_085 | Australia | Oceania | 42517 | Taouk_2022 | SS14 | Pop6 |
| SRR15440233 | PRJNA754263 | SAMN20751865 | TPAUS_690 | Australia | Oceania | 43111 | Taouk_2022 | SS14 | Pop6 |
| SRR15440234 | PRJNA754263 | SAMN20751864 | TPAUS_689 | Australia | Oceania | 43398 | Taouk_2022 | SS14 | Pop6 |
| SRR15440235 | PRJNA754263 | SAMN20751863 | TPAUS_688 | Australia | Oceania | 43350 | Taouk_2022 | SS14 | Pop8 |
| SRR15440236 | PRJNA754263 | SAMN20751862 | TPAUS_687 | Australia | Oceania | 43166 | Taouk_2022 | SS14 | Pop6 |
| SRR15440237 | PRJNA754263 | SAMN20751861 | TPAUS_686 | Australia | Oceania | 43243 | Taouk_2022 | SS14 | Pop6 |
| SRR15440238 | PRJNA754263 | SAMN20751860 | TPAUS_685 | Australia | Oceania | 43235 | Taouk_2022 | SS14 | Pop6 |
| SRR15440239 | PRJNA754263 | SAMN20751859 | TPAUS_684 | Australia | Oceania | 43199 | Taouk_2022 | SS14 | Pop6 |
| SRR15440240 | PRJNA754263 | SAMN20751858 | TPAUS_682 | Australia | Oceania | 43367 | Taouk_2022 | SS14 | Pop6 |
| SRR15440242 | PRJNA754263 | SAMN20751515 | TPAUS_084 | Australia | Oceania | 43168 | Taouk_2022 | SS14 | Pop6 |

| | | | | | | | | | |
|---|---|---|---|---|---|---|---|---|---|
| SRR15440243 | PRJNA754263 | SAMN20751856 | TPAUS_680 | Australia | Oceania | 43257 | Taouk_2022 | SS14 | Pop6 |
| SRR15440244 | PRJNA754263 | SAMN20751855 | TPAUS_679 | Australia | Oceania | 43151 | Taouk_2022 | SS14 | Pop6 |
| SRR15440245 | PRJNA754263 | SAMN20751854 | TPAUS_678 | Australia | Oceania | 43171 | Taouk_2022 | SS14 | Pop6 |
| SRR15440246 | PRJNA754263 | SAMN20751853 | TPAUS_677 | Australia | Oceania | 43029 | Taouk_2022 | SS14 | Pop6 |
| SRR15440247 | PRJNA754263 | SAMN20751852 | TPAUS_676 | Australia | Oceania | 42933 | Taouk_2022 | SS14 | Pop7 |
| SRR15440248 | PRJNA754263 | SAMN20751851 | TPAUS_675 | Australia | Oceania | 42578 | Taouk_2022 | Nichols | Pop1 |
| SRR15440249 | PRJNA754263 | SAMN20751850 | TPAUS_674 | Australia | Oceania | 43725 | Taouk_2022 | SS14 | Pop6 |
| SRR15440250 | PRJNA754263 | SAMN20751849 | TPAUS_672 | Australia | Oceania | 43871 | Taouk_2022 | Nichols | Pop2 |
| SRR15440251 | PRJNA754263 | SAMN20751848 | TPAUS_671 | Australia | Oceania | 43961 | Taouk_2022 | SS14 | Pop8 |
| SRR15440252 | PRJNA754263 | SAMN20751847 | TPAUS_670 | Australia | Oceania | 43961 | Taouk_2022 | Nichols | Pop2 |
| SRR15440253 | PRJNA754263 | SAMN20751514 | TPAUS_083 | Australia | Oceania | 42930 | Taouk_2022 | Nichols | Pop2 |
| SRR15440254 | PRJNA754263 | SAMN20751846 | TPAUS_668 | Australia | Oceania | 44099 | Taouk_2022 | Nichols | Pop2 |
| SRR15440255 | PRJNA754263 | SAMN20751845 | TPAUS_666 | Australia | Oceania | 44098 | Taouk_2022 | SS14 | Pop6 |
| SRR15440256 | PRJNA754263 | SAMN20751844 | TPAUS_664 | Australia | Oceania | 44097 | Taouk_2022 | Nichols | Pop2 |
| SRR15440257 | PRJNA754263 | SAMN20751843 | TPAUS_663 | Australia | Oceania | 44097 | Taouk_2022 | SS14 | Pop8 |
| SRR15440258 | PRJNA754263 | SAMN20751842 | TPAUS_660 | Australia | Oceania | 44182 | Taouk_2022 | SS14 | Pop6 |
| SRR15440259 | PRJNA754263 | SAMN20751841 | TPAUS_657 | Australia | Oceania | 43404 | Taouk_2022 | SS14 | Pop8 |
| SRR15440260 | PRJNA754263 | SAMN20751840 | TPAUS_644 | Australia | Oceania | 44155 | Taouk_2022 | SS14 | Pop8 |
| SRR15440261 | PRJNA754263 | SAMN20751839 | TPAUS_643 | Australia | Oceania | 44155 | Taouk_2022 | SS14 | Pop6 |
| SRR15440262 | PRJNA754263 | SAMN20751838 | TPAUS_640 | Australia | Oceania | 44085 | Taouk_2022 | SS14 | Pop6 |
| SRR15440263 | PRJNA754263 | SAMN20751837 | TPAUS_639 | Australia | Oceania | 43872 | Taouk_2022 | SS14 | Pop6 |
| SRR15440264 | PRJNA754263 | SAMN20751513 | TPAUS_082 | Australia | Oceania | 42818 | Taouk_2022 | Nichols | Pop2 |
| SRR15440265 | PRJNA754263 | SAMN20751836 | TPAUS_638 | Australia | Oceania | 43932 | Taouk_2022 | SS14 | Pop8 |
| SRR15440266 | PRJNA754263 | SAMN20751835 | TPAUS_637 | Australia | Oceania | 43932 | Taouk_2022 | SS14 | Pop6 |
| SRR15440267 | PRJNA754263 | SAMN20751834 | TPAUS_634 | Australia | Oceania | 44130 | Taouk_2022 | Nichols | Pop2 |
| SRR15440268 | PRJNA754263 | SAMN20751833 | TPAUS_631 | Australia | Oceania | 44114 | Taouk_2022 | SS14 | Pop8 |
| SRR15440269 | PRJNA754263 | SAMN20751832 | TPAUS_621 | Australia | Oceania | 44196 | Taouk_2022 | SS14 | Pop8 |
| SRR15440270 | PRJNA754263 | SAMN20751831 | TPAUS_620 | Australia | Oceania | 44195 | Taouk_2022 | Nichols | Pop2 |
| SRR15440271 | PRJNA754263 | SAMN20751830 | TPAUS_619 | Australia | Oceania | 44195 | Taouk_2022 | SS14 | Pop8 |
| SRR15440273 | PRJNA754263 | SAMN20751828 | TPAUS_616 | Australia | Oceania | 44147 | Taouk_2022 | Nichols | Pop2 |
| SRR15440274 | PRJNA754263 | SAMN20751827 | TPAUS_615 | Australia | Oceania | 44024 | Taouk_2022 | SS14 | Pop6 |
| SRR15440275 | PRJNA754263 | SAMN20751512 | TPAUS_081 | Australia | Oceania | 42811 | Taouk_2022 | SS14 | Pop6 |
| SRR15440276 | PRJNA754263 | SAMN20751826 | TPAUS_614 | Australia | Oceania | 43842 | Taouk_2022 | SS14 | Pop6 |
| SRR15440277 | PRJNA754263 | SAMN20751825 | TPAUS_613 | Australia | Oceania | 44162 | Taouk_2022 | SS14 | Pop6 |
| SRR15440278 | PRJNA754263 | SAMN20751824 | TPAUS_612 | Australia | Oceania | 44163 | Taouk_2022 | SS14 | Pop6 |
| SRR15440279 | PRJNA754263 | SAMN20751823 | TPAUS_610 | Australia | Oceania | 44151 | Taouk_2022 | SS14 | Pop8 |
| SRR15440280 | PRJNA754263 | SAMN20751822 | TPAUS_609 | Australia | Oceania | 44146 | Taouk_2022 | Nichols | Pop2 |
| SRR15440281 | PRJNA754263 | SAMN20751821 | TPAUS_607 | Australia | Oceania | 44085 | Taouk_2022 | Nichols | Pop2 |
| SRR15440282 | PRJNA754263 | SAMN20751820 | TPAUS_606 | Australia | Oceania | 43932 | Taouk_2022 | Nichols | Pop2 |
| SRR15440283 | PRJNA754263 | SAMN20751819 | TPAUS_605 | Australia | Oceania | 43932 | Taouk_2022 | Nichols | Pop2 |
| SRR15440284 | PRJNA754263 | SAMN20751818 | TPAUS_604 | Australia | Oceania | 44119 | Taouk_2022 | SS14 | Pop6 |
| SRR15440285 | PRJNA754263 | SAMN20751817 | TPAUS_603 | Australia | Oceania | 44022 | Taouk_2022 | SS14 | Pop6 |
| SRR15440287 | PRJNA754263 | SAMN20751816 | TPAUS_602 | Australia | Oceania | 43992 | Taouk_2022 | SS14 | Pop8 |
| SRR15440288 | PRJNA754263 | SAMN20751815 | TPAUS_601 | Australia | Oceania | 43871 | Taouk_2022 | Nichols | Pop2 |

| | | | | | | | | | |
|---|---|---|---|---|---|---|---|---|---|
| SRR15440289 | PRJNA754263 | SAMN20751814 | TPAUS_600 | Australia | Oceania | 44104 | Taouk_2022 | SS14 | Pop8 |
| SRR15440290 | PRJNA754263 | SAMN20751813 | TPAUS_599 | Australia | Oceania | 44103 | Taouk_2022 | SS14 | Pop6 |
| SRR15440291 | PRJNA754263 | SAMN20751812 | TPAUS_598 | Australia | Oceania | 44102 | Taouk_2022 | Nichols | Pop2 |
| SRR15440292 | PRJNA754263 | SAMN20751811 | TPAUS_578 | Australia | Oceania | 44056 | Taouk_2022 | Nichols | Pop2 |
| SRR15440293 | PRJNA754263 | SAMN20751810 | TPAUS_572 | Australia | Oceania | 44111 | Taouk_2022 | Nichols | Pop2 |
| SRR15440294 | PRJNA754263 | SAMN20751809 | TPAUS_569 | Australia | Oceania | 44057 | Taouk_2022 | Nichols | Pop2 |
| SRR15440295 | PRJNA754263 | SAMN20751808 | TPAUS_568 | Australia | Oceania | 43989 | Taouk_2022 | SS14 | Pop8 |
| SRR15440296 | PRJNA754263 | SAMN20751807 | TPAUS_564 | Australia | Oceania | 43836 | Taouk_2022 | Nichols | Pop2 |
| SRR15440297 | PRJNA754263 | SAMN20751510 | TPAUS_079 | Australia | Oceania | 42543 | Taouk_2022 | Nichols | Pop2 |
| SRR15440298 | PRJNA754263 | SAMN20751806 | TPAUS_563 | Australia | Oceania | 43897 | Taouk_2022 | Nichols | Pop2 |
| SRR15440299 | PRJNA754263 | SAMN20751805 | TPAUS_562 | Australia | Oceania | 44011 | Taouk_2022 | SS14 | Pop8 |
| SRR15440300 | PRJNA754263 | SAMN20751804 | TPAUS_559 | Australia | Oceania | 44090 | Taouk_2022 | SS14 | Pop6 |
| SRR15440301 | PRJNA754263 | SAMN20751803 | TPAUS_555 | Australia | Oceania | 44144 | Taouk_2022 | SS14 | Pop6 |
| SRR15440303 | PRJNA754263 | SAMN20751801 | TPAUS_552 | Australia | Oceania | 44064 | Taouk_2022 | SS14 | Pop8 |
| SRR15440304 | PRJNA754263 | SAMN20751800 | TPAUS_550 | Australia | Oceania | 44067 | Taouk_2022 | Nichols | Pop2 |
| SRR15440305 | PRJNA754263 | SAMN20751799 | TPAUS_549 | Australia | Oceania | 44042 | Taouk_2022 | Nichols | Pop2 |
| SRR15440306 | PRJNA754263 | SAMN20751798 | TPAUS_548 | Australia | Oceania | 44020 | Taouk_2022 | Nichols | Pop2 |
| SRR15440307 | PRJNA754263 | SAMN20751797 | TPAUS_547 | Australia | Oceania | 44173 | Taouk_2022 | Nichols | Pop2 |
| SRR15440308 | PRJNA754263 | SAMN20751509 | TPAUS_078 | Australia | Oceania | 42544 | Taouk_2022 | SS14 | Pop6 |
| SRR15440309 | PRJNA754263 | SAMN20751796 | TPAUS_546 | Australia | Oceania | 44060 | Taouk_2022 | Nichols | Pop2 |
| SRR15440310 | PRJNA754263 | SAMN20751795 | TPAUS_543 | Australia | Oceania | 43896 | Taouk_2022 | Nichols | Pop2 |
| SRR15440311 | PRJNA754263 | SAMN20751794 | TPAUS_541 | Australia | Oceania | 44041 | Taouk_2022 | SS14 | Pop6 |
| SRR15440312 | PRJNA754263 | SAMN20751793 | TPAUS_540 | Australia | Oceania | 44029 | Taouk_2022 | Nichols | Pop2 |
| SRR15440313 | PRJNA754263 | SAMN20751792 | TPAUS_539 | Australia | Oceania | 44025 | Taouk_2022 | Nichols | Pop3 |
| SRR15440314 | PRJNA754263 | SAMN20751791 | TPAUS_538 | Australia | Oceania | 43837 | Taouk_2022 | SS14 | Pop7 |
| SRR15440315 | PRJNA754263 | SAMN20751790 | TPAUS_537 | Australia | Oceania | 43837 | Taouk_2022 | Nichols | Pop3 |
| SRR15440316 | PRJNA754263 | SAMN20751789 | TPAUS_536 | Australia | Oceania | 44006 | Taouk_2022 | Nichols | Pop2 |
| SRR15440317 | PRJNA754263 | SAMN20751788 | TPAUS_535 | Australia | Oceania | 44001 | Taouk_2022 | SS14 | Pop6 |
| SRR15440318 | PRJNA754263 | SAMN20751787 | TPAUS_534 | Australia | Oceania | 44090 | Taouk_2022 | SS14 | Pop8 |
| SRR15440319 | PRJNA754263 | SAMN20751508 | TPAUS_077 | Australia | Oceania | 42403 | Taouk_2022 | Nichols | Pop2 |
| SRR15440320 | PRJNA754263 | SAMN20751786 | TPAUS_533 | Australia | Oceania | 44092 | Taouk_2022 | SS14 | Pop7 |
| SRR15440322 | PRJNA754263 | SAMN20751784 | TPAUS_531 | Australia | Oceania | 44083 | Taouk_2022 | SS14 | Pop6 |
| SRR15440323 | PRJNA754263 | SAMN20751783 | TPAUS_530 | Australia | Oceania | 44113 | Taouk_2022 | Nichols | Pop2 |
| SRR15440324 | PRJNA754263 | SAMN20751782 | TPAUS_529 | Australia | Oceania | 44083 | Taouk_2022 | SS14 | Pop6 |
| SRR15440325 | PRJNA754263 | SAMN20751781 | TPAUS_528 | Australia | Oceania | 44052 | Taouk_2022 | SS14 | Pop6 |
| SRR15440326 | PRJNA754263 | SAMN20751780 | TPAUS_527 | Australia | Oceania | 43899 | Taouk_2022 | Nichols | Pop2 |
| SRR15440327 | PRJNA754263 | SAMN20751779 | TPAUS_526 | Australia | Oceania | 44001 | Taouk_2022 | SS14 | Pop8 |
| SRR15440328 | PRJNA754263 | SAMN20751778 | TPAUS_525 | Australia | Oceania | 44061 | Taouk_2022 | SS14 | Pop6 |
| SRR15440329 | PRJNA754263 | SAMN20751777 | TPAUS_524 | Australia | Oceania | 44064 | Taouk_2022 | SS14 | Pop6 |
| SRR15440330 | PRJNA754263 | SAMN20751507 | TPAUS_076 | Australia | Oceania | 42514 | Taouk_2022 | SS14 | Pop8 |
| SRR15440331 | PRJNA754263 | SAMN20751480 | TPAUS_010 | Australia | Oceania | 43553 | Taouk_2022 | SS14 | Pop8 |
| SRR15440332 | PRJNA754263 | SAMN20751776 | TPAUS_523 | Australia | Oceania | 44063 | Taouk_2022 | Nichols | Pop2 |
| SRR15440333 | PRJNA754263 | SAMN20751775 | TPAUS_522 | Australia | Oceania | 44060 | Taouk_2022 | Nichols | Pop2 |
| SRR15440334 | PRJNA754263 | SAMN20751774 | TPAUS_521 | Australia | Oceania | 43836 | Taouk_2022 | Nichols | Pop3 |

| | | | | | | | | | |
|---|---|---|---|---|---|---|---|---|---|
| SRR15440336 | PRJNA754263 | SAMN20751772 | TPAUS_510 | Australia | Oceania | 43861 | Taouk_2022 | SS14 | Pop8 |
| SRR15440337 | PRJNA754263 | SAMN20751771 | TPAUS_508 | Australia | Oceania | 43844 | Taouk_2022 | Nichols | Pop2 |
| SRR15440338 | PRJNA754263 | SAMN20751770 | TPAUS_507 | Australia | Oceania | 43644 | Taouk_2022 | Nichols | Pop2 |
| SRR15440339 | PRJNA754263 | SAMN20751769 | TPAUS_506 | Australia | Oceania | 43640 | Taouk_2022 | SS14 | Pop6 |
| SRR15440340 | PRJNA754263 | SAMN20751768 | TPAUS_502 | Australia | Oceania | 44044 | Taouk_2022 | SS14 | Pop6 |
| SRR15440341 | PRJNA754263 | SAMN20751767 | TPAUS_501 | Australia | Oceania | 43830 | Taouk_2022 | Nichols | Pop3 |
| SRR15440343 | PRJNA754263 | SAMN20751766 | TPAUS_498 | Australia | Oceania | 43643 | Taouk_2022 | SS14 | Pop8 |
| SRR15440344 | PRJNA754263 | SAMN20751765 | TPAUS_497 | Australia | Oceania | 43909 | Taouk_2022 | Nichols | Pop2 |
| SRR15440345 | PRJNA754263 | SAMN20751764 | TPAUS_496 | Australia | Oceania | 43916 | Taouk_2022 | SS14 | Pop8 |
| SRR15440346 | PRJNA754263 | SAMN20751763 | TPAUS_495 | Australia | Oceania | 43889 | Taouk_2022 | SS14 | Pop8 |
| SRR15440347 | PRJNA754263 | SAMN20751762 | TPAUS_489 | Australia | Oceania | 43616 | Taouk_2022 | SS14 | Pop8 |
| SRR15440348 | PRJNA754263 | SAMN20751761 | TPAUS_485 | Australia | Oceania | 43907 | Taouk_2022 | SS14 | Pop7 |
| SRR15440349 | PRJNA754263 | SAMN20751760 | TPAUS_483 | Australia | Oceania | 43768 | Taouk_2022 | SS14 | Pop6 |
| SRR15440350 | PRJNA754263 | SAMN20751759 | TPAUS_480 | Australia | Oceania | 43847 | Taouk_2022 | Nichols | Pop2 |
| SRR15440351 | PRJNA754263 | SAMN20751758 | TPAUS_477 | Australia | Oceania | 43864 | Taouk_2022 | Nichols | Pop2 |
| SRR15440352 | PRJNA754263 | SAMN20751757 | TPAUS_469 | Australia | Oceania | 43892 | Taouk_2022 | Nichols | Pop3 |
| SRR15440353 | PRJNA754263 | SAMN20751505 | TPAUS_073 | Australia | Oceania | 43361 | Taouk_2022 | SS14 | Pop8 |
| SRR15440354 | PRJNA754263 | SAMN20751756 | TPAUS_465 | Australia | Oceania | 43562 | Taouk_2022 | Nichols | Pop2 |
| SRR15440355 | PRJNA754263 | SAMN20751755 | TPAUS_462 | Australia | Oceania | 43810 | Taouk_2022 | SS14 | Pop6 |
| SRR15440356 | PRJNA754263 | SAMN20751754 | TPAUS_453 | Australia | Oceania | 43753 | Taouk_2022 | Nichols | Pop2 |
| SRR15440357 | PRJNA754263 | SAMN20751753 | TPAUS_450 | Australia | Oceania | 43644 | Taouk_2022 | SS14 | Pop8 |
| SRR15440359 | PRJNA754263 | SAMN20751751 | TPAUS_447 | Australia | Oceania | 43640 | Taouk_2022 | Nichols | Pop2 |
| SRR15440360 | PRJNA754263 | SAMN20751750 | TPAUS_446 | Australia | Oceania | 43670 | Taouk_2022 | SS14 | Pop7 |
| SRR15440361 | PRJNA754263 | SAMN20751749 | TPAUS_441 | Australia | Oceania | 43910 | Taouk_2022 | SS14 | Pop8 |
| SRR15440363 | PRJNA754263 | SAMN20751747 | TPAUS_439 | Australia | Oceania | 43675 | Taouk_2022 | SS14 | Pop6 |
| SRR15440364 | PRJNA754263 | SAMN20751504 | TPAUS_062 | Australia | Oceania | 43395 | Taouk_2022 | SS14 | Pop6 |
| SRR15440365 | PRJNA754263 | SAMN20751746 | TPAUS_436 | Australia | Oceania | 43829 | Taouk_2022 | Nichols | Pop2 |
| SRR15440366 | PRJNA754263 | SAMN20751745 | TPAUS_435 | Australia | Oceania | 43815 | Taouk_2022 | SS14 | Pop6 |
| SRR15440367 | PRJNA754263 | SAMN20751744 | TPAUS_433 | Australia | Oceania | 43926 | Taouk_2022 | SS14 | Pop8 |
| SRR15440368 | PRJNA754263 | SAMN20751743 | TPAUS_432 | Australia | Oceania | 43835 | Taouk_2022 | Nichols | Pop2 |
| SRR15440369 | PRJNA754263 | SAMN20751742 | TPAUS_431 | Australia | Oceania | 43980 | Taouk_2022 | Nichols | Pop2 |
| SRR15440370 | PRJNA754263 | SAMN20751741 | TPAUS_428 | Australia | Oceania | 43920 | Taouk_2022 | SS14 | Pop6 |
| SRR15440371 | PRJNA754263 | SAMN20751740 | TPAUS_427 | Australia | Oceania | 43938 | Taouk_2022 | SS14 | Pop6 |
| SRR15440372 | PRJNA754263 | SAMN20751739 | TPAUS_425 | Australia | Oceania | 43943 | Taouk_2022 | SS14 | Pop8 |
| SRR15440373 | PRJNA754263 | SAMN20751738 | TPAUS_422 | Australia | Oceania | 43949 | Taouk_2022 | SS14 | Pop6 |
| SRR15440374 | PRJNA754263 | SAMN20751737 | TPAUS_421 | Australia | Oceania | 43910 | Taouk_2022 | SS14 | Pop6 |
| SRR15440375 | PRJNA754263 | SAMN20751503 | TPAUS_057 | Australia | Oceania | 43370 | Taouk_2022 | SS14 | Pop8 |
| SRR15440376 | PRJNA754263 | SAMN20751736 | TPAUS_419 | Australia | Oceania | 43909 | Taouk_2022 | SS14 | Pop6 |
| SRR15440377 | PRJNA754263 | SAMN20751735 | TPAUS_418 | Australia | Oceania | 43907 | Taouk_2022 | Nichols | Pop3 |
| SRR15440378 | PRJNA754263 | SAMN20751734 | TPAUS_415 | Australia | Oceania | 44138 | Taouk_2022 | SS14 | Pop8 |
| SRR15440379 | PRJNA754263 | SAMN20751733 | TPAUS_410 | Australia | Oceania | 43852 | Taouk_2022 | Nichols | Pop2 |
| SRR15440380 | PRJNA754263 | SAMN20751732 | TPAUS_405 | Australia | Oceania | 43850 | Taouk_2022 | SS14 | Pop8 |
| SRR15440381 | PRJNA754263 | SAMN20751731 | TPAUS_404 | Australia | Oceania | 43888 | Taouk_2022 | SS14 | Pop8 |
| SRR15440382 | PRJNA754263 | SAMN20751730 | TPAUS_403 | Australia | Oceania | 43847 | Taouk_2022 | SS14 | Pop6 |

| | | | | | | | | | |
|---|---|---|---|---|---|---|---|---|---|
| SRR15440383 | PRJNA754263 | SAMN20751729 | TPAUS_402 | Australia | Oceania | 43844 | Taouk_2022 | Nichols | Pop2 |
| SRR15440384 | PRJNA754263 | SAMN20751728 | TPAUS_398 | Australia | Oceania | 43983 | Taouk_2022 | SS14 | Pop6 |
| SRR15440385 | PRJNA754263 | SAMN20751727 | TPAUS_397 | Australia | Oceania | 43983 | Taouk_2022 | SS14 | Pop6 |
| SRR15440386 | PRJNA754263 | SAMN20751502 | TPAUS_056 | Australia | Oceania | 43403 | Taouk_2022 | SS14 | Pop6 |
| SRR15440387 | PRJNA754263 | SAMN20751726 | TPAUS_396 | Australia | Oceania | 43983 | Taouk_2022 | SS14 | Pop8 |
| SRR15440388 | PRJNA754263 | SAMN20751725 | TPAUS_395 | Australia | Oceania | 43830 | Taouk_2022 | SS14 | Pop8 |
| SRR15440389 | PRJNA754263 | SAMN20751724 | TPAUS_393 | Australia | Oceania | 43787 | Taouk_2022 | Nichols | Pop2 |
| SRR15440390 | PRJNA754263 | SAMN20751723 | TPAUS_390 | Australia | Oceania | 43476 | Taouk_2022 | SS14 | Pop6 |
| SRR15440391 | PRJNA754263 | SAMN20751722 | TPAUS_388 | Australia | Oceania | 43750 | Taouk_2022 | SS14 | Pop6 |
| SRR15440392 | PRJNA754263 | SAMN20751721 | TPAUS_386 | Australia | Oceania | 43826 | Taouk_2022 | SS14 | Pop6 |
| SRR15440393 | PRJNA754263 | SAMN20751720 | TPAUS_383 | Australia | Oceania | 43737 | Taouk_2022 | SS14 | Pop6 |
| SRR15440394 | PRJNA754263 | SAMN20751719 | TPAUS_382 | Australia | Oceania | 43534 | Taouk_2022 | Nichols | Pop3 |
| SRR15440395 | PRJNA754263 | SAMN20751718 | TPAUS_378 | Australia | Oceania | 43752 | Taouk_2022 | Nichols | Pop2 |
| SRR15440396 | PRJNA754263 | SAMN20751717 | TPAUS_377 | Australia | Oceania | 43768 | Taouk_2022 | Nichols | Pop2 |
| SRR15440397 | PRJNA754263 | SAMN20751501 | TPAUS_054 | Australia | Oceania | 43417 | Taouk_2022 | SS14 | Pop8 |
| SRR15440398 | PRJNA754263 | SAMN20751716 | TPAUS_374 | Australia | Oceania | 43700 | Taouk_2022 | SS14 | Pop6 |
| SRR15440399 | PRJNA754263 | SAMN20751715 | TPAUS_372 | Australia | Oceania | 43688 | Taouk_2022 | SS14 | Pop8 |
| SRR15440400 | PRJNA754263 | SAMN20751714 | TPAUS_371 | Australia | Oceania | 43752 | Taouk_2022 | Nichols | Pop3 |
| SRR15440401 | PRJNA754263 | SAMN20751713 | TPAUS_368 | Australia | Oceania | 43717 | Taouk_2022 | SS14 | Pop6 |
| SRR15440402 | PRJNA754263 | SAMN20751712 | TPAUS_364 | Australia | Oceania | 43601 | Taouk_2022 | SS14 | Pop7 |
| SRR15440403 | PRJNA754263 | SAMN20751711 | TPAUS_362 | Australia | Oceania | 43601 | Taouk_2022 | SS14 | Pop8 |
| SRR15440404 | PRJNA754263 | SAMN20751710 | TPAUS_356 | Australia | Oceania | 43800 | Taouk_2022 | SS14 | Pop8 |
| SRR15440405 | PRJNA754263 | SAMN20751709 | TPAUS_350 | Australia | Oceania | 43480 | Taouk_2022 | Nichols | Pop2 |
| SRR15440406 | PRJNA754263 | SAMN20751708 | TPAUS_345 | Australia | Oceania | 43307 | Taouk_2022 | SS14 | Pop6 |
| SRR15440407 | PRJNA754263 | SAMN20751707 | TPAUS_340 | Australia | Oceania | 43286 | Taouk_2022 | Nichols | Pop2 |
| SRR15440408 | PRJNA754263 | SAMN20751500 | TPAUS_053 | Australia | Oceania | 43111 | Taouk_2022 | Nichols | Pop2 |
| SRR15440409 | PRJNA754263 | SAMN20751706 | TPAUS_334 | Australia | Oceania | 43207 | Taouk_2022 | SS14 | Pop8 |
| SRR15440410 | PRJNA754263 | SAMN20751705 | TPAUS_331 | Australia | Oceania | 43185 | Taouk_2022 | Nichols | Pop2 |
| SRR15440411 | PRJNA754263 | SAMN20751704 | TPAUS_330 | Australia | Oceania | 43139 | Taouk_2022 | Nichols | Pop2 |
| SRR15440412 | PRJNA754263 | SAMN20751703 | TPAUS_326 | Australia | Oceania | 42918 | Taouk_2022 | Nichols | Pop2 |
| SRR15440413 | PRJNA754263 | SAMN20751702 | TPAUS_325 | Australia | Oceania | 43000 | Taouk_2022 | Nichols | Pop2 |
| SRR15440414 | PRJNA754263 | SAMN20751701 | TPAUS_322 | Australia | Oceania | 42826 | Taouk_2022 | SS14 | Pop6 |
| SRR15440415 | PRJNA754263 | SAMN20751700 | TPAUS_321 | Australia | Oceania | 42626 | Taouk_2022 | SS14 | Pop6 |
| SRR15440416 | PRJNA754263 | SAMN20751699 | TPAUS_319 | Australia | Oceania | 42464 | Taouk_2022 | SS14 | Pop6 |
| SRR15440417 | PRJNA754263 | SAMN20751698 | TPAUS_314 | Australia | Oceania | 42780 | Taouk_2022 | SS14 | Pop8 |
| SRR15440418 | PRJNA754263 | SAMN20751697 | TPAUS_313 | Australia | Oceania | 42780 | Taouk_2022 | SS14 | Pop8 |
| SRR15440419 | PRJNA754263 | SAMN20751499 | TPAUS_052 | Australia | Oceania | 43111 | Taouk_2022 | SS14 | Pop8 |
| SRR15440420 | PRJNA754263 | SAMN20751696 | TPAUS_312 | Australia | Oceania | 43027 | Taouk_2022 | Nichols | Pop2 |
| SRR15440421 | PRJNA754263 | SAMN20751695 | TPAUS_311 | Australia | Oceania | 43027 | Taouk_2022 | Nichols | Pop2 |
| SRR15440423 | PRJNA754263 | SAMN20751693 | TPAUS_308 | Australia | Oceania | 42765 | Taouk_2022 | SS14 | Pop8 |
| SRR15440424 | PRJNA754263 | SAMN20751692 | TPAUS_307 | Australia | Oceania | 42846 | Taouk_2022 | Nichols | Pop2 |
| SRR15440425 | PRJNA754263 | SAMN20751691 | TPAUS_306 | Australia | Oceania | 43096 | Taouk_2022 | SS14 | Pop6 |
| SRR15440426 | PRJNA754263 | SAMN20751690 | TPAUS_305 | Australia | Oceania | 42507 | Taouk_2022 | SS14 | Pop8 |
| SRR15440428 | PRJNA754263 | SAMN20751688 | TPAUS_302 | Australia | Oceania | 42555 | Taouk_2022 | SS14 | Pop6 |

| | | | | | | | | | |
|---|---|---|---|---|---|---|---|---|---|
| SRR15440429 | PRJNA754263 | SAMN20751687 | TPAUS_300 | Australia | Oceania | 42718 | Taouk_2022 | Nichols | Pop3 |
| SRR15440430 | PRJNA754263 | SAMN20751498 | TPAUS_051 | Australia | Oceania | 43402 | Taouk_2022 | SS14 | Pop8 |
| SRR15440432 | PRJNA754263 | SAMN20751685 | TPAUS_298 | Australia | Oceania | 42417 | Taouk_2022 | SS14 | Pop8 |
| SRR15440433 | PRJNA754263 | SAMN20751684 | TPAUS_295 | Australia | Oceania | 42856 | Taouk_2022 | SS14 | Pop8 |
| SRR15440434 | PRJNA754263 | SAMN20751683 | TPAUS_294 | Australia | Oceania | 42525 | Taouk_2022 | SS14 | Pop6 |
| SRR15440435 | PRJNA754263 | SAMN20751682 | TPAUS_293 | Australia | Oceania | 42431 | Taouk_2022 | SS14 | Pop6 |
| SRR15440436 | PRJNA754263 | SAMN20751681 | TPAUS_292 | Australia | Oceania | 42699 | Taouk_2022 | SS14 | Pop7 |
| SRR15440437 | PRJNA754263 | SAMN20751680 | TPAUS_291 | Australia | Oceania | 42525 | Taouk_2022 | SS14 | Pop8 |
| SRR15440438 | PRJNA754263 | SAMN20751679 | TPAUS_290 | Australia | Oceania | 42459 | Taouk_2022 | Nichols | Pop2 |
| SRR15440439 | PRJNA754263 | SAMN20751678 | TPAUS_289 | Australia | Oceania | 42616 | Taouk_2022 | SS14 | Pop6 |
| SRR15440441 | PRJNA754263 | SAMN20751497 | TPAUS_048 | Australia | Oceania | 43403 | Taouk_2022 | SS14 | Pop6 |
| SRR15440443 | PRJNA754263 | SAMN20751676 | TPAUS_283 | Australia | Oceania | 42179 | Taouk_2022 | SS14 | Pop8 |
| SRR15440444 | PRJNA754263 | SAMN20751675 | TPAUS_282 | Australia | Oceania | 42306 | Taouk_2022 | SS14 | Pop6 |
| SRR15440445 | PRJNA754263 | SAMN20751674 | TPAUS_279 | Australia | Oceania | 42106 | Taouk_2022 | SS14 | Pop6 |
| SRR15440446 | PRJNA754263 | SAMN20751673 | TPAUS_278 | Australia | Oceania | 43570 | Taouk_2022 | SS14 | Pop6 |
| SRR15440447 | PRJNA754263 | SAMN20751672 | TPAUS_277 | Australia | Oceania | 42823 | Taouk_2022 | SS14 | Pop8 |
| SRR15440448 | PRJNA754263 | SAMN20751671 | TPAUS_276 | Australia | Oceania | 42360 | Taouk_2022 | SS14 | Pop6 |
| SRR15440449 | PRJNA754263 | SAMN20751670 | TPAUS_275 | Australia | Oceania | 42849 | Taouk_2022 | SS14 | Pop6 |
| SRR15440450 | PRJNA754263 | SAMN20751669 | TPAUS_274 | Australia | Oceania | 42759 | Taouk_2022 | SS14 | Pop6 |
| SRR15440451 | PRJNA754263 | SAMN20751668 | TPAUS_272 | Australia | Oceania | 43146 | Taouk_2022 | Nichols | Pop2 |
| SRR15440452 | PRJNA754263 | SAMN20751667 | TPAUS_271 | Australia | Oceania | 43129 | Taouk_2022 | Nichols | Pop2 |
| SRR15440454 | PRJNA754263 | SAMN20751666 | TPAUS_270 | Australia | Oceania | 43344 | Taouk_2022 | SS14 | Pop6 |
| SRR15440455 | PRJNA754263 | SAMN20751665 | TPAUS_269 | Australia | Oceania | 43123 | Taouk_2022 | SS14 | Pop6 |
| SRR15440456 | PRJNA754263 | SAMN20751664 | TPAUS_268 | Australia | Oceania | 43145 | Taouk_2022 | SS14 | Pop6 |
| SRR15440457 | PRJNA754263 | SAMN20751663 | TPAUS_267 | Australia | Oceania | 43223 | Taouk_2022 | Nichols | Pop3 |
| SRR15440458 | PRJNA754263 | SAMN20751662 | TPAUS_266 | Australia | Oceania | 43337 | Taouk_2022 | SS14 | Pop6 |
| SRR15440459 | PRJNA754263 | SAMN20751661 | TPAUS_265 | Australia | Oceania | 43340 | Taouk_2022 | SS14 | Pop8 |
| SRR15440460 | PRJNA754263 | SAMN20751660 | TPAUS_264 | Australia | Oceania | 43340 | Taouk_2022 | Nichols | Pop2 |
| SRR15440461 | PRJNA754263 | SAMN20751659 | TPAUS_263 | Australia | Oceania | 43339 | Taouk_2022 | SS14 | Pop6 |
| SRR15440462 | PRJNA754263 | SAMN20751658 | TPAUS_262 | Australia | Oceania | 43337 | Taouk_2022 | SS14 | Pop6 |
| SRR15440463 | PRJNA754263 | SAMN20751657 | TPAUS_261 | Australia | Oceania | 43259 | Taouk_2022 | Nichols | Pop2 |
| SRR15440464 | PRJNA754263 | SAMN20751495 | TPAUS_045 | Australia | Oceania | 43446 | Taouk_2022 | SS14 | Pop6 |
| SRR15440465 | PRJNA754263 | SAMN20751656 | TPAUS_260 | Australia | Oceania | 43297 | Taouk_2022 | SS14 | Pop7 |
| SRR15440466 | PRJNA754263 | SAMN20751655 | TPAUS_259 | Australia | Oceania | 43305 | Taouk_2022 | SS14 | Pop6 |
| SRR15440467 | PRJNA754263 | SAMN20751654 | TPAUS_258 | Australia | Oceania | 43301 | Taouk_2022 | SS14 | Pop6 |
| SRR15440468 | PRJNA754263 | SAMN20751653 | TPAUS_257 | Australia | Oceania | 43306 | Taouk_2022 | SS14 | Pop8 |
| SRR15440469 | PRJNA754263 | SAMN20751652 | TPAUS_256 | Australia | Oceania | 43306 | Taouk_2022 | SS14 | Pop6 |
| SRR15440470 | PRJNA754263 | SAMN20751651 | TPAUS_255 | Australia | Oceania | 43304 | Taouk_2022 | SS14 | Pop8 |
| SRR15440471 | PRJNA754263 | SAMN20751650 | TPAUS_254 | Australia | Oceania | 43295 | Taouk_2022 | SS14 | Pop8 |
| SRR15440472 | PRJNA754263 | SAMN20751649 | TPAUS_253 | Australia | Oceania | 43298 | Taouk_2022 | SS14 | Pop6 |
| SRR15440473 | PRJNA754263 | SAMN20751648 | TPAUS_251 | Australia | Oceania | 43350 | Taouk_2022 | SS14 | Pop6 |
| SRR15440474 | PRJNA754263 | SAMN20751647 | TPAUS_250 | Australia | Oceania | 43166 | Taouk_2022 | SS14 | Pop6 |
| SRR15440475 | PRJNA754263 | SAMN20751494 | TPAUS_043 | Australia | Oceania | 43385 | Taouk_2022 | SS14 | Pop8 |
| SRR15440476 | PRJNA754263 | SAMN20751646 | TPAUS_248 | Australia | Oceania | 43257 | Taouk_2022 | Nichols | Pop3 |

| | | | | | | | | | |
|---|---|---|---|---|---|---|---|---|---|
| SRR15440478 | PRJNA754263 | SAMN20751644 | TPAUS_246 | Australia | Oceania | 43241 | Taouk_2022 | SS14 | Pop7 |
| SRR15440479 | PRJNA754263 | SAMN20751643 | TPAUS_245 | Australia | Oceania | 43237 | Taouk_2022 | SS14 | Pop7 |
| SRR2996724 | PRJNA305961 | SAMN04334598 | SHC-0 | China | Asia | 2015 | Sun_2016 | SS14 | Pop7 |
| SRR2996725 | PRJNA305961 | SAMN04334599 | SHD-R | China | Asia | 2015 | Sun_2016 | SS14 | Pop7 |
| SRR2996726 | PRJNA305961 | SAMN04334600 | SHE-V | China | Asia | 2015 | Sun_2016 | SS14 | Pop7 |
| SRR2996727 | PRJNA305961 | SAMN04334601 | SHG-I2 | China | Asia | 2015 | Sun_2016 | SS14 | Pop7 |
| SRR2996728 | PRJNA305961 | SAMN04334602 | B3 | China | Asia | 2015 | Sun_2016 | SS14 | Pop7 |
| SRR2996729 | PRJNA305961 | SAMN04334603 | C3 | China | Asia | 2015 | Sun_2016 | SS14 | Pop7 |
| SRR2996730 | PRJNA305961 | SAMN04334604 | K3 | China | Asia | 2015 | Sun_2016 | SS14 | Pop7 |
| SRR2996732 | PRJNA305961 | SAMN04334611 | Q3 | China | Asia | 2015 | Sun_2016 | SS14 | Pop7 |
| SRR3268682 | PRJNA313497 | SAMN04524046 | AR2 | Argentina | S.America | 2013 | Arora_2016 | SS14 | Pop8 |
| SRR3268685 | PRJNA313497 | SAMN04524042 | CZ27 | Czech | Europe | 2012 | Arora_2016 | SS14 | Pop8 |
| SRR3268696 | PRJNA313497 | SAMN04524084 | GRA2 | USA | N.America | 1980-1999 | Arora_2016 | SS14 | Pop8 |
| SRR3268701 | PRJNA313497 | SAMN04524027 | NE12 | Netherlands | Europe | 2013 | Arora_2016 | SS14 | Pop8 |
| SRR3268702 | PRJNA313497 | SAMN04524028 | NE13 | Netherlands | Europe | 2013 | Arora_2016 | SS14 | Pop8 |
| SRR3268703 | PRJNA313497 | SAMN04524029 | NE14 | Netherlands | Europe | 2013 | Arora_2016 | SS14 | Pop6 |
| SRR3268705 | PRJNA313497 | SAMN04524030 | NE15 | Netherlands | Europe | 2013 | Arora_2016 | SS14 | Pop6 |
| SRR3268707 | PRJNA313497 | SAMN04524032 | NE17 | Netherlands | Europe | 2013 | Arora_2016 | SS14 | Pop6 |
| SRR3268709 | PRJNA313497 | SAMN04524034 | NE19 | Netherlands | Europe | 2013 | Arora_2016 | SS14 | Pop7 |
| SRR3268710 | PRJNA313497 | SAMN04524035 | NE20 | Netherlands | Europe | 2013 | Arora_2016 | Nichols | Pop2 |
| SRR3268712 | PRJNA313497 | SAMN04524050 | NIC1 | USA | N.America | 1912 | Arora_2016 | Nichols | Pop1 |
| SRR3268713 | PRJNA313497 | SAMN04524060 | NIC2 | USA | N.America | 1912 | Arora_2016 | Nichols | Pop1 |
| SRR3268715 | PRJNA313497 | SAMN04524053 | BAL3 | USA | N.America | 1973 | Arora_2016 | Nichols | Pop1 |
| SRR3268718 | PRJNA313497 | SAMN04524026 | SW11 | Switzerland | Europe | 2013 | Arora_2016 | SS14 | Pop6 |
| SRR3268722 | PRJNA313497 | SAMN04524064 | AU15 | Austria | Europe | 2013 | Arora_2016 | SS14 | Pop6 |
| SRR3268723 | PRJNA313497 | SAMN04524065 | AU16 | Austria | Europe | 2013 | Arora_2016 | SS14 | Pop6 |
| SRR3268726 | PRJNA313497 | SAMN04524055 | BAL73 | USA | N.America | 1973 | Arora_2016 | Nichols | Pop1 |
| SRR3268730 | PRJNA313497 | SAMN04524019 | SW4 | Switzerland | Europe | 2012 | Arora_2016 | SS14 | Pop8 |
| SRR3268732 | PRJNA313497 | SAMN04524021 | SW6 | Switzerland | Europe | 2012 | Arora_2016 | SS14 | Pop7 |
| SRR3268736 | PRJNA313497 | SAMN04524023 | SW8 | Switzerland | Europe | 2012 | Arora_2016 | SS14 | Pop6 |
| SRR3268740 | PRJNA313497 | SAMN04524056 | SEA86 | USA | N.America | 1986 | Arora_2016 | Nichols | Pop2 |
| SRR3268758 | PRJNA313497 | SAMN04524058 | UW1 | USA | N.America | 2004 | Arora_2016 | SS14 | Pop6 |
| SRR3571774 | PRJNA322283 | SAMN05150260 | PT_SIF0697 | Portugal | Europe | 2009 | Pinto_2016 | SS14 | Pop7 |
| SRR3571775 | PRJNA322283 | SAMN05150353 | PT_SIF0877_3 | Portugal | Europe | 2010 | Pinto_2016 | SS14 | Pop6 |
| SRR3571776 | PRJNA322283 | SAMN05150354 | PT_SIF0908 | Portugal | Europe | 2010 | Pinto_2016 | SS14 | Pop8 |
| SRR3571778 | PRJNA322283 | SAMN05150355 | PT_SIF0954 | Portugal | Europe | 2010 | Pinto_2016 | SS14 | Pop6 |
| SRR3571783 | PRJNA322283 | SAMN05150356 | PT_SIF1002 | Portugal | Europe | 2011 | Pinto_2016 | SS14 | Pop7 |
| SRR3571785 | PRJNA322283 | SAMN05150357 | PT_SIF1020 | Portugal | Europe | 2011 | Pinto_2016 | SS14 | Pop7 |
| SRR3571786 | PRJNA322283 | SAMN05150358 | PT_SIF1063 | Portugal | Europe | 2012 | Pinto_2016 | SS14 | Pop7 |
| SRR3571794 | PRJNA322283 | SAMN05150778 | PT_SIF1127 | Portugal | Europe | 2013 | Pinto_2016 | SS14 | Pop8 |
| SRR3584837 | PRJNA322283 | SAMN05150382 | PT_SIF1135 | Portugal | Europe | 2013 | Pinto_2016 | SS14 | Pop7 |
| SRR3584838 | PRJNA322283 | SAMN05150594 | PT_SIF1140 | Portugal | Europe | 2013 | Pinto_2016 | SS14 | Pop8 |
| SRR3584839 | PRJNA322283 | SAMN05150624 | PT_SIF1142 | Portugal | Europe | 2013 | Pinto_2016 | SS14 | Pop8 |
| SRR3584840 | PRJNA322283 | SAMN05150625 | PT_SIF1156 | Portugal | Europe | 2013 | Pinto_2016 | SS14 | Pop7 |

| | | | | | | | | | |
|---|---|---|---|---|---|---|---|---|---|
| SRR3584841 | PRJNA322283 | SAMN05150626 | PT_SIF1167 | Portugal | Europe | 2013 | Pinto_2016 | SS14 | Pop7 |
| SRR3584842 | PRJNA322283 | SAMN05150627 | PT_SIF1183 | Portugal | Europe | 2013 | Pinto_2016 | SS14 | Pop6 |
| SRR3584843 | PRJNA322283 | SAMN05150628 | PT_SIF1196 | Portugal | Europe | 2013 | Pinto_2016 | SS14 | Pop7 |
| SRR3584844 | PRJNA322283 | SAMN05150632 | PT_SIF1200 | Portugal | Europe | 2013 | Pinto_2016 | SS14 | Pop6 |
| SRR3584845 | PRJNA322283 | SAMN05150648 | PT_SIF1242 | Portugal | Europe | 2014 | Pinto_2016 | SS14 | Pop7 |
| SRR3584871 | PRJNA322283 | SAMN05150673 | PT_SIF1252 | Portugal | Europe | 2014 | Pinto_2016 | SS14 | Pop6 |
| SRR3584875 | PRJNA322283 | SAMN05150674 | PT_SIF1261 | Portugal | Europe | 2014 | Pinto_2016 | SS14 | Pop6 |
| SRR3584879 | PRJNA322283 | SAMN05150675 | PT_SIF1278 | Portugal | Europe | 2014 | Pinto_2016 | SS14 | Pop6 |
| SRR3584882 | PRJNA322283 | SAMN05150676 | PT_SIF1280 | Portugal | Europe | 2014 | Pinto_2016 | SS14 | Pop6 |
| SRR3584886 | PRJNA322283 | SAMN05150777 | PT_SIF1348 | Portugal | Europe | 2014 | Pinto_2016 | SS14 | Pop6 |
| SRR3584962 | PRJNA322283 | SAMN05150273 | PT_SIF0751 | Portugal | Europe | 2009 | Pinto_2016 | SS14 | Pop8 |
| SRR3584965 | PRJNA322283 | SAMN05173064 | PT_SIF0857 | Portugal | Europe | 2010 | Pinto_2016 | SS14 | Pop7 |
| SRR8494413 | PRJNA508872 | SAMN10829684 | CW45 | Czech | Europe | 2013 | Grillova_2019 | SS14 | Pop8 |
| SRR8494414 | PRJNA508872 | SAMN10829685 | CW89 | Czech | Europe | 2017 | Grillova_2019 | SS14 | Pop8 |
| SRR8494415 | PRJNA508872 | SAMN10829686 | CW33 | Czech | Europe | 2012 | Grillova_2019 | SS14 | Pop8 |
| SRR8494416 | PRJNA508872 | SAMN10829687 | CW35 | Czech | Europe | 2013 | Grillova_2019 | Nichols | Pop2 |
| SRR8501161 | PRJNA508872 | SAMN10534861 | CW88 | Czech | Europe | 2017 | Grillova_2019 | SS14 | Pop6 |
| SRR8501162 | PRJNA508872 | SAMN10534852 | CW56 | Cuba | N.America | 2013 | Grillova_2019 | SS14 | Pop6 |
| SRR8501163 | PRJNA508872 | SAMN10534851 | CW30 | Czech | Europe | 2014 | Grillova_2019 | SS14 | Pop6 |
| SRR8501164 | PRJNA508872 | SAMN10534854 | CW65 | Australia | Oceania | 2014 | Grillova_2019 | Nichols | Pop1 |
| SRR8501165 | PRJNA508872 | SAMN10534853 | CW59 | France | Europe | 2012 | Grillova_2019 | Nichols | Pop2 |
| SRR8501166 | PRJNA508872 | SAMN10534856 | CW83 | Cuba | N.America | 2015 | Grillova_2019 | Nichols | Pop5 |
| SRR8501167 | PRJNA508872 | SAMN10534855 | CW82 | Cuba | N.America | 2016 | Grillova_2019 | Nichols | Pop1 |
| SRR8501168 | PRJNA508872 | SAMN10534858 | CW85 | France | Europe | 2016 | Grillova_2019 | SS14 | Pop6 |
| SRR8501169 | PRJNA508872 | SAMN10534857 | CW84 | France | Europe | 2015 | Grillova_2019 | SS14 | Pop6 |
| SRR8501170 | PRJNA508872 | SAMN10534860 | CW87 | France | Europe | 2016 | Grillova_2019 | SS14 | Pop8 |
| SRR8501171 | PRJNA508872 | SAMN10534859 | CW86 | France | Europe | 2013 | Grillova_2019 | Nichols | Pop2 |
| SRR21294940 | PRJNA815321 | SAMN30415008 | A0YIQC9601 | USA | N.America | 2020 | This study | SS14 | Pop8 |
| SRR18326767 | PRJNA815321 | SAMN26583528 | SS14600000 | Colombia | S.America | 2015 | This study | SS14 | Pop8 |
| SRR21294827 | PRJNA815321 | SAMN30415061 | SS15600000 | Colombia | S.America | 2016 | This study | SS14 | Pop8 |
| SRR18326766 | PRJNA815321 | SAMN26583606 | SS16700000 | Colombia | S.America | 2016 | This study | SS14 | Pop7 |
| SRR21294825 | PRJNA815321 | SAMN30415063 | SS19600000 | Colombia | S.America | 2020 | This study | SS14 | Pop8 |
| SRR21294828 | PRJNA815321 | SAMN30415060 | SS20800000 | Colombia | S.America | 2020 | This study | SS14 | Pop8 |
| SRR21294824 | PRJNA815321 | SAMN30415064 | SS21300000 | Colombia | S.America | 2019 | This study | SS14 | Pop8 |
| SRR21294823 | PRJNA815321 | SAMN30415065 | SS21500000 | Colombia | S.America | 2019 | This study | SS14 | Pop7 |
| SRR21294941 | PRJNA815321 | SAMN30415007 | SS2210000A | Colombia | S.America | 2020 | This study | SS14 | Pop8 |
| SRR18326765 | PRJNA815321 | SAMN26583608 | SS2220000A | Colombia | S.America | 2020 | This study | Nichols | Pop1 |
| RR21294820 | PRJNA815321 | SAMN30415067 | TPVCC00170 | Colombia | S.America | 2020 | This study | SS14 | Pop7 |
| SRR21294819 | PRJNA815321 | SAMN30415068 | TPVCC004D0 | Colombia | S.America | 2020 | This study | Nichols | Pop2 |
| SRR21294826 | PRJNA815321 | SAMN30415062 | TPVCC008N0 | Colombia | S.America | 2020 | This study | Nichols | Pop2 |
| SRR21294818 | PRJNA815321 | SAMN30415069 | TPVCC009R0 | Colombia | S.America | 2020 | This study | SS14 | Pop8 |
| SRR21294791 | PRJNA815321 | SAMN30415094 | TPVCC020D0 | Colombia | S.America | 2020 | This study | SS14 | Pop8 |
| SRR21294790 | PRJNA815321 | SAMN30415095 | TPVCC02450 | Colombia | S.America | 2020 | This study | SS14 | Pop7 |
| SRR21294785 | PRJNA815321 | SAMN30415098 | TPVCC03090 | Colombia | S.America | 2021 | This study | SS14 | Pop7 |

| | | | | | | | | | |
|---|---|---|---|---|---|---|---|---|---|
| SRR21294789 | PRJNA815321 | SAMN30415096 | TPVCC03770 | Colombia | S.America | 2021 | This study | SS14 | Pop6 |
| SRR21294786 | PRJNA815321 | SAMN30415097 | TPVCC038T0 | Colombia | S.America | 2021 | This study | Nichols | Pop2 |
| SRR21294954 | PRJNA815321 | SAMN30414898 | TPVCC046TB | Colombia | S.America | 2021 | This study | SS14 | Pop8 |
| SRR21294906 | PRJNA815321 | SAMN30414901 | TPVCC051LS | Colombia | S.America | 2021 | This study | Nichols | Pop2 |
| SRR21294917 | PRJNA815321 | SAMN30414900 | TPVCC060XS | Colombia | S.America | 2021 | This study | SS14 | Pop8 |
| SRR21294845 | PRJNA815321 | SAMN30414992 | TPVCN00490 | China | Asia | 2019 | This study | Nichols | Pop3 |
| SRR23948432 | PRJNA815321 | SAMN33864087 | TPVCN005BB | China | Asia | 2019 | This study | SS14 | Pop7 |
| SRR23948455 | PRJNA815321 | SAMN33864096 | TPVCN006DB | China | Asia | 2020 | This study | Nichols | Pop2 |
| SRR21294952 | PRJNA815321 | SAMN30414997 | TPVCN007F0 | China | Asia | 2020 | This study | SS14 | Pop7 |
| SRR21294951 | PRJNA815321 | SAMN30414998 | TPVCN008H0 | China | Asia | 2020 | This study | Nichols | Pop3 |
| SRR23948443 | PRJNA815321 | SAMN33864107 | TPVCN0105B | China | Asia | 2020 | This study | SS14 | Pop7 |
| SRR23948457 | PRJNA815321 | SAMN33864094 | TPVCN0117B | China | Asia | 2020 | This study | SS14 | Pop7 |
| SRR23948442 | PRJNA815321 | SAMN33864108 | TPVCN0121B | China | Asia | 2020 | This study | SS14 | Pop7 |
| SRR23948431 | PRJNA815321 | SAMN33864088 | TPVCN0133B | China | Asia | 2020 | This study | SS14 | Pop7 |
| SRR21294843 | PRJNA815321 | SAMN30414994 | TPVCN014D0 | China | Asia | 2020 | This study | SS14 | Pop7 |
| SRR23948441 | PRJNA815321 | SAMN33864109 | TPVCN0169B | China | Asia | 2020 | This study | Nichols | Pop3 |
| SRR21294883 | PRJNA815321 | SAMN30415031 | TPVCN017B0 | China | Asia | 2020 | This study | SS14 | Pop7 |
| SRR21294928 | PRJNA815321 | SAMN30415019 | TPVCN019R0 | China | Asia | 2020 | This study | SS14 | Pop7 |
| SRR23948440 | PRJNA815321 | SAMN33864110 | TPVCN0209B | China | Asia | 2020 | This study | SS14 | Pop7 |
| SRR21294887 | PRJNA815321 | SAMN30415027 | TPVCN021B0 | China | Asia | 2020 | This study | SS14 | Pop7 |
| SRR21294936 | PRJNA815321 | SAMN30415012 | TPVCN022D0 | China | Asia | 2020 | This study | SS14 | Pop7 |
| SRR21294932 | PRJNA815321 | SAMN30415016 | TPVCN02410 | China | Asia | 2020 | This study | Nichols | Pop3 |
| SRR23948456 | PRJNA815321 | SAMN33864095 | TPVCN0253B | China | Asia | 2020 | This study | SS14 | Pop7 |
| SRR21294933 | PRJNA815321 | SAMN30415015 | TPVCN02650 | China | Asia | 2020 | This study | SS14 | Pop7 |
| SRR23948427 | PRJNA815321 | SAMN33864092 | TPVCN0277B | China | Asia | 2020 | This study | SS14 | Pop7 |
| SRR23948430 | PRJNA815321 | SAMN33864089 | TPVCN028TS | China | Asia | 2020 | This study | SS14 | Pop7 |
| SRR21294939 | PRJNA815321 | SAMN30415009 | TPVCN029V0 | China | Asia | 2020 | This study | Nichols | Pop2 |
| SRR23948460 | PRJNA815321 | SAMN33864083 | TPVCN030DB | China | Asia | 2020 | This study | SS14 | Pop7 |
| SRR21294890 | PRJNA815321 | SAMN30415025 | TPVCN031F0 | China | Asia | 2020 | This study | SS14 | Pop7 |
| SRR23948448 | PRJNA815321 | SAMN33864085 | TPVCN0329B | China | Asia | 2020 | This study | SS14 | Pop7 |
| SRR23948439 | PRJNA815321 | SAMN33864111 | TPVCN033BB | China | Asia | 2020 | This study | SS14 | Pop7 |
| SRR23948438 | PRJNA815321 | SAMN33864112 | TPVCN0357B | China | Asia | 2021 | This study | SS14 | Pop7 |
| SRR21294891 | PRJNA815321 | SAMN30415024 | TPVCN03610 | China | Asia | 2021 | This study | SS14 | Pop8 |
| SRR21294934 | PRJNA815321 | SAMN30415014 | TPVCN03730 | China | Asia | 2021 | This study | SS14 | Pop7 |
| SRR21294889 | PRJNA815321 | SAMN30415026 | TPVCN038X0 | China | Asia | 2021 | This study | Nichols | Pop3 |
| SRR23948437 | PRJNA815321 | SAMN33864086 | TPVCN039ZS | China | Asia | 2021 | This study | SS14 | Pop7 |
| SRR23948436 | PRJNA815321 | SAMN33864113 | TPVCN040HB | China | Asia | 2021 | This study | SS14 | Pop7 |
| SRR21294754 | PRJNA815321 | SAMN30414904 | TPVCN041L0 | China | Asia | 2022 | This study | SS14 | Pop7 |
| SRR23948435 | PRJNA815321 | SAMN33864114 | TPVCN042NB | China | Asia | 2021 | This study | SS14 | Pop7 |
| SRR23948449 | PRJNA815321 | SAMN33864102 | TPVCN047Z1 | China | Asia | Unknown | This study | SS14 | Pop7 |
| SRR23948447 | PRJNA815321 | SAMN33864103 | TPVCN04931 | China | Asia | 2021 | This study | SS14 | Pop7 |
| SRR23948458 | PRJNA815321 | SAMN33864093 | TPVCN051RS | China | Asia | 2021 | This study | SS14 | Pop7 |
| SRR23948429 | PRJNA815321 | SAMN33864090 | TPVCN052HS | China | Asia | 2021 | This study | SS14 | Pop7 |
| SRR23948452 | PRJNA815321 | SAMN33864099 | TPVCN053LB | China | Asia | 2021 | This study | SS14 | Pop7 |

| | | | | | | | | | |
|---|---|---|---|---|---|---|---|---|---|
| SRR23948453 | PRJNA815321 | SAMN33864098 | TPVCN054XB | China | Asia | 2021 | This study | SS14 | Pop7 |
| SRR23948450 | PRJNA815321 | SAMN33864101 | TPVCN0585B | China | Asia | 2021 | This study | SS14 | Pop7 |
| SRR23948446 | PRJNA815321 | SAMN33864104 | TPVCN061V1 | China | Asia | 2021 | This study | SS14 | Pop7 |
| SRR23948434 | PRJNA815321 | SAMN33864115 | TPVCN062XB | China | Asia | 2021 | This study | SS14 | Pop7 |
| SRR23948454 | PRJNA815321 | SAMN33864097 | TPVCN063ZB | China | Asia | 2021 | This study | SS14 | Pop7 |
| SRR23948445 | PRJNA815321 | SAMN33864105 | TPVCN064H1 | China | Asia | 2021 | This study | SS14 | Pop7 |
| SRR23948428 | PRJNA815321 | SAMN33864091 | TPVCN067RB | China | Asia | 2021 | This study | Nichols | Pop3 |
| SRR23948451 | PRJNA815321 | SAMN33864100 | TPVCN0689B | China | Asia | 2021 | This study | Nichols | Pop3 |
| SRR21294953 | PRJNA815321 | SAMN30414907 | TPVCN070X0 | China | Asia | 2022 | This study | SS14 | Pop7 |
| SRR23948433 | PRJNA815321 | SAMN33864116 | TPVCN072TB | China | Asia | 2022 | This study | SS14 | Pop7 |
| SRR23948459 | PRJNA815321 | SAMN33864084 | TPVCN073VS | China | Asia | 2022 | This study | SS14 | Pop7 |
| SRR21294942 | PRJNA815321 | SAMN30414908 | TPVCN074N0 | China | Asia | 2022 | This study | SS14 | Pop7 |
| SRR23948444 | PRJNA815321 | SAMN33864106 | TPVCN075R1 | China | Asia | 2022 | This study | Nichols | Pop3 |
| SRR21294809 | PRJNA815321 | SAMN30415077 | TPVMW001M1 | Malawi | Africa | 2020 | This study | SS14 | Pop7 |
| SRR21294945 | PRJNA815321 | SAMN30415004 | TPVMW012P0 | Malawi | Africa | 2020 | This study | SS14 | Pop7 |
| SRR21294944 | PRJNA815321 | SAMN30415005 | TPVMW022U1 | Malawi | Africa | 2020 | This study | SS14 | Pop7 |
| SRR21294901 | PRJNA815321 | SAMN30414941 | TPVMW04300 | Malawi | Africa | 2020 | This study | SS14 | Pop8 |
| SRR21294899 | PRJNA815321 | SAMN30414943 | TPVMW04500 | Malawi | Africa | 2020 | This study | SS14 | Pop7 |
| SRR21294852 | PRJNA815321 | SAMN30414986 | TPVMW048P0 | Malawi | Africa | 2020 | This study | SS14 | Pop7 |
| SRR21294946 | PRJNA815321 | SAMN30415003 | TPVMW05340 | Malawi | Africa | 2020 | This study | SS14 | Pop7 |
| SRR21294898 | PRJNA815321 | SAMN30414944 | TPVMW061C0 | Malawi | Africa | 2020 | This study | SS14 | Pop7 |
| SRR21294896 | PRJNA815321 | SAMN30414946 | TPVMW06540 | Malawi | Africa | 2020 | This study | SS14 | Pop8 |
| SRR21294772 | PRJNA815321 | SAMN30414950 | TPVMW075O0 | Malawi | Africa | 2020 | This study | SS14 | Pop7 |
| SRR21294900 | PRJNA815321 | SAMN30414942 | TPVMW07660 | Malawi | Africa | 2020 | This study | SS14 | Pop7 |
| SRR21294949 | PRJNA815321 | SAMN30415000 | TPVMW07740 | Malawi | Africa | 2020 | This study | Nichols | Pop5 |
| SRR21294950 | PRJNA815321 | SAMN30414999 | TPVMW082H0 | Malawi | Africa | 2020 | This study | Nichols | Pop4 |
| SRR21294947 | PRJNA815321 | SAMN30415002 | TPVMW083L0 | Malawi | Africa | 2020 | This study | SS14 | Pop7 |
| SRR21294774 | PRJNA815321 | SAMN30414948 | TPVMW091L0 | Malawi | Africa | 2020 | This study | SS14 | Pop7 |
| SRR21294773 | PRJNA815321 | SAMN30414949 | TPVMW100Y0 | Malawi | Africa | 2020 | This study | SS14 | Pop7 |
| SRR21294771 | PRJNA815321 | SAMN30414951 | TPVMW114K0 | Malawi | Africa | 2020 | This study | SS14 | Pop7 |
| SRR21294775 | PRJNA815321 | SAMN30414947 | TPVMW124Y0 | Malawi | Africa | 2020 | This study | Nichols | Pop5 |
| SRR21294880 | PRJNA815321 | SAMN30415034 | TPVMW12863 | Malawi | Africa | 2020 | This study | SS14 | Pop7 |
| SRR21294770 | PRJNA815321 | SAMN30414952 | TPVMW14460 | Malawi | Africa | 2020 | This study | Nichols | Pop5 |
| SRR21294767 | PRJNA815321 | SAMN30414955 | TPVMW14540 | Malawi | Africa | 2020 | This study | SS14 | Pop8 |
| SRR21294879 | PRJNA815321 | SAMN30415035 | TPVMW149W0 | Malawi | Africa | 2020 | This study | SS14 | Pop7 |
| SRR21294878 | PRJNA815321 | SAMN30415036 | TPVMW16060 | Malawi | Africa | 2020 | This study | SS14 | Pop7 |
| SRR21294876 | PRJNA815321 | SAMN30415037 | TPVMW169M1 | Malawi | Africa | 2020 | This study | SS14 | Pop7 |
| SRR21294764 | PRJNA815321 | SAMN30414957 | TPVMW17100 | Malawi | Africa | 2020 | This study | SS14 | Pop7 |
| SRR21294769 | PRJNA815321 | SAMN30414953 | TPVMW174A0 | Malawi | Africa | 2020 | This study | SS14 | Pop8 |
| SRR21294763 | PRJNA815321 | SAMN30414958 | TPVMW182T0 | Malawi | Africa | 2020 | This study | Nichols | Pop4 |
| SRR21294762 | PRJNA815321 | SAMN30414959 | TPVMW186H0 | Malawi | Africa | 2020 | This study | SS14 | Pop8 |
| SRR21294873 | PRJNA815321 | SAMN30415040 | TPVMW187L1 | Malawi | Africa | 2020 | This study | SS14 | Pop7 |
| SRR21294871 | PRJNA815321 | SAMN30415042 | TPVMW22381 | Malawi | Africa | 2020 | This study | SS14 | Pop7 |
| SRR21294869 | PRJNA815321 | SAMN30415044 | TPVMW23741 | Malawi | Africa | 2020 | This study | SS14 | Pop7 |

| | | | | | | | | | |
|---|---|---|---|---|---|---|---|---|---|
| SRR21294768 | PRJNA815321 | SAMN30414954 | TPVMW238U0 | Malawi | Africa | 2020 | This study | SS14 | Pop7 |
| SRR21294761 | PRJNA815321 | SAMN30414960 | TPVMW244Y0 | Malawi | Africa | 2020 | This study | SS14 | Pop7 |
| SRR21294766 | PRJNA815321 | SAMN30414956 | TPVMW260Y0 | Malawi | Africa | 2020 | This study | SS14 | Pop7 |
| SRR21294867 | PRJNA815321 | SAMN30415046 | TPVMW268E1 | Malawi | Africa | 2020 | This study | SS14 | Pop7 |
| SRR21294864 | PRJNA815321 | SAMN30415048 | TPVMW284D0 | Malawi | Africa | 2020 | This study | SS14 | Pop7 |
| SRR21294840 | PRJNA815321 | SAMN30415049 | TPVMW30701 | Malawi | Africa | 2020 | This study | SS14 | Pop7 |
| SRR21294757 | PRJNA815321 | SAMN30414964 | TPVMW312E0 | Malawi | Africa | 2020 | This study | SS14 | Pop8 |
| SRR21294760 | PRJNA815321 | SAMN30414961 | TPVMW32060 | Malawi | Africa | 2020 | This study | SS14 | Pop7 |
| SRR21294758 | PRJNA815321 | SAMN30414963 | TPVMW325C0 | Malawi | Africa | 2020 | This study | SS14 | Pop7 |
| SRR21294838 | PRJNA815321 | SAMN30415051 | TPVMW337C1 | Malawi | Africa | 2020 | This study | SS14 | Pop7 |
| SRR21294834 | PRJNA815321 | SAMN30415055 | TPVMW339G2 | Malawi | Africa | 2020 | This study | SS14 | Pop7 |
| SRR21294756 | PRJNA815321 | SAMN30414965 | TPVMW343S0 | Malawi | Africa | 2020 | This study | Nichols | Pop5 |
| SRR21294831 | PRJNA815321 | SAMN30415057 | TPVMW351S0 | Malawi | Africa | 2020 | This study | SS14 | Pop7 |
| SRR21294804 | PRJNA815321 | SAMN30415082 | TPVMW39410 | Malawi | Africa | 2021 | This study | SS14 | Pop7 |
| SRR21294751 | PRJNA815321 | SAMN30414969 | TPVMW399V0 | Malawi | Africa | 2021 | This study | SS14 | Pop7 |
| SRR21294803 | PRJNA815321 | SAMN30415083 | TPVMW412N0 | Malawi | Africa | 2021 | This study | SS14 | Pop7 |
| SRR21294755 | PRJNA815321 | SAMN30414966 | TPVMW413R0 | Malawi | Africa | 2021 | This study | SS14 | Pop7 |
| SRR21294753 | PRJNA815321 | SAMN30414967 | TPVMW427L0 | Malawi | Africa | 2021 | This study | SS14 | Pop7 |
| SRR21294801 | PRJNA815321 | SAMN30415085 | TPVMW434H0 | Malawi | Africa | 2021 | This study | SS14 | Pop7 |
| SRR21294800 | PRJNA815321 | SAMN30415086 | TPVMW44050 | Malawi | Africa | 2021 | This study | Nichols | Pop4 |
| SRR21294796 | PRJNA815321 | SAMN30415089 | TPVMW445F0 | Malawi | Africa | 2021 | This study | SS14 | Pop7 |
| SRR21294750 | PRJNA815321 | SAMN30414970 | TPVMW45010 | Malawi | Africa | 2021 | This study | SS14 | Pop8 |
| SRR21294752 | PRJNA815321 | SAMN30414968 | TPVMW459L0 | Malawi | Africa | 2021 | This study | SS14 | Pop8 |
| SRR21294795 | PRJNA815321 | SAMN30415090 | TPVMW468X0 | Malawi | Africa | 2021 | This study | Nichols | Pop4 |
| SRR21294927 | PRJNA815321 | SAMN30414918 | TPVMW469Z0 | Malawi | Africa | 2021 | This study | Nichols | Pop5 |
| SRR21294794 | PRJNA815321 | SAMN30415091 | TPVMW47650 | Malawi | Africa | 2021 | This study | SS14 | Pop7 |
| SRR21294793 | PRJNA815321 | SAMN30415092 | TPVMW478T1 | Malawi | Africa | 2021 | This study | Nichols | Pop4 |
| SRR21294749 | PRJNA815321 | SAMN30414971 | TPVMW479V0 | Malawi | Africa | 2021 | This study | SS14 | Pop8 |
| SRR21294747 | PRJNA815321 | SAMN30414973 | TPVMW505R0 | Malawi | Africa | 2021 | This study | SS14 | Pop7 |
| SRR21294745 | PRJNA815321 | SAMN30414975 | TPVMW516N0 | Malawi | Africa | 2021 | This study | Nichols | Pop5 |
| SRR21294860 | PRJNA815321 | SAMN30414978 | TPVMW56330 | Malawi | Africa | 2021 | This study | Nichols | Pop5 |
| SRR21294863 | PRJNA815321 | SAMN30414976 | TPVMW569R0 | Malawi | Africa | 2021 | This study | SS14 | Pop7 |
| SRR21294861 | PRJNA815321 | SAMN30414977 | TPVMW57010 | Malawi | Africa | 2021 | This study | SS14 | Pop7 |
| SRR21294925 | PRJNA815321 | SAMN30414919 | TPVMW578H0 | Malawi | Africa | 2021 | This study | SS14 | Pop7 |
| SRR21294748 | PRJNA815321 | SAMN30414972 | TPVMW589C0 | Malawi | Africa | 2021 | This study | SS14 | Pop7 |
| SRR21294931 | PRJNA815321 | SAMN30414909 | TPVMW605F0 | Malawi | Africa | 2021 | This study | SS14 | Pop7 |
| SRR21294909 | PRJNA815321 | SAMN30414934 | TPVMW629Z2 | Malawi | Africa | 2021 | This study | SS14 | Pop8 |
| SRR21294856 | PRJNA815321 | SAMN30414982 | TPVMW631B0 | Malawi | Africa | 2021 | This study | SS14 | Pop7 |
| SRR21294888 | PRJNA815321 | SAMN30414910 | TPVMW632D0 | Malawi | Africa | 2021 | This study | SS14 | Pop7 |
| SRR21294924 | PRJNA815321 | SAMN30414920 | TPVMW63650 | Malawi | Africa | 2021 | This study | SS14 | Pop7 |
| SRR21294877 | PRJNA815321 | SAMN30414911 | TPVMW639V2 | Malawi | Africa | 2021 | This study | SS14 | Pop7 |
| SRR21294866 | PRJNA815321 | SAMN30414912 | TPVMW640N0 | Malawi | Africa | 2021 | This study | SS14 | Pop8 |
| SRR21294821 | PRJNA815321 | SAMN30414914 | TPVMW64970 | Malawi | Africa | 2021 | This study | SS14 | Pop7 |
| SRR21294921 | PRJNA815321 | SAMN30414923 | TPVMW654T0 | Malawi | Africa | 2021 | This study | SS14 | Pop7 |

| | | | | | | | | | |
|---|---|---|---|---|---|---|---|---|---|
| SRR21294857 | PRJNA815321 | SAMN30414981 | TPVMW708X0 | Malawi | Africa | 2020 | This study | Nichols | Pop4 |
| SRR21294855 | PRJNA815321 | SAMN30414983 | TPVMW709Z0 | Malawi | Africa | 2020 | This study | Nichols | Pop4 |
| SRR21294920 | PRJNA815321 | SAMN30414924 | TPVMW71530 | Malawi | Africa | 2020 | This study | SS14 | Pop7 |
| SRR21294854 | PRJNA815321 | SAMN30414984 | TPVMW71770 | Malawi | Africa | 2020 | This study | SS14 | Pop8 |
| SRR21294787 | PRJNA815321 | SAMN30414917 | TPVMW719V0 | Malawi | Africa | 2020 | This study | SS14 | Pop7 |
| SRR21294916 | PRJNA815321 | SAMN30414927 | TPVMW72170 | Malawi | Africa | 2020 | This study | SS14 | Pop8 |
| SRR21294915 | PRJNA815321 | SAMN30414928 | TPVMW724D0 | Malawi | Africa | 2020 | This study | SS14 | Pop7 |
| SRR21294914 | PRJNA815321 | SAMN30414929 | TPVMW735B0 | Malawi | Africa | 2020 | This study | SS14 | Pop7 |
| SRR21294912 | PRJNA815321 | SAMN30414931 | TPVMW736D0 | Malawi | Africa | 2020 | This study | SS14 | Pop7 |

**Supplementary Table 2:** Frequency of *TPA* populations sampled in this study by country.

| Population | Lineage | Overall, n | By site, n | | | |
|---|---|---|---|---|---|---|
| | | | China | Colombia | Malawi | USA |
| Pop1 | Nichols | 1 | 0 | 1 | 0 | 0 |
| Pop2 | Nichols | 6 | 2 | 4 | 0 | 0 |
| Pop3 | Nichols | 8 | 8 | 0 | 0 | 0 |
| Pop4 | Nichols | 7 | 0 | 0 | 7 | 0 |
| Pop5 | Nichols | 7 | 0 | 0 | 7 | 0 |
| Pop6 | SS14 | 1 | 0 | 1 | 0 | 0 |
| Pop7 | SS14 | 103 | 41 | 5 | 57 | 0 |
| Pop8 | SS14 | 25 | 1 | 10 | 13 | 1 |

**Supplementary Table 3:** Fixed SNVs identified during comparison of Nichols- (reference sequence) and SS14-lineage *TPA* strains, including functional annotation.

| Genome | Position | Reference | Alternate | Effect | Gene_name | Gene_ID | Nucleotide_change | AA_change |
|---|---|---|---|---|---|---|---|---|
| CP004010.2 | 6442 | G | A | synonymous_variant | gyrA | TPANIC_0005 | c.2052G>A | p.Glu684Glu |
| CP004010.2 | 41132 | C | T | missense_variant | TPANIC_0033 | TPANIC_0033 | c.563G>A | p.Cys188Tyr |
| CP004010.2 | 47527 | C | T | upstream_gene_variant | znuA | TPANIC_0034 | c.-4782G>A | NA |
| CP004010.2 | 73158 | A | G | upstream_gene_variant | dnaB | TPANIC_0058 | c.-4195T>C | NA |
| CP004010.2 | 79965 | T | C | missense_variant | TPANIC_0073 | TPANIC_0073 | c.673A>G | p.Ile225Val |
| CP004010.2 | 88008 | A | C | missense_variant | coxL | TPANIC_0079 | c.959A>C | p.Glu320Ala |
| CP004010.2 | 94075 | C | T | synonymous_variant | TPANIC_0083 | TPANIC_0083 | c.888C>T | p.Tyr296Tyr |
| CP004010.2 | 148115 | A | C | synonymous_variant | TPANIC_0126 | TPANIC_0126 | c.180T>G | p.Gly60Gly |
| CP004010.2 | 150557 | G | C | upstream_gene_variant | TPANIC_0126 | TPANIC_0126 | c.-2263C>G | NA |
| CP004010.2 | 155812 | C | T | missense_variant | TPANIC_0134 | TPANIC_0134 | c.898G>A | p.Ala300Thr |
| CP004010.2 | 156721 | G | A | upstream_gene_variant | TPANIC_0130 | TPANIC_0130 | c.-4479C>T | NAOn |
| CP004010.2 | 158193 | G | A | missense_variant | TPANIC_0136 | TPANIC_0136 | c.251G>A | p.Gly84Glu |
| CP004010.2 | 197405 | A | G | synonymous_variant | TPANIC_0178 | TPANIC_0178 | c.378T>C | p.Asp126Asp |
| CP004010.2 | 198040 | G | A | synonymous_variant | TPANIC_0179 | TPANIC_0179 | c.1626C>T | p.Ala542Ala |
| CP004010.2 | 198186 | C | T | missense_variant | TPANIC_0179 | TPANIC_0179 | c.1480G>A | p.Ala494Thr |
| CP004010.2 | 198269 | T | C | missense_variant | TPANIC_0179 | TPANIC_0179 | c.1397A>G | p.Asp466Gly |
| CP004010.2 | 198334 | A | G | synonymous_variant | TPANIC_0179 | TPANIC_0179 | c.1332T>C | p.Thr444Thr |
| CP004010.2 | 198352 | T | C | synonymous_variant | TPANIC_0179 | TPANIC_0179 | c.1314A>G | p.Val438Val |
| CP004010.2 | 198396 | C | T | missense_variant | TPANIC_0179 | TPANIC_0179 | c.1270G>A | p.Glu424Lys |
| CP004010.2 | 198428 | C | T | missense_variant | TPANIC_0179 | TPANIC_0179 | c.1238G>A | p.Gly413Glu |
| CP004010.2 | 237746 | T | G | upstream_gene_variant | TPANIC_0226 | TPANIC_0226 | c.-836A>C | NA |
| CP004010.2 | 302121 | G | A | missense_variant | TPANIC_0286 | TPANIC_0286 | c.736G>A | p.Gly246Ser |
| CP004010.2 | 312085 | T | A | missense_variant | TPANIC_0297 | TPANIC_0297 | c.742T>A | p.Trp248Arg |
| CP004010.2 | 312087 | G | T | missense_variant | TPANIC_0297 | TPANIC_0297 | c.744G>T | p.Trp248Cys |
| CP004010.2 | 320692 | C | T | missense_variant | TPANIC_0304 | TPANIC_0304 | c.1513G>A | p.Val505Ile |
| CP004010.2 | 320834 | T | C | synonymous_variant | TPANIC_0304 | TPANIC_0304 | c.1371A>G | p.Leu457Leu |
| CP004010.2 | 327426 | G | A | synonymous_variant | TPANIC_0309 | TPANIC_0309 | c.663G>A | p.Pro221Pro |
| CP004010.2 | 327602 | T | C | missense_variant | ssb2 | TPANIC_0310 | c.52T>C | p.Ser18Pro |
| CP004010.2 | 327666 | T | C | missense_variant | ssb2 | TPANIC_0310 | c.116T>C | p.Val39Ala |
| CP004010.2 | 347401 | T | G | missense_variant | tp92 | TPANIC_0326 | c.1967T>G | p.Phe656Cys |
| CP004010.2 | 353352 | T | C | missense_variant | TPANIC_0330 | TPANIC_0330 | c.1589A>G | p.Asn530Ser |
| CP004010.2 | 354963 | G | A | upstream_gene_variant | TPANIC_0330 | TPANIC_0330 | c.-23C>T | NA |
| CP004010.2 | 370050 | A | G | synonymous_variant | trcF | TPANIC_0344 | c.990T>C | p.His330His |
| CP004010.2 | 413262 | A | G | missense_variant | ftsW | TPANIC_0387 | c.601A>G | p.Met201Val |
| CP004010.2 | 421764 | T | C | missense_variant | topA | TPANIC_0394 | c.2000T>C | p.Phe667Ser |
| CP004010.2 | 428595 | C | T | missense_variant | fliI | TPANIC_0402 | c.401C>T | p.Pro134Leu |
| CP004010.2 | 460450 | A | G | missense_variant | TPANIC_0431 | TPANIC_0431 | c.226A>G | p.Ile76Val |
| CP004010.2 | 477634 | G | A | missense_variant | TPANIC_0449 | TPANIC_0449 | c.427G>A | p.Val143Met |
| CP004010.2 | 492772 | A | G | missense_variant | TPANIC_0462 | TPANIC_0462 | c.151A>G | p.Asn51Asp |
| CP004010.2 | 492773 | A | G | missense_variant | TPANIC_0462 | TPANIC_0462 | c.152A>G | p.Asn51Ser |
| CP004010.2 | 503421 | T | C | missense_variant | TPANIC_0473 | TPANIC_0473 | c.633A>G | p.Ile211Met |

| | | | | | | | | |
|---|---|---|---|---|---|---|---|---|
| CP004010.2 | 521090 | C | A | upstream_gene_variant | TPANIC_0484 | TPANIC_0484 | c.-3856G>T | NA |
| CP004010.2 | 521150 | G | A | upstream_gene_variant | TPANIC_0484 | TPANIC_0484 | c.-3916C>T | NA |
| CP004010.2 | 521754 | C | T | upstream_gene_variant | TPANIC_0484 | TPANIC_0484 | c.-4520G>A | NA |
| CP004010.2 | 528814 | A | G | synonymous_variant | dnaG | TPANIC_0492 | c.1152A>G | p.Ser384Ser |
| CP004010.2 | 537571 | C | T | missense_variant | pbp1 | TPANIC_0500 | c.1691C>T | p.Pro564Leu |
| CP004010.2 | 537846 | G | T | missense_variant | rodA | TPANIC_0501 | c.95G>T | p.Gly32Val |
| CP004010.2 | 544427 | G | A | synonymous_variant | clpP1 | TPANIC_0507 | c.390G>A | p.Pro130Pro |
| CP004010.2 | 550331 | T | C | synonymous_variant | ispDF | TPANIC_0512 | c.1122T>C | p.Leu374Leu |
| CP004010.2 | 555872 | G | A | missense_variant | TPANIC_0515 | TPANIC_0515 | c.949G>A | p.Gly317Arg |
| CP004010.2 | 556275 | A | G | missense_variant | TPANIC_0515 | TPANIC_0515 | c.1352A>G | p.Asn451Ser |
| CP004010.2 | 557598 | C | T | missense_variant | TPANIC_0515 | TPANIC_0515 | c.2675C>T | p.Ser892Leu |
| CP004010.2 | 557603 | C | A | missense_variant | TPANIC_0515 | TPANIC_0515 | c.2680C>A | p.Gln894Lys |
| CP004010.2 | 557612 | C | A | missense_variant | TPANIC_0515 | TPANIC_0515 | c.2689C>A | p.Gln897Lys |
| CP004010.2 | 557619 | A | G | missense_variant | TPANIC_0515 | TPANIC_0515 | c.2696A>G | p.Lys899Arg |
| CP004010.2 | 557670 | A | G | missense_variant | TPANIC_0515 | TPANIC_0515 | c.2747A>G | p.Gln916Arg |
| CP004010.2 | 557768 | T | C | missense_variant | TPANIC_0515 | TPANIC_0515 | c.2845T>C | p.Ser949Pro |
| CP004010.2 | 557771 | T | C | missense_variant | TPANIC_0515 | TPANIC_0515 | c.2848T>C | p.Ser950Pro |
| CP004010.2 | 592614 | T | C | synonymous_variant | ispH | TPANIC_0547 | c.513A>G | p.Pro171Pro |
| CP004010.2 | 594571 | C | T | synonymous_variant | TPANIC_0548 | TPANIC_0548 | c.1302C>T | p.Asn434Asn |
| CP004010.2 | 602216 | C | T | missense_variant | gph | TPANIC_0554 | c.445C>T | p.Pro149Ser |
| CP004010.2 | 606165 | T | G | synonymous_variant | nicO | TPANIC_0558 | c.408T>G | p.Val136Val |
| CP004010.2 | 606171 | C | T | synonymous_variant | nicO | TPANIC_0558 | c.414C>T | p.Gly138Gly |
| CP004010.2 | 606229 | T | C | synonymous_variant | nicO | TPANIC_0558 | c.472T>C | p.Leu158Leu |
| CP004010.2 | 606552 | T | C | synonymous_variant | nicO | TPANIC_0558 | c.795T>C | p.Tyr265Tyr |
| CP004010.2 | 606591 | C | T | synonymous_variant | nicO | TPANIC_0558 | c.834C>T | p.Leu278Leu |
| CP004010.2 | 606939 | A | G | missense_variant | thiI | TPANIC_0559 | c.212A>G | p.Tyr71Cys |
| CP004010.2 | 607169 | C | T | missense_variant | thiI | TPANIC_0559 | c.442C>T | p.His148Tyr |
| CP004010.2 | 608437 | C | T | missense_variant | tktA | TPANIC_0560 | c.1535G>A | p.Ser512Asn |
| CP004010.2 | 619959 | T | C | synonymous_variant | pepP | TPANIC_0569 | c.1128A>G | p.Ala376Ala |
| CP004010.2 | 622191 | T | C | missense_variant | TPANIC_0571 | TPANIC_0571 | c.458A>G | p.Glu153Gly |
| CP004010.2 | 623981 | G | T | missense_variant | TPANIC_0574 | TPANIC_0574 | c.1182C>A | p.Ser394Arg |
| CP004010.2 | 630004 | G | A | missense_variant | TPANIC_0577 | TPANIC_0577 | c.1612G>A | p.Ala538Thr |
| CP004010.2 | 642894 | T | C | missense_variant | hprK | TPANIC_0591 | c.676A>G | p.Ile226Val |
| CP004010.2 | 652210 | T | C | missense_variant | TPANIC_0598 | TPANIC_0598 | c.259A>G | p.Thr87Ala |
| CP004010.2 | 667340 | C | T | synonymous_variant | sufB | TPANIC_0612 | c.1413C>T | p.Ser471Ser |
| CP004010.2 | 708563 | G | A | missense_variant | lysS1 | TPANIC_0644 | c.64C>T | p.Arg22Cys |
| CP004010.2 | 730935 | C | T | missense_variant | TPANIC_0667 | TPANIC_0667 | c.437G>A | p.Arg146Gln |
| CP004010.2 | 749757 | A | G | upstream_gene_variant | gcp | TPANIC_0680 | c.-3191T>C | NA |
| CP004010.2 | 799933 | A | G | upstream_gene_variant | flgE | TPANIC_0727 | c.-4996T>C | NA |
| CP004010.2 | 803821 | A | G | synonymous_variant | TPANIC_0736 | TPANIC_0736 | c.49T>C | p.Leu17Leu |
| CP004010.2 | 821958 | T | G | synonymous_variant | fmt | TPANIC_0756 | c.933A>C | p.Leu311Leu |
| CP004010.2 | 874149 | C | T | missense_variant | TPANIC_0804 | TPANIC_0804 | c.80C>T | p.Ala27Val |
| CP004010.2 | 916216 | T | C | missense_variant | htrA2 | TPANIC_0841 | c.21A>G | p.Ile7Met |
| CP004010.2 | 936178 | G | A | synonymous_variant | TPANIC_0858 | TPANIC_0858 | c.168G>A | p.Arg56Arg |
| CP004010.2 | 937180 | C | T | synonymous_variant | TPANIC_0858 | TPANIC_0858 | c.1170C>T | p.Gly390Gly |

| | | | | | | | | |
|---|---|---|---|---|---|---|---|---|
| CP004010.2 | 948473 | A | G | missense_variant | TPANIC_0867 | TPANIC_0867 | c.113T>C | p.Val38Ala |
| CP004010.2 | 948666 | T | C | upstream_gene_variant | TPANIC_0864 | TPANIC_0864 | c.-3733A>G | NA |
| CP004010.2 | 951595 | G | A | missense_variant | fliD | TPANIC_0872 | c.548G>A | p.Ser183Asn |
| CP004010.2 | 955916 | T | C | synonymous_variant | TPANIC_0877 | TPANIC_0877 | c.651T>C | p.Phe217Phe |
| CP004010.2 | 956280 | C | A | missense_variant | TPANIC_0877 | TPANIC_0877 | c.1015C>A | p.Gln339Lys |
| CP004010.2 | 977900 | A | G | synonymous_variant | recB | TPANIC_0898 | c.3258T>C | p.Leu1086Leu |
| CP004010.2 | 992764 | A | G | missense_variant | TPANIC_0912 | TPANIC_0912 | c.967A>G | p.Asn323Asp |
| CP004010.2 | 992765 | A | C | missense_variant | TPANIC_0912 | TPANIC_0912 | c.968A>C | p.Asn323Thr |
| CP004010.2 | 1050223 | G | A | missense_variant | TPANIC_0966 | TPANIC_0966 | c.560C>T | p.Thr187Ile |
| CP004010.2 | 1050442 | G | A | missense_variant | TPANIC_0966 | TPANIC_0966 | c.341C>T | p.Ala114Val |
| CP004010.2 | 1059461 | T | C | synonymous_variant | pheS | TPANIC_0973 | c.1506T>C | p.Gly502Gly |
| CP004010.2 | 1081597 | G | A | missense_variant | TPANIC_0995 | TPANIC_0995 | c.478G>A | p.Ala160Thr |
| CP004010.2 | 1102208 | A | G | upstream_gene_variant | gyrB | TPANIC_1006 | c.-3666T>C | NA |
| CP004010.2 | 1103911 | T | G | upstream_gene_variant | thyX | TPANIC_1007 | c.-4343A>C | NA |
| CP004010.2 | 1125380 | C | T | synonymous_variant | TPANIC_1029 | TPANIC_1029 | c.51G>A | p.Gln17Gln |
| CP004010.2 | 1125555 | A | G | upstream_gene_variant | uvrD | TPANIC_1028 | c.-894T>C | NA |
| CP004010.2 | 1125741 | C | G | upstream_gene_variant | uvrD | TPANIC_1028 | c.-1080G>C | NA |

**Supplementary Table 4:** Highly differentiated (Fisher exact $p < 4.451 \times 10^{-6}$) SNVs identified during comparison of Nichols- (reference sequence) and SS14-lineage *TPA* strains, including functional annotation.

| Genome | Position | Ref | Alt | Effect | Gene_name | Gene_ID | Nucleotide_change | AA_change | Fisher P_value |
|---|---|---|---|---|---|---|---|---|---|
| CP004010.2 | 1453 | G | A | upstream_gene_variant | dnaN | TPANIC_0002 | c.-188G>A | NA | 2.60E-06 |
| CP004010.2 | 7984 | C | T | missense_variant | TPANIC_0007 | TPANIC_0007 | c.668C>T | p.Ala223Val | 1.12E-80 |
| CP004010.2 | 25345 | G | A | missense_variant | TPANIC_0021 | TPANIC_0021 | c.233G>A | p.Arg78His | 1.61E-79 |
| CP004010.2 | 92037 | T | C | upstream_gene_variant | TPANIC_0083 | TPANIC_0083 | c.-1151T>C | NA | 9.59E-38 |
| CP004010.2 | 92232 | G | A | synonymous_variant | fhlA | TPANIC_0082 | c.210C>T | p.Leu70Leu | 8.81E-38 |
| CP004010.2 | 99385 | G | A | synonymous_variant | murB | TPANIC_0090 | c.435C>T | p.Ile145Ile | 2.65E-20 |
| CP004010.2 | 101869 | A | G | missense_variant | rpoE | TPANIC_0092 | c.236A>G | p.Glu79Gly | 9.71E-65 |
| CP004010.2 | 126059 | A | G | missense_variant | TPANIC_0110 | TPANIC_0110 | c.868A>G | p.Asn290Asp | 5.57E-61 |
| CP004010.2 | 128280 | A | G | synonymous_variant | rpoN | TPANIC_0111 | c.1296A>G | p.Gln432Gln | 1.44E-10 |
| CP004010.2 | 146729 | C | T | upstream_gene_variant | pckG | TPANIC_0122 | c.-3837G>A | NA | 2.24E-54 |
| CP004010.2 | 158138 | A | G | missense_variant | TPANIC_0136 | TPANIC_0136 | c.196A>G | p.Ser66Gly | 3.92E-48 |
| CP004010.2 | 158149 | C | T | synonymous_variant | TPANIC_0136 | TPANIC_0136 | c.207C>T | p.Tyr69Tyr | 4.51E-55 |
| CP004010.2 | 158161 | C | T | synonymous_variant | TPANIC_0136 | TPANIC_0136 | c.219C>T | p.Gly73Gly | 6.30E-57 |
| CP004010.2 | 158989 | C | A | missense_variant | TPANIC_0136 | TPANIC_0136 | c.1047C>A | p.Asp349Glu | 2.86E-54 |
| CP004010.2 | 159162 | C | T | missense_variant | TPANIC_0136 | TPANIC_0136 | c.1220C>T | p.Ser407Leu | 1.62E-54 |
| CP004010.2 | 168817 | T | C | missense_variant | fadD1 | TPANIC_0145 | c.1763T>C | p.Val588Ala | 2.20E-45 |
| CP004010.2 | 169148 | G | A | upstream_gene_variant | TPANIC_0142 | TPANIC_0142 | c.-4942C>T | NA | 1.16E-168 |
| CP004010.2 | 170012 | T | C | upstream_gene_variant | TPANIC_0143 | TPANIC_0143 | c.-4032A>G | NA | 1.04E-35 |
| CP004010.2 | 174137 | C | T | missense_variant | mfD | TPANIC_0151 | c.778G>A | p.Val260Ile | 3.76E-216 |
| CP004010.2 | 179167 | A | G | synonymous_variant | TPANIC_0155 | TPANIC_0155 | c.888A>G | p.Ala296Ala | 2.43E-79 |
| CP004010.2 | 184144 | C | A | missense_variant | proS | TPANIC_0160 | c.280G>T | p.Ala94Ser | 4.97E-152 |
| CP004010.2 | 187064 | A | G | synonymous_variant | troB | TPANIC_0164 | c.228A>G | p.Lys76Lys | 1.90E-168 |
| CP004010.2 | 187073 | A | G | synonymous_variant | troB | TPANIC_0164 | c.237A>G | p.Arg79Arg | 1.90E-168 |
| CP004010.2 | 187177 | T | G | missense_variant | troB | TPANIC_0164 | c.341T>G | p.Leu114Arg | 1.16E-168 |
| CP004010.2 | 189935 | A | G | missense_variant | troR | TPANIC_0167 | c.314A>G | p.Glu105Gly | 6.60E-21 |
| CP004010.2 | 198194 | G | C | missense_variant | TPANIC_0179 | TPANIC_0179 | c.1472C>G | p.Ala491Gly | 1.39E-168 |
| CP004010.2 | 202514 | A | G | missense_variant | lepB1 | TPANIC_0185 | c.290A>G | p.Asn97Ser | 8.40E-129 |
| CP004010.2 | 205724 | T | A | upstream_gene_variant | TPANIC_0183 | TPANIC_0183 | c.-4056A>T | NA | 1.86E-54 |
| CP004010.2 | 228320 | G | C | missense_variant | TPANIC_0222 | TPANIC_0222 | c.199G>C | p.Ala67Pro | 1.11E-129 |
| CP004010.2 | 263037 | G | A | synonymous_variant | flaA1 | TPANIC_0249 | c.438G>A | p.Gln146Gln | 1.44E-10 |
| CP004010.2 | 279291 | A | G | synonymous_variant | TPANIC_0265 | TPANIC_0265 | c.138T>C | p.Thr46Thr | 2.75E-168 |
| CP004010.2 | 287491 | C | T | missense_variant | pcnB1 | TPANIC_0270 | c.232C>T | p.His78Tyr | 1.95E-08 |
| CP004010.2 | 288142 | C | T | missense_variant | pcnB1 | TPANIC_0270 | c.883C>T | p.Pro295Ser | 1.98E-79 |
| CP004010.2 | 288872 | C | A | missense_variant | parB | TPANIC_0271 | c.796G>T | p.Asp266Tyr | 1.90E-168 |
| CP004010.2 | 299301 | G | T | missense_variant | coaD | TPANIC_0283 | c.151G>T | p.Asp51Tyr | 7.07E-15 |
| CP004010.2 | 319648 | C | T | missense_variant | TPANIC_0304 | TPANIC_0304 | c.2557G>A | p.Val853Met | 3.30E-168 |
| CP004010.2 | 324869 | A | G | missense_variant | TPANIC_0307 | TPANIC_0307 | c.140A>G | p.Asp47Gly | 1.47E-80 |
| CP004010.2 | 336910 | C | T | synonymous_variant | tmpC | TPANIC_0319 | c.930C>T | p.Asp310Asp | 4.49E-24 |
| CP004010.2 | 344101 | G | A | missense_variant | TPANIC_0324 | TPANIC_0324 | c.3055G>A | p.Val1019Met | 7.34E-38 |
| CP004010.2 | 347052 | C | T | missense_variant | tp92 | TPANIC_0326 | c.1618C>T | p.His540Tyr | 2.51E-15 |
| CP004010.2 | 358153 | A | G | missense_variant | TPANIC_0334 | TPANIC_0334 | c.622A>G | p.Thr208Ala | 2.43E-79 |

| | | | | | | | | | |
|---|---|---|---|---|---|---|---|---|---|
| CP004010.2 | 372926 | A | G | missense_variant | TPANIC_0346 | TPANIC_0346 | c.487A>G | p.Lys163Glu | 1.29E-128 |
| CP004010.2 | 384678 | G | T | synonymous_variant | TPANIC_0361 | TPANIC_0361 | c.255C>A | p.Pro85Pro | 2.31E-45 |
| CP004010.2 | 395778 | T | C | synonymous_variant | TPANIC_0370 | TPANIC_0370 | c.186A>G | p.Ala62Ala | 1.12E-140 |
| CP004010.2 | 401798 | C | T | upstream_gene_variant | fliL1 | TPANIC_0377 | c.-1188C>T | NA | 1.60E-21 |
| CP004010.2 | 407061 | C | A | missense_variant | TPANIC_0380 | TPANIC_0380 | c.1181G>T | p.Cys394Phe | 2.65E-79 |
| CP004010.2 | 415693 | G | A | missense_variant | ftsA | TPANIC_0389 | c.1072G>A | p.Gly358Ser | 6.26E-164 |
| CP004010.2 | 420034 | A | C | missense_variant | topA | TPANIC_0394 | c.270A>C | p.Gln90His | 6.36E-167 |
| CP004010.2 | 444557 | C | T | synonymous_variant | ffh | TPANIC_0416 | c.648G>A | p.Leu216Leu | 7.86E-14 |
| CP004010.2 | 463703 | A | G | missense_variant | nlpE | TPANIC_0435 | c.374T>C | p.Ile125Thr | 2.90E-79 |
| CP004010.2 | 492583 | A | G | upstream_gene_variant | TPANIC_0456 | TPANIC_0456 | c.-4609T>C | NA | 9.79E-108 |
| CP004010.2 | 492826 | T | C | missense_variant | TPANIC_0462 | TPANIC_0462 | c.205T>C | p.Ser69Pro | 8.29E-84 |
| CP004010.2 | 492905 | A | T | missense_variant | TPANIC_0462 | TPANIC_0462 | c.284A>T | p.Tyr95Phe | 7.51E-44 |
| CP004010.2 | 493030 | G | A | missense_variant | TPANIC_0462 | TPANIC_0462 | c.409G>A | p.Asp137Asn | 1.31E-104 |
| CP004010.2 | 493057 | G | A | missense_variant | TPANIC_0462 | TPANIC_0462 | c.436G>A | p.Glu146Lys | 2.94E-128 |
| CP004010.2 | 493060 | C | T | missense_variant | TPANIC_0462 | TPANIC_0462 | c.439C>T | p.Leu147Phe | 2.56E-128 |
| CP004010.2 | 493061 | T | C | missense_variant | TPANIC_0462 | TPANIC_0462 | c.440T>C | p.Leu147Pro | 4.38E-128 |
| CP004010.2 | 493112 | A | G | missense_variant | TPANIC_0462 | TPANIC_0462 | c.491A>G | p.Asp164Gly | 7.92E-156 |
| CP004010.2 | 493204 | A | G | missense_variant | TPANIC_0462 | TPANIC_0462 | c.583A>G | p.Ser195Gly | 4.38E-157 |
| CP004010.2 | 493216 | A | G | missense_variant | TPANIC_0462 | TPANIC_0462 | c.595A>G | p.Ser199Gly | 4.38E-157 |
| CP004010.2 | 493237 | G | A | missense_variant | TPANIC_0462 | TPANIC_0462 | c.616G>A | p.Gly206Ser | 1.52E-153 |
| CP004010.2 | 493285 | G | A | missense_variant | TPANIC_0462 | TPANIC_0462 | c.664G>A | p.Gly222Arg | 3.67E-146 |
| CP004010.2 | 493288 | A | G | missense_variant | TPANIC_0462 | TPANIC_0462 | c.667A>G | p.Lys223Glu | 3.67E-146 |
| CP004010.2 | 493308 | A | G | synonymous_variant | TPANIC_0462 | TPANIC_0462 | c.687A>G | p.Leu229Leu | 6.68E-146 |
| CP004010.2 | 493358 | A | G | missense_variant | TPANIC_0462 | TPANIC_0462 | c.737A>G | p.Asn246Ser | 2.64E-158 |
| CP004010.2 | 493374 | T | C | synonymous_variant | TPANIC_0462 | TPANIC_0462 | c.753T>C | p.Ala251Ala | 6.87E-161 |
| CP004010.2 | 493394 | C | A | missense_variant | TPANIC_0462 | TPANIC_0462 | c.773C>A | p.Thr258Asn | 1.32E-160 |
| CP004010.2 | 493531 | A | G | missense_variant | TPANIC_0462 | TPANIC_0462 | c.910A>G | p.Thr304Ala | 1.18E-159 |
| CP004010.2 | 493549 | G | C | missense_variant | TPANIC_0462 | TPANIC_0462 | c.928G>C | p.Glu310Gln | 9.06E-155 |
| CP004010.2 | 493577 | G | A | missense_variant | TPANIC_0462 | TPANIC_0462 | c.956G>A | p.Gly319Asp | 4.57E-150 |
| CP004010.2 | 493582 | G | A | missense_variant | TPANIC_0462 | TPANIC_0462 | c.961G>A | p.Asp321Asn | 4.57E-150 |
| CP004010.2 | 493601 | A | G | missense_variant | TPANIC_0462 | TPANIC_0462 | c.980A>G | p.Asp327Gly | 4.57E-150 |
| CP004010.2 | 493602 | T | G | missense_variant | TPANIC_0462 | TPANIC_0462 | c.981T>G | p.Asp327Glu | 5.38E-150 |
| CP004010.2 | 493605 | G | C | missense_variant | TPANIC_0462 | TPANIC_0462 | c.984G>C | p.Arg328Ser | 1.57E-152 |
| CP004010.2 | 502857 | A | C | missense_variant | uvrC | TPANIC_0472 | c.478T>G | p.Leu160Val | 4.38E-61 |
| CP004010.2 | 506432 | C | G | missense_variant | pgi | TPANIC_0475 | c.172G>C | p.Glu58Gln | 2.90E-79 |
| CP004010.2 | 514619 | T | C | missense_variant | TPANIC_0483 | TPANIC_0483 | c.614A>G | p.His205Arg | 2.90E-79 |
| CP004010.2 | 514720 | G | A | synonymous_variant | TPANIC_0483 | TPANIC_0483 | c.513C>T | p.Asn171Asn | 3.17E-79 |
| CP004010.2 | 514823 | G | A | missense_variant | TPANIC_0483 | TPANIC_0483 | c.410C>T | p.Thr137Met | 4.27E-78 |
| CP004010.2 | 514885 | C | T | synonymous_variant | TPANIC_0483 | TPANIC_0483 | c.348G>A | p.Pro116Pro | 1.98E-79 |
| CP004010.2 | 514894 | C | T | synonymous_variant | TPANIC_0483 | TPANIC_0483 | c.339G>A | p.Gly113Gly | 2.16E-79 |
| CP004010.2 | 514897 | T | C | synonymous_variant | TPANIC_0483 | TPANIC_0483 | c.336A>G | p.Gly112Gly | 2.16E-79 |
| CP004010.2 | 515326 | C | T | missense_variant | TPANIC_0484 | TPANIC_0484 | c.1909G>A | p.Glu637Lys | 1.61E-79 |
| CP004010.2 | 523234 | T | C | missense_variant | mcp2 | TPANIC_0488 | c.776T>C | p.Val259Ala | 2.04E-126 |
| CP004010.2 | 538516 | G | T | missense_variant | rodA | TPANIC_0501 | c.765G>T | p.Leu255Phe | 3.23E-164 |
| CP004010.2 | 554556 | C | A | missense_variant | uvrA | TPANIC_0514 | c.2512C>A | p.Arg838Ser | 2.60E-06 |

| | | | | | | | | | |
|---|---|---|---|---|---|---|---|---|---|
| CP004010.2 | 556289 | C | T | missense_variant | TPANIC_0515 | TPANIC_0515 | c.1366C>T | p.Arg456Cys | 3.03E-218 |
| CP004010.2 | 573135 | C | T | missense_variant | ntpB2 | TPANIC_0528 | c.817G>A | p.Gly273Ser | 6.79E-78 |
| CP004010.2 | 573412 | C | T | missense_variant | ntpB2 | TPANIC_0528 | c.540G>A | p.Met180Ile | 9.83E-122 |
| CP004010.2 | 574682 | C | T | missense_variant | ntpA2 | TPANIC_0529 | c.1084G>A | p.Glu362Lys | 1.43E-126 |
| CP004010.2 | 579791 | T | C | missense_variant | TPANIC_0535 | TPANIC_0535 | c.161A>G | p.Glu54Gly | 1.90E-168 |
| CP004010.2 | 583866 | C | T | synonymous_variant | era | TPANIC_0541 | c.87C>T | p.Ala29Ala | 1.90E-130 |
| CP004010.2 | 593477 | C | A | missense_variant | TPANIC_0548 | TPANIC_0548 | c.208C>A | p.Gln70Lys | 1.60E-114 |
| CP004010.2 | 594930 | G | T | missense_variant | clpA1 | TPANIC_0549 | c.259G>T | p.Val87Phe | 2.99E-170 |
| CP004010.2 | 600356 | G | A | upstream_gene_variant | trmE | TPANIC_0550 | c.-1712C>T | NA | 1.77E-21 |
| CP004010.2 | 623599 | C | A | synonymous_variant | TPANIC_0572 | TPANIC_0572 | c.150G>T | p.Gly50Gly | 8.25E-132 |
| CP004010.2 | 626141 | G | A | missense_variant | ptsI | TPANIC_0575 | c.868G>A | p.Val290Met | 2.65E-79 |
| CP004010.2 | 657057 | C | A | synonymous_variant | uppS | TPANIC_0603 | c.135G>T | p.Val45Val | 5.23E-44 |
| CP004010.2 | 659548 | G | A | missense_variant | rpsB | TPANIC_0606 | c.143C>T | p.Thr48Met | 4.14E-44 |
| CP004010.2 | 665921 | A | G | upstream_gene_variant | TPANIC_0608 | TPANIC_0608 | c.-4975T>C | NA | 2.27E-54 |
| CP004010.2 | 670222 | A | G | missense_variant | TPANIC_0615 | TPANIC_0615 | c.419A>G | p.His140Arg | 2.65E-79 |
| CP004010.2 | 678701 | T | C | missense_variant | TPANIC_0623 | TPANIC_0623 | c.626T>C | p.Leu209Ser | 2.56E-130 |
| CP004010.2 | 686052 | G | A | synonymous_variant | sbcC | TPANIC_0627 | c.2283G>A | p.Ala761Ala | 1.33E-72 |
| CP004010.2 | 698264 | T | C | upstream_gene_variant | recO | TPANIC_0636 | c.-1010A>G | NA | 9.12E-154 |
| CP004010.2 | 710654 | T | C | synonymous_variant | serS | TPANIC_0647 | c.753A>G | p.Leu251Leu | 3.61E-13 |
| CP004010.2 | 737426 | C | T | missense_variant | TPANIC_0671 | TPANIC_0671 | c.674G>A | p.Arg225His | 1.36E-167 |
| CP004010.2 | 753234 | G | A | missense_variant | mglC | TPANIC_0686 | c.538G>A | p.Ala180Thr | 1.56E-79 |
| CP004010.2 | 760212 | T | C | missense_variant | scpA | TPANIC_0691 | c.89A>G | p.Lys30Arg | 8.79E-212 |
| CP004010.2 | 773215 | T | C | missense_variant | mrcA | TPANIC_0705 | c.1873A>G | p.Met625Val | 1.85E-261 |
| CP004010.2 | 773571 | G | A | missense_variant | mrcA | TPANIC_0705 | c.1517C>T | p.Ala506Val | 2.63E-35 |
| CP004010.2 | 773572 | C | T | missense_variant | mrcA | TPANIC_0705 | c.1516G>A | p.Ala506Thr | 3.68E-229 |
| CP004010.2 | 779707 | C | T | missense_variant | TPANIC_0710 | TPANIC_0710 | c.1423C>T | p.Arg475Trp | 4.99E-39 |
| CP004010.2 | 800209 | A | G | missense_variant | TPANIC_0733 | TPANIC_0733 | c.217A>G | p.Asn73Asp | 2.31E-45 |
| CP004010.2 | 800227 | G | A | missense_variant | TPANIC_0733 | TPANIC_0733 | c.235G>A | p.Gly79Ser | 1.81E-21 |
| CP004010.2 | 800587 | T | C | missense_variant | TPANIC_0733 | TPANIC_0733 | c.595T>C | p.Ser199Pro | 1.11E-33 |
| CP004010.2 | 818570 | A | G | synonymous_variant | TPANIC_0752 | TPANIC_0752 | c.846T>C | p.Ser282Ser | 7.01E-15 |
| CP004010.2 | 825069 | A | T | missense_variant | pbp2 | TPANIC_0760 | c.1243A>T | p.Ile415Phe | 5.46E-08 |
| CP004010.2 | 829582 | A | G | missense_variant | TPANIC_0764 | TPANIC_0764 | c.766A>G | p.Arg256Gly | 2.16E-45 |
| CP004010.2 | 834017 | T | C | synonymous_variant | fusA2 | TPANIC_0767 | c.720T>C | p.Ile240Ile | 4.14E-44 |
| CP004010.2 | 842145 | A | G | missense_variant | TPANIC_0772 | TPANIC_0772 | c.811A>G | p.Ser271Gly | 8.01E-151 |
| CP004010.2 | 856051 | T | C | synonymous_variant | TPANIC_0789 | TPANIC_0789 | c.585A>G | p.Lys195Lys | 2.90E-79 |
| CP004010.2 | 860410 | G | A | synonymous_variant | flaB1 | TPANIC_0792 | c.717G>A | p.Gln239Gln | 9.07E-18 |
| CP004010.2 | 867908 | A | G | synonymous_variant | metG | TPANIC_0798 | c.198T>C | p.Pro66Pro | 8.39E-38 |
| CP004010.2 | 868731 | C | T | missense_variant | dacC | TPANIC_0800 | c.299C>T | p.Ala100Val | 1.54E-130 |
| CP004010.2 | 873743 | C | A | synonymous_variant | TPANIC_0803 | TPANIC_0803 | c.960C>A | p.Ile320Ile | 1.13E-20 |
| CP004010.2 | 877816 | C | T | synonymous_variant | TPANIC_0806 | TPANIC_0806 | c.1041G>A | p.Ser347Ser | 4.34E-196 |
| CP004010.2 | 906101 | C | A | missense_variant | TPANIC_0835 | TPANIC_0835 | c.577G>T | p.Ala193Ser | 2.69E-167 |
| CP004010.2 | 919401 | A | G | missense_variant | gap | TPANIC_0844 | c.1046A>G | p.Lys349Arg | 1.81E-79 |
| CP004010.2 | 930862 | T | C | missense_variant | TPANIC_0854 | TPANIC_0854 | c.328A>G | p.Arg110Gly | 3.15E-11 |
| CP004010.2 | 936177 | G | A | missense_variant | TPANIC_0858 | TPANIC_0858 | c.167G>A | p.Arg56Gln | 3.25E-112 |
| CP004010.2 | 936431 | G | A | missense_variant | TPANIC_0858 | TPANIC_0858 | c.421G>A | p.Asp141Asn | 3.07E-06 |
| CP004010.2 | 936841 | C | A | missense_variant | TPANIC_0858 | TPANIC_0858 | c.831C>A | p.Asn277Lys | 6.50E-170 |

| | | | | | | | | | |
|---|---|---|---|---|---|---|---|---|---|
| CP004010.2 | 936851 | A | T | missense_variant | TPANIC_0858 | TPANIC_0858 | c.841A>T | p.Thr281Ser | 6.50E-170 |
| CP004010.2 | 937077 | T | C | missense_variant | TPANIC_0858 | TPANIC_0858 | c.1067T>C | p.Met356Thr | 4.58E-14 |
| CP004010.2 | 942290 | G | A | missense_variant | nifS | TPANIC_0863 | c.340G>A | p.Val114Ile | 2.27E-54 |
| CP004010.2 | 945224 | C | T | missense_variant | TPANIC_0865 | TPANIC_0865 | c.1183G>A | p.Asp395Asn | 2.75E-168 |
| CP004010.2 | 945308 | A | G | missense_variant | TPANIC_0865 | TPANIC_0865 | c.1099T>C | p.Ser367Pro | 2.75E-168 |
| CP004010.2 | 945331 | T | C | missense_variant | TPANIC_0865 | TPANIC_0865 | c.1076A>G | p.Asn359Ser | 2.29E-168 |
| CP004010.2 | 945520 | C | G | missense_variant | TPANIC_0865 | TPANIC_0865 | c.887G>C | p.Gly296Ala | 1.90E-168 |
| CP004010.2 | 945526 | A | G | missense_variant | TPANIC_0865 | TPANIC_0865 | c.881T>C | p.Met294Thr | 9.62E-169 |
| CP004010.2 | 945527 | T | C | missense_variant | TPANIC_0865 | TPANIC_0865 | c.880A>G | p.Met294Val | 9.62E-169 |
| CP004010.2 | 945541 | T | C | missense_variant | TPANIC_0865 | TPANIC_0865 | c.866A>G | p.Asn289Ser | 1.16E-168 |
| CP004010.2 | 945542 | T | C | missense_variant | TPANIC_0865 | TPANIC_0865 | c.865A>G | p.Asn289Asp | 1.16E-168 |
| CP004010.2 | 945838 | T | G | missense_variant | TPANIC_0865 | TPANIC_0865 | c.569A>C | p.Glu190Ala | 8.47E-162 |
| CP004010.2 | 945845 | T | G | missense_variant | TPANIC_0865 | TPANIC_0865 | c.562A>C | p.Thr188Pro | 7.10E-162 |
| CP004010.2 | 945851 | A | G | missense_variant | TPANIC_0865 | TPANIC_0865 | c.556T>C | p.Tyr186His | 7.10E-162 |
| CP004010.2 | 945853 | G | A | missense_variant | TPANIC_0865 | TPANIC_0865 | c.554C>T | p.Pro185Leu | 5.00E-162 |
| CP004010.2 | 946138 | A | G | missense_variant | TPANIC_0865 | TPANIC_0865 | c.269T>C | p.Val90Ala | 8.01E-122 |
| CP004010.2 | 946142 | G | A | missense_variant | TPANIC_0865 | TPANIC_0865 | c.265C>T | p.Pro89Ser | 8.01E-122 |
| CP004010.2 | 951392 | T | C | synonymous_variant | fliD | TPANIC_0872 | c.345T>C | p.Pro115Pro | 4.42E-167 |
| CP004010.2 | 952802 | T | C | synonymous_variant | fliD | TPANIC_0872 | c.1755T>C | p.Ile585Ile | 1.54E-130 |
| CP004010.2 | 994358 | A | G | synonymous_variant | TPANIC_0915 | TPANIC_0915 | c.243A>G | p.Thr81Thr | 8.90E-18 |
| CP004010.2 | 998026 | A | C | missense_variant | TPANIC_0919 | TPANIC_0919 | c.74T>G | p.Phe25Cys | 1.42E-153 |
| CP004010.2 | 1018974 | C | T | synonymous_variant | TPANIC_0935 | TPANIC_0935 | c.54G>A | p.Lys18Lys | 8.81E-38 |
| CP004010.2 | 1043721 | C | T | synonymous_variant | flgG1 | TPANIC_0960 | c.195G>A | p.Gly65Gly | 1.23E-80 |
| CP004010.2 | 1050211 | G | A | missense_variant | TPANIC_0966 | TPANIC_0966 | c.572C>T | p.Ser191Leu | 1.56E-167 |
| CP004010.2 | 1050214 | G | A | missense_variant | TPANIC_0966 | TPANIC_0966 | c.569C>T | p.Pro190Leu | 1.25E-152 |
| CP004010.2 | 1050217 | G | A | missense_variant | TPANIC_0966 | TPANIC_0966 | c.566C>T | p.Thr189Ile | 1.48E-152 |
| CP004010.2 | 1050335 | C | T | missense_variant | TPANIC_0966 | TPANIC_0966 | c.448G>A | p.Gly150Arg | 3.61E-70 |
| CP004010.2 | 1050712 | G | T | missense_variant | TPANIC_0966 | TPANIC_0966 | c.71C>A | p.Ala24Glu | 1.56E-45 |
| CP004010.2 | 1050800 | T | C | upstream_gene_variant | TPANIC_0962 | TPANIC_0962 | c.-4676A>G | NA | 7.71E-38 |
| CP004010.2 | 1051947 | G | T | missense_variant | TPANIC_0967 | TPANIC_0967 | c.436C>A | p.Gln146Lys | 6.68E-44 |
| CP004010.2 | 1054981 | A | G | synonymous_variant | TPANIC_0969 | TPANIC_0969 | c.637T>C | p.Leu213Leu | 1.81E-79 |
| CP004010.2 | 1057406 | C | G | missense_variant | ftr1 | TPANIC_0972 | c.463G>C | p.Asp155His | 8.81E-38 |
| CP004010.2 | 1060933 | G | A | missense_variant | TPANIC_0976 | TPANIC_0976 | c.61G>A | p.Val21Met | 8.75E-38 |
| CP004010.2 | 1072433 | T | C | missense_variant | TPANIC_0986 | TPANIC_0986 | c.250T>C | p.Phe84Leu | 1.90E-168 |
| CP004010.2 | 1085138 | G | A | missense_variant | sppA | TPANIC_0997 | c.922C>T | p.Arg308Cys | 9.19E-38 |
| CP004010.2 | 1093589 | G | A | synonymous_variant | TPANIC_1003 | TPANIC_1003 | c.477G>A | p.Leu159Leu | 1.65E-21 |
| CP004010.2 | 1118569 | C | T | missense_variant | recX | TPANIC_1023 | c.4G>A | p.Gly2Ser | 1.04E-06 |

**Supplementary Table 5:** Genes with multiple distinct missense mutations by *TPA* lineage and population, and including within-population allele frequencies.

| Gene | Position | Nucleotide_change | AA_change | FastBAPS_population | Lineage | Allele_frq | Allele_count | Total_count |
|---|---|---|---|---|---|---|---|---|
| TP0136 | 158138 | c.196A>G | p.Ser66Gly | Pop1 | Nichols | 0.115 | 3 | 26 |
| | | | | Pop2 | Nichols | 0 | 0 | 124 |
| | | | | Pop3 | Nichols | 0 | 0 | 95 |
| | | | | Pop4 | Nichols | 0 | 0 | 43 |
| | | | | Pop5 | Nichols | 0.918 | 56 | 61 |
| | | | | Pop6 | SS14 | 0 | 0 | 362 |
| | | | | Pop7 | SS14 | 0.034 | 11 | 327 |
| | | | | Pop8 | SS14 | 0.011 | 4 | 365 |
| TP0136 | 158989 | c.1047C>A | p.Asp349Glu | Pop1 | Nichols | 0 | 0 | 26 |
| | | | | Pop2 | Nichols | 0 | 0 | 123 |
| | | | | Pop3 | Nichols | 0 | 0 | 96 |
| | | | | Pop4 | Nichols | 0.976 | 42 | 43 |
| | | | | Pop5 | Nichols | 0 | 0 | 61 |
| | | | | Pop6 | SS14 | 0 | 0 | 362 |
| | | | | Pop7 | SS14 | 0 | 0 | 329 |
| | | | | Pop8 | SS14 | 0 | 0 | 365 |
| TP0136 | 159162 | c.1220C>T | p.Ser407Leu | Pop1 | Nichols | 0 | 0 | 24 |
| | | | | Pop2 | Nichols | 0 | 0 | 124 |
| | | | | Pop3 | Nichols | 0 | 0 | 96 |
| | | | | Pop4 | Nichols | 0.976 | 42 | 43 |
| | | | | Pop5 | Nichols | 0 | 0 | 61 |
| | | | | Pop6 | SS14 | 0 | 0 | 362 |
| | | | | Pop7 | SS14 | 0 | 0 | 329 |
| | | | | Pop8 | SS14 | 0 | 0 | 370 |
| TP0462 | 492826 | c.205T>C | p.Ser69Pro | Pop1 | Nichols | 0 | 0 | 26 |
| | | | | Pop2 | Nichols | 0.936 | 58 | 62 |
| | | | | Pop3 | Nichols | 0 | 0 | 96 |
| | | | | Pop4 | Nichols | 0 | 0 | 43 |
| | | | | Pop5 | Nichols | 0 | 0 | 61 |
| | | | | Pop6 | SS14 | 0 | 0 | 362 |
| | | | | Pop7 | SS14 | 0 | 0 | 329 |
| | | | | Pop8 | SS14 | 0 | 0 | 371 |
| TP0462 | 492905 | c.284A>T | p.Tyr95Phe | Pop1 | Nichols | 0 | 0 | 26 |
| | | | | Pop2 | Nichols | 0.833 | 30 | 36 |
| | | | | Pop3 | Nichols | 0 | 0 | 96 |
| | | | | Pop4 | Nichols | 0 | 0 | 43 |

| | | | | | | | | |
|---|---|---|---|---|---|---|---|---|
| | | | | Pop5 | Nichols | 0 | 0 | 61 |
| | | | | Pop6 | SS14 | 0 | 0 | 362 |
| | | | | Pop7 | SS14 | 0 | 0 | 329 |
| | | | | Pop8 | SS14 | 0 | 0 | 366 |
| TP0462 | 493030 | c.409G>A | p.Asp137Asn | Pop1 | Nichols | 0 | 0 | 26 |
| | | | | Pop2 | Nichols | 0.938 | 75 | 80 |
| | | | | Pop3 | Nichols | 0 | 0 | 95 |
| | | | | Pop4 | Nichols | 0 | 0 | 43 |
| | | | | Pop5 | Nichols | 0 | 0 | 61 |
| | | | | Pop6 | SS14 | 0 | 0 | 362 |
| | | | | Pop7 | SS14 | 0 | 0 | 329 |
| | | | | Pop8 | SS14 | 0 | 0 | 364 |
| TP0462 | 493057 | c.436G>A | p.Glu146Lys | Pop1 | Nichols | 0 | 0 | 26 |
| | | | | Pop2 | Nichols | 0.969 | 93 | 96 |
| | | | | Pop3 | Nichols | 0 | 0 | 95 |
| | | | | Pop4 | Nichols | 0 | 0 | 43 |
| | | | | Pop5 | Nichols | 0 | 0 | 61 |
| | | | | Pop6 | SS14 | 0 | 0 | 362 |
| | | | | Pop7 | SS14 | 0 | 0 | 329 |
| | | | | Pop8 | SS14 | 0 | 0 | 363 |
| TP0462 | 493060 | c.439C>T | p.Leu147Phe | Pop1 | Nichols | 0 | 0 | 26 |
| | | | | Pop2 | Nichols | 0.969 | 93 | 96 |
| | | | | Pop3 | Nichols | 0 | 0 | 95 |
| | | | | Pop4 | Nichols | 0 | 0 | 43 |
| | | | | Pop5 | Nichols | 0 | 0 | 61 |
| | | | | Pop6 | SS14 | 0 | 0 | 362 |
| | | | | Pop7 | SS14 | 0 | 0 | 329 |
| | | | | Pop8 | SS14 | 0 | 0 | 364 |
| TP0462 | 493061 | c.440T>C | p.Leu147Pro | Pop1 | Nichols | 0 | 0 | 26 |
| | | | | Pop2 | Nichols | 0.959 | 93 | 97 |
| | | | | Pop3 | Nichols | 0 | 0 | 95 |
| | | | | Pop4 | Nichols | 0 | 0 | 43 |
| | | | | Pop5 | Nichols | 0 | 0 | 61 |
| | | | | Pop6 | SS14 | 0 | 0 | 362 |
| | | | | Pop7 | SS14 | 0 | 0 | 329 |
| | | | | Pop8 | SS14 | 0 | 0 | 364 |
| TP0462 | 493112 | c.491A>G | p.Asp164Gly | Pop1 | Nichols | 0 | 0 | 26 |
| | | | | Pop2 | Nichols | 0.983 | 114 | 116 |
| | | | | Pop3 | Nichols | 0 | 0 | 95 |

| | | | | | | | | |
|---|---|---|---|---|---|---|---|---|
| | | | | Pop4 | Nichols | 0 | 0 | 43 |
| | | | | Pop5 | Nichols | 0 | 0 | 61 |
| | | | | Pop6 | SS14 | 0 | 0 | 362 |
| | | | | Pop7 | SS14 | 0 | 0 | 329 |
| | | | | Pop8 | SS14 | 0 | 0 | 362 |
| TP0462 | 493204 | c.583A>G | p.Ser195Gly | Pop1 | Nichols | 0 | 0 | 26 |
| | | | | Pop2 | Nichols | 0.991 | 115 | 116 |
| | | | | Pop3 | Nichols | 0 | 0 | 96 |
| | | | | Pop4 | Nichols | 0 | 0 | 43 |
| | | | | Pop5 | Nichols | 0 | 0 | 61 |
| | | | | Pop6 | SS14 | 0 | 0 | 362 |
| | | | | Pop7 | SS14 | 0 | 0 | 329 |
| | | | | Pop8 | SS14 | 0 | 0 | 370 |
| TP0462 | 493216 | c.595A>G | p.Ser199Gly | Pop1 | Nichols | 0 | 0 | 26 |
| | | | | Pop2 | Nichols | 0.991 | 115 | 116 |
| | | | | Pop3 | Nichols | 0 | 0 | 96 |
| | | | | Pop4 | Nichols | 0 | 0 | 43 |
| | | | | Pop5 | Nichols | 0 | 0 | 61 |
| | | | | Pop6 | SS14 | 0 | 0 | 362 |
| | | | | Pop7 | SS14 | 0 | 0 | 329 |
| | | | | Pop8 | SS14 | 0 | 0 | 370 |
| TP0462 | 493237 | c.616G>A | p.Gly206Ser | Pop1 | Nichols | 0 | 0 | 26 |
| | | | | Pop2 | Nichols | 0.991 | 112 | 113 |
| | | | | Pop3 | Nichols | 0 | 0 | 96 |
| | | | | Pop4 | Nichols | 0 | 0 | 42 |
| | | | | Pop5 | Nichols | 0 | 0 | 61 |
| | | | | Pop6 | SS14 | 0 | 0 | 362 |
| | | | | Pop7 | SS14 | 0 | 0 | 329 |
| | | | | Pop8 | SS14 | 0 | 0 | 368 |
| TP0462 | 493285 | c.664G>A | p.Gly222Arg | Pop1 | Nichols | 0 | 0 | 26 |
| | | | | Pop2 | Nichols | 0.982 | 106 | 108 |
| | | | | Pop3 | Nichols | 0 | 0 | 96 |
| | | | | Pop4 | Nichols | 0 | 0 | 42 |
| | | | | Pop5 | Nichols | 0 | 0 | 61 |
| | | | | Pop6 | SS14 | 0 | 0 | 362 |
| | | | | Pop7 | SS14 | 0 | 0 | 329 |
| | | | | Pop8 | SS14 | 0 | 0 | 372 |
| TP0462 | 493288 | c.667A>G | p.Lys223Glu | Pop1 | Nichols | 0 | 0 | 26 |
| | | | | Pop2 | Nichols | 0.982 | 106 | 108 |

| | | | | | | | | |
|---|---|---|---|---|---|---|---|---|
| | | | | Pop3 | Nichols | 0 | 0 | 96 |
| | | | | Pop4 | Nichols | 0 | 0 | 42 |
| | | | | Pop5 | Nichols | 0 | 0 | 61 |
| | | | | Pop6 | SS14 | 0 | 0 | 362 |
| | | | | Pop7 | SS14 | 0 | 0 | 329 |
| | | | | Pop8 | SS14 | 0 | 0 | 372 |
| TP0462 | 493358 | c.737A>G | p.Asn246Ser | Pop1 | Nichols | 0 | 0 | 26 |
| | | | | Pop2 | Nichols | 0.992 | 116 | 117 |
| | | | | Pop3 | Nichols | 0 | 0 | 96 |
| | | | | Pop4 | Nichols | 0 | 0 | 43 |
| | | | | Pop5 | Nichols | 0 | 0 | 61 |
| | | | | Pop6 | SS14 | 0 | 0 | 362 |
| | | | | Pop7 | SS14 | 0 | 0 | 329 |
| | | | | Pop8 | SS14 | 0 | 0 | 370 |
| TP0462 | 493394 | c.773C>A | p.Thr258Asn | Pop1 | Nichols | 0 | 0 | 26 |
| | | | | Pop2 | Nichols | 0.983 | 118 | 120 |
| | | | | Pop3 | Nichols | 0 | 0 | 96 |
| | | | | Pop4 | Nichols | 0 | 0 | 43 |
| | | | | Pop5 | Nichols | 0 | 0 | 61 |
| | | | | Pop6 | SS14 | 0 | 0 | 362 |
| | | | | Pop7 | SS14 | 0 | 0 | 329 |
| | | | | Pop8 | SS14 | 0 | 0 | 372 |
| TP0462 | 493531 | c.910A>G | p.Thr304Ala | Pop1 | Nichols | 0 | 0 | 26 |
| | | | | Pop2 | Nichols | 0.991 | 116 | 117 |
| | | | | Pop3 | Nichols | 0 | 0 | 96 |
| | | | | Pop4 | Nichols | 0 | 0 | 43 |
| | | | | Pop5 | Nichols | 0 | 0 | 61 |
| | | | | Pop6 | SS14 | 0 | 0 | 362 |
| | | | | Pop7 | SS14 | 0 | 0 | 329 |
| | | | | Pop8 | SS14 | 0 | 0 | 370 |
| TP0462 | 493549 | c.928G>C | p.Glu310Gln | Pop1 | Nichols | 0 | 0 | 26 |
| | | | | Pop2 | Nichols | 0.991 | 112 | 113 |
| | | | | Pop3 | Nichols | 0 | 0 | 96 |
| | | | | Pop4 | Nichols | 0 | 0 | 43 |
| | | | | Pop5 | Nichols | 0 | 0 | 61 |
| | | | | Pop6 | SS14 | 0 | 0 | 361 |
| | | | | Pop7 | SS14 | 0 | 0 | 329 |
| | | | | Pop8 | SS14 | 0 | 0 | 370 |
| TP0462 | 493577 | c.956G>A | p.Gly319Asp | Pop1 | Nichols | 0 | 0 | 26 |

| | | | | | | | | |
|---|---|---|---|---|---|---|---|---|
| | | | | Pop2 | Nichols | 0.991 | 108 | 109 |
| | | | | Pop3 | Nichols | 0 | 0 | 96 |
| | | | | Pop4 | Nichols | 0 | 0 | 43 |
| | | | | Pop5 | Nichols | 0 | 0 | 61 |
| | | | | Pop6 | SS14 | 0 | 0 | 362 |
| | | | | Pop7 | SS14 | 0 | 0 | 329 |
| | | | | Pop8 | SS14 | 0 | 0 | 372 |
| TP0462 | 493582 | c.961G>A | p.Asp321Asn | Pop1 | Nichols | 0 | 0 | 26 |
| | | | | Pop2 | Nichols | 0.991 | 108 | 109 |
| | | | | Pop3 | Nichols | 0 | 0 | 96 |
| | | | | Pop4 | Nichols | 0 | 0 | 43 |
| | | | | Pop5 | Nichols | 0 | 0 | 61 |
| | | | | Pop6 | SS14 | 0 | 0 | 362 |
| | | | | Pop7 | SS14 | 0 | 0 | 329 |
| | | | | Pop8 | SS14 | 0 | 0 | 372 |
| TP0462 | 493601 | c.980A>G | p.Asp327Gly | Pop1 | Nichols | 0 | 0 | 26 |
| | | | | Pop2 | Nichols | 0.991 | 108 | 109 |
| | | | | Pop3 | Nichols | 0 | 0 | 96 |
| | | | | Pop4 | Nichols | 0 | 0 | 43 |
| | | | | Pop5 | Nichols | 0 | 0 | 61 |
| | | | | Pop6 | SS14 | 0 | 0 | 362 |
| | | | | Pop7 | SS14 | 0 | 0 | 329 |
| | | | | Pop8 | SS14 | 0 | 0 | 372 |
| TP0462 | 493602 | c.981T>G | p.Asp327Glu | Pop1 | Nichols | 0 | 0 | 26 |
| | | | | Pop2 | Nichols | 0.991 | 108 | 109 |
| | | | | Pop3 | Nichols | 0 | 0 | 96 |
| | | | | Pop4 | Nichols | 0 | 0 | 43 |
| | | | | Pop5 | Nichols | 0 | 0 | 61 |
| | | | | Pop6 | SS14 | 0 | 0 | 362 |
| | | | | Pop7 | SS14 | 0 | 0 | 329 |
| | | | | Pop8 | SS14 | 0 | 0 | 371 |
| TP0462 | 493605 | c.984G>C | p.Arg328Ser | Pop1 | Nichols | 0 | 0 | 26 |
| | | | | Pop2 | Nichols | 0.982 | 109 | 111 |
| | | | | Pop3 | Nichols | 0 | 0 | 96 |
| | | | | Pop4 | Nichols | 0 | 0 | 43 |
| | | | | Pop5 | Nichols | 0 | 0 | 61 |
| | | | | Pop6 | SS14 | 0 | 0 | 362 |
| | | | | Pop7 | SS14 | 0 | 0 | 329 |
| | | | | Pop8 | SS14 | 0 | 0 | 372 |

| | | | | | | | | |
|---|---|---|---|---|---|---|---|---|
| TP0483 | 514619 | c.614A>G | p.His205Arg | Pop1 | Nichols | 0 | 0 | 26 |
| | | | | Pop2 | Nichols | 0 | 0 | 124 |
| | | | | Pop3 | Nichols | 0 | 0 | 96 |
| | | | | Pop4 | Nichols | 0 | 0 | 43 |
| | | | | Pop5 | Nichols | 0.984 | 60 | 61 |
| | | | | Pop6 | SS14 | 0 | 0 | 362 |
| | | | | Pop7 | SS14 | 0 | 0 | 328 |
| | | | | Pop8 | SS14 | 0 | 0 | 371 |
| TP0483 | 514823 | c.410C>T | p.Thr137Met | Pop1 | Nichols | 0 | 0 | 26 |
| | | | | Pop2 | Nichols | 0 | 0 | 124 |
| | | | | Pop3 | Nichols | 0 | 0 | 96 |
| | | | | Pop4 | Nichols | 0 | 0 | 43 |
| | | | | Pop5 | Nichols | 0.966 | 58 | 60 |
| | | | | Pop6 | SS14 | 0 | 0 | 362 |
| | | | | Pop7 | SS14 | 0 | 0 | 328 |
| | | | | Pop8 | SS14 | 0 | 0 | 372 |
| TP0515 | 556289 | c.1366C>T | p.Arg456Cys | Pop1 | Nichols | 0 | 0 | 26 |
| | | | | Pop2 | Nichols | 0 | 0 | 124 |
| | | | | Pop3 | Nichols | 0 | 0 | 96 |
| | | | | Pop4 | Nichols | 0 | 0 | 43 |
| | | | | Pop5 | Nichols | 0 | 0 | 61 |
| | | | | Pop6 | SS14 | 1 | 362 | 362 |
| | | | | Pop7 | SS14 | 0.003 | 1 | 328 |
| | | | | Pop8 | SS14 | 0.692 | 252 | 364 |
| TP0858 | 936177 | c.167G>A | p.Arg56Gln | Pop1 | Nichols | 0 | 0 | 25 |
| | | | | Pop2 | Nichols | 0 | 0 | 123 |
| | | | | Pop3 | Nichols | 1 | 95 | 95 |
| | | | | Pop4 | Nichols | 0 | 0 | 43 |
| | | | | Pop5 | Nichols | 0 | 0 | 61 |
| | | | | Pop6 | SS14 | 0.003 | 1 | 361 |
| | | | | Pop7 | SS14 | 0.006 | 2 | 329 |
| | | | | Pop8 | SS14 | 0.003 | 1 | 363 |
| TP0858 | 936431 | c.421G>A | p.Asp141Asn | Pop1 | Nichols | 0.269 | 7 | 26 |
| | | | | Pop2 | Nichols | 0 | 0 | 124 |
| | | | | Pop3 | Nichols | 0 | 0 | 96 |
| | | | | Pop4 | Nichols | 0 | 0 | 43 |
| | | | | Pop5 | Nichols | 0 | 0 | 61 |
| | | | | Pop6 | SS14 | 0 | 0 | 362 |
| | | | | Pop7 | SS14 | 0.009 | 3 | 329 |
| | | | | Pop8 | SS14 | 0 | 0 | 370 |

| | | | | | | | | | |
|---|---|---|---|---|---|---|---|---|---|
| TP0858 | 936841 | c.831C>A | p.Asn277Lys | | Pop1 | Nichols | 0 | 0 | 17 |
| | | | | | Pop2 | Nichols | 0.992 | 123 | 124 |
| | | | | | Pop3 | Nichols | 0 | 0 | 96 |
| | | | | | Pop4 | Nichols | 0 | 0 | 41 |
| | | | | | Pop5 | Nichols | 0 | 0 | 61 |
| | | | | | Pop6 | SS14 | 0 | 0 | 355 |
| | | | | | Pop7 | SS14 | 0 | 0 | 329 |
| | | | | | Pop8 | SS14 | 0 | 0 | 371 |
| TP0858 | 936851 | c.841A>T | p.Thr281Ser | | Pop1 | Nichols | 0 | 0 | 17 |
| | | | | | Pop2 | Nichols | 0.992 | 123 | 124 |
| | | | | | Pop3 | Nichols | 0 | 0 | 96 |
| | | | | | Pop4 | Nichols | 0 | 0 | 41 |
| | | | | | Pop5 | Nichols | 0 | 0 | 61 |
| | | | | | Pop6 | SS14 | 0 | 0 | 355 |
| | | | | | Pop7 | SS14 | 0 | 0 | 329 |
| | | | | | Pop8 | SS14 | 0 | 0 | 371 |
| TP0858 | 937077 | c.1067T>C | p.Met356Thr | | Pop1 | Nichols | 0 | 0 | 26 |
| | | | | | Pop2 | Nichols | 0 | 0 | 124 |
| | | | | | Pop3 | Nichols | 0 | 0 | 96 |
| | | | | | Pop4 | Nichols | 0 | 0 | 43 |
| | | | | | Pop5 | Nichols | 0 | 0 | 61 |
| | | | | | Pop6 | SS14 | 0.116 | 42 | 362 |
| | | | | | Pop7 | SS14 | 0 | 0 | 329 |
| | | | | | Pop8 | SS14 | 0.030 | 11 | 365 |
| TP0865 | 945224 | c.1183G>A | p.Asp395Asn | | Pop1 | Nichols | 0 | 0 | 26 |
| | | | | | Pop2 | Nichols | 0.992 | 123 | 124 |
| | | | | | Pop3 | Nichols | 0 | 0 | 96 |
| | | | | | Pop4 | Nichols | 0 | 0 | 43 |
| | | | | | Pop5 | Nichols | 0 | 0 | 61 |
| | | | | | Pop6 | SS14 | 0 | 0 | 362 |
| | | | | | Pop7 | SS14 | 0 | 0 | 329 |
| | | | | | Pop8 | SS14 | 0 | 0 | 370 |
| TP0865 | 945308 | c.1099T>C | p.Ser367Pro | | Pop1 | Nichols | 0 | 0 | 26 |
| | | | | | Pop2 | Nichols | 0.992 | 123 | 124 |
| | | | | | Pop3 | Nichols | 0 | 0 | 96 |
| | | | | | Pop4 | Nichols | 0 | 0 | 43 |
| | | | | | Pop5 | Nichols | 0 | 0 | 61 |
| | | | | | Pop6 | SS14 | 0 | 0 | 362 |
| | | | | | Pop7 | SS14 | 0 | 0 | 329 |

| | | | | | | | | |
|---|---|---|---|---|---|---|---|---|
| | | | | Pop8 | SS14 | 0 | 0 | 370 |
| TP0865 | 945331 | c.1076A>G | p.Asn359Ser | Pop1 | Nichols | 0 | 0 | 26 |
| | | | | Pop2 | Nichols | 0.992 | 123 | 124 |
| | | | | Pop3 | Nichols | 0 | 0 | 96 |
| | | | | Pop4 | Nichols | 0 | 0 | 43 |
| | | | | Pop5 | Nichols | 0 | 0 | 61 |
| | | | | Pop6 | SS14 | 0 | 0 | 362 |
| | | | | Pop7 | SS14 | 0 | 0 | 329 |
| | | | | Pop8 | SS14 | 0 | 0 | 371 |
| TP0865 | 945520 | c.887G>C | p.Gly296Ala | Pop1 | Nichols | 0 | 0 | 26 |
| | | | | Pop2 | Nichols | 0.992 | 123 | 124 |
| | | | | Pop3 | Nichols | 0 | 0 | 96 |
| | | | | Pop4 | Nichols | 0 | 0 | 43 |
| | | | | Pop5 | Nichols | 0 | 0 | 61 |
| | | | | Pop6 | SS14 | 0 | 0 | 362 |
| | | | | Pop7 | SS14 | 0 | 0 | 329 |
| | | | | Pop8 | SS14 | 0 | 0 | 372 |
| TP0865 | 945526 | c.881T>C | p.Met294Thr | Pop1 | Nichols | 0 | 0 | 26 |
| | | | | Pop2 | Nichols | 0.992 | 123 | 124 |
| | | | | Pop3 | Nichols | 0 | 0 | 96 |
| | | | | Pop4 | Nichols | 0 | 0 | 43 |
| | | | | Pop5 | Nichols | 0 | 0 | 61 |
| | | | | Pop6 | SS14 | 0 | 0 | 362 |
| | | | | Pop7 | SS14 | 0 | 0 | 329 |
| | | | | Pop8 | SS14 | 0 | 0 | 372 |
| TP0865 | 945527 | c.880A>G | p.Met294Val | Pop1 | Nichols | 0 | 0 | 26 |
| | | | | Pop2 | Nichols | 0.992 | 123 | 124 |
| | | | | Pop3 | Nichols | 0 | 0 | 96 |
| | | | | Pop4 | Nichols | 0 | 0 | 43 |
| | | | | Pop5 | Nichols | 0 | 0 | 61 |
| | | | | Pop6 | SS14 | 0 | 0 | 362 |
| | | | | Pop7 | SS14 | 0 | 0 | 329 |
| | | | | Pop8 | SS14 | 0 | 0 | 372 |
| TP0865 | 945541 | c.866A>G | p.Asn289Ser | Pop1 | Nichols | 0 | 0 | 26 |
| | | | | Pop2 | Nichols | 0.992 | 123 | 124 |
| | | | | Pop3 | Nichols | 0 | 0 | 96 |
| | | | | Pop4 | Nichols | 0 | 0 | 43 |
| | | | | Pop5 | Nichols | 0 | 0 | 61 |
| | | | | Pop6 | SS14 | 0 | 0 | 362 |

| | | | | | | | | | |
|---|---|---|---|---|---|---|---|---|---|
| | | | | | Pop7 | SS14 | 0 | 0 | 329 |
| | | | | | Pop8 | SS14 | 0 | 0 | 371 |
| TP0865 | 945542 | c.865A>G | p.Asn289Asp | | Pop1 | Nichols | 0 | 0 | 26 |
| | | | | | Pop2 | Nichols | 0.992 | 123 | 124 |
| | | | | | Pop3 | Nichols | 0 | 0 | 96 |
| | | | | | Pop4 | Nichols | 0 | 0 | 43 |
| | | | | | Pop5 | Nichols | 0 | 0 | 61 |
| | | | | | Pop6 | SS14 | 0 | 0 | 362 |
| | | | | | Pop7 | SS14 | 0 | 0 | 329 |
| | | | | | Pop8 | SS14 | 0 | 0 | 371 |
| TP0865 | 945838 | c.569A>C | p.Glu190Ala | | Pop1 | Nichols | 0 | 0 | 26 |
| | | | | | Pop2 | Nichols | 0.992 | 119 | 120 |
| | | | | | Pop3 | Nichols | 0 | 0 | 96 |
| | | | | | Pop4 | Nichols | 0 | 0 | 43 |
| | | | | | Pop5 | Nichols | 0 | 0 | 61 |
| | | | | | Pop6 | SS14 | 0 | 0 | 362 |
| | | | | | Pop7 | SS14 | 0 | 0 | 329 |
| | | | | | Pop8 | SS14 | 0 | 0 | 368 |
| TP0865 | 945845 | c.562A>C | p.Thr188Pro | | Pop1 | Nichols | 0 | 0 | 26 |
| | | | | | Pop2 | Nichols | 0.992 | 119 | 120 |
| | | | | | Pop3 | Nichols | 0 | 0 | 96 |
| | | | | | Pop4 | Nichols | 0 | 0 | 43 |
| | | | | | Pop5 | Nichols | 0 | 0 | 61 |
| | | | | | Pop6 | SS14 | 0 | 0 | 362 |
| | | | | | Pop7 | SS14 | 0 | 0 | 329 |
| | | | | | Pop8 | SS14 | 0 | 0 | 369 |
| TP0865 | 945851 | c.556T>C | p.Tyr186His | | Pop1 | Nichols | 0 | 0 | 26 |
| | | | | | Pop2 | Nichols | 0.992 | 119 | 120 |
| | | | | | Pop3 | Nichols | 0 | 0 | 96 |
| | | | | | Pop4 | Nichols | 0 | 0 | 43 |
| | | | | | Pop5 | Nichols | 0 | 0 | 61 |
| | | | | | Pop6 | SS14 | 0 | 0 | 362 |
| | | | | | Pop7 | SS14 | 0 | 0 | 329 |
| | | | | | Pop8 | SS14 | 0 | 0 | 369 |
| TP0865 | 945853 | c.554C>T | p.Pro185Leu | | Pop1 | Nichols | 0 | 0 | 26 |
| | | | | | Pop2 | Nichols | 0.992 | 119 | 120 |
| | | | | | Pop3 | Nichols | 0 | 0 | 96 |
| | | | | | Pop4 | Nichols | 0 | 0 | 43 |
| | | | | | Pop5 | Nichols | 0 | 0 | 61 |

| | | | | | | | | |
|---|---|---|---|---|---|---|---|---|
| | | | | Pop6 | SS14 | 0 | 0 | 362 |
| | | | | Pop7 | SS14 | 0 | 0 | 329 |
| | | | | Pop8 | SS14 | 0 | 0 | 371 |
| TP0865 | 946138 | c.269T>C | p.Val90Ala | Pop1 | Nichols | 0 | 0 | 26 |
| | | | | Pop2 | Nichols | 0.967 | 87 | 90 |
| | | | | Pop3 | Nichols | 0 | 0 | 96 |
| | | | | Pop4 | Nichols | 0 | 0 | 43 |
| | | | | Pop5 | Nichols | 0 | 0 | 61 |
| | | | | Pop6 | SS14 | 0 | 0 | 362 |
| | | | | Pop7 | SS14 | 0 | 0 | 329 |
| | | | | Pop8 | SS14 | 0 | 0 | 372 |
| TP0865 | 946142 | c.265C>T | p.Pro89Ser | Pop1 | Nichols | 0 | 0 | 26 |
| | | | | Pop2 | Nichols | 0.967 | 87 | 90 |
| | | | | Pop3 | Nichols | 0 | 0 | 96 |
| | | | | Pop4 | Nichols | 0 | 0 | 43 |
| | | | | Pop5 | Nichols | 0 | 0 | 61 |
| | | | | Pop6 | SS14 | 0 | 0 | 362 |
| | | | | Pop7 | SS14 | 0 | 0 | 329 |
| | | | | Pop8 | SS14 | 0 | 0 | 372 |
| TP0966 | 1050211 | c.572C>T | p.Ser191Leu | Pop1 | Nichols | 0 | 0 | 25 |
| | | | | Pop2 | Nichols | 0.991 | 122 | 123 |
| | | | | Pop3 | Nichols | 0 | 0 | 96 |
| | | | | Pop4 | Nichols | 0 | 0 | 43 |
| | | | | Pop5 | Nichols | 0 | 0 | 61 |
| | | | | Pop6 | SS14 | 0 | 0 | 362 |
| | | | | Pop7 | SS14 | 0 | 0 | 329 |
| | | | | Pop8 | SS14 | 0 | 0 | 372 |
| TP0966 | 1050214 | c.569C>T | p.Pro190Leu | Pop1 | Nichols | 0 | 0 | 25 |
| | | | | Pop2 | Nichols | 0.919 | 113 | 123 |
| | | | | Pop3 | Nichols | 0 | 0 | 96 |
| | | | | Pop4 | Nichols | 0 | 0 | 43 |
| | | | | Pop5 | Nichols | 0 | 0 | 61 |
| | | | | Pop6 | SS14 | 0 | 0 | 362 |
| | | | | Pop7 | SS14 | 0 | 0 | 329 |
| | | | | Pop8 | SS14 | 0 | 0 | 372 |
| TP0966 | 1050217 | c.566C>T | p.Thr189Ile | Pop1 | Nichols | 0 | 0 | 25 |
| | | | | Pop2 | Nichols | 0.919 | 113 | 123 |
| | | | | Pop3 | Nichols | 0 | 0 | 96 |
| | | | | Pop4 | Nichols | 0 | 0 | 43 |

| | | | | | | | | | |
|---|---|---|---|---|---|---|---|---|---|
| | | | | | Pop5 | Nichols | 0 | 0 | 61 |
| | | | | | Pop6 | SS14 | 0 | 0 | 362 |
| | | | | | Pop7 | SS14 | 0 | 0 | 329 |
| | | | | | Pop8 | SS14 | 0 | 0 | 371 |
| TP0966 | 1050335 | c.448G>A | p.Gly150Arg | | Pop1 | Nichols | 0 | 0 | 26 |
| | | | | | Pop2 | Nichols | 0 | 0 | 123 |
| | | | | | Pop3 | Nichols | 0 | 0 | 96 |
| | | | | | Pop4 | Nichols | 0 | 0 | 43 |
| | | | | | Pop5 | Nichols | 0.902 | 55 | 61 |
| | | | | | Pop6 | SS14 | 0 | 0 | 362 |
| | | | | | Pop7 | SS14 | 0 | 0 | 329 |
| | | | | | Pop8 | SS14 | 0 | 0 | 372 |
| TP0966 | 1050712 | c.71C>A | p.Ala24Glu | | Pop1 | Nichols | 0 | 0 | 25 |
| | | | | | Pop2 | Nichols | 0 | 0 | 123 |
| | | | | | Pop3 | Nichols | 0 | 0 | 96 |
| | | | | | Pop4 | Nichols | 0 | 0 | 43 |
| | | | | | Pop5 | Nichols | 0.610 | 36 | 59 |
| | | | | | Pop6 | SS14 | 0 | 0 | 359 |
| | | | | | Pop7 | SS14 | 0 | 0 | 328 |
| | | | | | Pop8 | SS14 | 0 | 0 | 369 |

**Supplementary Figure 1:** Participant characteristics by *TPA* lineage across clinical sites.

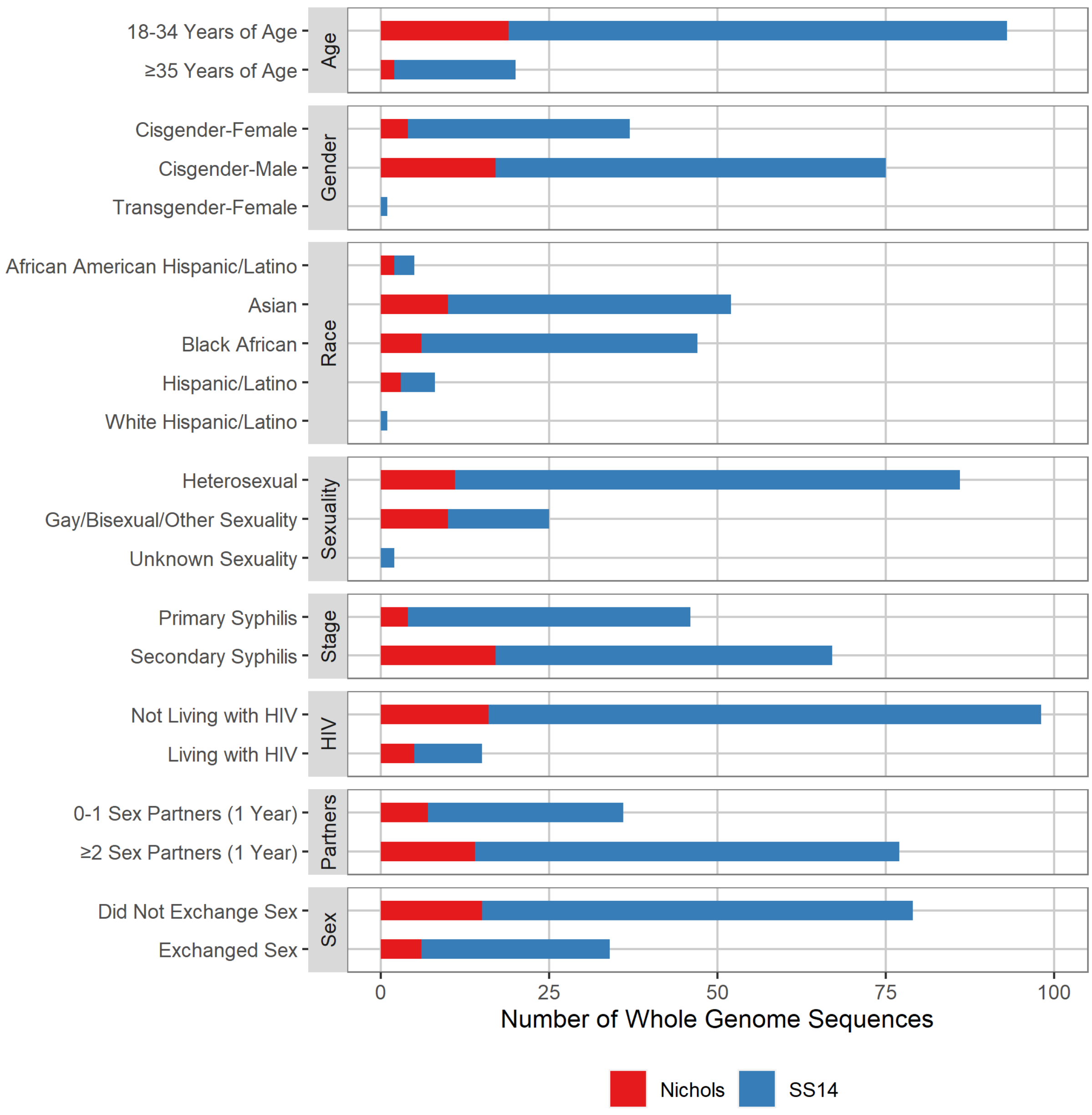

**Note:** Sex exchanged for goods/money/favors was assessed by partner for up to 5 sexual partners from the past year. If no partners named, classified as no.

**Supplementary Figure 2:** Principal components analyses illustrating relationship between *TPA* strains sequenced as part of this study (n=166) and global *TPA* genomic diversity (n=1,413). Points are not offset; thus, samples with identical SNVs overlap.

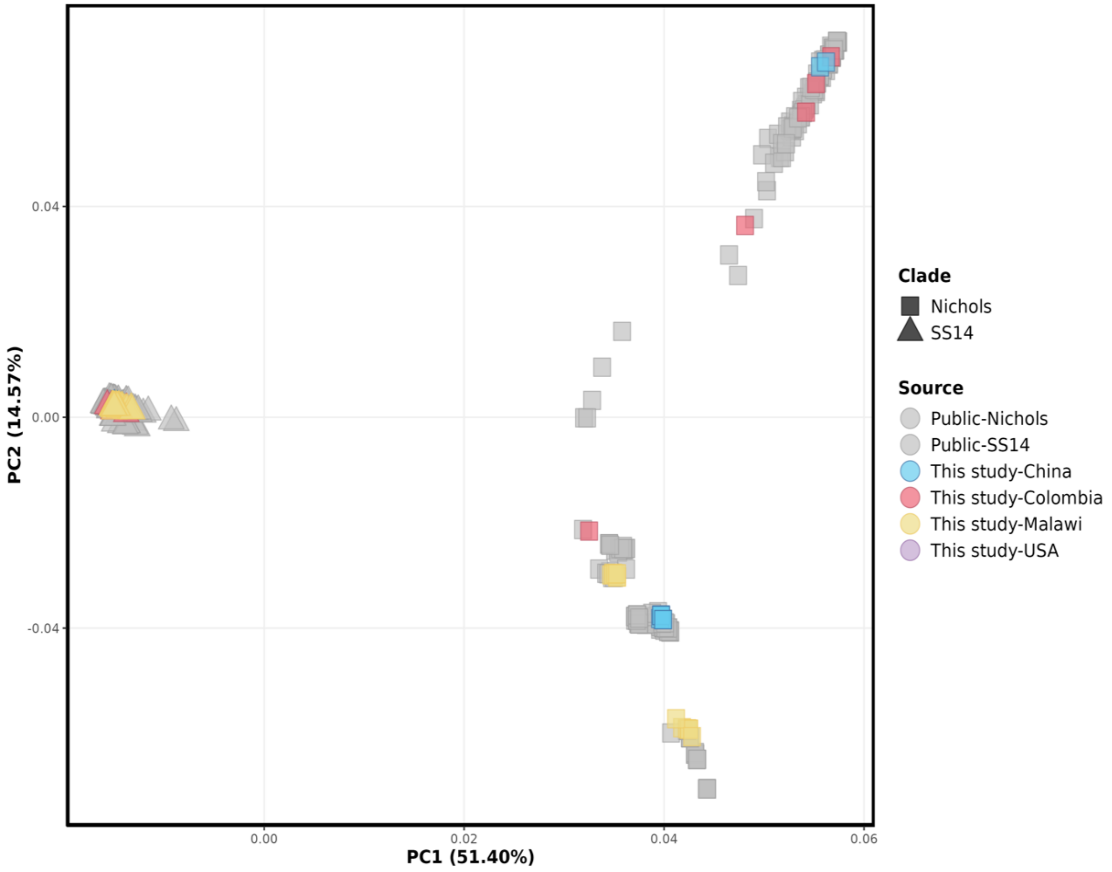

**Supplementary Figure 3:** Recombination-masked *TPA* whole-genome phylogeny (Figure 3), annotated by previously published sublineage and by *TPA* population determined using *baps* Bayesian modeling. Phylogeny derived from 166 individuals in this study,[a] 62 recently published genomes, and 5 reference genomes (*TPA*, red; *TPE*, blue; *TEN*, green). Nodes with >80% bootstrap support are highlighted with a black circle.

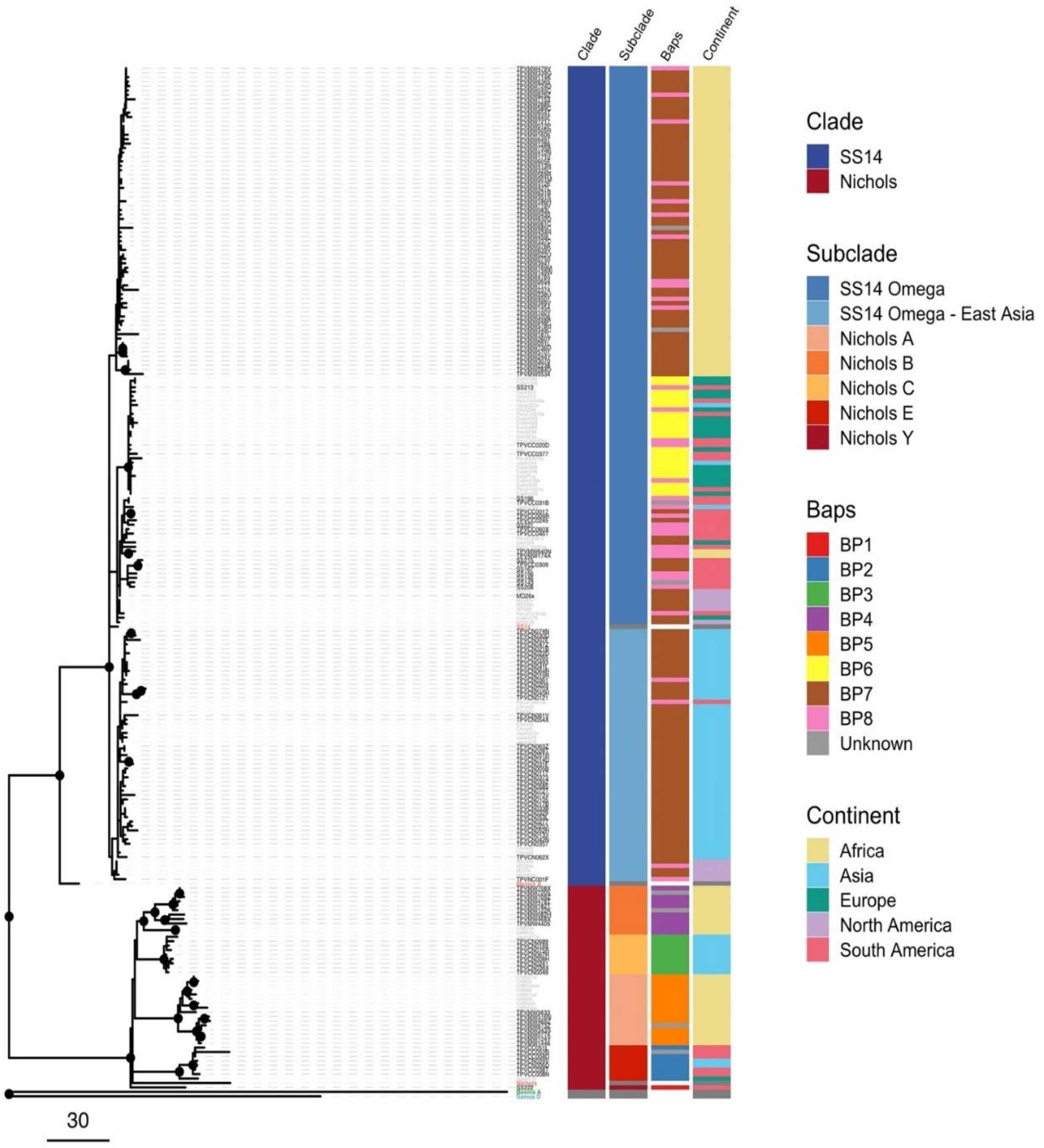

*TPA = Treponemal pallidum* sub. *pallidum; TPE = Treponema pallidum* subsp. *pertenue*; *TEN = Treponema pallidum* subsp. *endemicum*,
[a] Includes 43 DFM- negative specimens from Lilongwe, Malawi, and 10 specimens from Cali, Colombia, collected in a prior study.

**Supplementary Figure 4:** Allele frequencies of lineage-informative, highly differentiated (Fisher exact $p < 4{\cdot}451 \times 10^{-6}$) SNVs identified during analysis by lineage and geographical region, excluding *tpr* family, *tp0470*, and *arp* genes, with gene annotation by *SnpEff*. SNV coordinates reference the Nichols genome.

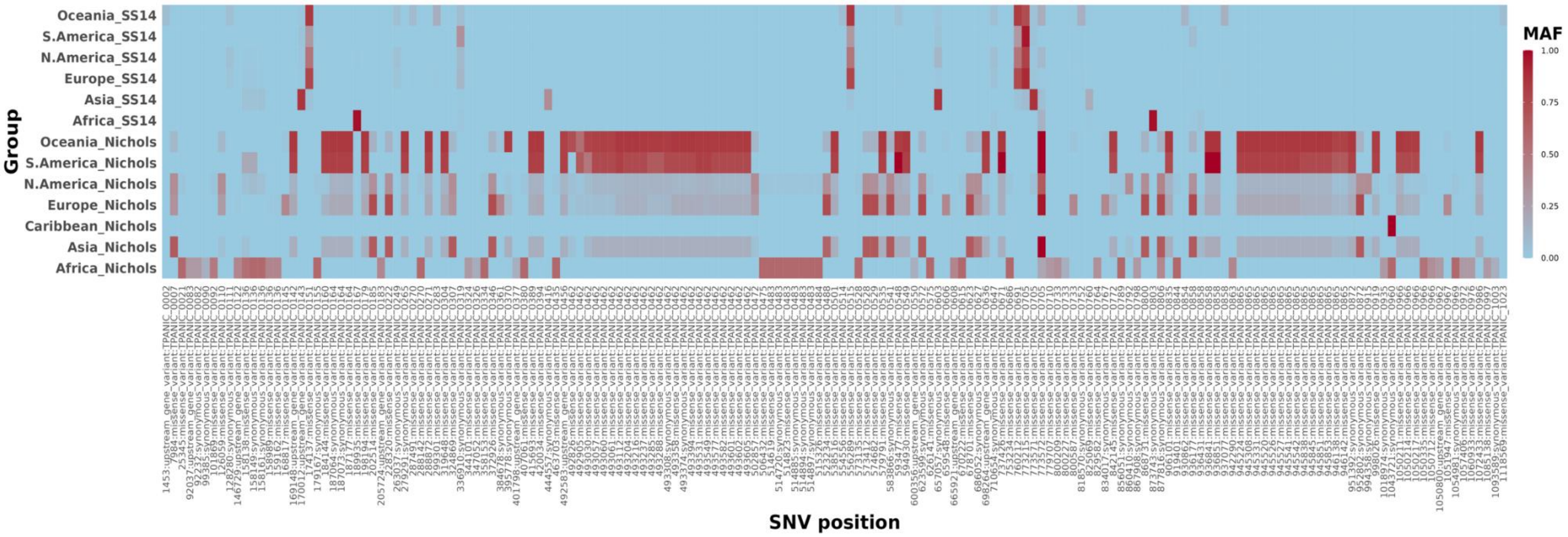

SNV = single nucletotide variant

**Supplementary Figure 5:** Allele frequencies of lineage-informative, highly differentiated (Fisher exact $p < 4{\cdot}451 \times 10^{-6}$) SNVs identified during recombination-masked analysis by *baps* TPA population membership, excluding *tpr* family, *tp0470*, and *arp* genes, annotated using *SnpEff*. SNV coordinates reference the Nichols genome.

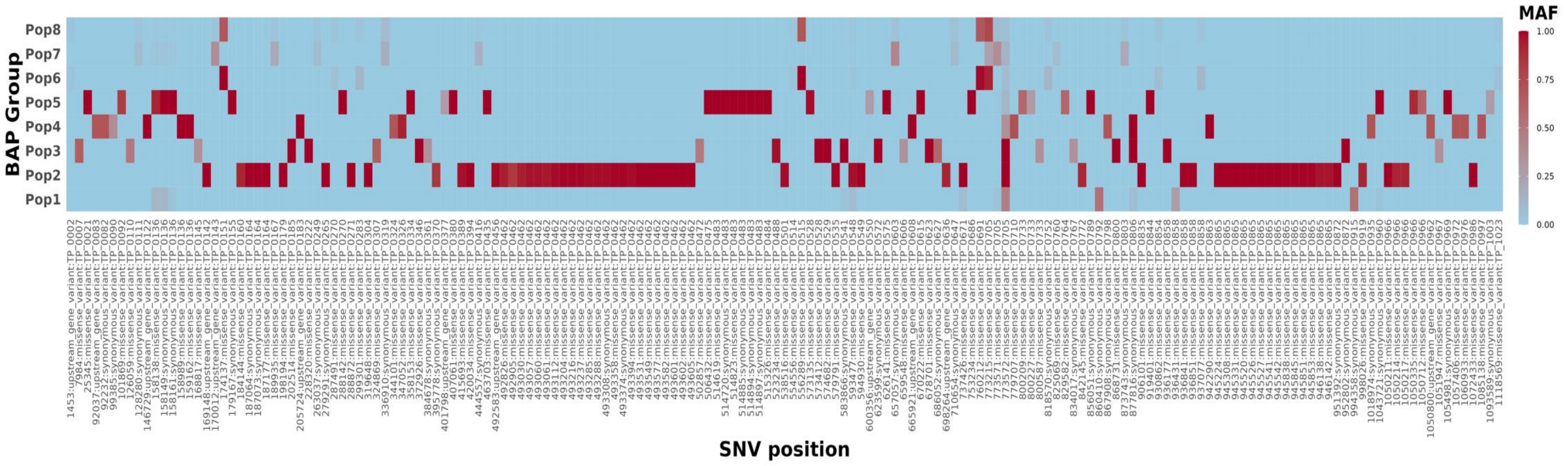

*TPA* = *Treponemal pallidum* sub. *pallidum; SNV* = single nucleotide variant

**References**

1. Delgado KN, Montezuma-Rusca JM, Orbe IC, et al. Extracellular loops of the *Treponema pallidum* FadL orthologs TP0856 and TP0858 elicit IgG antibodies and IgG(+)-specific B-cells in the rabbit model of experimental syphilis. *mBio* 2022; **13**: e0163922.

2. Chen W, Smajs D, Hu Y, et al. Analysis of *Treponema pallidum* strains from China using improved methods for whole-genome sequencing from primary syphilis chancres. *J Infect Dis* 2021; **223**(5): 848-53.

3. Lieberman NAP, Lin MJ, Xie H, et al. Treponema pallidum genome sequencing from six continents reveals variability in vaccine candidate genes and dominance of Nichols clade strains in Madagascar. *PLoS Negl Trop Dis* 2021; **15**(12): e0010063.

4. Hunt M, Mather AE, Sanchez-Buso L, et al. ARIBA: rapid antimicrobial resistance genotyping directly from sequencing reads. *Microb Genom* 2017; **3**(10): e000131.

5. Corander J, Marttinen P, Siren J, Tang J. Enhanced Bayesian modelling in BAPS software for learning genetic structures of populations. *BMC Bioinformatics* 2008; **9**: 539.

6. Nguyen LT, Schmidt HA, von Haeseler A, Minh BQ. IQ-TREE: a fast and effective stochastic algorithm for estimating maximum-likelihood phylogenies. *Mol Biol Evol* 2015; **32**(1): 268-74.

7. Hawley KL, Montezuma-Rusca JM, Delgado KN, et al. Structural modeling of the *Treponema pallidum* OMPeome: a roadmap for deconvolution of syphilis pathogenesis and development of a syphilis vaccine. *J Bacteriol* 2021; **203**(15): e0008221.

8. Gotelli NJ, Colwell RK. Quantifying biodiversity: procedures and pitfalls in the measurement and comparison of species richness. *Ecology Letters* 2001; **4**(4): 379-91.

9. Radolf JD, Kumar S. The *Treponema pallidum* outer membrane. *Curr Top Microbiol Immunol* 2017.
